# A train-and-assist device that upskills novices to strengthen the workforce and expand diagnostic access

**DOI:** 10.1101/2025.11.11.25339793

**Authors:** Minkyo Lee, Xinyue (Penny) Pei, Si Hyung Jin, Natasha Shelby, Rani Gera, Alexander Viloria Winnett, Colin F. Camerer, Mahbubur Rahman, Nils Pilotte, Steven A. Williams, Rustem F. Ismagilov

## Abstract

Artificial intelligence and automation technologies are displacing millions of workers across industries in developed countries, while many developing nations continue to grapple with chronically high unemployment. Meanwhile, healthcare laboratories—particularly in resource-limited settings (including rural and community sites within high-income countries)—face acute shortages of trained staff and the high cost of molecular diagnostics. Here, we propose a “train-and-assist” class of devices that aims to both (i) upskill—rather than replace—workers and (ii) expand diagnostic capacity in a cost-effective way. We describe a device that trains and assists laboratory-inexperienced personnel to perform sample-pooling procedures, which enable high-performance molecular testing at lower costs and higher throughput. A 48-participant user study demonstrated that the device enabled both skill acquisition and high-accuracy pooling. A device-validation study using clinical stool specimens demonstrated that device-assisted pooling agreed 100% with individual assays for soil-transmitted helminths, which affect more than 1.5 billion people worldwide.

**Teaser:** We propose a new class of technologies that train and assist personnel to learn skills for complex laboratory tasks, with the goal of strengthening the diagnostics workforce and expanding healthcare access.

## Introduction

Labor market imbalances present a key challenge in both developing and developed countries. Many developing countries have high unemployment and lack training opportunities. In developed countries, advances in artificial intelligence (AI) and automation are reshaping the labor market across multiple sectors^1,2^, with projections that millions of workers may be displaced by AI within the next decade^2,3,4^. Healthcare, in contrast, has historically suffered from a global shortage of skilled workers in both developing and developed countries, particularly for clinical diagnostic services^5,6^. Persistent vacancies are documented in both low-income regions and high-income countries such as the United States^8,9^, Canada^10^, and the United Kingdom^11^. The reasons for the lack of skilled clinical laboratorians are complex, but two known drivers are the limited training and certification programs and the challenges and burdens associated with entry-level requirements and licensure.^8^ Although automation and AI will offset some healthcare labor needs^7^, they will not offset the growing demand for healthcare services that require skilled workers^1,2^. Specifically, rural hospitals, community pharmacies, and long-term-care facilities remain diagnostically underserved even within wealthy health systems. In both developing and developed countries, the coexistence of surplus labor in some sectors and persistent shortages in diagnostics highlights an urgent need for innovative technologies that can rapidly upskill staff and enable newcomers to perform critical tasks with confidence **(Fig. 1*A*)**.

**Figure 1.**
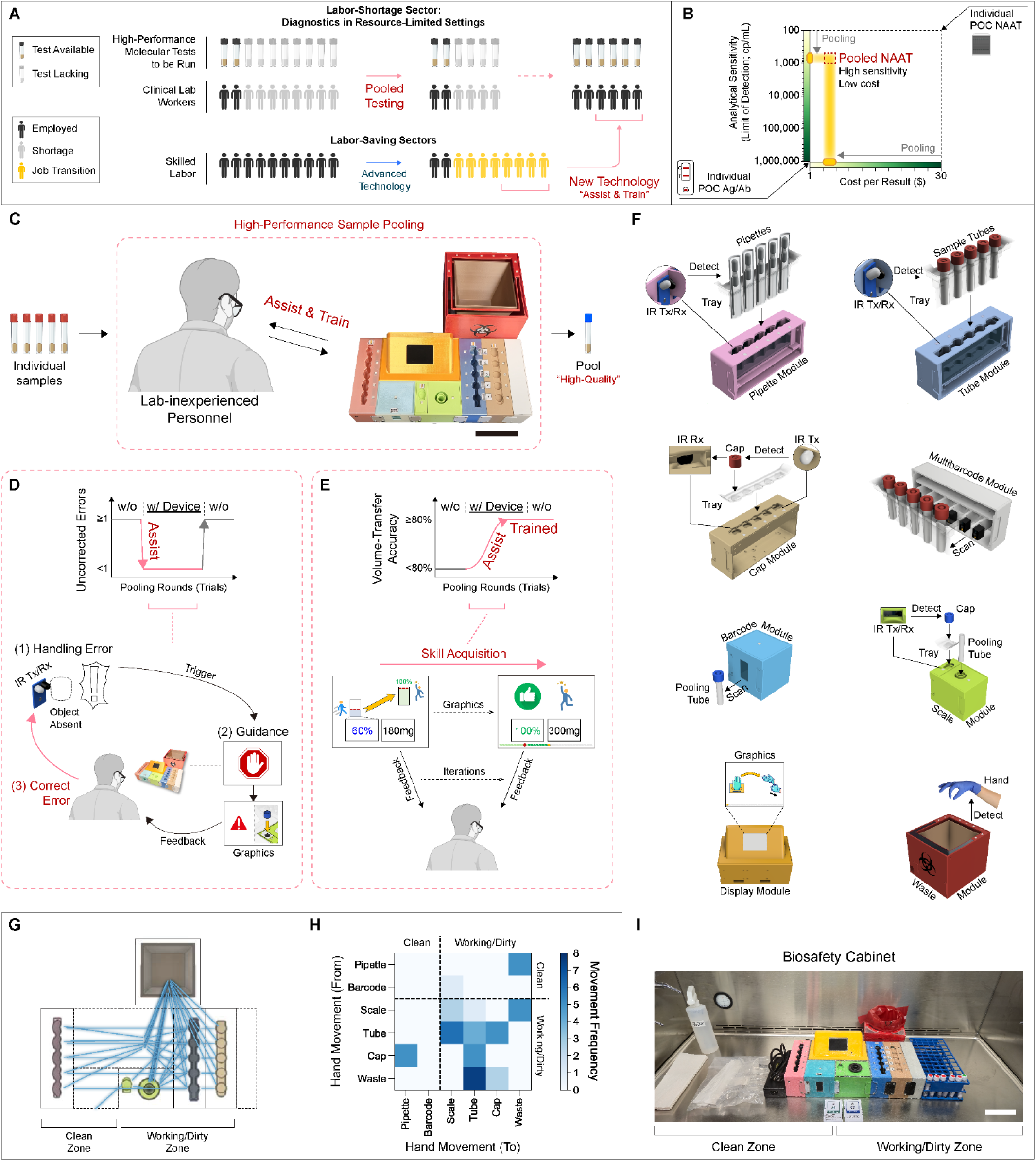
An interactive and instructional device for assisting and training users to perform diagnostic sample-pooling. (***A***) Clinical laboratories suffer from shortages of trained workers (top), while other workplace sectors experience reduced labor needs after implementation of advanced technologies (bottom). A novel class of assistive/training technology enabling pooled nucleic acid amplification testing (NAAT) would resolve the labor shortage and scale-up high-performance testing in resource-limited settings. (***B***) High-analytical-sensitivity pooled NAATs provide decreased cost-per-result levels comparable to low-analytical-sensitivity POC Ag/Ab tests. (***C***) Instructional device to assist and train laboratory-inexperienced personnel in pooling. Scale bar: 10 cm. (***D***) Device-assistive effect reducing user errors with use of the device. (***E***) Device-training effect improving volume-transfer skills over multiple pooling iterations. (***F***) Device modules house electronic sensors to monitor the handling workflow. (***G***) Expected movement of materials by user (blue lines) while using the device. (***H***) Matrix of expected hand movement frequency between two device modules during the pooling. Dashed lines separate clean and working/dirty zones. (***I***) Device within a biosafety cabinet alongside materials (e.g., towels, disinfectants, etc.) for clinical sample pooling. Scale bar: 10cm.

Beyond workforce shortages, the diagnostics field has faced another critical challenge: limited access to high-performance testing in resource-limited settings (RLS)^12^. Nucleic acid amplification tests (NAAT) are essential tools to combat high-burden infectious diseases globally, including human immunodeficiency virus (HIV)^13^, hepatitis C virus (HCV)^14^, tuberculosis (TB)^15^, neglected tropical diseases (NTDs)^16,17,18^, and respiratory viruses such as influenza^19^, respiratory syncytial virus (RSV)^20^ and severe acute respiratory syndrome (SARS)^21^. NAATs with high analytical sensitivity and specificity that can be used at the point-of-care (POC) are key, particularly for underserved populations without access to clinical laboratories^22^. POC testing can enable patients to swiftly receive the correct treatment, reducing the risk of being lost to follow-up^23^. Yet, despite their high utility, POC NAATs are either unavailable or prohibitively expensive in many contexts – both in high-income countries (HICs)^24,25,26^ and in low-and-middle income countries (LMICs)^14,15,27^. For example, integrated sample-to-answer molecular POC tests exist for HCV RNA, but they are not as widely used as HCV POC antibody tests due to the high cost per test^14^.

Sample pooling is a laboratory technique that reduces per-sample diagnostic testing costs and increases the total number of patients that can be tested^28,29,30,31,32,33,34^. In sample pooling, equal volumes from specimens from different individuals are pooled together and input into a single diagnostic test. Pooling thereby reduces the number of total tests required to be run and subsequent cost for each result. For high-sensitivity POC NAATs, the cost per sample with pooling becomes similar to low-sensitivity POC immunoassays^31,35,36,37^. Although pooling dilutes individual specimens, high-analytical-sensitivity molecular tests still yield the correct qualitative results for the pools^28,29,30,31,32,33,34^ and in many cases achieve far superior analytical sensitivity^31,32,38^ than POC immunoassays **(Fig. 1*B*)**.

Pooling specimens for use with NAAT diagnostics can enable public-health surveillance on a scale that is currently cost-prohibitive^33,34,39,40,41^. One example is the surveillance for soil-transmitted helminths (STHs), an NTD that affects >1.5 billion people^18^. Mass drug administration (MDA) using anti-helminth preventive chemotherapy (PC) has reduced disease prevalence; however, to identify when MDA can be stopped (when prevalence of moderate-to-heavy intensity infections is below 2%^18^), large-scale diagnostic testing must be performed^42,43^. Current STH testing is performed by manual DNA extraction followed by qPCR^16,34^; therefore pooling of raw samples upstream of DNA extraction would reduce the cost of and increase the throughput of testing^34^. Because most STH-vulnerable regions are in LMICs where testing resources are limited, sample-pooling for NAATs could logistically and financially enable testing at the scale needed to inform cessation of MDA^34^.

However, the manual sample-pooling process is error-prone^29,32,33,44^. Skilled personnel must safely handle multiple clinical specimens^33,44^, transfer accurate amounts of samples to each pool^29^, avoid cross-contamination^32^, and maintain documentation^32,33,44^. Automated liquid handlers for pooling exist^33^, but their high cost and a lack of laboratory infrastructure make them infeasible in most RLS^45^. Sample pooling is therefore inaccessible in regions where it is most critically needed to expand testing.

Here, we propose a new class of technologies that train and assist personnel to learn skills for complex laboratory tasks, with the goal of strengthening the diagnostics workforce and expanding healthcare access. As the initial example of technology in this class, we designed a low-cost, instructional pooling device that assists and trains laboratory-inexperienced personnel to pool clinical specimens for subsequent diagnostic testing **(Fig. 1*C* and Supplementary Movie S1),** which we illustrate with STH testing. We anticipate at least three contexts in which the instructional pooling device would have high impact: (i) rapid and widespread molecular diagnostic testing, such as during outbreaks and other testing surges, (ii) manual pooling tasks in RLS where automated equipment such as liquid handlers are cost-prohibitive or otherwise impractical, and (iii) programmatic screening/surveillance on a broad scale (such as for disease-elimination efforts).

Pooling was an important strategy to mitigate testing surges during the COVID-19 pandemic^33^, but was largely limited to labs that could perform automated pooling, whereas with the instructional device, pooling could be used on a massive scale, even in labs with limited resources and regions with minimally trained personnel. In decentralized RLS (e.g., mobile clinics) that collect specimens, the device is designed to help minimally trained workers perform high-quality, on-site pooling of aliquots with minimal human error. The resulting pooled tubes could be tested locally on POC NAAT platforms (e.g., Cepheid GeneXpert) or using existing manual workflows. In centralized RLS that must process larger numbers of specimens manually for screening/surveillance—as is the case with NTD testing—the device is intended to assist with pooling of primary specimens while automatically logging sample barcodes and pool-quality metrics, significantly increasing number of results that can be obtained with the manual NAAT workflows.

To evaluate the utility of the device, we conducted a user study with a cross-over design to demonstrate device usability, and to qualitatively and quantitatively determine the device’s assistive **(Fig. 1*D*)** and training effects **(Fig. 1*E*)** compared with traditional printed (paper) instructions. We also conducted a device-validation study using STH clinical stool samples to demonstrate the utility of the device for pooled molecular testing that would enable public health surveillance of NTDs.

## Results

### Design features of instructional device for POC pooling

The instructional pooling device is built with open-source embedded systems, interfacing with eight 3D-printed, independently-replaceable modules for the application of 5-sample pooling (combining five individual samples into a single pool) **(Fig. 1*F*).** The device modules are equipped with low-cost, off-the-shelf electronic components, including 940 nm infrared diodes for object detection and 640×480-resolution barcode scanners for sample identification (**Supplementary Figs. S1–2**; see Methods for electronic components details), to monitor objects such as sample tubes, caps, and individually wrapped pipettes **(Fig. 1*F* and Supplementary Fig. S3)**.

An optimized layout for left-to-right workflow separates a clean zone, where non-biohazardous materials are placed, and a working/dirty zone **(Fig. 1*G*)**. This layout reduces the frequency of hand movement across clean and working/dirty zones to prevent cross-contamination and minimize biosafety risks **(Fig. 1*H*)** within a limited space, such as a small table or biosafety cabinet **(Fig. 1*I*)**.

To enable accurate transfer of sample volumes in RLS where calibrated lab-grade pipettes are not available, the device is designed to work with disposable dual-bulb pipettes that transfer set volumes (this study: 300 µL) of liquid **(Supplementary Fig. S4a)**. To protect the device against contamination from clinical specimens during the handling workflow, device-protective disposable polypropylene trays manufactured via vacuum-forming technology **(Supplementary Fig. S4b)** are placed over certain areas of the device modules.

The device incorporates several user-friendly design features, including colorblind-friendly color-coding and shape-based designs **(Supplementary Fig. S4c)**. Protective trays have unique shapes which pair exclusively to the corresponding module, similar to shape-sorting blocks **(Supplementary Fig. S4d)**. The device layout uses intuitive symbolic indicators to guide users through proper pooling procedures **(Supplementary Fig. S4e).**

### Device functionalities for easy-to-perform POC pooling

The device is designed for deployment in public-health laboratories, mobile clinics, or other near-patient settings that routinely use cartridge-based, sample-to-answer NAAT platforms (e.g., Cepheid GeneXpert) or standard benchtop PCR thermocyclers. To make the pooling workflow easy to perform, the device features step-by-step guidance, real-time monitoring of the workflow, and quality-control documentation. Instructional guidance via language-agnostic graphics **(Fig. 2*A* and Supplementary Text S1)**, and visual and audible cues are provided by an algorithm **(Fig. 2*B* and Supplementary Movie S2)** at every step. In parallel, the device monitors the user’s actions and detects errors in real time through dynamically configured, scenario-based interrupt handlers **(Fig. 2*C*)**. The device helps users fix correctable mistakes **(Fig. 2*D* and Supplementary Text S2 and Movie S3)**; if a detected mistake is uncorrectable, the device will guide the user to terminate the process **(Fig. 2*B* and Supplementary Text S2 and Movie S4)**.

**Figure 2.**
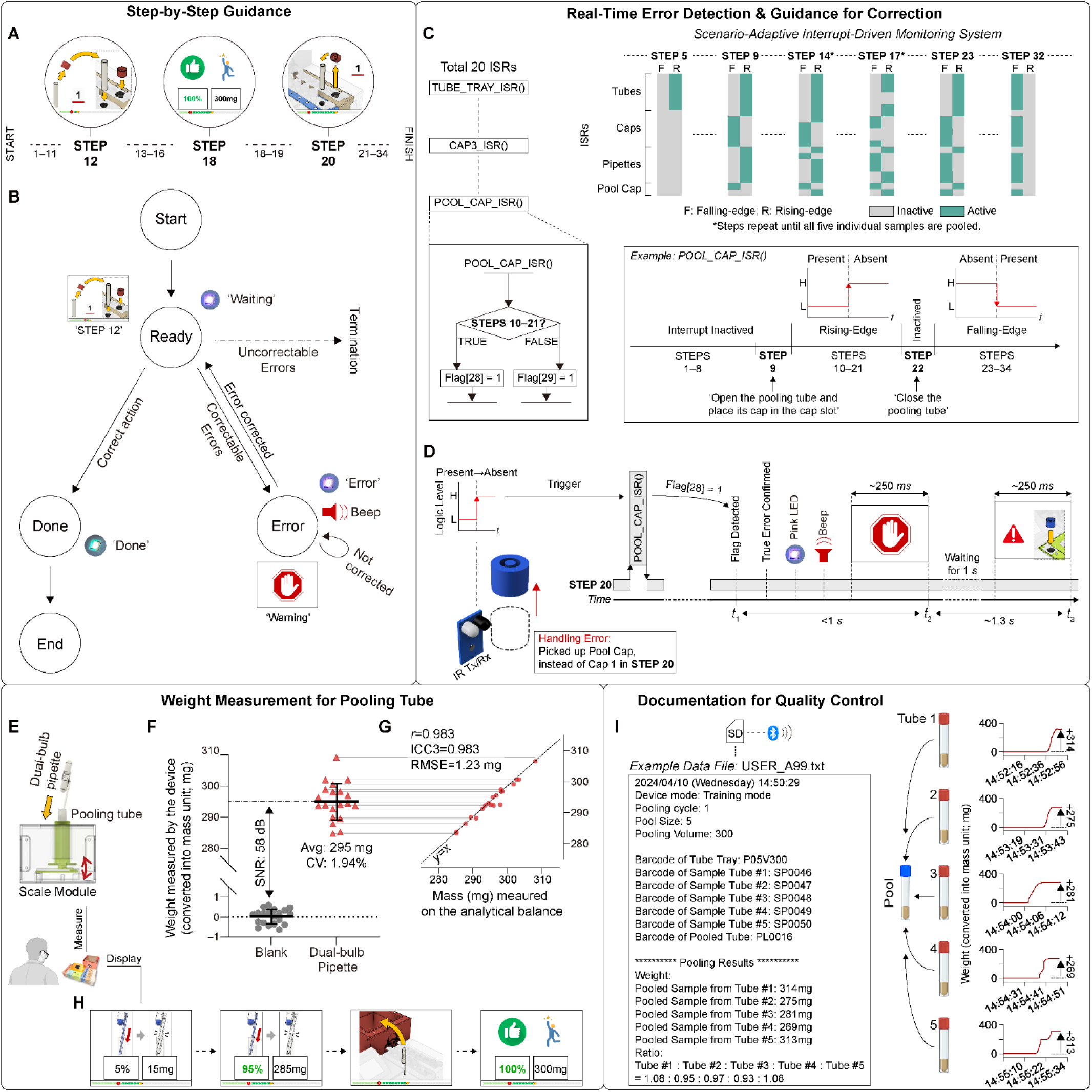
Device functionalities to enable easy-to-perform POC pooling. (***A***) Step-by-step guidance using language-agnostic graphics displayed on the device screen. (***B***) State diagram of algorithm operated at every guidance step. (***C***) Illustration of scenario-adaptive interrupt-driven system for real-time error monitoring. During the process, the system dynamically reconfigures trigger types between rising and falling edges for 20 interrupt service routines (ISRs). As an example, POOL_CAP_ISR() is inactive until STEP 9 that guides users to open the pooling tube and put its cap into the empty cap slot. During STEPS 10–21, the rising-edge is set to detect errors removing the cap from the slot. After STEP 22, the falling-edge is set until the end of process. When executed, the ISR raises a flag, based on scenarios. (***D***) Example process of real-time error detection and correction guidance when a pool cap is removed in STEP 20 which guides to pick up the cap of Tube 1. Rising-edge trigger causes the system to immediately pause the workflow and execute POOL_CAP_ISR(). The system detects the raised flag and provides guidance. (***E***) Illustration of Scale Module measuring the weight of pooling tube during the volume-transfer. (***F***) Scatter plot of weight measured on the device scale before and after the volume-transfer (n=20) of 300-μL water. ‘Blank’: the weight on the tared scale; ‘Dual-bulb pipette’: the weight of water transferred by pipettes; Avg, average; CV, coefficient of variation. (***G***) Correlation plot comparing measurements (n=20, red) between device and laboratory-grade analytical balance. Coefficients *r* and ICC3 were acquired through Pearson’s (*P*<0.00001) and interclass correlation analyses (*P*<0.00001). RMSE, root means squared error. (***H***) Real-time feedback of transferred sample amount and percentage on the screen. (***I***) Documentation feature. The example file shows recorded data, including scanned barcodes and weight of transferred samples. Inlet plots show the weight measurement during the volume-transfer.

Equipped with load cells and strain sensors **(Fig. 2*E* and Supplementary Fig. S5)**, the device measures the weight of transferred sample with >98% accuracy and precision (1.94%CV) during volume transfer steps, when users transfer liquid from individual tubes to the pooling tube using 300-μL dual-bulb pipettes **(Fig. 2*F*)**. The device measurement shows strong agreement with laboratory-grade analytical balance-based measurements (Pearson’s *r*: 0.983) **(Fig. 2*G*)**. During the volume-transfer activities, the weight values of each transferred sample and the percentages (relative to the target 300 µL) are shown to the user in real time, to reinforce the quality of pooled sample volumes **(Fig. 2*H*)**. All weight data are paired with tube barcodes and recorded on a removable Secure Digital (SD) card in the device, with wireless transmission capability **(Fig. 2*I* and Supplementary Fig. S1)**.

### User study design

We conducted a within-subject user study to evaluate two primary questions: (1) whether an instructional pooling device could assist laboratory-inexperienced personnel in improving their performance when handling challenging sample types, and (2) whether the device could effectively train these personnel to maintain high performance levels in sample pooling without the assistance from the device. We enrolled 48 participants, 37 (77%) of whom had limited or no prior laboratory experience **(Supplementary Fig. S6)**.

Participants were allocated to one of two training protocols (Protocols 1 and 2**; Supplementary Tables S1–5**). Protocol 1 was structured to examine the effectiveness of paper instructions in training participants, and the subsequent impact of introducing the device. Protocol 2 was designed to investigate the device’s effectiveness in improving performance during device use and maintaining performance (skill acquisition) after device removal. In each round of pooling, participants were guided to pool 300 µL of artificial respiratory or stool samples **(Supplementary Fig. S7)** with either paper instructions **(Supplementary Text S3)** or the device. Each participant completed 16 total rounds of 5-sample pooling over two non-consecutive days (“Day 1” and “Day 2”) **(Fig. 3*A*,*B*)**. After the exercises each day, participants evaluated the usability of instructional tools (paper or device) and perceived training effectiveness via questionnaires: post-study system usability questionnaire (PSSUQ; **Supplementary Tables S6–7**), Kirkpatrick training evaluation questionnaire (KTEQ; **Supplementary Fig. S8c**), and post-training effectiveness questionnaire (PTEQ; **Supplementary Fig. S8d–f**). See Methods for details.

**Figure 3.**
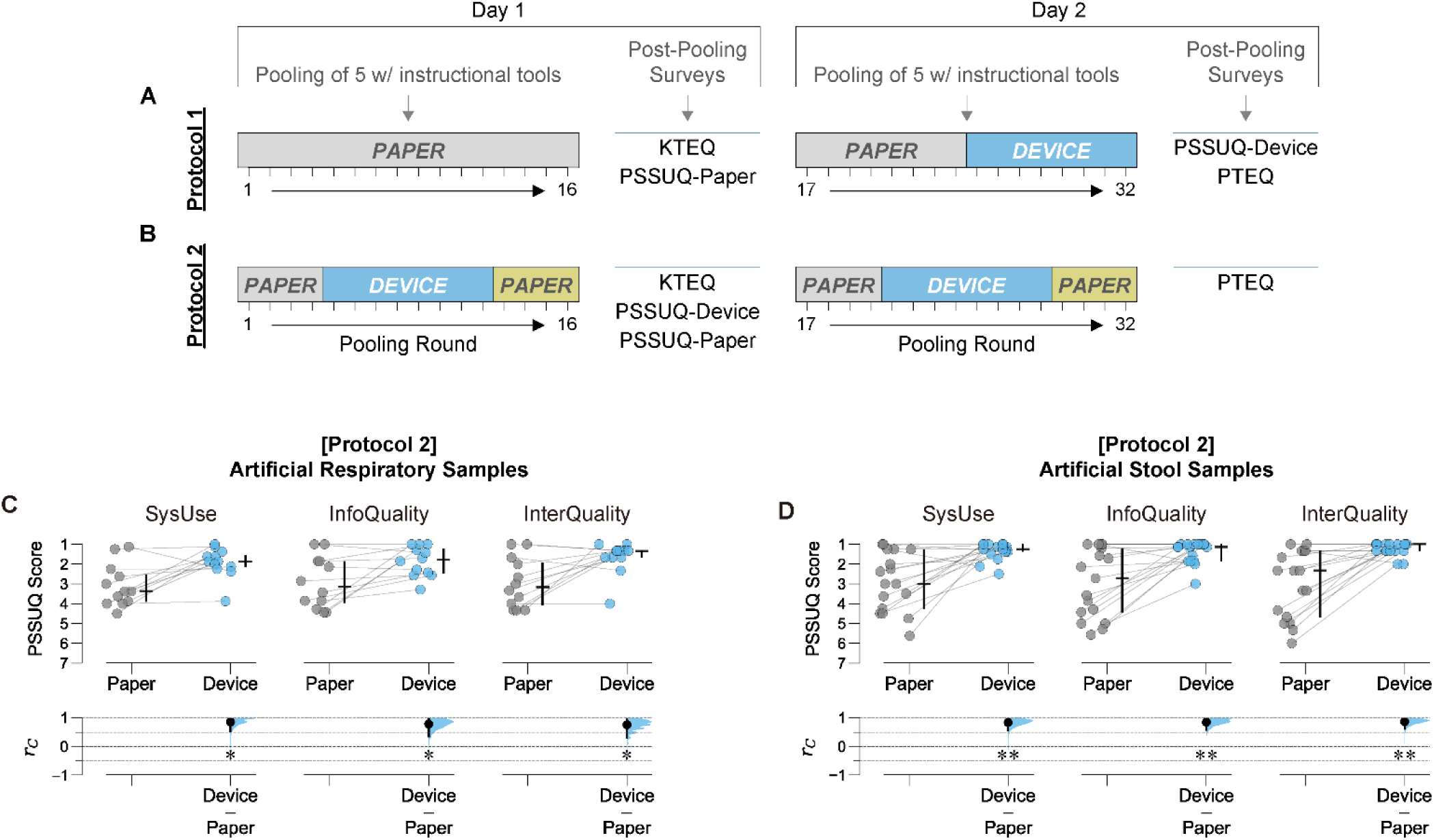
User study overview and self-reported results on usability surveys for instructional tools. (***A****,**B***), Diagrams of pooling training workflows for Protocol 1 (***A***) and Protocol 2 (***B***) and subsequent surveys (KTEQ, Kirkpatrick training evaluation questionnaire; PSSUQ-Paper, post-study system usability questionnaire for paper instructions; PSSUQ-Device, post-study system usability questionnaire for instructional pooling device; PTEQ, post-training effectiveness questionnaire). PAPER, paper-assisted pooling; DEVICE, device-assisted pooling. (***C***,***D***) PSSUQ scores for system usefulness, information quality, and interface quality of Day-1 paper instructions and instructional pooling device in Protocol 2 groups using artificial respiratory (***C***) and artificial stool samples (***D***). The score follows a 7-point Likert scale; lower scores indicate better usability. In the upper graphs, horizontal and vertical bold lines represent median and quantile ranges (Q1–Q3), respectively. Individual data points represent individual participants. In the lower graphs, effect sizes *r_C_*(the matched-pairs rank biserial correlation coefficients^58^) were calculated using one-sided Wilcoxon signed-rank tests comparing PSSUQ scores between the paper instructions and the device. The effect sizes were displayed as black dots with 95% confidence intervals (CI) (vertical lines) and resampled distributions of effect sizes (blue curves) given the observed data. The magnitude of the effect using *r_C_* is described by its distance from zero. *|r_C_|*^58,59^: 0.1=small; 0.2=medium; 0.3=large; ≥0.4=very large effect. Statistical significance was extracted from the one-sided Wilcoxon signed-rank tests with Benjamini-Hochberg correction (5% false discovery rate). Asterisks indicate significance levels: \**P*<0.05; \*\**P*<0.01. SysUse, system usefulness; InfoQual, information quality; InterQual, interface quality.

### Self-reported surveys: Users prefer the instructional device over paper instructions

Protocol 1 participants, who evaluated the usability of paper instructions on Day 1 and the device on Day 2 **(Fig. 3*A*)**, reported that both paper and device instructions were useful and provided high information and interface quality **(Supplementary Figs. S8a,b and S9)**. In contrast, Protocol 2 participants, who were exposed to both the paper instructions and the device on Day 1 **(Fig. 3*B*)**, reported that the device was more useful and provided better quality of information and interface than the paper instructions **(Fig. 3*C*,*D* and Supplementary Fig. S9)**.

Across both protocols, most users (≥75%) reported the perception of knowledge gain (Likert score ≥4; **Supplementary Fig. S8c**), skill acquisition (Likert score ≥4; **Supplementary Fig. S8c**), and skill utilization **(Supplementary Fig. S8d and Table S9)**. However, participants felt more confident performing high-performance pooling using the device compared with the paper instructions **(Supplementary Fig. S8e,f and Table S9)**.

### Protocol 1: Assistive effects of the device resulting in improved volume-transfer accuracy and handling error reduction

To investigate assistive effects of the device on sample-handling competency, we analyzed quantitative performance metrics measured during the pooling process in the Protocol 1 group. These metrics included the total number of uncorrected handling errors (*err*_all_; **Supplementary Text S4**), categorized as severe (*err*_severe_) and minor (*err*_minor_) **(Supplementary Table S10)**, and volume-transfer accuracy (Acc_pool_). The handling errors were recorded through researcher observation, while scanned barcodes and weight values were recorded via a customized data logger (paper-assisted pooling) or the instructional device (device-assisted pooling) (see Methods for details).

User performance was poor with paper instructions but improved significantly with the device. In Protocol 1, participants showed poor performance (average *err*_all_≥1, average Acc_pool_<80%) during paper-assisted pooling on both Day 1 and Day 2 **(Fig. 4*A*–*D*)**. Repetitive pooling with paper instructions did not result in improved performance (*r_C_*∼0 and Hedges’ g∼0) over all 24 rounds in either sample type **(Fig. 4*E*–*H* and Supplementary Tables S11–12)**, even in the analyses controlling for completion time spent on pooling process **(Supplementary Fig. S10, Text S5, and Tables S13–17)**. These objective measures contradict the subjective survey (KTEQ) results wherein participants reported skill acquisition from pooling with paper instructions on Day 1.

**Figure 4.**
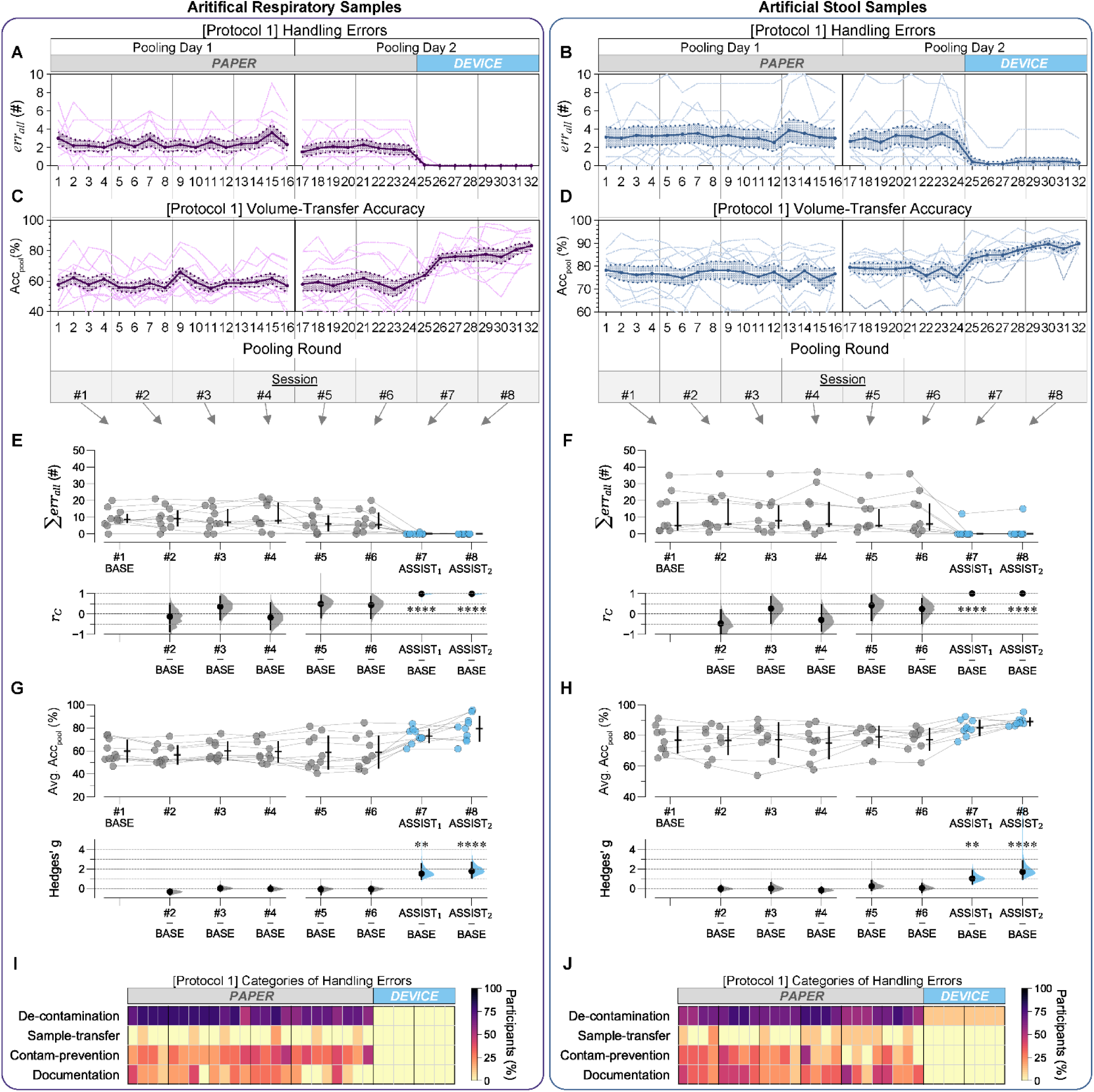
Assistive effects of instructional pooling device on user handling performance (errors and volume-transfer accuracy) in the Protocol 1 group. (***A*–*D***) Time-course of all uncorrected handling errors (***A*,*B***) and volume-transfer accuracy (***C*,*D***) for artificial respiratory (***A*,*C***) and stool (***B*,*D***) samples across pooling rounds for Protocol 1 group. Acc_pool_, volume-transfer accuracy; *err_all_*, all uncorrected handling errors. Semitransparent lines: individual participants; Bold lines: the means of *err_all_*(***A*,*B***) and Acc_pool_ (***C*,*D***) across participants; Shaded area borders: ±1 standard error of the mean (SEM). **e**–**h**, Gardner-Altman estimation plots comparing the sum of *err_all_* (***E*,*F***) and average Acc_pool_ (***G*,*H***) across pooling sessions (1 session=4 pooling rounds). In the upper graphs, horizontal bold lines represent median for non-normal data (***E*,*F***) and mean for normal data (***G*,*H***). Vertical lines represent quartile ranges (***E*,*F***) or ±1 SEM (***G*,*H***). Data points represent participants. Σ*err*_all,_ sum of *err_all_*; Avg. Acc_pool_, average Acc_pool_. In the lower graphs, effect sizes *r_C_* (***E*,*F***) and Hedges’ g (***G*,*H***) are shown from the comparison of the performance between the first session (#1, BASE) and each of the other sessions (#2–#8). *r_C_* (***E*,*F***) and Hedges’ g (***G*,*H***) were calculated using one-sided Wilcoxon signed-rank tests and one-sided, paired Student’s t-tests, respectively. The magnitude of the effect using *r_C_* and Hedges’ g is described by its distance from zero. *|r_C_|*^58,59^: 0.1=small; 0.2=medium; 0.3=large; ≥0.4=very large effect. Hedges’ g^58^: 0.2=small; 0.5=medium; 0.8=large effect. Effect sizes are displayed as black dots with 95% CI (vertical lines) and resampled distributions (curves). Statistical significance levels: \**P*<0.05; \*\**P*<0.01; \*\*\**P*<0.001; \*\*\*\**P*<0.0001 (extracted from the fixed effects of linear mixed-effect models using dummy coding). (***I*,*J***) Heatmaps showing the proportion of Protocol 1 participants making errors in four categories across 32 pooling rounds over two non-consecutive days. Color intensity indicates the percentage of participants making errors in each category per round.

Compared with the paper instructions, Protocol 1 users exhibited improved performance when using the device. During the device-assisted pooling, all participants had significantly (*r_C_*∼1) lower numbers of all uncorrected errors (∑*err*_all_) and significantly better (Hedges’ g≥1.5) average volume-transfer accuracy (Avg. Acc_pool_) **(Fig. 4*E*–*H* and Supplementary Tables S11–12)**.

To identify what kinds of errors were prevalent, we classified errors into four categories: decontamination, sample-transfer, contamination-prevention, and documentation **(Supplementary Table S18)**, and as either minor or severe. In the paper-assisted pooling, the most frequent errors observed (in ∼60% of users in each round) fell within the de-contamination category **(Fig. 4*I*,*J*)**, most of which were classified as minor **(Supplementary Fig. S11a and Table S18)**. The second most frequent errors occurred in the documentation category, all of which were considered severe **(Supplementary Fig. S11a and Table S18)**. When using the device, nearly all users (respiratory: 10/10; stool: 9/10) avoided or corrected these minor and severe errors **(Fig. 4*I*,*J* and Supplementary Fig. 12a–d and Tables S19–20)**.

### Protocol 2: Device-enabled training for high-performance volume transfer skills that persist after device removal

The quantitative performance metrics (i.e., *err*_all_, *err*_minor_, *err_severe_*, and Acc_pool_) were also analyzed for Protocol 2 participants. The repeat cross-over design of Protocol 2 clearly demonstrates significantly improved performance (fewer human errors and higher volume transfer accuracy) during device-assisted pooling, compared to paper-assisted pooling (*r_C_*∼1; Hedges’ g>0.9) for both sample types **(Fig. 5*A*–*H* and Supplementary Tables S21–22)**.

**Figure 5.**
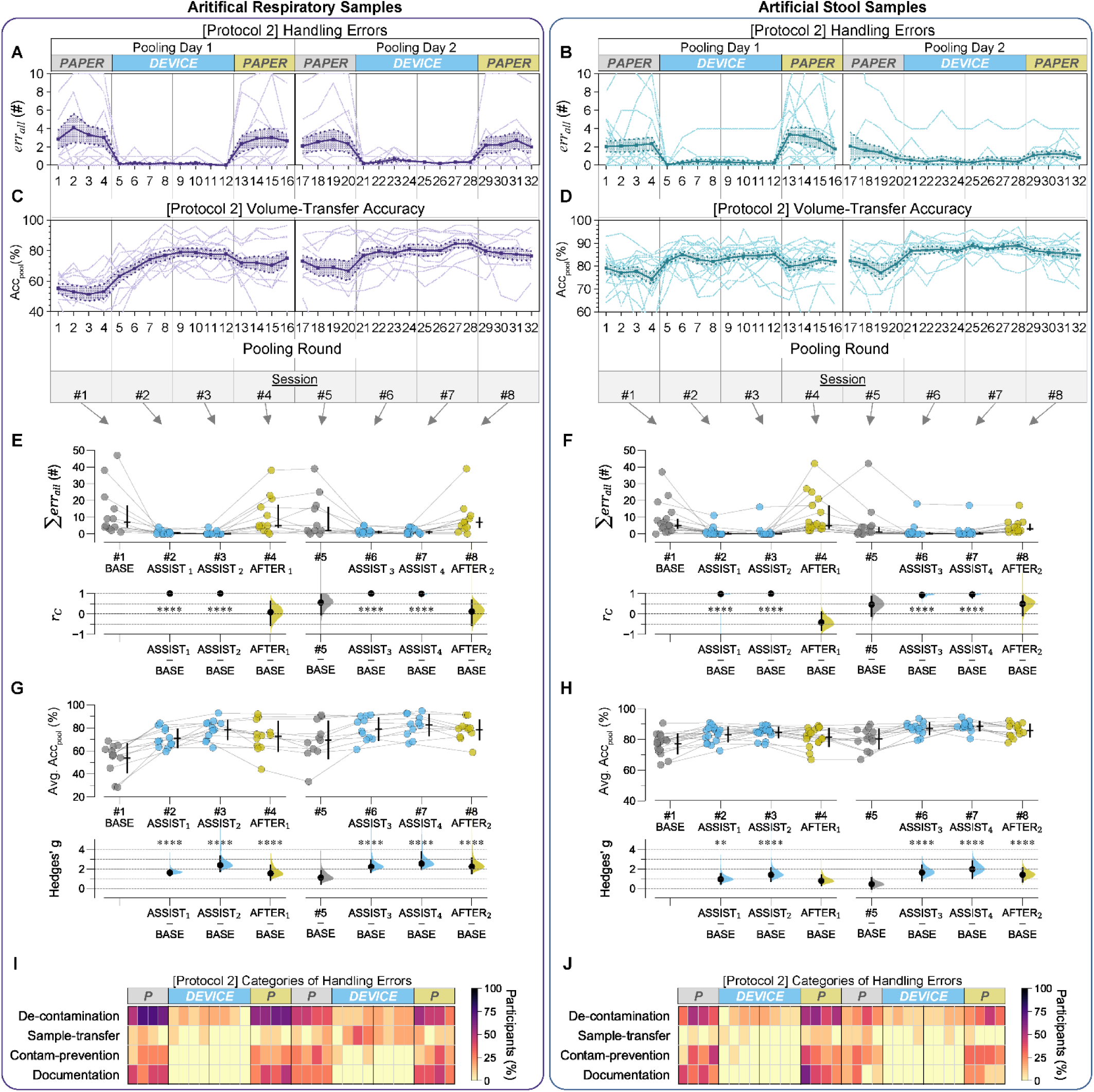
Assistive and training effects of instructional pooling device on user performance (errors and volume-transfer accuracy) in the Protocol 2 group. (***A*–*D***) Time-course of all uncorrected handling errors (***A*,*B***) and volume-transfer accuracy (***C*,*D***) for artificial respiratory (***A*,*C***) and stool (***B*,*D***) samples across pooling rounds for Protocol 2 group. Semitransparent lines: individual participants; Bold lines: the means of handling errors (***A*,*B***) and accuracy (***C*,*D***) across participants; Shaded area borders: ±1 standard error of the mean (SEM). (***E*–*H***) Gardner-Altman estimation plots comparing the sum of all uncorrected errors (***E*,*F***) and average volume-transfer accuracy (***G*,*H***) across pooling sessions. In the upper graphs, horizontal bold lines represent median for non-normal data (***E*,*F***) and mean for normal data (***G*,*H***). Vertical lines represent quartile ranges (***E*,*F***) or ±1 SEM (***G*,*H***). Data points represent participants. In the lower graphs, effect sizes *r_C_*(***E*,*F***) and Hedges’ g (***G*,*H***) are shown from the comparison of the performance between the first session (#1, BASE) and each of the other sessions (#2–#8). *r_C_* (***E*,*F***) and Hedges’ g (***G*,*H***) were calculated using one-sided Wilcoxon signed-rank tests and one-sided, paired student’s t-tests, respectively. The magnitude of the effect using *r_C_* and Hedges’ g is described by its distance from zero. *|r_C_|*^58,59^: 0.1=small; 0.2=medium; 0.3=large; ≥0.4=very large effect. Hedges’ g^58^: 0.2=small; 0.5=medium; 0.8=large effect. Effect sizes are displayed as black dots with 95% CI (vertical lines) and resampled distributions (curves) given the observed data. Statistical significance levels: \**P*<0.05; \*\**P*<0.01; \*\*\**P*<0.001; \*\*\*\**P*<0.0001 (extracted from results of linear mixed-effect models using dummy coding). (***I*,*J***) Heatmaps showing the proportion of Protocol 2 participants making handling errors in four categories across 32 pooling rounds over two non-consecutive days. Color intensity indicates the percentage of participants making errors in each category per round.

The reduction in handling errors was observed only during device-assisted pooling. Except for severe errors from Protocol 2/stool group, error counts (∑*err*_minor_, ∑*err*_severe_, and ∑*err*_all_) increased when participants used paper instructions, even after using the device (|*r*_C_|<0.5; BASE vs. AFTER_2_) **(Fig. 5*E*,*F* and Supplementary Figs. S12e–h and Tables S21, S23–24)**. Error patterns remained consistent for individual participants before and after the device use **(Fig. 5*I*,*J* and Supplementary Figs. S11b and S13)**. These results indicate that device-assisted pooling in Protocol 2 was insufficient to train users to avoid handling errors when not using the device.

In contrast, the device produced a training effect on volume-transfer accuracy, even after the participants were no longer using the device. Individual participants showed improved volume-transfer accuracy (Avg. Acc_pool_; Hedges’ g>1) during device-assisted pooling (ASSIST_1-4_) and maintained high accuracy (Hedges’ g>0.5) in the post-device session (AFTER_1,2_) **(Fig. 5*G*,*H* and Supplementary Table S19)**.

Although participants performed slowly with the device **(Supplementary Fig. S10)**, these assistive and training effects of the device on error metrics and Acc_pool_ in Protocol 2 remained statistically significant even in the analyses controlling for completion time as a covariate **(Supplementary Text S5 and Tables S25–29)**. There was no statistically significant difference in the baseline performance (for all metrics; *err*_all_, *err*_minor_, *err_severe_*, and Acc_pool_) between Protocol 1 and 2 groups (*P*>0.05; interaction terms comparing Session #1 in the linear mixed-effect analyses). Both accuracy and handling errors were also independent of the interval between Day 1 and Day 2 **(Supplementary Fig. S14)**, which differed among participants (median [Q1, Q3]: 7 [4, 8] days).

Use of the device resulted in high-performance volume-transfer, which was determined by the number of high-quality pools showing high accuracy (Acc_pool_≥80%) and precision (CV_pool_≤25%) per pooling round **(Supplementary Text S6 and Fig. S15)**. Participants produced high-quality pools in both device-assisted (ASSIST_1-4_) and post-device sessions (AFTER_1-2_) significantly more (*r_C_<*–0.5) than those in the first paper-assisted session (BASE) **(Supplementary Fig. S16a–d and Tables S30–31)**. In Protocol 2, 15 participants (72.4% in respiratory; 53.8% in stool) showed significant performance improvements (adjusted *P*<0.05; BASE vs. ASSIST_4_), with 14 of these participants maintained their high performance in the post-device session (AFTER_2_) **(Supplementary Fig. S16e,f and Table S32)**. These results suggest both assistive and training effects for volume-transfer skills.

### The device enables users to achieve high-performance pooling of artificial clinical samples

We next investigated whether the device could enable users to achieve high-performance pooling, defined as high-accuracy and high-precision volume transfer (Acc_pool_≥80% & CV_pool_≤25%) and perfect handling (*err*_all_=0).

Considering all quantitative metrics, the device significantly (*r_C_*∼–1) assisted users in achieving high-performance pooling for both sample types **(Fig. 6*A*–*D* and Supplementary Figs. S17–19 and Tables S33–34)**. The assistive effects were significant (adjusted *P*<0.05) for most individuals (58.3–72.7% in the respiratory group; 76.5–76.9% in the stool group) shown in device-assisted sessions #3 and #7 **(Fig. 6*E*,*F* and Supplementary Table S35)**. Consequently, up to 92.8% of participants achieved high-performance pooling in ≥2 of 4 pooling rounds during the last device-assisted session #7 (ASSIST_4_).

**Figure 6.**
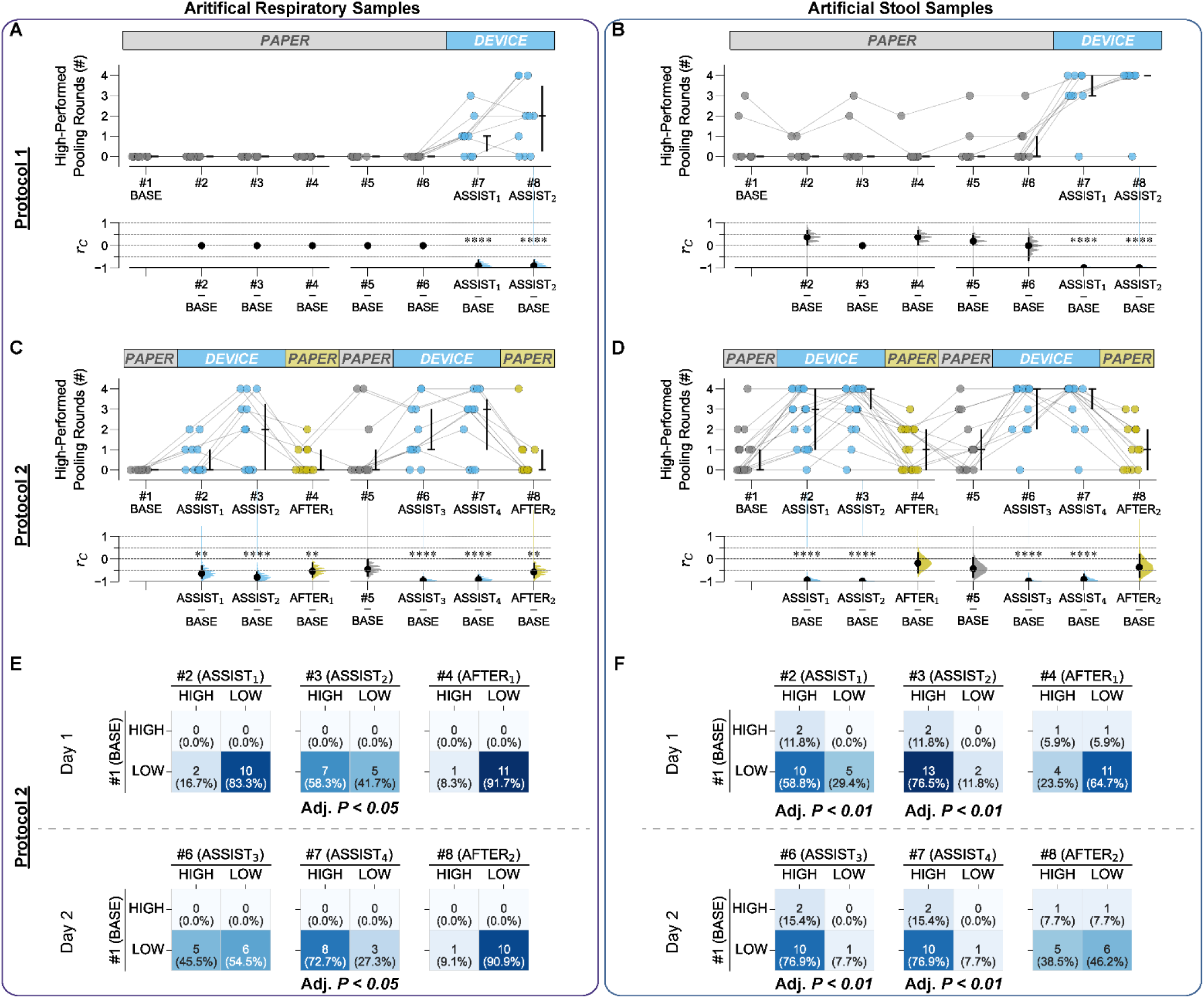
Device-assisted high-performance sample pooling, defined as pooling with volume-transfer accuracy Acc_pool_≥80%, precision ≤25% CV_pool_ and perfect handling (i.e., *err*_all_=0). (***A*–*D***) Gardner-Altman estimation plots comparing the number of high-performance pooling rounds between the first pooling session (#1, BASE) and subsequent sessions (#2–#8) in Protocol 1 (***A*,*B***) and Protocol 2 groups (***C*,*D***). Upper graphs: Data points with median (horizontal bold lines) and quartile ranges (vertical lines) for individual participants. Lower graphs: Effect sizes *r_C_* (black dots) from one-sided Wilcoxon signed-rank tests with 95% CI (vertical lines) and resampled distributions (curves). The magnitude of the effect using *r_C_* is described by its distance from zero. *|r_C_|*^58,59^: 0.1=small; 0.2=medium; 0.3=large; ≥0.4=very large effect. Statistical significance levels: \**P*<0.05; \*\**P*<0.01; \*\*\**P*<0.001; \*\*\*\**P*<0.0001 (extracted from results of linear mixed-effect models using dummy coding). (***E*,*F***) 2×2 contingency tables for McNemar’s tests comparing sample-pooling performance of Protocol 2 participants between the first pooling session #1 (BASE) and one of sessions #2–#8 separated by (“HIGH”) and low (“LOW”) performers. Color intensity (light to dark blue) highlights the relative distribution of participants within each individual 2×2 table, with darker blue indicating cells containing higher percentages. Note that the color scale is applied across all contingency tables within each sample type. Cell values: Number of participants in each performance category combination. HIGH, high performers who achieved ≥2 rounds of high-performance pooling during a 4-round session. LOW, low performers who achieved <2 rounds of high-performance pooling during a 4-round session. *P* values for statistical significance were determined by McNemar’s tests with Benjamini-Hochberg correction (5% false discovery rate).

### The device helps users create trackable pools with five consistent sample volumes and avoid poor-quality pools

Despite the significant assistive/training effects, a critical question remained: would these improvements translate to pools suitable for trackable and reliable pooled molecular testing? To address this question, we categorized pools into three classifications, defined in **Fig. 7*A***: *Correct*, *Incorrect*, and *Invalid* with additional sub-classifications for types. *Correct* pools are generated when participants accurately follow instructions and properly transfer individual samples. *Incorrect* pools result from procedural user errors that are not corrected nor avoided. *Invalid* pools result from the poor pool quality. They include those with a maximum mass pooling ratio (MPR_max_) ≥10, where MPR_max_ is defined as the ratio of the pool’s final weight to the minimum weight among any individual sample within the pool. A common cause of *Invalid* pools is inconsistent sample volumes, which could cause false diagnostic negative results, particularly when an insufficient volume of a weak-positive sample is pooled with negative samples.

**Figure 7.**
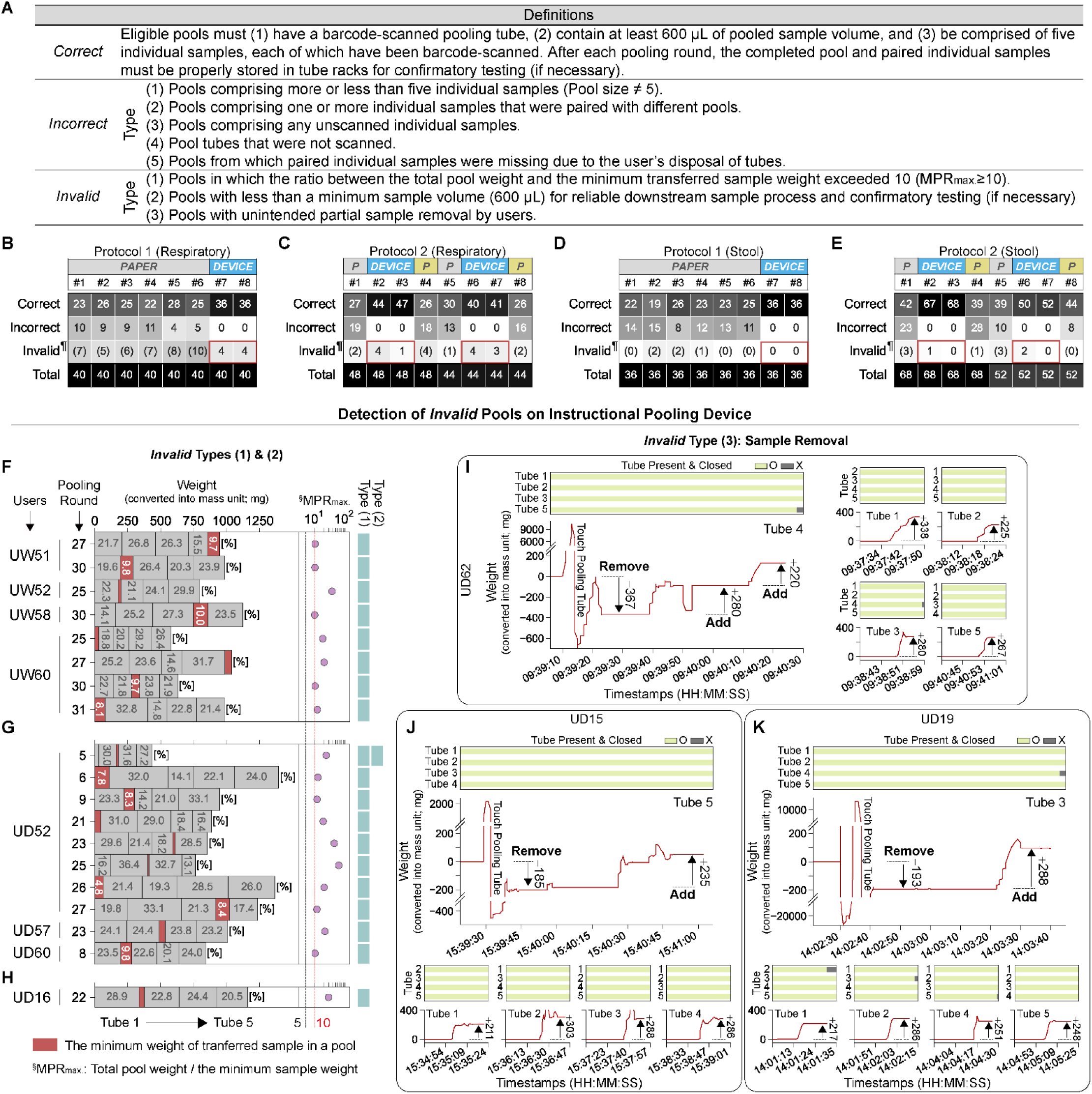
*Correct*, *Incorrect*, and *Invalid* pools produced during the user study. (***A***) Definitions of *Correct*, *Incorrect*, and *Invalid* pools. *Correct* pools meet all specified criteria. This category includes pools where users corrected procedural errors. Pools are classified as *Incorrect* or *Invalid* if they meet any subtype condition. (***B***–***E***), Number of pools produced across pooling sessions #1–#8 in Protocol 1 (***B***,***D***) and 2 (***C***,***F***) groups using artificial respiratory (***B***,***C***) and stool samples (***D***,***E***). ^¶^Pools that fall into both *Incorrect* and *Invalid* categories are counted as *Incorrect*. (***F***–***H***), Device-detected *Invalid* Types (1) and (2) pools produced by Protocol 1 (***F***) and 2 (***G***,***H***) groups using artificial respiratory (***F***,***G***) and stool samples (***H***). Each horizontal bar represents a pool of five samples (Tubes 1–5), with weight percentages. Maximum mass pooling ratio (MPR_max_), defined as the ratio of total pool weight to the minimum sample weight, is shown for each *Invalid* pool. Red: the lowest weight sample from each pool. Turquoise: *Invalid* subtype classifications. (***I***–***K***), Device-detected *Invalid* Type (3) pools through real-time monitoring of volume-transfer activities by users UD62 (***I***), UD15 (***J***), and UD19 (***K***) during Rounds 23rd, 5th, and 24th, respectively. The weight-time plots in each panel show measured weight changes in the pooling tube during the volume transfer in a single round. The large plot shows inappropriate activities where sample was removed from the pooling tube (Remove) before adding sample (Add) from an opened tube: Tubes 4 (***I***), 5 (***J***), or 3 (***K***). Small inlet plots show proper activities where samples were correctly transferred from other tubes to the pooling tube. Horizontal bars indicate real-time monitoring of tube presence and cap status in the device (green bar with O: tube present and capped; grey bar with X: tube absent or uncapped) for all other tubes. Numbers adjacent to arrows indicate the weight (mg) of transferred sample.

Device-assisted pooling significantly reduced *Incorrect* pools and increased the proportion of *Correct* pools compared with paper-assisted pooling **(Fig. 7*B*–*E*)**. When using the device, Protocol 2 users produced zero *Incorrect* pools (out of 424 pools), whereas with paper-assisted pooling they produced 135 *Incorrect* pools (31.8%) out of 424 total pools (Fisher’s Exact test*, P*<0.0001; **Supplementary Table S36**).

Crucially, the device was able to identify *Invalid* pools of the three types **(Fig. 7*A*)**. Through weight measurement, the device tracked individual pooled sample weights, total pooled sample weights, and volume-transfer activities during pooling. In parallel, the real-time monitoring of all tubes and caps verified that sample was correctly transferred from an opened individual tube into the pooling tube. As a result, total of 22 *Invalid* pools were identified: 18 *Invalid* Type (1) pools with large maximum mass pooling ratios (MPR_max_: 10–100) from 8 participants; one combined *Invalid* Type (1) & Type (2) pool (total volume: 516 µL; MPR_max_: 24.3) from one participant; and three *Invalid* Type (3) pools from participants who accidentally removed sample from pooling tubes **(Fig. 7*F*–*K*)**. In contrast, nearly three times as many (64) *Invalid* pools were generated during the paper-assisted pooling **(Fig. 7*B*–*E* and Supplementary Figs. S20–21)**; as paper-assisted pooling does not have built-in verification of sample transfer, these *Invalid* pools would likely go unrecognized in a clinical laboratory setting without the device.

### The device supports reliable pooling of clinical STH samples to enable high-performance pooled NAAT

To demonstrate the utility of the device for the pooling of clinical samples prior to NAAT, we used the device to create 5-sample pools using 42 archived clinical stool specimens from the previous field study of STH infections in children in Bangladesh (see Methods)^46^. Among these 42 archived specimens, 21 had previously tested positive for the STH *Ascaris lumbricoides* DNA **(Fig. 8*A*)**. Each individual sample was randomly selected and a biosafety-trained researcher used the instructional pooling device to pool each clinical sample with four (control) stool samples from healthy human donors.

**Figure 8.**
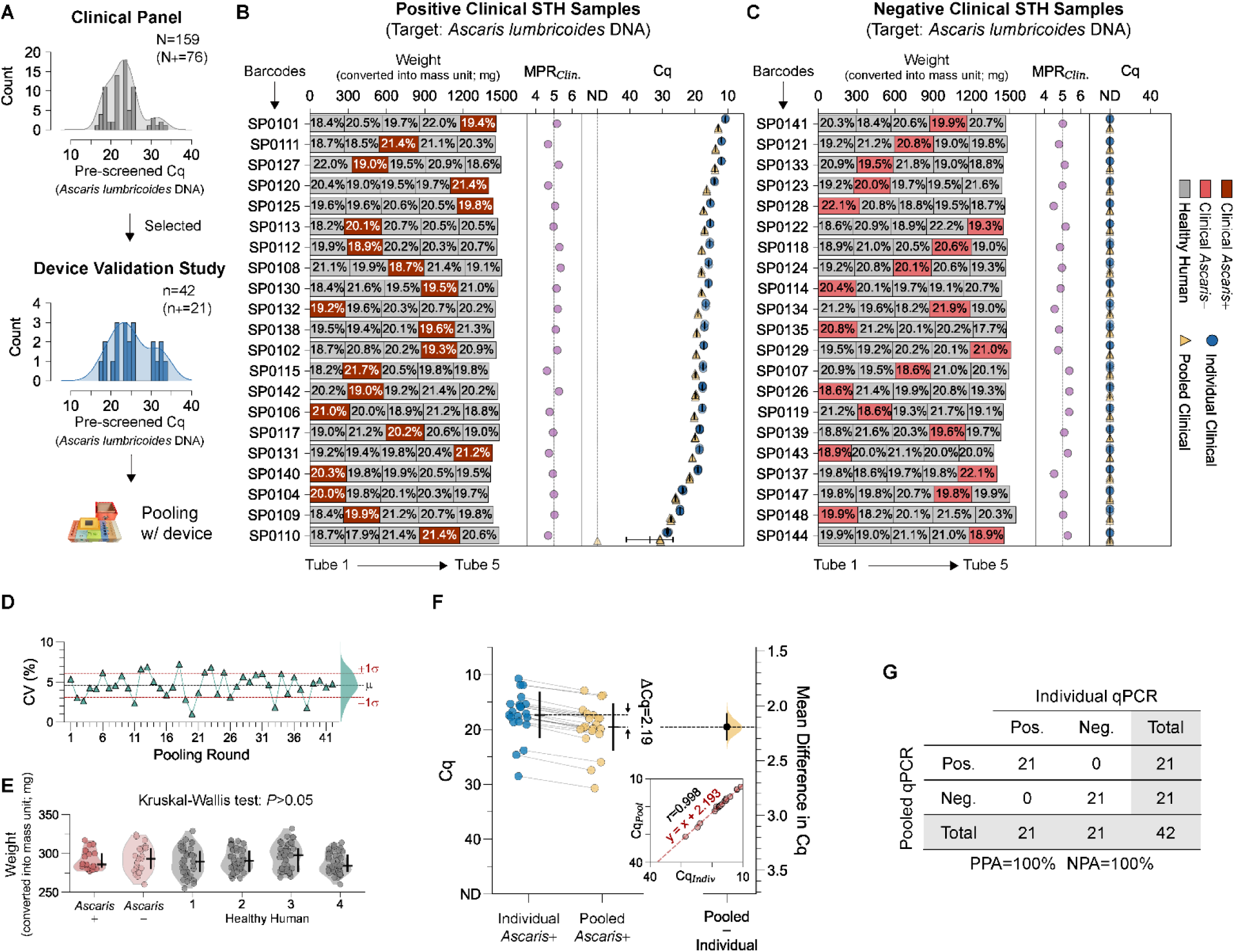
Device validation study for clinical sample pooling to detect *Ascaris lumbricoides* DNA from human stools collected from children in STH-prevalent regions. (*A*) Distribution of pre-screened Cq values of *Ascaris lumbricoides* DNA from archived clinical samples (N=159; 76 positives, 83 negatives). For this study, 42 samples (21 positives, 21 negatives) were selected through stratified random sampling to obtain a range of Cq values. **(*B*,*C*)** Weight distribution of five individual stool suspensions (one clinical and four healthy human) per pool. Barcodes represent unique identifiers for individual clinical samples, arranged by ascending Cq values. Horizontal bars represent a pool of five individual samples (Tubes 1–5), with the weight percentages. Red segments: clinical positive **(*B*)** and negative **(*C*)** stool suspensions. Grey segments: commercial stool suspensions from healthy human donors. The sequential position of the clinical STH sample was randomized among the four commercial samples in each pooling round. MPR_Clin._: mass pooling ratio in respect to each clinical sample (total pool weight/individual clinical sample weight). Cq values of all qPCR replicates in this study are shown for individual clinical (blue circle) and pooled samples (yellow triangle). Vertical lines: mean; horizontal lines: standard deviation. **(*D*)** CV values across pooling rounds. Green curve: CV density. Black line: mean (µ=4.56%). Red lines: ±1 standard deviation (s=1.47%). **(*E*)** Weight distributions of suspensions transferred from 21 clinical positives (Ascaris+), 21 clinical negatives (Ascaris-), and four unique healthy human samples to pools. Horizontal lines: median; vertical lines: quantile ranges (Q1–Q3). **(*F*)** Estimation plot comparing paired Cq values between individual (Cq_Indiv_) and pooled positive samples (Cq_Pool_). Horizontal lines: mean; vertical lines: standard deviations. Mean Cq difference (ΔCq=2.19) shown with 95% CI (vertical line) and resampled distributions of difference (curves). Inlet: correlation plot (Pearson’s *r*: 0.998). **(*G*)** 2×2 contingency table showing positive percent agreement (PPA) and negative percent agreement (NPA) of pooled versus individual qPCR testing.

The device demonstrated high-precision pooling (average CV<5% in volume transfer) of homogenized human stool suspensions **(Fig. 8*D*)**. The mass pooling ratio (MPR) of each clinical sample within a pool was close to the ideal value of 5 (mean: 4.998; std: 0.2459) **(Fig. 8*B*,*C*)**. No significant differences were observed between the pooled weight of individual STH clinical and control stool suspensions (Kruskal–Wallis test, *P=*0.314) **(Fig. 8*E* and Supplementary Table S37)**. For *A. lumbricoides* detection by NAAT (qPCR), the device-assisted pooling showed a mean cycle quantification shift (ΔCq; Cq_Pool_ – Cq_Indiv_) of 2.19 (95%CI=[2.06, 2.31]); this Cq shift is close to the expected shift (log_2_5 ∼ 2.32) for pools of 5, leading to a strong linear relationship between Cq_indiv_ and Cq_pool_ for STH positive specimens **(Fig. 8*F*)**. Perfect agreement was observed between STH diagnostic results from individual testing and 5-sample pools generated using the device **(Fig. 8*G*)**.

## Discussion

The rapid advancement of AI and automation technologies is reshaping global labor markets, creating an urgent need to develop effective strategies for workforce adaptation. Simultaneously, the persistent inaccessibility of high-performance molecular diagnostic testing remains one of the greatest barriers to global infectious-disease eradication efforts, particularly in RLS. Diagnostic shortages for major diseases (e.g., bloodborne, respiratory, STH), and testing backlogs during surges (e.g., COVID-19 pandemic) stem from the high cost-per-test, error-prone workflows, and a lack of trained laboratory personnel.

This work **(Fig. 1)** addresses all of these challenges by both assisting and training laboratory-naïve personnel to perform the complex workflow of sample-pooling, which has the potential to reduce the per-test diagnostic cost (without sacrificing test performance) and improve the workforce in RLS. The device **(Fig. 2)** is designed for a low-cost, open-source platform (∼$600; **Supplementary Table S38**), with further cost reductions possible through mass production and local sourcing. It features a user-friendly design for seamless workflows, real-time error monitoring, pool quality control, and documentation capabilities.

The user study **(Figs. 3–7)** demonstrated the utility of the device for assisting and training laboratory-inexperienced personnel to perform sample-pooling. Compared with paper instructions, device use reduced uncorrected handling errors (|*r*_C_|∼1) and raised mean volume-transfer accuracy (Hedges’ g>1). Device users retained better volume-transfer skills even after device removal, supporting training features of the device beyond its procedural assistance. Analyses controlling for completion time suggest that performance gain and skill acquisition were primarily driven by the device, rather than slower pacing alone **(Supplementary Text S5)**. Notably, the Dunning–Kruger effect was evident among Protocol 1 participants, who reported strong perceived skill gains despite no significant improvements in objective performance during the paper-assisted pooling on Day 1. This discordance highlights the importance of objective analytics and real-time quantitative feedback to foster true skill acquisition and prevent false confidence^47^ that can cause diagnostic errors^48^. Finally, unlike low-cost liquid handlers, the device recorded the actual weight of each pooled sample during pooling, enabling detection of *Invalid* pools that could otherwise lead to diagnostic errors.

The device validation study **(Fig. 8)** using 42 remnant clinical STH samples demonstrated the utility of the device for clinical sample pooling in disease surveillance or screening in RLS. Device-assisted 5-sample pooling yielded 100% positive/negative agreement for pooled qPCR in this sample set, suggesting the potential to transform STH-surveillance economics^34^ by driving down per-sample analysis costs while yielding diagnostic performance superior to the gold-standard Kato-Katz technique^16^. Current STH analyses are performed by manual DNA extraction followed by qPCR assays^16,34,46^; therefore pooling of raw samples upstream of DNA extraction would reduce the cost of analysis (e.g., from $11 to <$3) and increase the throughput of analysis^34^. By facilitating easier adoption of pooling in RLS, device-assisted high-performance surveillance would facilitate timely decisions on whether to cease mass drug administration where STH prevalence is sufficiently low.

Beyond STH, the same instructional pooling paradigm could be adapted for universal screening: by pairing with POC molecular diagnostics (e.g., Cepheid GeneXpert^31,32,33^), the device provides a pathway to increase testing throughput for bloodborne pathogens (e.g., HIV^28,29^, HCV^30,31^), respiratory diseases (e.g., TB^32^, viral infections^33^), and other infections in RLS. The device could also be invaluable during testing surges, so that even minimally trained personnel could be enlisted to help meet increased testing demands. This work exemplifies how new technologies, with affordability and usability, can extend healthcare capacity while employing a newly trained workers for healthcare services, rather than replacing them^49,50,51^.

This work highlights the potential of this class of devices for use in RLS, where automated instruments are often impractical for pre-analytical sample processing—tasks that humans can perform more efficiently with appropriate guidance^52^. Diagnostic workflows involving clinical specimens, such as stool-homogenization (STH)^34^, blood/plasma-separation (HIV/HCV)^53^, and sputum-evaluation (TB)^54^, frequently involve complex manual steps, precise volume-transfer skills, and visual judgement. These activities are challenging for low-cost automated systems due to pipetting failures, clogging, and inability to perform visual judgments^55,56,57^. By assisting and training laboratory-inexperienced personnel to successfully perform complex tasks (e.g., pooling), this class of devices can facilitate task-shifting in settings where individuals often perform multiple roles. This integration of technology with human capabilities supports more sustainable and scalable delivery of diagnostic services at the point of need.

We acknowledge several limitations of this work. First, the user-study cohort of 48 participants was skewed toward young adults. Training protocols spanned two nonconsecutive days, so longer-term skill retention remains to be studied. Artificial samples were prepared to approximate, but may not have fully replicated, the physical properties of actual clinical specimens. Printed instructions were chosen as the control because paper-based instructions remain the primary training resource in many resource-limited laboratories; however, richer comparators (e.g., a one-time instructional video) could better mirror real-world onboarding and should be evaluated in future work. Furthermore, Caltech policies prohibit untrained people to handle biohazardous materials, such as clinical samples, as they can contain unknown entities; for example, these samples came from a region where poliovirus is a concern and are known to be infected with STH pathogens. Therefore, the device validation studies with STH were performed by an engineer who underwent proper biosafety training. This study should be followed by multi-location field trials with RLS-relevant personnel and conditions.

This class of train-and-assist biomedical devices sits at the intersection of two domains: labor and human health. Beyond pooling, broader development and adoption of devices in this class could help mitigate both the labor crisis and healthcare inequities. Importantly, as new technologies disrupt labor markets, there remains a critical time window to implement scalable, pragmatic retraining solutions before inequalities widen further. We hope that this publication, the publicly available data, and open-source designs accompanying this work will stimulate development of innovative devices in this class to strengthen workforce and expand healthcare access.

## Materials and Methods

In the **Supplementary Methods**, we provide a comprehensive list of materials for the fabrication of the eight-module instructional pooling device and its disposable trays, detailing polymer filaments, sensor packages, micro-electronics, and vacuum forming. We then discuss descriptions of device fabrication for each module and its polypropylene device-protective trays, as well as the dual-microcontroller architecture that coordinates display, barcode scanning, object detection, precision weighing, and Bluetooth data export. We then outline the methods for expected handling movement and the calibration of the strain-gauge microbalance of the device. We present the design of the user study with underlying hypotheses and the details for participant recruitment, eligibility, withdrawal, compensation, and demographic statistics. We describe the details for preparing the user study: the preparation of customized data logger, sample-collection tubes, and artificial samples. Experimental settings, data collection methods, and detailed procedures for pooling exercises in the user study are also described. Details for data analysis of the user study data are described—paired statistical tests, linear mixed-effects modeling, and effect-size estimation, which were used to analyze the assistive and training effects on accuracy, precision, error counts, and task duration. Finally, we describe the methods for the device-validation study that was conducted for a 5-sample pooling validation of clinical STH stool samples on the device, detailing clinical sample preparation, pooling procedure, DNA extraction, qPCR conditions, and positive/negative percent agreement calculations. Full details for Materials and Methods are described in **Supplementary Methods S1**─**S22**. Inclusion & ethics statements are available in **Supplementary Text S8**. The text of the manuscript was proofread and revised by using OpenAI GPT-4o and o3 models and the Claud Sonnet 3.7 model.

## Data, Materials, and Software Availability

The datasets generated and analyzed during the current study, including all device designs and related codes, are available at CaltechDATA: https://doi.org/10.22002/wnh8n-mh110. Four videos are available at the link. All statistical analysis results are available in the Supporting Information Tables.

## Author Contributions

R.F.I. and M.L. initiated the concept and designed all studies; R.F.I. supervised the overall work; M.L. led the experiments, collected the data, and analyzed for the device design, system integration, device characterization, the user study, and clinical STH pooling on the device at California Institute of Technology; X.P.P. led the experiments, collected the data, and analyzed for overall clinical STH sample extraction and qPCR analysis at California Institute of Technology; N.P. and S.A.W. supervised experiments, data acquisition and analysis at Smith College for the clinical samples collected from Bangladesh local areas, the design and validation of the clinical STH sample homogenization, extraction, and pooling protocols, and qPCR assay characterization/analysis at Smith College; M.R. oversaw the clinical sample acquisition from Bangladesh local areas, through designing, planning, and managing field implementation that includes protocol development, IRB approval, training for sample/data collectors, sample quality assurance and tracking, sample preservation, and facilitation of sample shipment. S.H,J., contributed to device & tray design and fabrication; R.F.I., M.L., N.S., A.V.W., and C.F.C. contributed to the design of the user study; M.L., R.G., and C.F.C. contributed to the analysis of the user study; M.L. prepared all figures, tables, notes, and videos with input from R.F.I., S.H.J., X.P.P., A.V.W., and N.S.; M.L. wrote the original draft of the paper; M.L., N.S., and A.V.W. edited. All authors contributed to the data analysis and provided feedback on the manuscript. Detailed author contribution statements are available in **Supplementary Text S9**.

## Acknowledgments

We thank A. E. Romano (Caltech), M. Cooper (Caltech), R. Lazarovits (Caltech), O. Pradhan (Caltech), N. J. Wu-Woods (Caltech), R. Akana (Caltech), M. K. Porter (Caltech), M. K. Kim (Caltech), S. Haynes (Smith College), and A. M. Gonzalez (Smith College) for helping this work. This work was funded in part by an Innovation Seed Grant from the Merkin Institute for Translational Research (Caltech) and the Jacobs Institute for Molecular Engineering for Medicine (Caltech). Detailed statements are available in **Supplementary Text S10**.

