## Supplementary Information for "A train-and-assist device that upskills novices to strengthen the workforce and expand diagnostic access"

A class of train-and-assist biomedical devices
to strengthen the workforce and expand diagnostic access

Minkyo Lee^a^, Xinyue (Penny) Pei^b^, Si Hyung Jin^b^, Natasha Shelby^b^, Rani Gera^c^, Alexander Viloria Winnett^b^, Colin F. Camerer^c^, Mahbubur Rahman^d^, Nils Pilotte^e^, Steven A. Williams^f,g^, and Rustem F. Ismagilov^a,b,h,*^

^a^ Andrew and Peggy Cherng Department of Medical Engineering, Division of Engineering and Applied Science, California Institute of Technology, Pasadena, CA, United States of America

^b^ Division of Chemistry and Chemical Engineering, California Institute of Technology, Pasadena, CA, United States of America

^c^ Division of the Humanities and Social Sciences, California Institute of Technology, Pasadena, CA, United States of America

^d^ Environmental Health and WASH, Health Systems and Population Studies Division, International Centre for Diarrhoeal Disease Research, Bangladesh (icddr,b), Dhaka, Bangladesh

^e^ Department of Biological Sciences, Quinnipiac University, Hamden, CT, United States of America

^f^ Department of Biological Sciences, Smith College, Northampton, MA, United States of America

^g^ Molecular and Cellular Biology Program, University of Massachusetts, Amherst, MA, United States of America

^h^ Division of Biology and Biological Engineering, California Institute of Technology, Pasadena, CA, United States of America

**Contents**

**Supplementary Note S1 |** Step-by-step instructions through language-agnostic images provided by the instructional pooling device.

**Supplementary Note S2 |** Classification and management of correctable/uncorrectable errors on the instructional pooling device.

**Supplementary Note S3 |** Written paper instructions provided to participants at the beginning of pooling.

**Supplementary Note S4 |** Classification and examples of uncorrected handling errors in the user study.

**Supplementary Note S5 |** Effect of completion time versus device intervention on volume-transfer accuracy and handling errors.

**Supplementary Note S6 |** Establishing criteria for high-quality sample pools to assess high-performance volume transfer in pooling procedures.

**Supplementary Note S7 |** Quick information sheets provided to participants before starting pooling on Day 1.

**Supplementary Note S8 |** Inclusion & ethics statements.

**Supplementary Note S9 |** Detailed author contribution statements and ORCiDs.

**Supplementary Note S10 |** Detailed acknowledgement statements.

**Supplementary Fig. S1 |** Detailed circuit schematics of Display and Waste Modules of the instructional pooling device.

**Supplementary Fig. S2 |** Detailed circuit schematics of Pipette, Tube, Multibarcode, Cap, Barcode, and Scale Modules of the instructional pooling device.

**Supplementary Fig. S3 |** Detailed physical design of instructional pooling device for the detection of various objects.

**Supplementary Fig. S4 |** Laboratory materials and design components to support the instructional pooling device.

**Supplementary Fig. S5 |** Average accuracy and inter-variability of weight measurement (n=20 for each weight) of the device for known mass (1 mg to 50 g).

**Supplementary Fig. S6 |** A CONSORT flow diagram shows participant recruitment, eligibility, enrollment, and assignment to the study protocols.

**Supplementary Fig. S7 |** Determination of weight for 300 uL of artificial samples.

**Supplementary Fig. S8 |** Self-reported, post-pooling surveys on usability, training experience and effectiveness.

**Supplementary Fig. S9 |** Raw PSSUQ score data for 19 questionnaire items.

**Supplementary Fig. S10 |** Completion time in each pooling round using artificial respiratory and stool samples for Protocol 1 and 2 groups.

**Supplementary Fig. S11 |** Uncorrected handling errors of participants during pooling exercises across two training days.

**Supplementary Fig. S12 |** Assistive effects of instructional pooling device on reducing both minor and severe handling errors in both Protocol 1 and 2 groups for both sample types.

**Supplementary Fig. S13 |** Assessment of handling error recurrence in the same categories before (session #1; BASE) and after (session #8; AFTER_2_) the use of instructional pooling device.

**Supplementary Fig. S14 |** Impact of time gaps between experimental Day 1 and Day 2 on participants' handling performance metrics (volume-transfer accuracy and the number of all uncorrected handling errors) comparing the final pooling round on Day 1 (i.e., Round 16th) and initial round on Day 2 (i.e., Round 17th).

**Supplementary Fig. S15 |** Determination of Acc_pool_ and CV_pool_ thresholds through Monte Carlo simulations.

**Supplementary Fig. S16 |** High-quality pools produced through high-performance volume-transfer, defined as pools with volume-transfer accuracy (Acc_pool_) ≥80% and precision ≤25% CV_pool_.

**Supplementary Fig. S17 |** High-quality pools through high-performance sample pooling (Acc_pool_≥80%, CV_pool_≤25%, *err*_all_=0) from Protocol 1 participants.

**Supplementary Fig. S18 |** High-quality pools through high-performance sample pooling (Acc_pool_≥80%, CV_pool_≤25%, *err*_all_=0) from Protocol 2 participants using artificial respiratory samples.

**Supplementary Fig. S19 |** High-quality pools through high-performance sample pooling (Acc_pool_≥80%, CV_pool_≤25%, *err*_all_=0) from Protocol 2 participants using artificial stool samples.

**Supplementary Fig. S20 |** The number of *Incorrect* and *Invalid* pools across pooling sessions.

**Supplementary Fig. S21 |** *Invalid* pools produced during paper-assisted pooling.

**Supplementary Fig. S22 |** Circuit schematics of customized data logger used in paper-assisted pooling.

**Supplementary Fig. S23 |** Handling error check sheet used to assess user’s handling errors during the pooling.

**Supplementary Fig. S24 |** Sample processing, extraction control, and limit of blank for 5-sample pooled qPCR testing in the device validation study.

**Supplementary Table S1 |** Summary demographic and prior training data for participants in the usability studies.

**Supplementary Table S2 |** Demographic data for participants of Protocol 1 Group using artificial respiratory samples.

**Supplementary Table S3 |** Demographic data for participants of Protocol 1 Group using artificial stool samples. HS, high school; AP, advanced placement.

**Supplementary Table S4 |** Demographic data for participants of Protocol 2 Group using artificial respiratory samples.

**Supplementary Table S5 |** Demographic data for participants of Protocol 2 Group using artificial stool samples.

**Supplementary Table S6 |** A modified post-study system usefulness questionnaire (PSSUQ) to evaluate the usability of written paper instructions.

**Supplementary Table S7 |** A modified post-study system usefulness questionnaire (PSSUQ) to evaluate the usability of instructional pooling device.

**Supplementary Table S8 |** Statistical analysis results for PSSUQ surveys.

**Supplementary Table S9 |** Statistical analysis results for KTEQ and PTEQ surveys.

**Supplementary Table S10 |** Severity level of user handling errors.

**Supplementary Table S11 |** Statistical analysis results for uncorrected handling errors (∑*err*_all_) across pooling sessions for Protocol 1 group.

**Supplementary Table S12 |** Statistical analysis results for average volume-transfer accuracy (Avg. Acc_pool_) across pooling sessions for Protocol 1 group.

**Supplementary Table S13 |** Linear mixed model analyses (with continuous pooling round variable) for completion time in the Protocol 1 group.

**Supplementary Table S14 |** Linear mixed model analyses (with continuous pooling round variable) for volume-transfer accuracy (Acc_pool_), controlling for completion time in the Protocol 1 group.

**Supplementary Table S15 |** Linear mixed model analyses (with continuous pooling round variable) for all handling (*err*_all_), minor (*err*_minor_), and severe errors (*err*_severe_), controlling for completion time in the Protocol 1 group.

**Supplementary Table S16 |** Linear mixed model analyses (with dummy-coded pooling sessions and group effects) for all uncorrected handling errors (∑*err*_all_), controlling for completion time in the Protocol 1 group.

**Supplementary Table S17 |** Linear mixed model analyses (with dummy-coded pooling sessions and group effects) for average volume-transfer accuracy (Avg. Acc_pool_), controlling for completion time in the Protocol 1 group.

**Supplementary Table S18 |** Categories of user handling errors: De-contamination, Sample-Transfer, Contamination-Prevention, and Documentation.

**Supplementary Table S19 |** Statistical analysis results for minor handling errors (∑*err*_minor_) across pooling sessions for Protocol 1 group.

**Supplementary Table S20 |** Statistical analysis results for severe handling errors (∑*err*_severe_) across pooling sessions for Protocol 1 group.

**Supplementary Table S21 |** Statistical analysis results for uncorrected handling errors (∑*err*_all_) across pooling sessions for Protocol 2 group.

**Supplementary Table S22 |** Statistical analysis results for average volume-transfer accuracy (Avg. Acc_pool_) across pooling sessions for Protocol 2 group.

**Supplementary Table S23 |** Statistical analysis results for minor handling errors (∑*err*_minor_) across pooling sessions for Protocol 2 group.

**Supplementary Table S24 |** Statistical analysis results for severe handling errors (∑*err*_severe_) across pooling sessions for Protocol 2 group.

**Supplementary Table S25 |** Linear mixed model analyses (with continuous pooling round variable) for completion time in the Protocol 2 group.

**Supplementary Table S26 |** Linear mixed model analyses (with continuous pooling round variable) for volume-transfer accuracy (Acc_pool_), controlling for completion time in the Protocol 2 group.

**Supplementary Table S27 |** Linear mixed model analyses (with continuous pooling round variable) for all handling (*err*_all_), minor (*err*_minor_), and severe errors (*err*_severe_), controlling for completion time in the Protocol 2 group.

**Supplementary Table S28 |** Linear mixed model analyses (with dummy-coded pooling sessions and group effects) for all uncorrected handling errors (∑*err*_all_), controlling for completion time in the Protocol 2 group.

**Supplementary Table S29 |** Linear mixed model analyses (with dummy-coded pooling sessions and group effects) for average volume-transfer accuracy (Avg. Acc_pool_), controlling for completion time in the Protocol 2 group.

**Supplementary Table S30 |** Statistical analysis results for the number of high-quality pools across pooling sessions for Protocol 1 group.

**Supplementary Table S31 |** Statistical analysis results for the number of high-quality pools across pooling sessions for Protocol 2 group.

**Supplementary Table S32 |** Statistical analysis results for the number of Protocol 2 participants generating high-quality pools across pooling sessions.

**Supplementary Table S33 |** Statistical analysis results for the number of high-performed pooling rounds across pooling sessions for Protocol 1 group.

**Supplementary Table S34 |** Statistical analysis results for the number of high-performed pooling rounds across pooling sessions for Protocol 2 group.

**Supplementary Table S35 |** Statistical analysis results for the number of Protocol 2 participants achieving high-performance pooling across pooling sessions.

**Supplementary Table S36 |** Statistical analysis results for the rates of *Incorrect* pools between paper-assisted and device-assisted pooling among Protocol 2 participants (N=29; Respiratory: 12, Stool: 17).

**Supplementary Table S37 |** Statistical analysis results for the weight of transferred liquid across homogenized stool suspension groups.

**Supplementary Table S38 |** Bill of materials for the instructional sample-pooling device.

**Supplementary Method S1 |** Materials for device fabrication

**Supplementary Method S2 |** Fabrication of instructional pooling device and trays

**Supplementary Method S3 |** System architecture of instructional pooling device

**Supplementary Method S4 |** Expected handling movement on the instructional pooling device

**Supplementary Method S5 |** Characterization of strain gauge-based weight measurement system of instructional pooling device

**Supplementary Method S6 |** Design of user study

**Supplementary Method S7 |** Participants for the user study

**Supplementary Method S8 |** Preparation of customized data logger

**Supplementary Method S9 |** Preparation of sample-collection tubes

**Supplementary Method S10 |** Preparation of artificial samples

**Supplementary Method S11 |** Determination of mass for 300 µL of artificial samples

**Supplementary Method S12 |** Setting for pooling exercises (user study)

**Supplementary Method S13 |** Initial study phase: orientation and practice on Day 1

**Supplementary Method S14 |** Pooling exercises

**Supplementary Method S15 |** Data collection during pooling exercises

**Supplementary Method S16 |** Post-experimental procedures

**Supplementary Method S17 |** Data analysis in the user study

**Supplementary Method S18 |** Clinical STH samples and commercial stool samples

**Supplementary Method S19 |** 5-sample pooling and DNA extraction

**Supplementary Method S20 |** qPCR analysis

**Supplementary Method S21 |** Data analysis in the device validation study

**Supplementary Method S22 |** Determination of PPA and NPA

**Supplementary Movie S1 (separate file)** | Introduction of instructional sample-pooling device

**Supplementary Movie S2 (separate file)** | Demonstration of instructional sample-pooling device that provides language-agnostic step-by-step instructions.

**Supplementary Movie S3 (separate file)** | Demonstration of device instructions for correcting user handling errors.

**Supplementary Movie S4 (separate file)** | Demonstration of device instructions for terminating the pooling process in response to uncorrectable handling errors.

**Note S1. Step-by-step instructions using language-agnostic images provided by the instructional pooling device.** Language-agnostic images shown below were saved on microSD card of the device. In each pooling round, the device displays the saved images on the LCD screen step-by-step to guide the users through the 5-sample pooling.

| 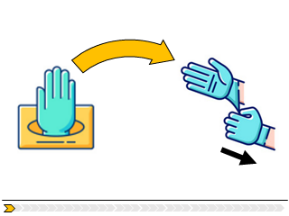 | 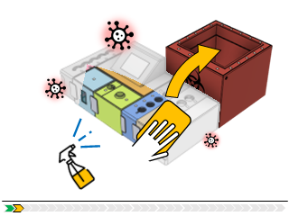 | 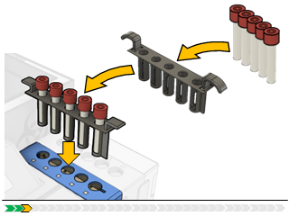 | 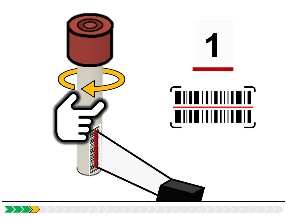 |
| --- | --- | --- | --- |
| **Step 1.** Wear gloves. | **Step 2.** Disinfect the workplace. | **Step 3.** Put the tube tray with sample tubes into Tube module. | **Step 4.** The device scans the barcodes of tubes #1 - #5. |
| 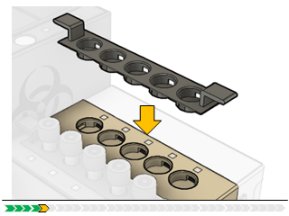 | 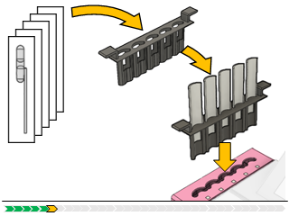 | 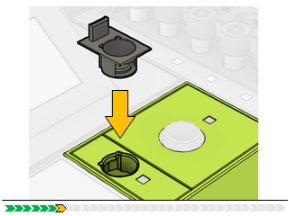 | 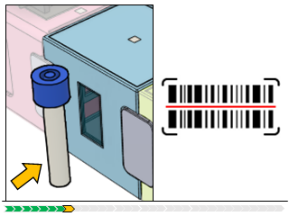 |
| **Step 5.** Put the cap tray into the Cap module. | **Step 6.** Put the pipette tray into the Pipette module. | **Step 7.** Put the pooling cap tray into the Pooling module. | **Step 8.** Scan the barcode of the pooling tube through the Single barcode module. |
| 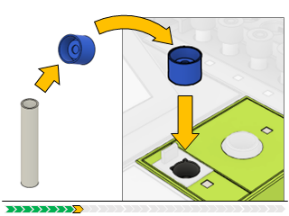 | 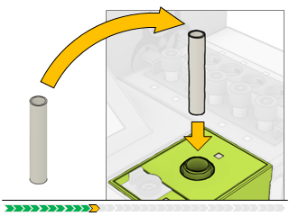 | 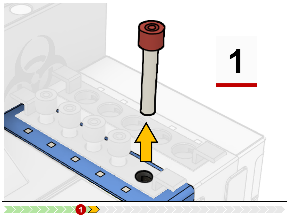 | 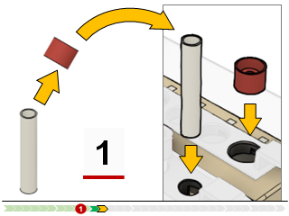 |
| **Step 9.** Open the pooling tube and put the cap into the detection well. | **Step 10.** Put the tube into the pooling tube holder. | **Step 11***: Pick up one individual tube (#1-#5) of sample to be pooled | **Step 12*.** Open the tube and place it and its cap into their slots. |
| 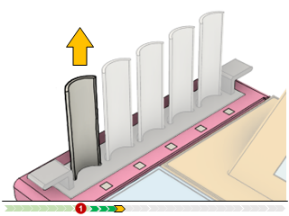 | 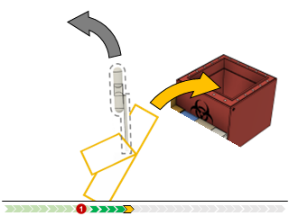 | 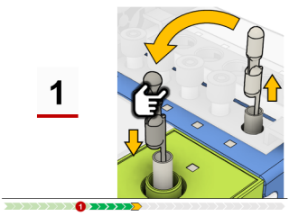 | 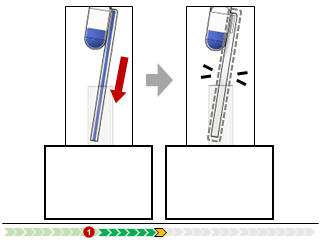 |
| **Step 13*.** Pick up a pipette. | **Step 14*.** Peel off the plastic bag of the wrapped pipette. | **Step 15*.** Aspirate the liquid sample from the sample tube. | **Step 16*.** Dispense the liquid sample into the pooling tube. |
| 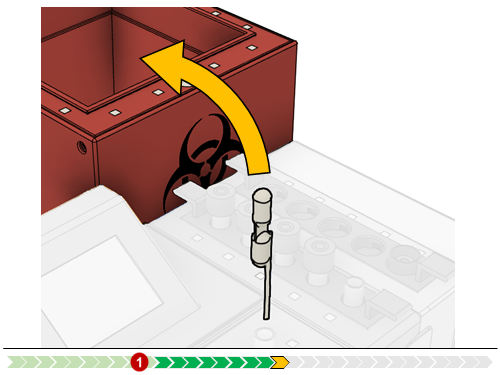 | 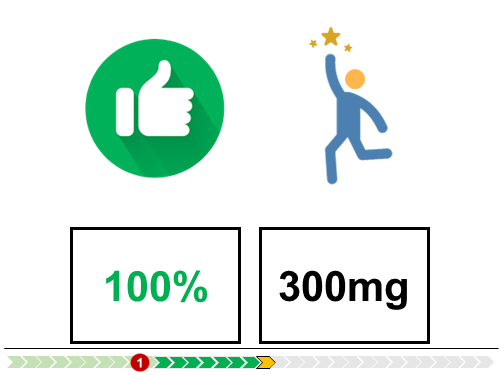 | 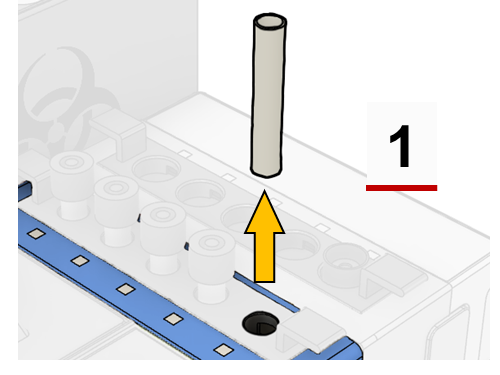 | 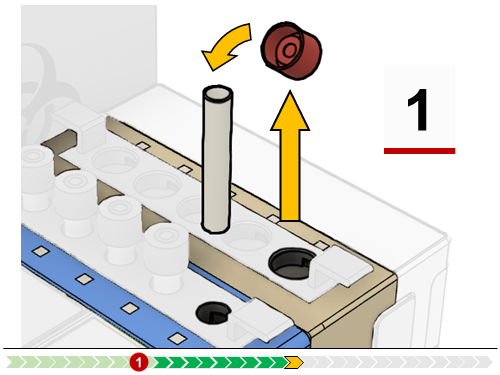 |
| **Step 17*.** Dispose of the used pipette. | **Step 18*.** The device checks the pooled volume of the sample. | **Step 19*.** Pick up the opened collection tube for closing | **Step 20*.** Pick up the cap and close the collection tube. |
| 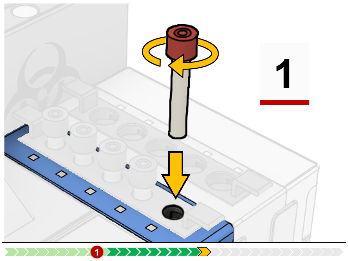 | 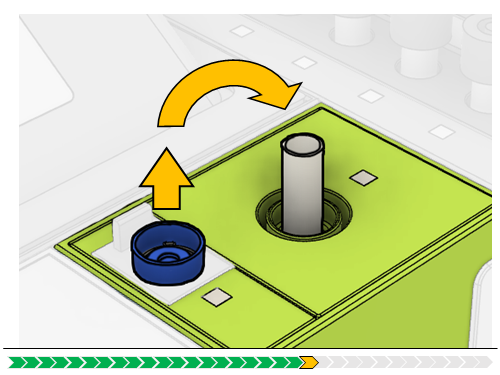 | 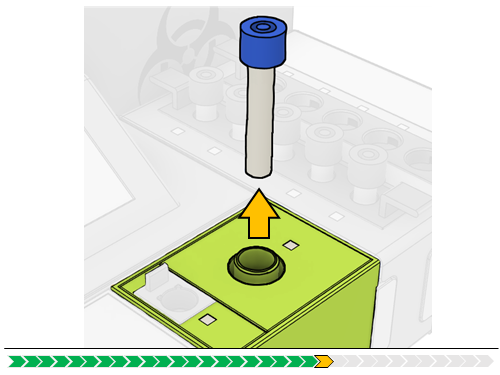 | 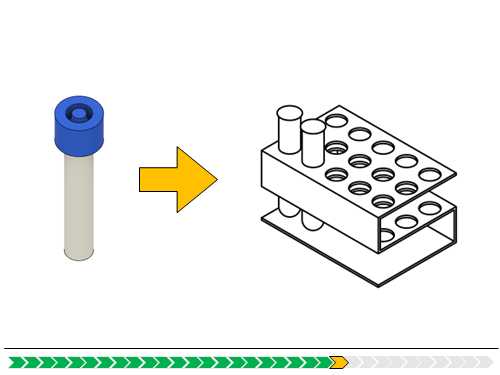 |
| **Step 21*.** Close the tube tightly and place it back to the module. | **Step 22.** Close the pooling tube. | **Step 23.** Remove the pooling tube from the device. | **Step 24.** Store the pooling tube to run the diagnostic testing. |
| 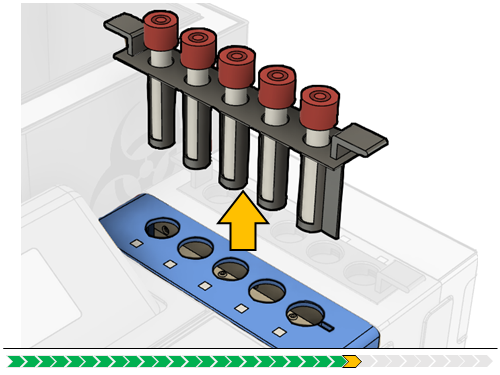 | 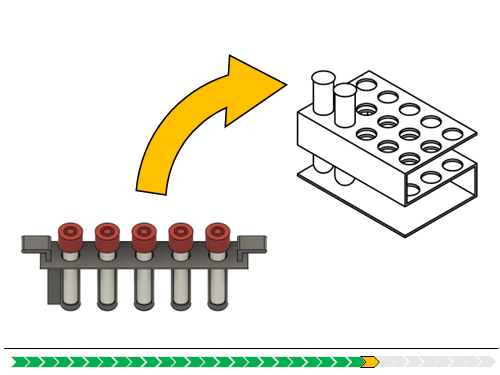 | 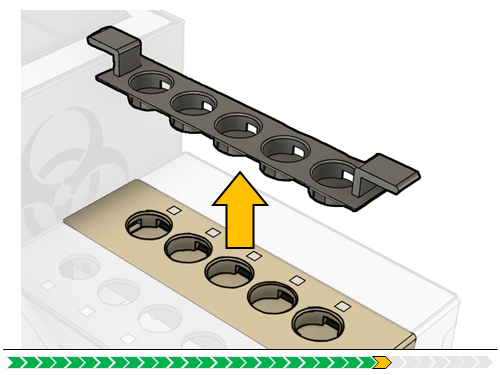 | 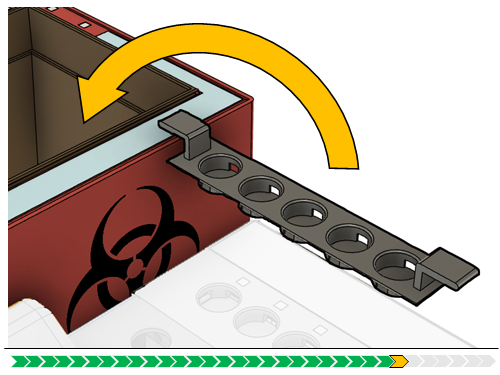 |
| **Step 25.** Remove the tube tray from the device. | **Step 26.** Store the tube tray for confirmatory testing. | **Step 27.** Remove the cap tray from the device. | **Step 28.** Dispose of the cap tray into the waste bin. |
| 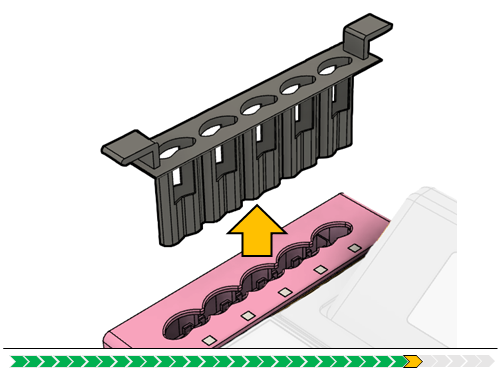 | 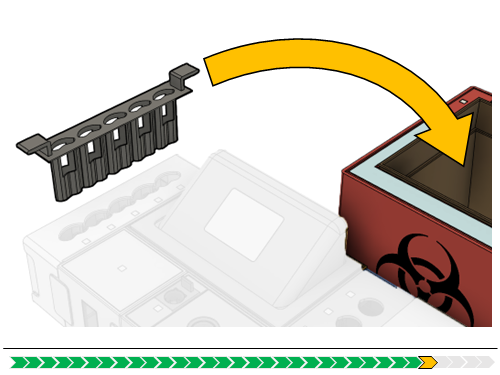 | 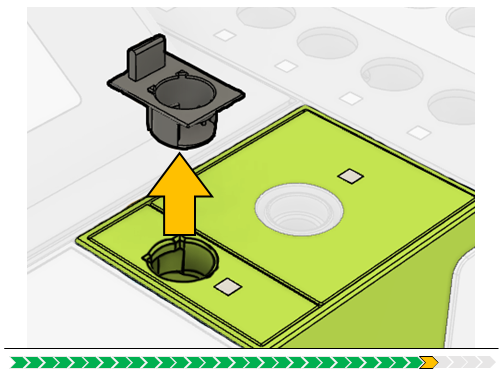 | 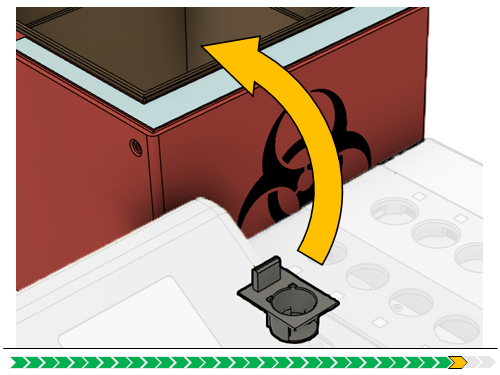 |
| **Step 29.** Remove the pipette tray from the device. | **Step 30.** Dispose of the pipette tray into the waste bin. | **Step 31.** Remove the pooling cap tray from the device. | **Step 32.** Dispose of the pooling cap tray into the waste bin. |
| 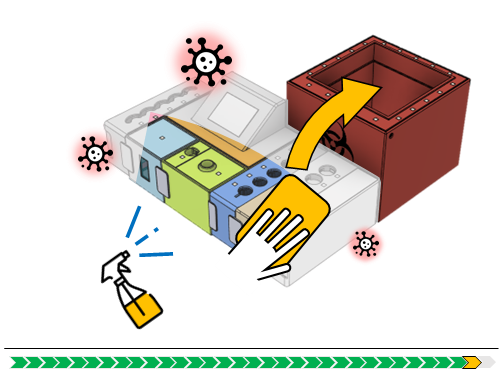 | 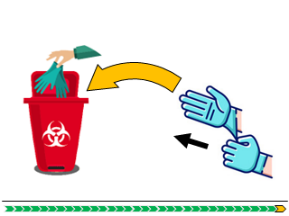 | 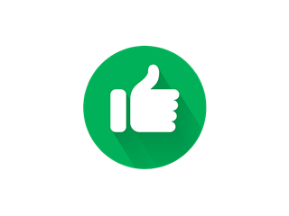 |  |
| **Step 33.** Disinfect the workplace. | **Step 34.** Take off the gloves and dispose of them in the waste bin. | **Step 35.** Pooling has been done. |  |

*Steps 11–21: In each pooling round, these steps were repeated until all five individual samples were pooled to the pooling tube.

**Note S2. Classification and management of correctable/uncorrectable errors on the instructional pooling device.** Handling errors on the instructional pooling device are classified into correctable and uncorrectable errors. Correctable errors are those that device users can remedy before compromising the biosafety and sample integrity, while uncorrectable errors are those that inherently compromise sample traceability, integrity, and reliability and cannot be remedied once they occur. The device incorporates four key functionalities to manage these errors: sensor-guided interactive instructions **(Fig. 2a,b)**, scenario-adaptive interrupt-driven real-time monitoring **(Fig. 2c,d)**, weight-based volume transfer monitoring **(Fig. 2e,f,g)**, and barcode-based sample tacking **(Fig. 2h)**.

Each functionality contributes to error management in distinct ways. Sensor-guided instructions ensure step-by-step completion of actions through visual guidance and monitoring. The scenario-adaptive monitoring system detects potential errors in real-time using interrupt service routines. Weight-based monitoring tracks volume transfers for accuracy, while barcode tracking ensures correct tube identification and usage throughout the process.

*1. Correctable errors on the device*

A. Errors detectable and preventable through sensor-guided instructions and interrupt-driven monitoring system

The sensor-guided interactive instructions are designed to ensure step-by-step completion of user actions, preventing users from skipping or forgetting steps. During each step, the device maintains vigilance by displaying blinking pink LEDs and monitoring object-detection slots for potential human errors **(Fig. 2b)**. When a correctable error is detected, the device immediately alerts users with a warning sign graphics and beeping sound, followed by another language-agnostic graphics for correction guidance **(Fig. 2b,d and Supplementary Movie S3)**. Users can correct their mistake by following the correction guidance from the device. The detection capability is enabled by the scenario-adaptive interrupt-driven monitoring system using interrupt service routines **(Fig. 2c)**. The following examples illustrate correctable errors that the device can detect and prevent through its interactive instructions and monitoring system.

(a) *Skipping barcode scanning*: Users may forget the barcode scanning of tubes. The sensor-guided instructions can help users complete barcode scanning steps. At STEP 4, the device displays the action graphics and waits until users scan each individual sample tube. Similarly, at STEP 8, the device prompts users for pooling tube scanning before proceeding to tube opening and cap placement. These features are designed to play a role in preventing the following handling errors in the user study **(Supplementary Fig. S11)**: ‘Did not scan the pooling tube,’ ‘Did not scan the new sample tube #1,’ ‘Did not scan the new sample tube #2,’ ‘Did not scan the new sample tube #3,’ ‘Did not scan the new sample tube #4,’ and ‘Did not scan the new sample tube #5.’

(b) *Forgetting the tube opening*: Users may forget the tube opening during the pooling process. The sensor-guided instructions can help users complete tube opening steps successfully. At STEP 9, the device waits for pooling tube opening and cap placement in the empty cap slot of Scale Module. Similarly, at STEP 12, the system guides users through individual sample tube opening with cap placement in the empty slot of Cap Module. These features are designed to play a role in preventing the following handling errors in the user study **(Supplementary Fig. S11)**: ‘Did not open the pooling tube until dispensing sample into the tube,’ and ‘Did not open the sample tube and attempted to aspire sample from it.’

(c) *Using used pipettes*: Users may attempt to use used pipettes. The device incorporates multiple procedural steps to prevent the use of used pipettes through sensor-guided interactive instructions: monitoring placement of unpacked dual-bulb pipettes in Pipette Module (STEP 6), tracking pickup of unpacked pipettes paired with individual tubes (STEP 13), and monitoring disposal of used pipettes (STEP 18). These features are designed to play a role in preventing the handling error, ‘Did not use a new pipette,’ in the user study **(Supplementary Fig. S11)**.

(d) *Confusing which sample tube to use for volume transfer*: Users may be confused about the sequence of pooling from individual tubes. At STEP 11, users may attempt to transfer sample from a tube that has already been pooled (e.g., trying to use tube #1 again instead of moving on to tube #2). When this happens, the device alerts users with the warning sign and beeping sound and guides users to put back the incorrect tube and pick up the next tube in sequence. This feature is designed to play a role in preventing the following handling errors in the user study **(Supplementary Fig. S11)**: ‘Did not transfer sample from the sample tube #1 to the pooling tube,’ ‘Did not transfer sample from the sample tube #2 to the pooling tube,’ ‘Did not transfer sample from the sample tube #3 to the pooling tube,’ ‘Did not transfer sample from the sample tube #4 to the pooling tube,’ and ‘Did not transfer sample from the sample tube #5 to the pooling tube.’

(e) *Forgetting the disposal of used pipettes:* Users may forget the disposal of used pipettes after the volume transfer. At STEP 18, the device waits for the disposal of pipette after the volume transfer while blinking pink LEDs and displaying the action graphics. This feature is designed to play a role in preventing the handling error, ‘Did not dispose of the used pipette,’ in the user study **(Supplementary Fig. S11)**.

(f) *Forgetting the closing of individual sample or pooling tubes:* Users may forget the tube closing during the pooling. The sensor-guided instructions can help users complete tube closing steps successfully. At STEP 20, the device waits for sample tube closing and cap removal from the cap slot of Cap Module. Similarly, at STEP 22, the system guides users through pooling tube closing by removing the cap from the cap slot of Scale Module. These features are designed to play a role in preventing the following handling errors in the user study **(Supplementary Fig. S11)**: ‘Did not close the sample tube until cleaning workspace,’ ‘Did not close the pooling tube until the end of pooling process,’ and ‘Skipped closing the pooling tube and clean-up to pool from the next set.’

B. Errors detectable and preventable through barcode-based sample tracking system

Barcode-based sample tracking can also enable the device to guide users in correct tube identification. When incorrect barcodes are scanned, the device alerts with the warning sign and beeping sound, followed by language-agnostic graphics to guide proper scanning. The following example illustrates how the device's barcode tracking features are designed to help users avoid scanning errors **(Supplementary Movie S3)**.

(a) *Confusing which tube to scan*: Users may scan incorrect tubes during the pooling process. At STEPS 4 and 8, when users scan an incorrect tube (such as a previously used sample tube or wrong type of tube), the device displays graphics guiding users to scan the correct tube. For individual sample tubes at STEP 4, the device verifies that each scanned tube is unused and from the correct set. For pooling tubes at STEP 8, the device verifies that a new, unused pooling tube is scanned. These features are designed to play a role in preventing the following handling errors in the user study **(Supplementary Fig. S11)**: 'Did not use a new pooling tube,' ‘Did not identify a new set of samples to be pooled,’ and 'Scanned the wrong sample tube(s) from the wrong set(s).'

C. Errors preventable but not detectable

Through sensor-guided instructions and the layout optimized for the pooling process, the device is also designed to prevent a few correctable errors that are not detectable. The following examples illustrate correctable errors that are not detectable but preventable by the device.

(a) *Forgetting the cleaning of workspace*: Users may forget to clean the workspace either at the beginning or the end of the pooling round. Although the device cannot detect the human action of cleaning directly, the device waits for the cleaning for a few seconds, displays the graphics, and waits for users’ action that disposes of the wipes into Waste Module. If users do not dispose of the wipe, the process will not move to the next step. This feature is designed to play a role in preventing the following handling errors in the user study **(Supplementary Fig. S11)**: ‘Did not clean the workspace with new wipes at the beginning’ and ‘Did not clean the workspace with new wipes at the end.’

(b) *Forgetting the preparation of new pipettes*: Users may forget to prepare new pipettes at the beginning of pooling round. While the device cannot directly detect the human action of preparing new pipettes with the pipette tray, the device displays the graphics at STEP 6 guiding users to put new individually wrapped pipettes in the pipette tray and insert them into Pipette Module. If users do not prepare new pipettes in the device module, the process will not move to the next step. This feature is designed to play a role in preventing the handling error, ‘Did not prepare new pipettes before opening sample tubes,’ in the user study **(Supplementary Fig. S11)**.

(c) *Not removing used gloves*: Users may attempt to proceed without disposing of their used gloves. Although the device cannot directly detect the human action of wearing off gloves, it displays the graphics prompting glove disposal and waits for users to complete this step before moving to the next step. This feature is designed to play a role in preventing the handling error, ‘Did not remove used gloves,’ in the user study **(Supplementary Fig. S11)**.

D. Errors neither detectable nor preventable

One correctable error ─ not wearing new gloves at the beginning of pooling round ─ cannot be directly detected nor prevented by the device. This error is indirectly addressed through the device's guidance for removing used gloves at the end of each pooling round. When users follow these instructions to dispose of used gloves from the previous round, they are naturally guided to wear new gloves for the next round.

*2. Uncorrectable errors on the device*

A. Errors undetectable but preventable through sensor-guided instructions and interrupt-driven monitoring system

The device also incorporates features designed to detect or prevent certain uncorrectable errors. While many uncorrectable errors cannot be directly detected, they can be proactively reduced through sensor-guided instructions and interrupt-driven monitoring system. The following examples illustrate how the device's interactive instructions and monitoring system work to prevent uncorrectable errors.

(a) *Transfer of sample from the wrong sample tube(s)*: Users may become confused about which individual tube of sample to be pooled. Although the device can detect volume transfer activity at the pooling tube, it cannot directly detect which individual sample tube is being used for sample removal. Despite this limitation, the device incorporates multiple preventive measures during STEPS 11–21. The scenario-adaptive interrupt-driven system monitors all individual sample tubes in Tube Module as well as empty cap slots of Cap Module. When any action (e.g., removing incorrect tubes or placing caps into the empty slots) is detected, the device alerts users with the warning sign and beeping sound and provides correction guidance. These features are designed to play a role in preventing the following handling errors in the user study **(Supplementary Fig. S11)**: ‘Transferred sample from a sample tube(s) that had already been pooled’ and ‘Transferred sample from a sample tube(s) from the wrong set(s).’ However, the device cannot prevent transfers that occur outside its monitoring area.

(b) *Transfer of sample to a wrong tube(s)*: Users may become confused about which tube should receive the sample transfer and attempt to use incorrect tubes. The device incorporates multiple preventive measures against such incorrect transfers. At STEP 8, barcode scanning verification helps prevent transfers to incorrect pooling tubes (such as previously used pooling tubes). The interrupt-driven monitoring system also alerts users with the warning sign and beeping sound when users pick up incorrect sample tubes from Tube Module, helping prevent transfers between individual sample tubes. These features are designed to play a role in preventing the following handling errors in the user study **(Supplementary Fig. S11)**: ‘Transferred sample to the pooling tube of the wrong set’ and ‘Transferred sample from a sample tube to another sample tube.’ However, the device cannot prevent transfers that occur outside its monitoring area.

(c) *Closing tubes with wrong caps*: Users may become confused about which cap to use for closing tubes. While the device cannot directly detect the human action of closing, the interrupt-driven monitoring system monitors the cap placement in its cap slot of Cap Module. When an error (e.g., picking up a wrong cap) occurs, the system alerts users with the warning sign and beeping sound and provides correction guidance. These features are designed to play a role in preventing the following handling errors in the user study **(Supplementary Fig. S11)**: ‘Closed the sample tube with the pooling tube cap’ and ‘Closed the pooling tube with the sample tube cap.’

B. Errors detectable but not preventable

A few uncorrectable errors can be detected but not prevented by the device. For these errors, the device alerts users with the warning sign and beeping sound, then provides guidance to terminate the entire pooling process, to prevent further contamination risks. The following examples illustrate how the device’s weight-based volume transfer monitoring system detects these errors and guides process termination **(Supplementary Movie S4)**.

(a) *Transfer of sample from a pooling tube to a sample tube*: Users may incorrectly attempt to transfer sample from the pooling tube to an individual sample tube, reversing the intended transfer direction. The device detects the sample removal through weight measurement of the pooling tube. When the removal is detected, the device alerts users with the warning sign and beeping sound and guides users to close and dispose of all tubes to prevent further contamination risks. These features are designed to play a role in preventing the following handling errors in the real-world clinical settings: ‘Transferred sample from the pooling tube to a sample tube’ and ‘Removed sample from the pooling tube.’ Please note that the termination guidance was inactivated during the user study to allow the observation of complete handling performance of users throughout the entire process in each pooling round.

(b) *Multiple sample transfers from individual sample tubes*: Users may attempt multiple sample transfers using the same pipette from the same individual sample tube, particularly when concerned about insufficient volume transfer. The multiple transfer of sample between individual sample and pooling tubes can increase the risk of cross-contamination. While the device cannot directly detect the volume transfer activities (such as re-aspiration) from individual sample tubes, it can detect the abnormal weight of transferred sample in the pooling tube (e.g., 150% of the 300 μL target weight). This excess weight occurs when users perform multiple re-aspirations, filling both the pipette channel and bottom bulb of dual-bulb pipettes with sample, which then dispense together into the pooling tube. Upon detecting excess weight, the device alerts users with the warning sign and beeping, then guides users to close and dispose of all tubes to prevent further contamination risks. These features are designed to play a role in preventing the handling error, ‘Re-aspirated with a used pipette that had been dipped in the pooling tube,’ in the real-world clinical settings. However, the termination guidance was inactive during the user study to allow the observation of complete handling performance of users throughout the entire process in each pooling round.

C. Errors neither detectable nor preventable

Two uncorrectable errors cannot be detected nor prevented by the device: ‘Dispensing liquid sample outside of the pooling tube’ and ‘Disposal of tubes after closing.’ These errors are unlikely to occur during routine pooling procedures and can be prevented when users are trained having good handling practices.

**Note S3. Written paper instructions provided to participants at the beginning of pooling.** All participants were provided the paper instructions with action diagrams combining graphics and texts for 5-sample pooling, as shown below. The instructions were designed and provided for all participants to maximize understandability and usability, incorporating step-by-step guidance with action diagrams that combined graphics and texts^1,2^. The procedural flow closely mirrored that of the instructional device to ensure a fair comparison between the paper instructions and the device instructions, preventing potential bias that could arise from significant differences in workflow. While maintaining the structural similarity, the paper instructions intentionally used different graphics from those on the instructional device. This choice was made to ensure participants had no prior exposure to the device's language-agnostic graphics and to avoid another potential source of bias and maintain the authenticity of participants' first interaction with the device instructions. Following is the instructions provided to participants in the user study.

**********

**Written Instruction for 5-Sample Pooling**

Repeat the steps below until all individual samples are pooled.

**<SET UP>**

| STEP #1 |  | STEP #2 |
| --- | --- | --- |
| 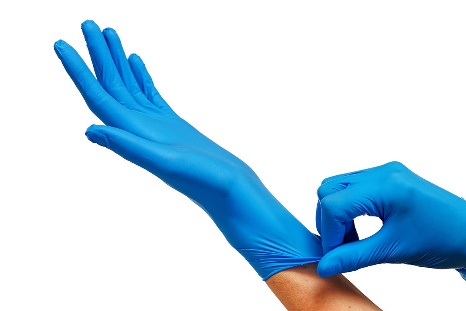 |  | 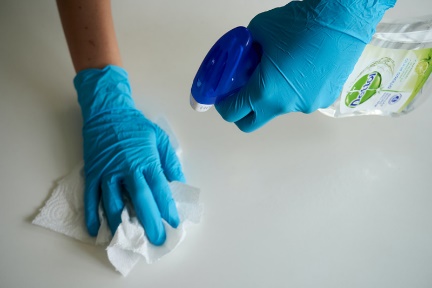 |
| Put on new gloves |  | Clean the workspace (or table) with the spray bottle and paper towels |

| STEP #3 |  | STEP #4 |  | STEP #5 |
| --- | --- | --- | --- | --- |
| 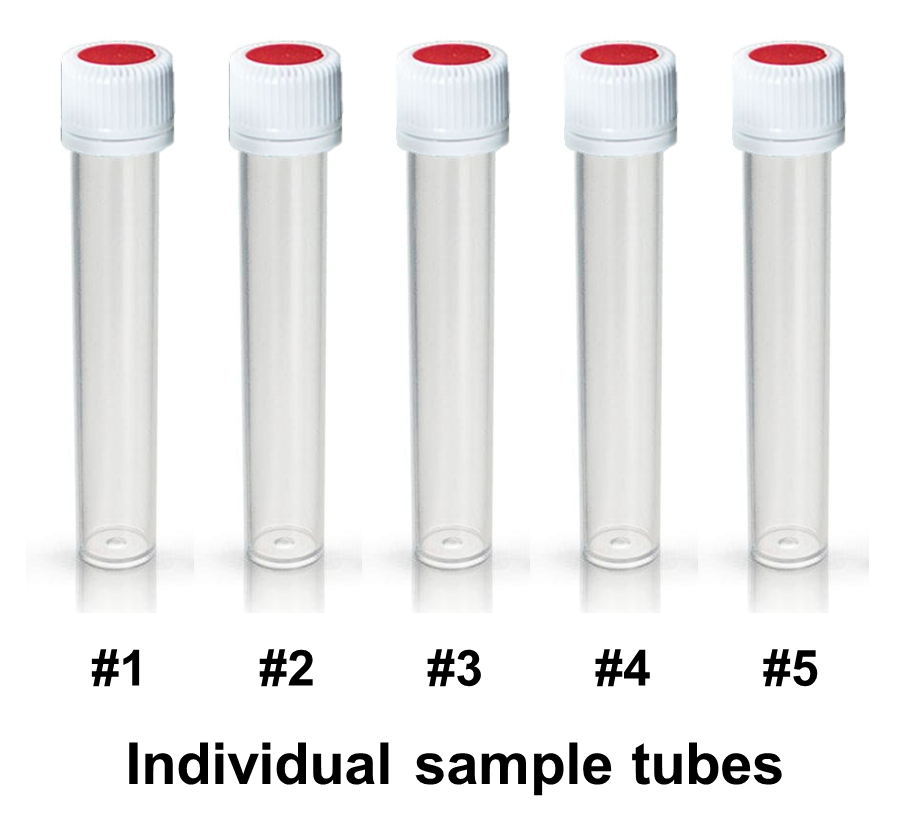 |  | 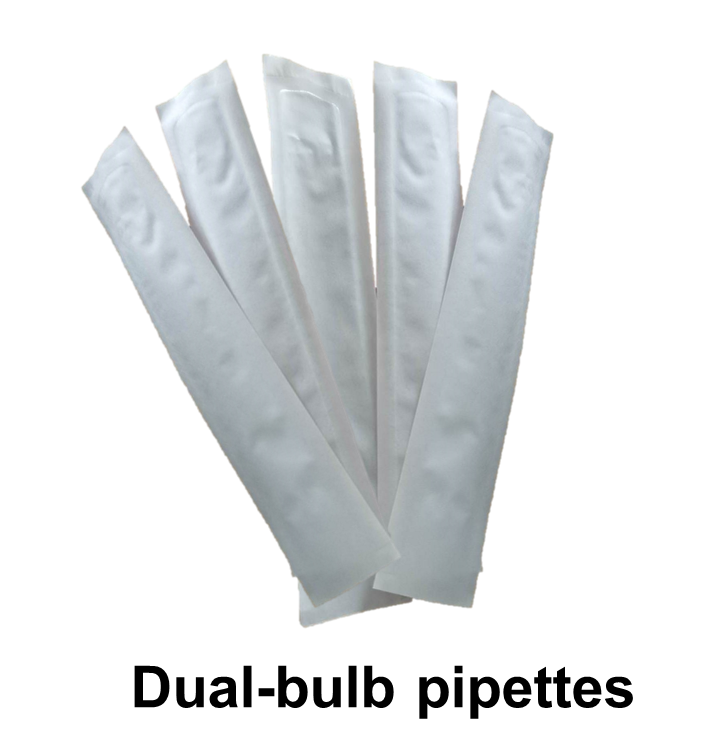 |  | 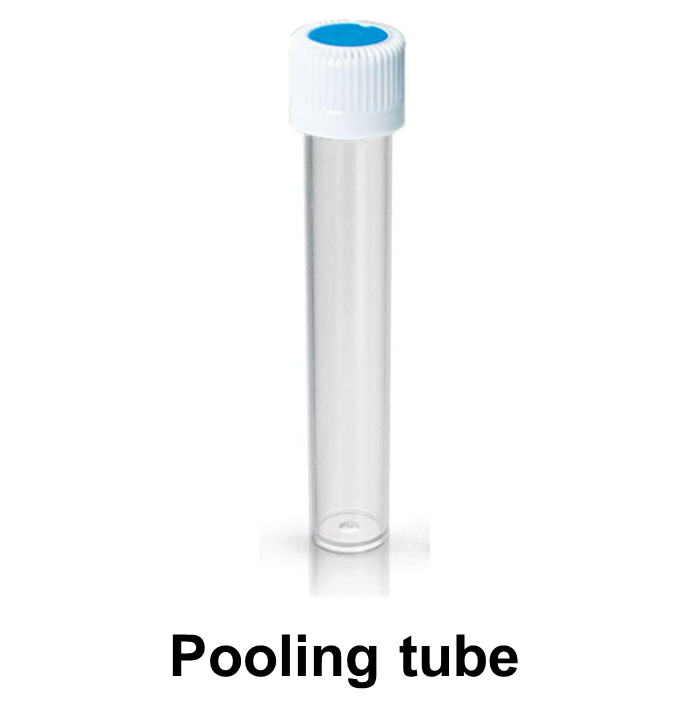 |
| Identify five un-pooled individual samples that will be pooled in this pooling cycle |  | Count out five new dual-bulb pipettes and place them in the clean zone |  | Remove one new, empty pooling tube, paired with individual samples, from the tube rack and place it in the pooling zone. |

| STEP #6 |  | STEP #7 |  | STEP #8 |
| --- | --- | --- | --- | --- |
| 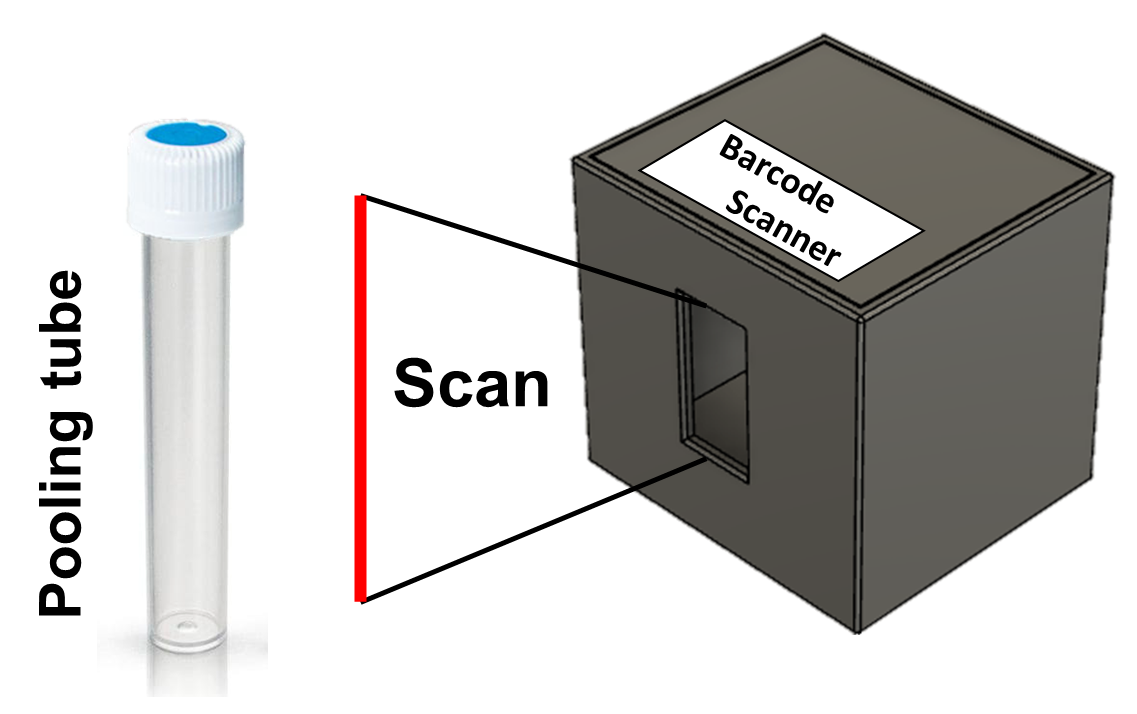 |  | 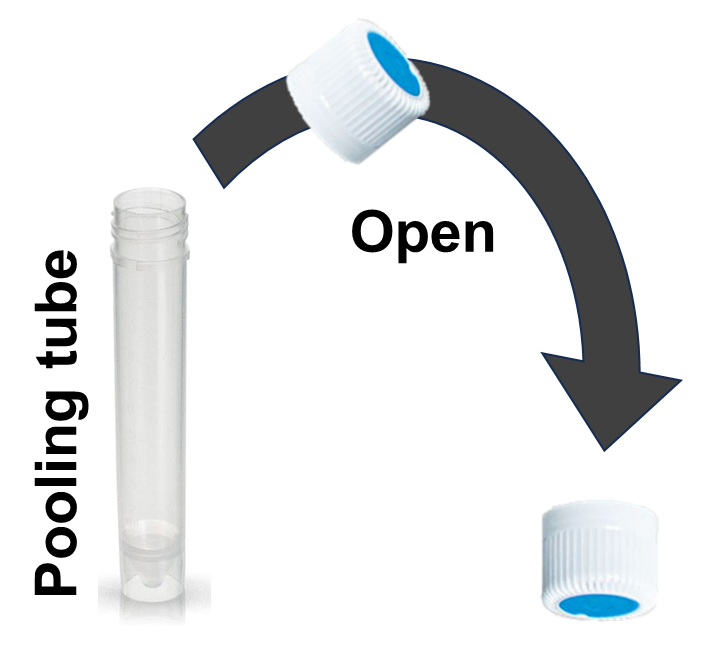 |  | 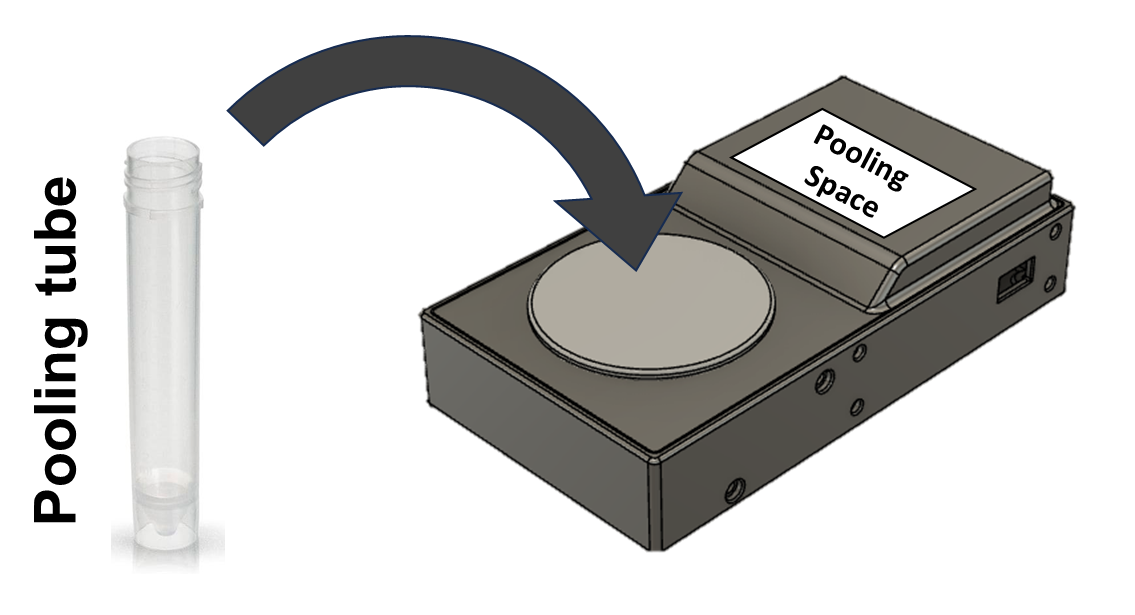 |
| Scan the pooling tube |  | Open the pooling tube and place the cap on the table, open side up |  | Place the tube on the pooling space |

**<POOLING>**

Repeat <POOLING> until all samples for a single pooling set are pooled.

| STEP #9 |  | STEP #10 |  | STEP #11 |
| --- | --- | --- | --- | --- |
| 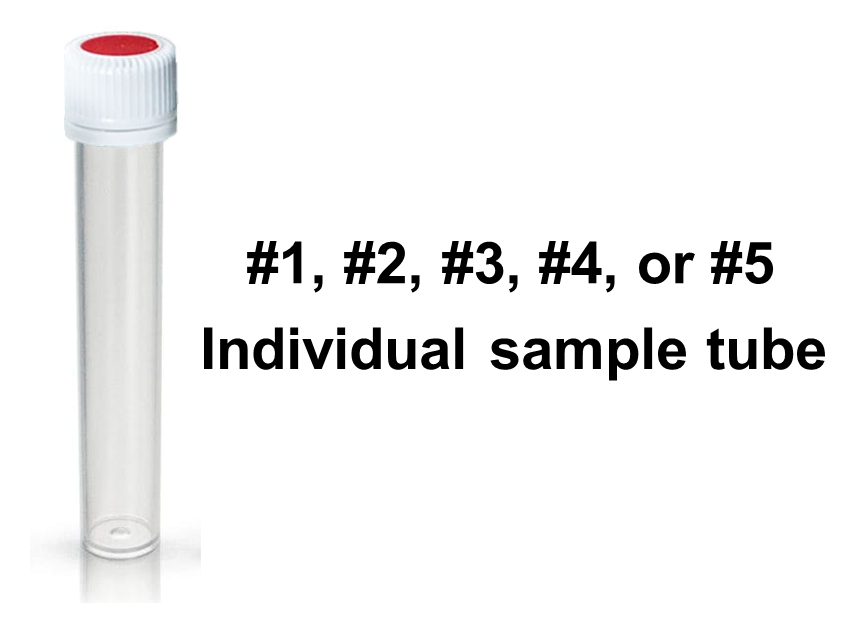 |  | 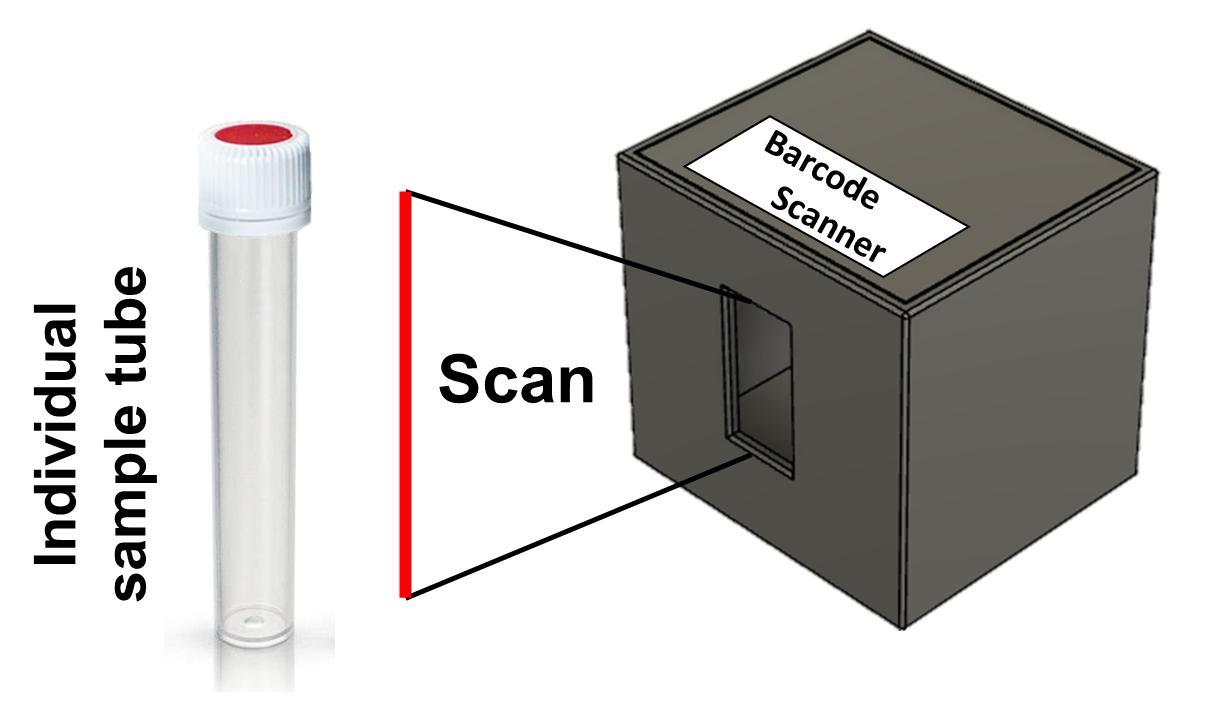 |  | 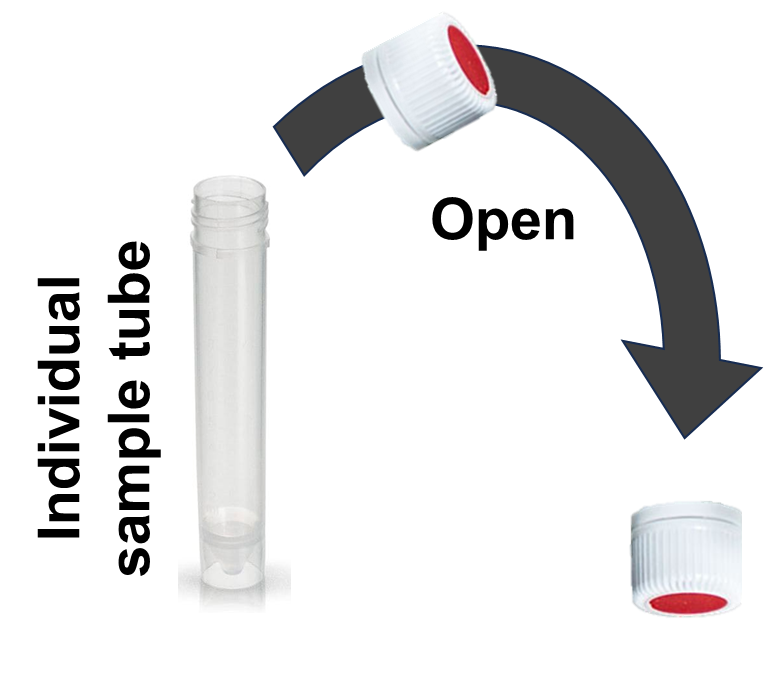 |
| Take the sample tube that you want to pool |  | Scan the sample tube |  | Open the sample tube and place the cap on the table, open side up |

| STEP #12 |  | STEP #13 |  | STEP #14 |
| --- | --- | --- | --- | --- |
| 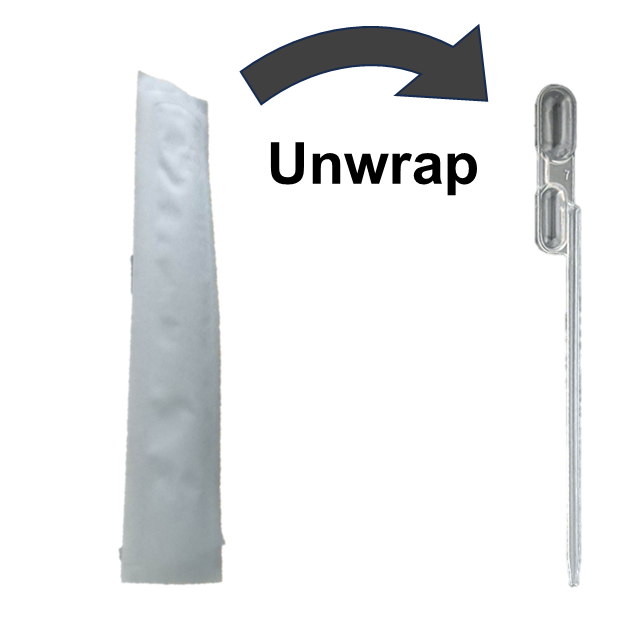 |  | 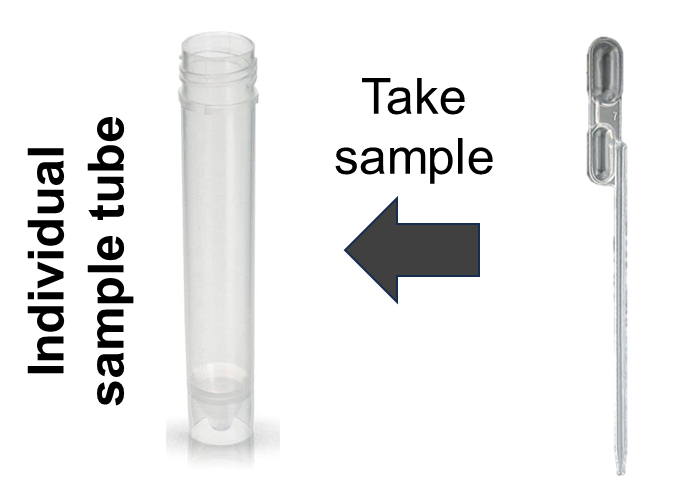 |  | 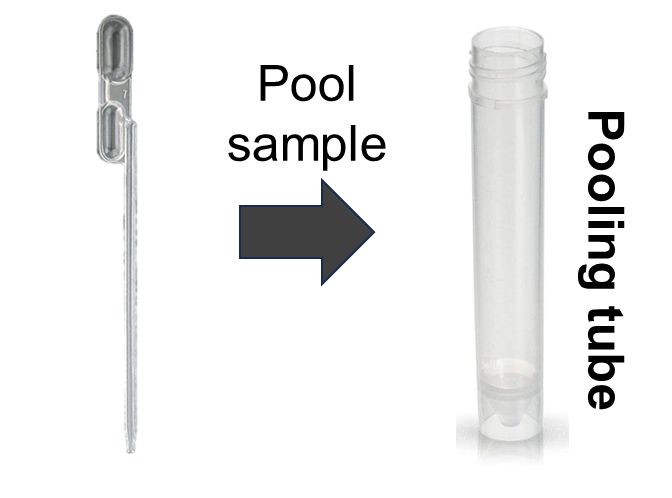 |
| Unwrap one pipette. Dispose of its plastic container into the waste bin |  | Draw the sample from the opened sample tube until the liquid fills the entire pipette tip |  | Carefully remove the pipette from the individual sample tube and transfer the liquid into the pooling tube |

| STEP #15 |  | STEP #16 |
| --- | --- | --- |
| 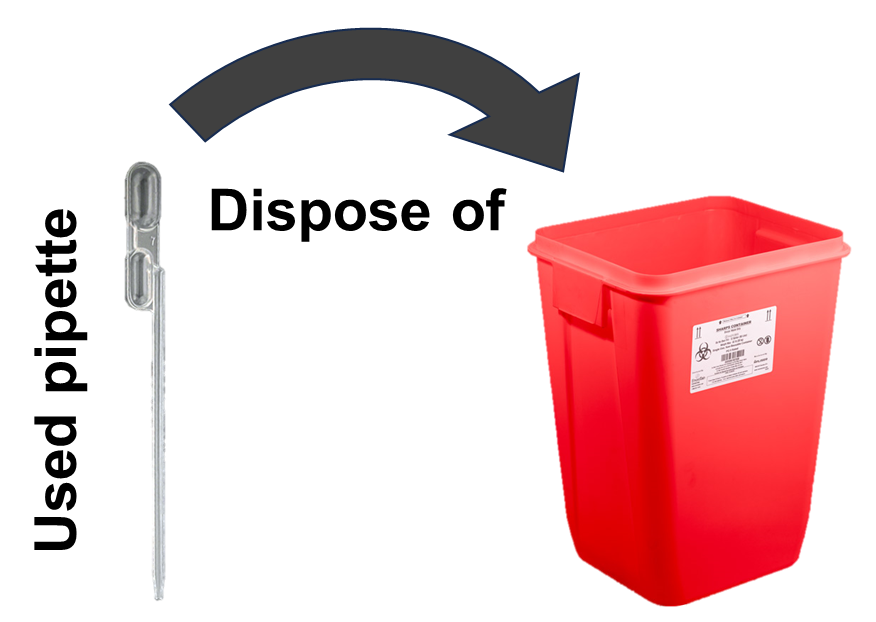 |  | 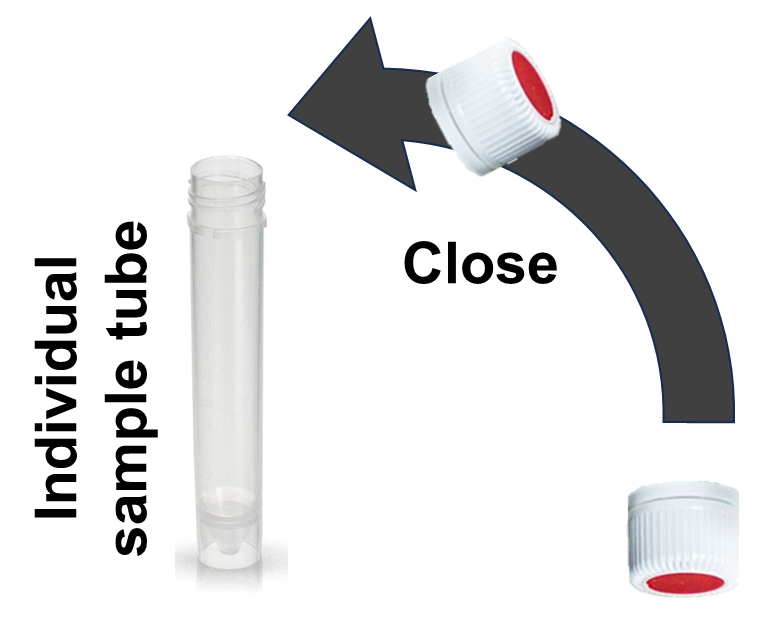 |
| Dispose of the pipette into the waste bin |  | Close the sample tube |

**<WRAP UP>**

PERFORM THE STEPS BELOW AFTER ALL 5 SAMPLES HAVE BEEN POOLED INTO THE POOLING TUBE

| STEP #17 |  | STEP #18 |  | STEP #19 |
| --- | --- | --- | --- | --- |
| 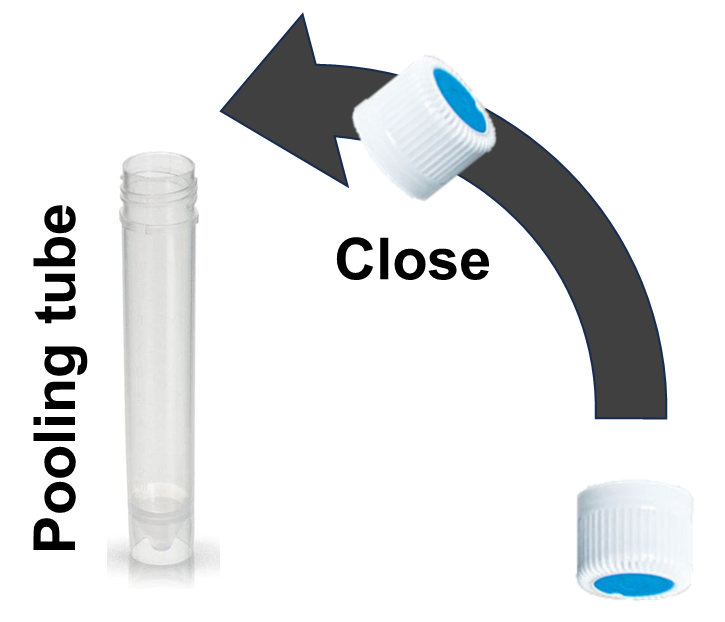 |  | 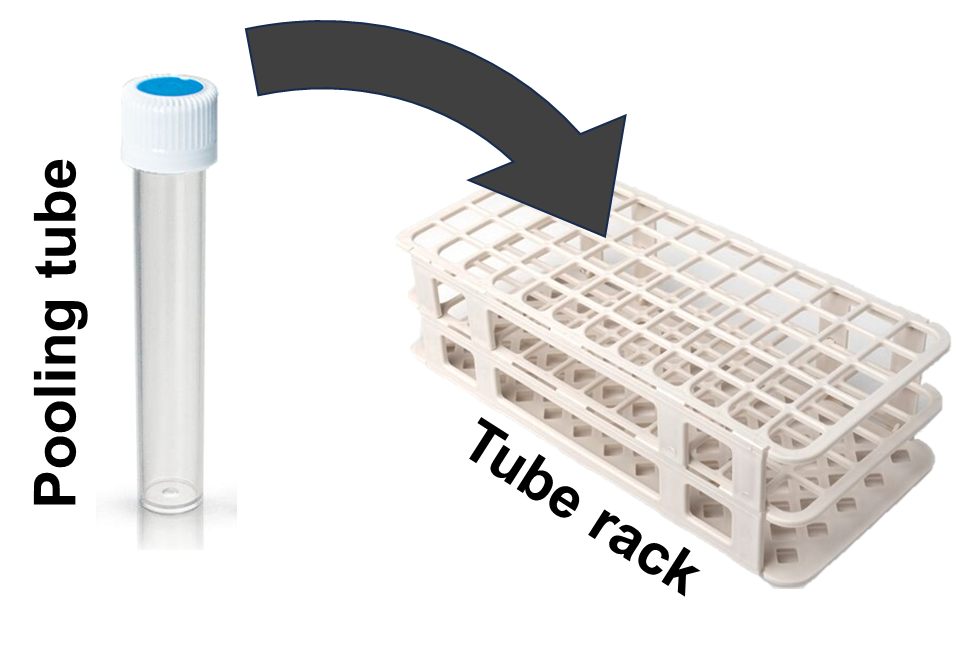 |  | 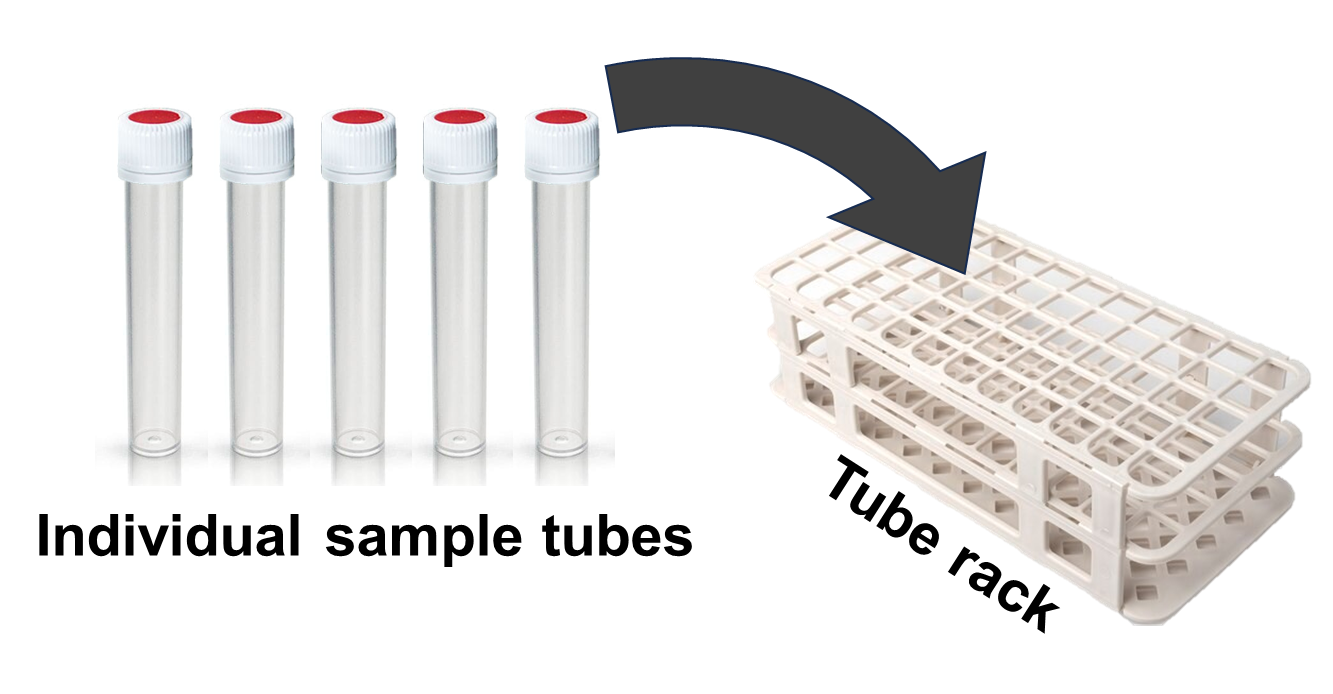 |
| Close the pooling tube |  | Put back the pooling tube into the tube rack |  | Make sure the individual sample tubes are placed in the tube rack |

| STEP #20 |  | STEP #21 |
| --- | --- | --- |
| 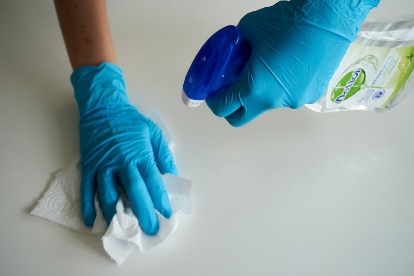 |  | 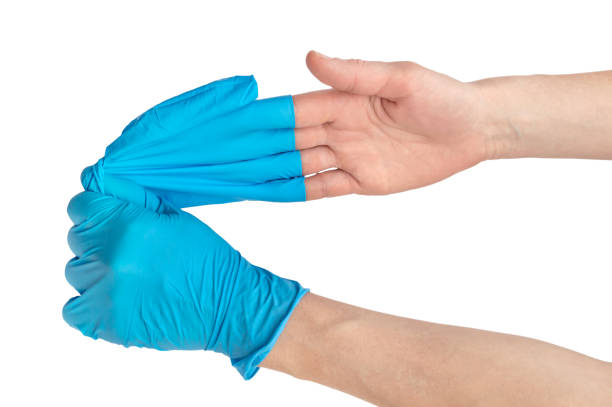 |
| Clean the workspace (or table) with the spray bottle and paper towels |  | Take off the gloves and dispose of them into the waste bin |

**Note S4. Classification and examples of uncorrected handling errors in the user study.** Sample pooling procedures require strict adherence to biosafety protocols to maintain sample integrity and worker safety. Uncorrected handling errors in the user study encompass three distinct categories: (1) correctable errors that were not corrected by participants or were corrected only after compromising the biosafety and sample integrity, (2) fundamentally uncorrectable errors that permanently compromise sample traceability, integrity, and reliability, and (3) correctable errors that were not corrected during the procedure, though they could have been safely remedied without compromising biosafety or sample integrity.

The first category includes errors that either remained uncorrected or were corrected too late to prevent potential contamination or biosafety risks. This classification reflects that in clinical settings where samples contain biohazardous materials, even corrected procedural deviations could have already led to pathogen transfer through cross-contamination between samples, contamination of clean materials, or exposure of workers to biohazardous materials. Additionally, the correction attempts themselves might introduce new risks by spreading contamination or compromising sample integrity. This approach to error classification acknowledges that in clinical settings, certain procedural mistakes can have significant consequences for sample integrity, worker safety, and pooling reliability ─ consequences that cannot be undone by subsequent corrections.

Following the definition, here are examples where participants attempted to correct their procedural mistakes, but these incidents were still recorded as uncorrected handling errors. Each example demonstrates why corrections, though made, cannot mitigate the potential biosafety risks that would arise in real-world clinical settings:

1. ‘Did not wear new gloves.’: During multiple pooling rounds, some participants reused their gloves instead of disposing of them between rounds. Even when participants recognized their mistake and changed gloves after cleaning the workspace, these incidents were recorded as uncorrected errors. This classification was made because in real-world clinical settings, gloves used in previous pooling rounds could potentially carry pathogens and compromise safety and integrity of pooling process through cross-contamination.

2. ‘Did not clean the workspace with new wipes at the beginning of pooling round.’: Some participants started new pooling rounds without cleaning the workspace first. Even when participants cleaned after already opening specimen tubes (pooling or individual), these incidents were recorded as uncorrected errors. This classification was made because in real-world clinical settings, two potential risks could arise: (1) gloves could become contaminated from the uncleaned workspace and transfer pathogens to open tubes; or (2) cleaning around open tubes could expose samples to disinfectants. Both scenarios could compromise sample integrity through contamination or degradation.

3. ‘Did not identify a new set of samples to be pooled’: Some participants failed to identify the correct sample set before beginning sample transfer, attempting to use samples from the wrong set. Even when participants recognized their mistake and switched to the correct set after opening tubes from the wrong set, these incidents were recorded as uncorrected errors. This classification was made because in real-world clinical settings, handling tubes from the wrong set could contaminate gloves with pathogens that might then be transferred to tubes from the correct set, compromising both biosafety and pool integrity.

4. ‘Did not prepare new pipettes before opening sample tubes.’: The instructions guide participants to take five unwrapped pipettes from a bulk package and prepare them in the working zone before scanning and opening the sample tubes. Even when participants prepared the pipettes after opening the sample tubes, these incidents were recorded as uncorrected errors. This classification was made because in real-world clinical settings, gloves potentially contaminated from handling open sample tubes could transfer pathogens to the bulk package of clean pipettes, compromising the integrity of remaining supplies and pooling through cross-contamination.

5. ‘Did not use a new pooling tube’: Some participants incorrectly opened used pooling tubes instead of using new ones for sample transfer. Even when participants later closed the used tubes and switched to new ones, these incidents were recorded as uncorrected errors. This classification was made because in real-world clinical settings, handling used pooling tubes could contaminate gloves with pathogens that might then be transferred to new tubes, compromising both biosafety and pool integrity.

6, ‘Did not open the sample tube and attempted to aspire sample from it.’: Some participants attempted to aspirate from tubes before checking if they were open, discovering they were closed during the attempt. Even when participants then opened the tube while holding the pipette or placing it on the workspace, these incidents were recorded as uncorrected errors. This classification was made because in real-world clinical settings, multiple contamination risks could arise in different contexts: (1) positive sample tube caps and tube openings could transfer pathogens to gloves and the clean pipette, (2) contaminated gloves could transfer pathogens to the negative sample in the tube and the clean pipette, and (3) the pipette placed on contaminated workspace surfaces could become contaminated. Any of these scenarios could compromise both sample integrity and pooling safety.

7. ‘Did not open the pooling tube until dispensing sample into the tube’: Some participants did not open the pooling tube before handling individual sample tubes. Even though they opened the pooling tube after already having opened an individual tube, these incidents were recorded as uncorrected errors in two scenarios: (1) when the pooling tube was opened after opening the individual tube but before aspiration, and (2) when the pooling tube was opened while holding a pipette containing the aspirated sample. This classification was made because in real-world clinical settings, both scenarios risk cross-contamination: contaminated gloves could transfer pathogens to the pooling tube cap, and in the second scenario, handling a filled pipette while opening the pooling tube creates additional risk of pathogen transfer and biosafety. Either situation could compromise the safety and integrity of the pooling process.

8. ‘Tried to transfer/remove sample from the pooling tube.’: Some participants incorrectly attempted to remove samples from the pooling tube instead of from individual sample tubes. These attempts included grabbing the pooling tube, positioning the pipette tip near the pooling tube opening, or inserting the pipette into the pooling tube. Even when no sample was actually removed, these incidents were recorded as uncorrected errors. This classification was made because in real-world clinical settings, such attempts could risk pathogen transfer through contaminated pipettes or gloves, compromising the safety and integrity of the pooling process.

9. ‘Did not close the sample tube until cleaning workspace’ and ‘Did not close the pooling tube until the end of pooling process’: Some participants did not close tubes until cleaning workspace. Even when participants later closed tubes during or after cleaning, these incidents were recorded as uncorrected errors. This classification was made because in real-world clinical settings, open tubes could create three potential risks: (1) sample spillage during cleaning activities, (2) sample degradation from exposure to disinfectants, and (3) contamination to cleaned workspace, gloves, or workers. All scenarios could compromise the safety and integrity of the pooling process.

10. ‘Closed the sample tube with the pooling tube cap’ and ‘Closed the pooling tube with the sample tube cap’: Some participants incorrectly switched tube caps between pooling and individual sample tubes (using pooling tube caps for individual tubes or vice versa). Even when participants later corrected this mistake, these incidents were recorded as uncorrected errors. This classification was made because in real-world clinical settings, contaminated caps could potentially transfer pathogens to individual negative samples in two ways: (1) when pooling tube caps placed in the working zone are used for closing the individual sample tubes, or when caps temporarily used on positive pooling tubes are later used to close individual negative samples. Both scenarios could transfer pathogens and compromise sample integrity through cross-contamination.

11. ‘Disposed of the individual sample tubes after closing it’: Some participants accidentally disposed of individual sample tubes after sample transfer and closing. Even when participants later retrieved these tubes from the waste bin, these incidents were recorded as uncorrected errors. This classification was made because in real-world clinical settings, retrieving tubes from medical waste creates serious biosafety risks through potential exposure to contaminated materials, compromising both worker safety and pooling integrity.

12. ‘Skipped closing the pooling tube and clean-up to pool from the next set.’: Some participants, after completing a set of five samples, failed to properly conclude that pooling set and incorrectly continued with samples from the next set while keeping the original pooling tube open. When pooling tubes remained open until participants grabbed and opened samples from the next set, these incidents were recorded as uncorrected errors, even if participants later closed the pooling tubes. This classification was made because in real-world clinical settings, this procedural confusion leads to critical errors by potentially mixing samples from different sets in the same pooling tube, compromising the integrity of the pooling process.

13. ‘Did not clean the workspace with new wipes at the end of pooling round.’ Some participants finished pooling rounds without cleaning the workspace. Even when participants performed the cleaning after removing their gloves, these incidents were recorded as uncorrected errors. This classification was made for two reasons: (1) pooling procedures require workspace cleaning while wearing gloves as part of the defined protocol (between wearing gloves at the start and removing them at the end), and (2) in real-world clinical settings, cleaning potentially contaminated surfaces with bare hands poses serious biosafety risks to workers through direct exposure to pathogens, compromising both worker safety and pooling integrity.

The secondary category comprises fundamentally uncorrectable errors that directly compromise sample integrity and pooling reliability. These errors include: ‘Scanned the wrong sample tube(s) from the wrong set(s),’ ‘Did not use a new pipette,’ ‘Transferred sample from a sample tube(s) that had already been pooled,’ Transferred sample from a sample tube(s) from the wrong set(s),’ ‘Transferred sample to the pooling tube of the wrong set.’ ‘Transferred sample from a sample tube to another sample tube,’ ‘Transferred sample from the pooling tube to a sample tube,’ ‘Removed sample from the pooling tube,’ ‘Re-aspirated with a used pipette that had been dipped in the pooling tube,’ and ‘Dispensed sample outside of the pooling tube.’ These errors represent fundamental breaches of pooling protocols that cannot be corrected once they occur, as they permanently compromise sample traceability, integrity, and reliability.

The third category of uncorrected handling errors includes procedural mistakes that could have been safely corrected without compromising biosafety or sample integrity, but remained uncorrected during the procedure. Unlike the previous categories, these errors were classified as uncorrected only when participants failed to correct them during the procedure, as their correction would not introduce contamination risks in real-world clinical settings. These errors include: ‘Did not scan the new sample tube #1,’ ‘Did not scan the new sample tube #2,’ ‘Did not scan the new sample tube #3,’ ‘Did not scan the new sample tube #4,’ ‘Did not scan the new sample tube #5,’ ‘Did not scan the pooling tube,’ ‘Did not transfer sample from the sample tube #1 to the pooling tube,’ ‘Did not transfer sample from the sample tube #2 to the pooling tube,’ ‘Did not transfer sample from the sample tube #3 to the pooling tube,’ ‘Did not transfer sample from the sample tube #4 to the pooling tube,’ and ‘Did not transfer sample from the sample tube #5 to the pooling tube.’

Through this classification of uncorrected handling errors, we recorded procedural mistakes in the user study that have varying degrees of consequences in real-world clinical settings where samples contain biohazardous materials.

**Note S5. Effect of completion time versus device intervention on volume-transfer accuracy and handling errors.** In the user study, participants performed pooling slowly with the device assistance compared to the paper instructions **(Supplementary Fig. S12)**. It is plausible that spending longer time could increase volume-transfer accuracy and reduce handling errors, regardless of device intervention. To disentangle the effects of completion time from true performance improvement as well as skill acquisition, linear mixed-effects analyses were performed across the 32 pooling rounds within each training protocol group (see **Supplementary Method S13**).

Impact of completion time (*t*_pool_) on volume-transfer accuracy (Acc_pool_)

*Protocol 1*

At the start of Protocol 1, participants’ completion time (*t*_pool_) was high **(Supplementary Fig. S10)**. As the pooling rounds progressed, *t*_pool_ decreased gradually (respiratory: *β*=−0.0525, *P*<0.0001; stool: *β*=−0.0463, *P*<0.0001), demonstrating a learning process within the paper-assisted pooling workflow **(Supplementary Fig. S10 and Table S13)**. Despite getting faster, their volume-transfer accuracy (Acc_pool_) plateaued (respiratory: *β*=−0.00365, *P*>0.05; stool: *β*=0.00481, *P*>0.05) at relatively low levels (respiratory~60%; stool~79%) throughout the 24 paper-assisted rounds, suggesting a potential floor effect **(Fig 4c,d,g,h and** **Supplementary Table S14)**. This pattern indicates that participants may have improved speed through procedural familiarity, without enhancing accuracy in sample transfer.

During paper use, a significant positive relationship between *t*_pool_ and Acc_pool_ was observed for artificial stool samples (*β*=0.0924, *P*<0.05; **Supplementary Table S14**), but not for artificial respiratory samples (*β*=−0.0875, *P*>0.05; **Supplementary Table S14**). Given that *t*_pool_ was initially high and decreased over the rounds while Acc_pool_ remained flat, this positive relationship for artificial stool pooling likely reflects incidental variation—participants happened to be more accurate when performing slowly, rather than due to a deliberate tradeoff between speed and accuracy across the pooling rounds.

When participants began the device-assisted pooling at Round 25, *t*_pool_ increased sharply, then gradually declined over subsequent rounds (respiratory: combined *β*=−0.0950, *P*<0.0001; stool: combined *β*=−0.179, *P*<0.0001), indicating a new learning curve specific to the device-assisted pooling workflow **(Supplementary Fig. S10 and Table S13)**. In parallel, Acc_pool_ improved significantly during the device-assisted pooling (respiratory: combined *β*=0.107, *P*<0.0001; stool: combined *β*=0.0718, *P*<0.001), suggesting the strong assistive effect of the device **(Fig. 4c,d,g,h and Supplementary Table S14)**.

During device use, a significant negative relationship between *t*_pool_ and Acc_pool_ was found for artificial respiratory samples (combined effect *β*=−0.250, *P*<0.01), but not for artificial stool samples (combined effect *β*=−0.0311, *P*>0.05). Since *t*_pool_ decreased while Acc_pool_ increased during device-assisted pooling, this negative relationship for artificial respiratory pooling likely reflects a true learning effect—participants became both faster and more accurate as they adapted to the device.

*Protocol 2*

During the first four rounds of paper-assisted pooling, participants showed similar patterns to those observed in Protocol 1: *t*_pool_ declined as participants became familiar with the workflow, while Acc_pool_ remained relatively stable **(Fig. 5c,d and Supplementary Fig. S10)**. Across all paper-assisted pooling sessions (i.e., Rounds 1─4, Rounds 13─16, Rounds 17─20, and Rounds 29─32), participants consistently became faster (respiratory: *β*=−0.0253, *P*<0.0001; stool: *β*=−0.0324, *P*<0.0001; **Supplementary Table S25**) and also gradually improved in Acc_pool_ (respiratory: *β*=0.0479, *P*<0.0001; stool: *β*=0.0236, *P*<0.0001; **Supplementary Table S26**). A significant positive relationship between *t*_pool_ and Acc_pool_ was observed in both sample types (respiratory: *β*=0.181, *P*<0.01; stool: *β*=0.149, *P*<0.0001), suggesting that participants tended to achieve higher volume-transfer accuracy when performing slowly **(Supplementary Table S26)**.

When the device was introduced at Round 5, participants initially required more time to complete the device-assisted pooling workflow, reflecting a new learning curve. As they adapted to the workflow, *t*_pool_ declined (respiratory: combined *β*=−0.0319, *P*<0.0001; stool: combined *β*=−0.0377, *P*<0.0001; **Supplementary Table S25**), while Acc_pool_ improved significantly (respiratory: combined *β*=0.0277, *P*<0.0001; stool: combined *β*=0.0235, *P*<0.0001; **Supplementary Table S26**) throughout Rounds 5─12 and Rounds 21─28. A significant positive relationship between *t*_pool_ and Acc_pool_ was observed for artificial stool samples (combined *β*=0.105, *P*<0.01), indicating that participants showed more accurate volume transfer when taking more time with the artificial stool samples under the device assistance **(Supplementary Table S26)**. However, no such relationship was found for artificial respiratory samples (combined *β*=−0.0593, *P*>0.05; **Supplementary Table S26**).

Across Protocol 1 and 2, slower pacing may have occasionally helped improve volume-transfer accuracy for artificial stool samples, likely because these samples were relatively easy to transfer, as reflected in their higher baseline accuracy (stool~79% versus respiratory~60%) In contrast, for artificial respiratory samples which were relatively difficult to transfer, slower pacing without the device intervention did not lead to improvements in Acc_pool_. Substantial gains in Acc_pool_ (e.g., from ~60% to >80%) were only observed when the device was introduced **(Figs. 4c,d,g,h and 5c,d,g,h)**, suggesting that these improvements were primarily driven by the assistive effects of the device, rather than by increased completion time alone.

This interpretation is further supported by performance in the post-device paper-assisted sessions (Round 13─16 & Round 29─32), where participants maintained high Acc_pool_ without device assistance, even as *t*_pool_ dropped immediately and continued to decline over rounds **(Fig. 5c,d,g,h and Supplementary Fig. S10, and Tables S25–26)**. The sustained high Acc_pool_, despite faster faster workflow, indicates that participants acquired deliberate volume-transfer skills through device-assisted pooling that effectively transferred to paper-based contexts. Notably, the significant positive relationships between *t*_pool_ and Acc_pool_ for both sample types during paper-assisted pooling in Protocol 2 suggest that participants who spent more time likely applied these learned skills more carefully, rather than simply slowing down without purpose. In this way, the device enabled not only immediate performance improvement but also skill acquisition beyond its direct use. These results point to the device's role as more than instructional assistance: it functioned as an assistive & training platform that guides participants through procedural steps and enables the development and transfer of volume-transfer skills.

Overall, no strong evidence was found that slower pooling workflow alone, independent of device assistance, led to substantial improvements in volume-transfer accuracy. Slower pacing may have modestly contributed to better accuracy for easier-to-transfer samples, reflecting incidental speed-accuracy tradeoffs. However, for more challenging samples like artificial respiratory specimens, slower workflow without device intervention was insufficient to drive meaningful gains. In contrast, the introduction of the device led to immediate and substantial improvements in accuracy for both sample types, indicating that device-guided support was essential for developing and executing accurate volume-transfer techniques. Therefore, device-assisted pooling not only enhanced immediate performance but also fostered lasting skill acquisition, enabling participants to apply deliberate, learned strategies beyond the device, rather than relying solely on slower pacing to achieve higher accuracy.

Impact of completion time (*t*_pool_) on the number of all uncorrected handling errors (*err*_all_), which is the sum of minor errors (*err*_minor_) and severe errors (*err*_severe_)

*Protocol 1*

During the 24-round paper-assisted pooling, *err*_all_ remained stable for both sample types (respiratory: *β*_all_=−0.00317, *P*>0.05; stool: *β*_all_=−0.00342, *P*>0.05) **(Fig. 4a,b, and Supplementary Table S15)**, while specific error types showed gradual improvements for certain sample types. *err*_minor_ and *err*_severe_ slightly decreased during the pooling with artificial stool (stool: *β*_minor_=−0.00417, *P*<0.05) and respiratory samples (respiratory: *β*_severe_=−0.00353, *P*<0.05), respectively **(Supplementary Table S15)**. Additionally, all error metrics (*err*_all_, *err*_minor_, and *err*_severe_) showed non-significant (*P*>0.05) relationships with *t*_pool_ for both sample types, except for *err*_minor_ showing a significant negative relationship with *t*_pool_ for artificial stool samples (*β*_minor_=−0.0492, *P*<0.01) **(Supplementary Table S15)**. Given that both *t*_pool_ and *err*_minor_ declined during paper-assisted pooling with artificial stool samples, this negative relationship likely reflects a true learning effect—participants occasionally tended to make fewer minor errors when doing the pooling more slowly.

When participants began the device-assisted pooling, all error metrics (*err*_all_, *err*_minor_, and *err*_severe_) dropped sharply and remained consistently low for both sample types (all *β*~0; *P*>0.05) across the last eight rounds **(Fig. 4a,b,e,f and Supplementary Fig. S12a–d and Table S15)**. Interestingly, significant positive relationships between *t*_pool_ and all error metrics (*err*_all_, *err*_minor_, and *err*_severe_) were observed for artificial stool samples (combined *β*_all_=0.104, *P*<0.01; combined *β*_minor_=0.0867, *P*<0.001; combined *β*_severe_=0.0475, *P*<0.05), whereas no such significant relationships were found for artificial respiratory samples (all *β*>0; *P*>0.05). Given that errors (*err*_all_, *err*_minor_, and *err*_severe_) remained low while *t*_pool_ gradually decreased during the device-assisted pooling, the significant positive relationships for artificial stool samples likely reflect that participants occasionally made more errors when it took longer to complete the pooling with the device.

*Protocol 2*

During the first 4-round paper-assisted pooling, the uncorrected handling errors did not change significantly among participants **(Fig. 5a,b)**. Across all paper-assisted pooling sessions (i.e., Rounds 1─4, Rounds 13─16, Rounds 17─20, and Rounds 29─32), participants exhibited sample- and error-type-specific patterns of error reduction. For respiratory sample pooling, *err*_minor_ gradually decreased (*β*_minor_=−0.00404, *P*<0.01), while the other metrics (*err*_all_ and *err*_severe_) did not change significantly (*β*_all_=−0.00408, *P*>0.05; *β*_severe_=−0.000437, *P*>0.05) **(Supplementary Table S27)**. For stool pooling, *err*_all_ and *err*_severe_ gradually declined (*β*_all_=−0.00695, *P*<0.0001; *β*_severe_=−0.00521, *P*<0.0001), whereas *err*_minor_ remained stable (*β*_minor_=−0.00124, *P*>0.05). Notably, significant negative relationships between *t*_pool_ and certain error metrics were observed: *err*_minor_ for artificial respiratory (*β*_minor_=−0.0436, *P*<0.05); both *err*_all_ and *err*_minor_ for artificial stool samples (*β*_all_=−0.0605, *P*<0.01; *β*_minor_=−0.0590, *P*<0.001). Given that *t*_pool_ gradually decreased across paper-assisted pooling rounds, the negative relationships indicate that participants occasionally tended to make fewer errors, particularly *err*_minor_ for artificial respiratory samples, when performing more slowly.

When the device was introduced at Round 5 (and 21), handling errors (*err*_all_, *err*_minor_, and *err*_severe_) decreased significantly and did not change significantly (all combined *β*~0, *P*>0.05) throughout device-assisted pooling sessions (i.e., Rounds 5─12 and Rounds 21─28) for both sample types **(Fig. 5a,b,e,f and Supplementary Fig. S12e–h and Table S27)**. Notably, significant negative relationships between *t*_pool_ and certain error metrics were observed during device use: *err*_all_ and *err*_minor_ for artificial respiratory samples (combined *β*_all_=−0.0755, *P*<0.05; combined *β*_minor_=−0.0732, *P*<0.001); *err*_severe_ for artificial stool samples showed (combined *β*_severe_=−0.0281, *P*<0.05) **(Supplementary Table S27)**. Given that *t*_pool_ gradually decreased while *err*_all_, *err*_minor_, and *err*_severe_ remained low, these negative relationships likely reflect that participants occasionally tended to make fewer minor or severe errors when it took longer to complete the pooling with the device.

Across Protocol 1 and 2, no strong evidence was found that slower pooling workflow alone, independent of device assistance, led to substantial reductions in handling errors (*err*_all_, *err*_minor_, and *err*_severe_). Slower pacing may have modestly helped reduce certain errors, particularly minor errors during the paper-assisted pooling; however, it was insufficient to drive the significant reduction. In contrast, the introduction of the device was associated with a sharp and sustained reduction in handling errors across all error types and sample types. Therefore, the significant reduction in handling errors was not simply a consequence of slower pooling workflow during device use but rather reflected improved procedural execution assisted by the device guidance.

Overall, substantial and sustained reductions in handling errors were achieved through use of the device, which provided real-time procedural instructions and error-correction support, rather than by slower handling alone.

**Note S6. Establishing criteria for high-quality sample pools to assess high-performance volume transfer in pooling procedures.** We investigated quality criteria for sample pools used in diagnostic testing, where one positive sample is typically combined with four negative samples. Precision is crucial for maintaining analytical sensitivity, since the positive sample may be pooled with several negative samples during the pooling process. The mass pooling ratio (MPR) exceeding 10 can significantly alter the pathogen load in the pool by more than one order of magnitude and cause a substantial Cq shift in qPCR analysis from the original positive sample's Cq value. This Cq shift, theoretically calculated as log_2_(MPR), would be approximately 3.32 cycles when MPR equals 10. Such a Cq shift can compromise the reliable detection of weak positive pools when the signal approaches or falls below the limit of detection (LOD), potentially leading to false negative results.

To prevent such diagnostic errors, we needed to establish a precision (CV_pool_) threshold that would ensure the maximum MPR (MPR_max_; the ratio of the pool's total weight to the minimum weight among the pooled samples) does not exceed 10. To identify the threshold, we derived a theoretical relationship between CV_pool_ and MPR_max_. The relationship was derived as follows:

$$\mu_{pool}=\frac{1}{5}\sum_{i=1}^{5} x_{i} (x_{1}\leq\cdots\leq x_{5})$$

where μ_pool_ is the mean volume of the five samples. Assuming the sample volumes are normally distributed, we can express the minimum volume in terms of the distribution parameters using a one-sided confidence limit:

$${MPR}_{max}=\frac{5\mu_{pool}}{x_{1}}\approx\frac{5\mu_{pool}}{\mu_{pool}-k\cdot\sigma_{pool}}=\frac{5\mu_{pool}}{\mu_{pool}-k\cdot\mu_{pool}\cdot{CV}_{pool}}=\frac{5}{1-k\cdot{CV}_{pool}}$$

where *k* is the z-score for a one-sided normal distribution and σ_pool_ is the standard deviation of the sample volumes. To identify the minimum value of CV_pool_ as a threshold, we picked *k* = 1.96, which ensures with 97.5% confidence that no sample volume will fall below the minimum volume ($x_{1}=\mu_{pool}-1.96\cdot\sigma_{pool}$). Solving the equation for CV_pool_ yields:

$${CV}_{pool}\left[ \% \right]=\frac{{MPR}_{max}-5}{{k\cdot MPR}_{max}}\times100 [\%]$$

At the critical MPR_max_ value of 10, this equation yields CV_pool,97.5%_=25.51%. We implemented a slightly more conservative threshold of CV_pool_=25% for practical applications, providing an additional margin of safety. Thus, we established CV_pool_≤25% as the first criterion for high-quality pools.

To validate this precision threshold (CV_pool_=25%), we conducted extensive Monte Carlo simulations (n=20,000) examining the relationship between MPR_max_ and CV_pool_ of simulated pools across different pipetting performance of personnel. We modeled pipetting performance using truncated normal distributions of sample volume (population mean μ_pipette_=300 uL) transferrable by dual-bulb pipettes. Since individual participants have different inherent variability of volume-transfer, we simulated with different CV_pipette_ (i.e., σ_pipette_/μ_pipette_ ×100) ranging from 5% to 45%. The result **(Supplementary Fig. S15a,b)** showed that simulated data at CV_pool_=25% fell within MPR<10, confirming the suitability of our CV_pool_ threshold.

Next, we investigated what accuracy threshold would complete our criteria for high-quality pools, focusing on how these criteria together could effectively identify pools created with high-performance volume transfer. A key hypothesis of our study was that participants could develop effective volume-transfer skills using 300-μL dual-bulb pipettes through either paper or device-based training, with emphasis on controlled (slow, gentle, and smooth) aspiration and dispensing. To establish baseline performance associated with different pipetting techniques, we first conducted preliminary experiments comparing ideal (slow, controlled dispensing) and suboptimal (rapid dispensing) volume-transfer techniques. For artificial respiratory samples of 300 μL, ideal technique achieved 92.21% accuracy (Acc_pipette_) with 6.819% CV_pipette_ precision, while suboptimal technique showed 49.74% accuracy with 37.42% CV_pipette_ precision **(Supplementary Fig. S15c)**. For artificial stool samples of 300 μL, ideal technique demonstrated 99.76% accuracy with 2.95% CV_pipette_ precision, compared to suboptimal technique's 76.85% accuracy with 20.35% CV_pipette_ precision **(Supplementary Fig. S15c)**. These results indicate that untrained personnel with poor volume-transfer techniques may need to improve in both accuracy (Acc_pipette_; 49.74%~76.85% vs. 92.21%~99.76%) and precision (CV_pipette_; 20.35%~37.42% vs. 2.95%~6.819%) to gain ideal volume-transfer techniques.

Based on these preliminary results showing the need for improvements in both metrics, we developed three models using truncated normal distributions to simulate two different potential improvement scenarios: improvement in accuracy alone and improvement in both accuracy and precision from the baseline before training. We did not consider the improvement in precision alone, since dual-bulb pipettes used in this study are designed for users to transfer 300 uL of liquid sample. We assumed that during the training, untrained personnel tend to achieve the transfer of 300 uL accurately initially and then achieve it precisely once they transfer the sample close to 300 uL. Below are the models for three different performance levels **(Supplementary Fig. S15d)**.

- Pre-training baseline (BF_25%_): μ_pipette_=202.5 μL [Acc_pipette_=67.5%], CV_pipette_=25%
- Accuracy improvement (AF_25%_): μ_pipette_=285 μL [Acc_pipette_=95%], CV_pipette_=25%
- Both accuracy and precision improvement (AF_5%_): μ_pipette_=285 μL [Acc_pipette_=95%], CV_pipette_=5%

Using these models, we performed Monte Carlo simulations (n=20,000) to identify the optimal accuracy threshold that would complete our criteria for high-quality pools. We evaluated pass rates (the percentage of pools meeting both accuracy and precision criteria) across different accuracy thresholds (70-90%) to answer critical questions:

1. What accuracy threshold, combined with our established CV_pool_ threshold, best differentiates between poor and high-performance volume transfer?
2. Are pools meeting these combined criteria achievable as volume-transfer skills improve?

Based on the simulation results **(Supplementary Fig. S15e)**, we established Acc_pool_=80% as the second criterion for high-quality pools. Together with our previously established CV_pool_ ≤25%, this accuracy threshold created a comprehensive framework for identifying high-quality pools. With these combined criteria, the simulation predicts pass rates of 3.52% from poor performance (BF_25%_), increasing to 55.03% from improved accuracy (AF_25%_), and reaching 100% from improved accuracy and precision (AF_5%_). This graduated improvement (3.52%→55.03%→100%) effectively captures different levels of technical proficiency, compared to other criteria showing either too lenient passing rates (Acc_pool_≥70%: 30.05%→69.45%→100%) or overly stringent requirements (Acc_pool_≥85%: 0.50%→26.70%→100%; Acc_pool_≥90%: 0.03%→6.05%→100%) that fail to capture meaningful performance improvements. These distinct performance tiers demonstrate that our criteria for high-quality pools effectively identify improved volume-transfer skills, as they are challenging for untrained personnel yet increasingly achievable with proper skill development.

The suitability of these criteria is further supported by effect size analysis **(Supplementary Fig. S15e)**. At the 80% accuracy threshold, the analysis reveals comparable improvements for accuracy improvement alone (Cohen's h=1.294) versus the additional benefit of precision improvement (incremental Cohen's h=1.470), suggesting balanced sensitivity to both metrics. In contrast, at the ≥85% threshold, the incremental improvement from precision substantially exceeds that from accuracy improvement alone (Cohen's h=0.951), indicating potential bias toward precision. Considering both the simulation results and effect size analysis, we established our comprehensive criteria for high-quality pools as Acc_pool_ ≥80% and CV_pool_ ≤25%. These criteria provide the most balanced and practical approach for identifying pools created with high-performance volume transfer, making them valuable metrics for assessing training outcomes in our study.

**Note S7. Quick information sheets provided to participants before starting pooling on Day 1.** All participants were provided the quick information sheets, which describe the motivation and purpose of the user study, the value and mechanics of sample pooling, specific learning/training objectives from the pooling exercise, the mechanism of dual-bulb pipettes, possible causes and effects of cross-contamination, and key handling skills to avoid cross-contamination. We explained to individuals step-by-step with the information sheets below.

**********

**Quick Info Sheets: Manual Sample Pooling**

Sample pooling is a strategy used to enhance efficiency and cost-effectiveness when testing large quantities of samples. The method involves combining multiple individual samples into a single “pool,” which is then subjected to a diagnostic test. If the pooled sample is negative, it implies that all the individual samples within that pool are also negative, significantly reducing the number of tests needed. However, if the pool is positive, all the individual samples can then be tested to pinpoint which were positive. Sample pooling has been valuable during public health crises, such as the COVID-19 pandemic, where widespread testing is essential for early detection and containment.

This manual will guide you to perform 5-sample pooling (combining five individual samples into a single pool) safely with biological samples, including, but not limited to: urine, sputum, saliva, swabs in transport media, and other biofluids from humans (however the samples used do not contain any human material).

You will be asked to perform these steps for multiple rounds of pooling and a researcher will observe you. Afterward, you will be given a short questionnaire about your experience. The researcher may additionally ask you a few questions about your experience.

Note:

- This usability study is intended for research only.
- This usability study is NOT intended for evaluating participants’ abilities.
- Results may be published but participants will remain anonymous; data collected during recruitment and in the questionnaires will not be tied to your identity.

If you have any questions, please feel free to reach out to Minkyo Lee.

Minkyo Lee (Principal Investigator: Rustem F. Ismagilov)

A graduate student in Andrew and Peggy Cherng Department of Medical Engineering,

California Institute of Technology (Caltech)

Phone: XXX-XXX-XXXX

Caltech IRB Protocol #23-1366

The materials you will use in this study are as follows:

- Deionized water
- 70% ethanol
- Contrived respiratory samples
  - Components: water, polyethylene glycol, mucin from porcine stomach, and Lambda gDNA
- Contrived stool samples
  - Components: water, polyethylene glycol, cellulose, psyllium husk, sodium (NaCl), potassium chloride (KCl), calcium hydroxide (CaCl_2_), miso paste, and oleic acid
- Disposable dual-bulb pipettes, 2 mL sample collection tube, Tube racks, Protective rubber gloves, Protective eyeglasses, Wipes
- **Objectives of the practice of manual sample pooling**

By the end of the practice of manual sample pooling, participants will be able to:

1. Describe sample pooling and provide examples of its application.
2. Generate pools from five individual contrived clinical samples without supervision.
3. Transfer the fixed volume of viscous liquid accurately and reliably using dual-bulb pipettes.
4. Handle potential biohazardous materials safely.

- **Biohazards**

None of the materials you will use are biohazardous. Instead, we use contrived (fake) samples that resemble real clinical samples (stool, respiratory samples). For this usability study, you will treat all samples as if they are real clinical samples and thus biohazardous (samples from patients, such as those with COVID-19). The instructions are designed for a participant to perform a single cycle for 5-sample pooling (combining five samples into a single pool).

- **Cross-contamination & Proper Handling skills**

Cross-contamination refers to the unintended transfer of material (such as microorganisms or DNA) from one sample to another. This transfer can occur during various stages of handling, processing, and analysis of samples. Cross-contamination can lead to inaccurate test results, misdiagnosis, or **false positives**.

Several factors can contribute to cross-contamination:

1. **Improper handling of samples**: Using the same materials (pipettes, tubes, or containers) without proper cleaning or decontamination between samples, can lead to cross-contamination.
2. **Proximity during processing: When multiple samples are processed close by, there's a risk of material transferring between them through aerosols, droplets, or contact.**
3. **Laboratory environment**: Contaminants in the lab environment, including airborne particles, aerosols, or residues from previous analyses, can be deposited on samples.
4. **Human error**: Mistakes made by personnel, such as inadequate handwashing, improper use of personal protective equipment (PPE), or accidental spills, can result in cross-contamination.

- To prevent cross-contamination, laboratories follow strict protocols and quality control measures:
- **Clean environment**: Instruments, containers, and surfaces used for sample processing are properly cleaned and sterilized.
- **Separation and isolation**: Samples are physically separated, we use separate workspaces, and we handle each sample with dedicated equipment.

1. We compartmentalize the working area into three zones: dirty zone, clean zone, and pooling zone.
   a. Dirty zone: All samples will be placed in this zone.
   b. Clean zone: All materials that should not be contaminated will be placed in this zone.
   c. Pooling zone: We will do pooling in this zone.
2. Please assign the above zones appropriately. The dirty zone should be the farthest from the clean zone. The area between the dirty zone and the clean zone would become the pooling zone.
3. Place the glove box, pipettes, wipes, spray, and empty tubes in the clean zone.
4. Place the individual sample tubes with the tube rack in the dirty zone.

- **Proper handling skills**: Touch only the outside of the cap and body of the tube. Do not touch the inside of the tube, the mouth of the tube, or the inside of the cap.

| 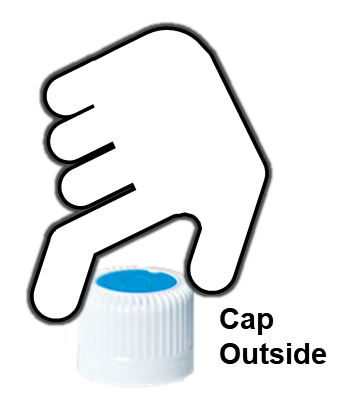 |  | 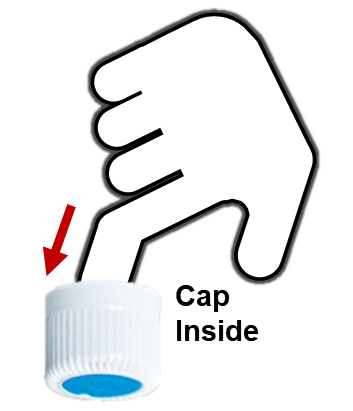 |  | 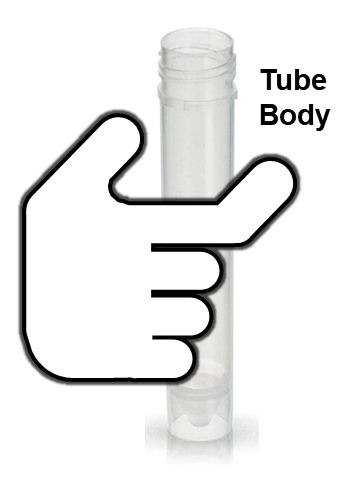 |  | 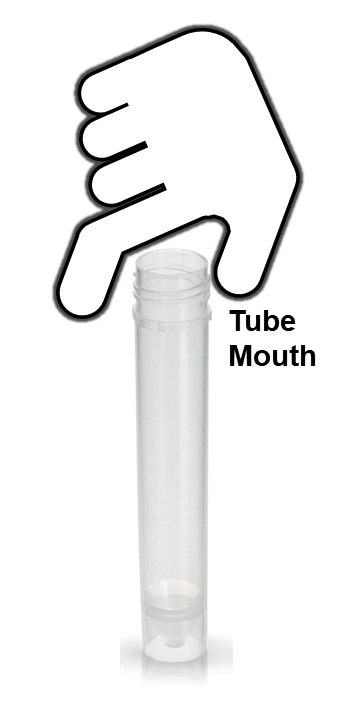 |
| --- | --- | --- | --- | --- | --- | --- |
| **Good** |  | **Bad** |  | **Good** |  | **Bad** |

**Note S8. Inclusion & ethics statements**

All authors of this study have fulfilled the criteria for authorship as their contributions were critical for design, implementation, and interpretation of the study. This work includes findings that are locally relevant, which have been determined in collaboration with local partners, including the International Centre for Diarrhoeal Disease Research, Bangladesh (icddr,b). Roles and responsibilities were agreed among collaborators ahead of the research. In addition, capacity-building plans were discussed and encouraged to lead to manuscripts. Detailed author contribution statements are available in **Supplementary Note S9**. This work was not severely restricted or prohibited in the setting of the researchers, and did not result in stigmatization, incrimination, discrimination or personal risk to participants. Local and regional research relevant to our study was taken into account in citations.

This study included human participants research conducted in Pasadena, CA, United States of America. The research was reviewed by the California Institute of Technology Institutional Review Board (IRB) and determined meet the criteria for exemption pursuant to (45 C.F.R. § 46.104(d)(2)(i),(ii)): Research that only includes interactions involving educational tests (e.g., cognitive, diagnostic, aptitude, achievement tests), survey procedures, interview procedures or observation of public behavior (IRB protocol #23-1366). In addition, this study utilized archived clinical samples collected in Bangladesh (study sites: Gazipur, Kishoreganj, Mymensingh, and Tangail districts) during 2015-2016 from research conducted at icddr,b. The sample collection was approved by the Ethical Review Committee at icddr,b (PR-11063), the Committee for the Protection of Human Subjects at the University of California, Berkeley (2011-09-3652), and the institutional review board at Stanford University (25863). The local researchers identified eligible communities through area surveys, traveled to the eligible communities, asked community leaders for permission to conduct research within their community, and recruited participants with formal informed consent for the study in the communities under the permission from the leaders.

**Note S9. Detailed author contribution statements and ORCiDs.**

Minkyo Lee (M.L.): Conceptualized the study with R.F.I.; major contributor to all device development and demonstrations with input from R.F.I. and S.H.J., including the device layout design and module fabrication, the system architecture (hardware and software) design, integration, and demonstrations for instructional guidance, real-time error monitoring, weight measurement, documentation, and wireless data transmission; major contributor to all device designs with input from R.F.I., N.S., S.H.J. and A.V.W., including language-agnostic graphics, device color-coding, and device module shapes compatible with device-protective polypropylene trays; major contributor to the selection of laboratory materials (e.g., specimen tubes, individually wrapped dual-bulb pipettes) with input from R.F.I., S.H.J., and A.V.W.; major contributor to the user study design and implementation with input from R.F.I., N.S., C.F.C., and A.V.W., including optimization and preparation of artificial respiratory and stool samples, development of a customized data logger and a barcode scanner, preparation of original draft of paper instructions with texts and graphics for action diagrams, preparation of original drafts of quick info sheet and handling error check sheet, preparation of original drafts of modified versions of user surveys (i.e., PSSUQ, KTEQ, PTEQ), establishment of the eligibility criteria of participants, participant recruitment, enrollment, random assignment, compensation, and follow-up, and user data acquisition and extraction from the error sheet, data logger, and instructional device; major contributor to the user data analysis and interpretation with input from R.F.I., R.G., C.F.C., N.S., and A.V.W.; wrote original drafts of SOPs with biosafety guidelines for clinical STH stool homogenization, pooling, and extraction; collaborated with R.F.I., N.P., S.A.W., and X.P.P. on the design and implementation of clinical STH pooling for the device validation clinical study at Caltech; co-designed the clinical STH pooling and extraction procedures for the device validation study with X.P.P.; determined the random selection of clinical samples for the device validation study; performed all clinical STH sample homogenization and pooling on the device for the device validation study; acquired and extracted all weight data measured by the device during the clinical pooling; contributed to securing funding; generated and edited all figures, tables, notes, and videos in the Main text, Expanded Data, and Supplementary Information with input from all authors; outlined and wrote the original draft of manuscript with input from R.F.I.; edited the manuscript with input from all authors.

Xinyue (Penny) Pei (X.P.P.): Major contributor to all clinical STH sample processing (i.e., storage, homogenization, DNA extraction, and qPCR analysis) at Caltech; modified the STH stool homogenization and DNA extraction protocols for biosafety at Caltech; edited and finalized SOPs with biosafety guidelines for clinical STH stool homogenization, pooling, and extraction at Caltech; wrote and established SOPs with biosafety guidelines for qPCR analysis from clinical stool elution at Caltech; introduced the Beadbug6 homogenizer and evaluated its performance for the modified homogenization and extraction protocols at Caltech; evaluated extraction yields of the modified protocols at Caltech; performed preliminary individual STH stool homogenization and extraction protocols, tested the detection of *Ascaris lumbricoides* DNA on qPCR, and established the correlation of Cq values between Caltech and Smith College; established storage process of clinical stool suspensions and elutions and thawing protocols for the device validation study; collaborated with R.F.I., M.L., N.P., and S.A.W. on the design and implementation of clinical STH pooling for the device validation clinical study at Caltech; co-designed the clinical STH pooling and extraction procedures for the device validation study with M.L.; performed all nucleic acid extractions and qPCR analysis for the device validation study; extracted and analyzed all qPCR data; contributed to visualization of Supplementary Fig. 14a,c; wrote ‘qPCR analysis’ in the Methods section; reviewed and provided feedback on the manuscript.

Si Hyung Jin (S.H.J.): Contributed to the device design; major contributor to the design and manufacturing of polypropylene device-protective trays using vacuum-forming technology; provided expertise on designs for easy-to-understand language-agnostic graphics, device module architectures for contamination-preventive hand movements, and error-preventive device pooling procedure; contributed to designs of device modules compatible with trays; contributed to the visualization of Fig. 1; reviewed and provided feedback on the manuscript.

Natasha Shelby (N.S.): Provided feedback on designs of language-agnostic graphics; contributed to the user study design; collaborated with M.L. on recruitment strategies; wrote the original and revised IRB protocols; contributed to editing paper instructions, quick info sheet, handling-error check sheet, and modified versions of user survey questionnaires (i.e., PSSUQ, KTEQ, and PTEQ); established demographic questionnaire and enrollment questionnaire; collaborated with R.F.I. and M.L. to establish the eligibility criteria of participants for the user study; managed questionnaires in Qualtrics; contributed to user data management; contributed to data interpretation of the user study; assisted with securing funding; managed the overall study budget; created the original **Supplementary Fig. S6**; contributed to making the device demonstration video **(Supplementary Movies 1 and 2)**; major contributor to writing and editing all sections of the manuscript.

Rani Gera (R.G.): Major contributor to data analysis and interpretation of the user study; provided technical guidance and expertise on selecting statistical models (e.g., linear mixed-effect regressions using dummy coding) and the analysis of every user data set; contributed to data presentation with statistical results in Figs. 3c-d, 4-6, and Extended Data Figs. 2e-f, 4, 6, and 7; reviewed and provided feedback on the manuscript.

Alexander Viloria Winnett (A.V.W.): Provided guidance on clinical microbiology laboratory processes and sample handling for the device design (including device layout, language-agnostic graphics, color coding, specimen collection tube compatibility, device features) and pooling procedure (including sample homogenization and transfer, contamination avoidance, biosafety); pilot user for iterations of device design; contributed to the design of the user study (including cross-over design to assess assistive vs. training functions of the device, measures to collect, participant compensation scheme, artificial human sample selection, and analysis approach with selection of statistical models for analyses shown in Fig. 3a-b and Extended Data Fig. 2a-b); assisted M.L. in interpretation of user study results; major contributor to the visualization of Fig. 1a; contributed to data presentation in Figs. 4, 5, and 8b-c,f and Extended Data Fig. 2c; major contributor to the outline of the Introduction section, including relevant background literature; contributed to editing Introduction, Results, Discussion, and Outlook of the manuscript.

Colin F. Camerer (C.F.C.): Co-investigator; provided guidance and expertise on user study design and data analysis; reviewed and provided feedback on the manuscript.

Mahbubur Rahman (M.R.): Co-investigator; major contributor to all research work regarding clinical sample acquisition and management in Bangladesh; contributed to design and plan for the WASH Benefits Bangladesh trial; prepared IRB protocols for the study in Bangladesh; managed the field implementation and trained local researchers for sample/data collection from children in rural Bangladesh; managed quality assurance and tracking of collected clinical samples; developed protocols for sample preservation, storage, and transport; collaborated with N.P. and S.A.W. on sample storage and shipment to Smith College; reviewed and provided feedback on the manuscript.

Nils Pilotte (N.P.): Co-investigator; major contributor to clinical sample characterization, development of DNA extraction and sample pooling protocols, and qPCR analysis for pre-screening clinical samples at Smith College; collaborated with S.A.W. and M.R. on sample shipment from Bangladesh to Smith College; collaborated with R.F.I., M.L., X.P.P., and N.P. on the design and implementation of clinical STH pooling for the device validation clinical study at Caltech; provided technical guidance and expertise on sample extraction and pooling experiments performed at Caltech; reviewed and provided feedback on the manuscript.

Steven A. Willams (S.A.W.): Co-investigator; major contributor to clinical sample characterization, development of DNA extraction and sample pooling protocols, and qPCR analysis for pre-screening clinical samples at Smith College; collaborated with N.P. and M.R. on sample shipment from Bangladesh to Smith College; collaborated with R.F.I., M.L., X.P.P., and N.P. on the design and implementation of clinical STH pooling for the device validation clinical study at Caltech; provided technical guidance and expertise on sample extraction and pooling experiments performed at Caltech; contributed to establishing the eligibility criteria of participants for the user study; reviewed and provided feedback on the manuscript.

Rustem F. Ismagilov (R.F.I.): Principal investigator; conceptualized the study with M.L.; provided technical guidance, oversight of all studies and analyses, including device development, the user study, and the device validation study with clinical STH samples, and was responsible for obtaining the primary funding for the study.

ORCiDs

Minkyo Lee,, 0000-0002-2714-179X

Xinyue (Penny) Pei,, 0009-0003-9840-6243

Si Hyung Jin,, 0000-0003-3376-2549

Natasha Shelby,, 0000-0001-9097-3663

Rani Gera,, 0000-0003-1888-4337

Alexander Viloria Winnett,, 0000-0002-7338-5605

Colin F. Camerer,, 0000-0003-4049-1871

Mahbubur Rahman,, 0000-0003-0520-2683

Nils Pilotte,, 0000-0002-8447-7425

Steven A. Williams,, 0000-0002-4881-7496

Rustem F. Ismagilov,, 0000-0002-3680-4399

**Note S10. Detailed acknowledgement statements.**

We thank A. E. Romano (Caltech) for her feedback on device layout and pooling procedure, revision of Institutional Review Board protocols for the user study, acquisition of key laboratory materials (e.g., tube specimens and dual-bulb pipettes), her expertise and advice on clinical STH sample processing (e.g., homogenization, extraction, pooling, and DNA detection), and establishment of standard operating procedures (SOP) with biosafety guidelines for implementation of clinical STH experiments. We thank M. Cooper (Caltech) for the ideation of pooling device, feedback on device layout and pooling procedure, biosafety paperwork, SOP establishment, and procedural demonstrations for clinical STH experiments. We thank R. Lazarovits (Caltech) for the ideation of device modules and feedback on device layout and pooling procedure. We thank O. Pradhan (Caltech) for his feedback on device layout and pooling procedure, biosafety paperwork, SOP establishment, and procedural demonstrations for clinical STH experiments. We thank N. J. Wu-Woods (Caltech) and R. Akana (Caltech) for providing feedback on device layout and pooling procedure. We thank M. K. Porter (Caltech) for his feedback on device layout and pooling procedure, his expertise on stool sample homogenization, and advice on the outline of manuscript. We thank M. K. Kim (Caltech) for organizing the compensation of participants in the user study. We thank S. Haynes (Smith College) for her administrative assistance and advice on institutional biosafety guidelines for clinical STH experiments, and shipment of clinical STH stool samples. We thank A. M. Gonzalez (Smith College) for his assistance in the processing of clinical human stool aliquots and the interpretation of clinical STH sample data.

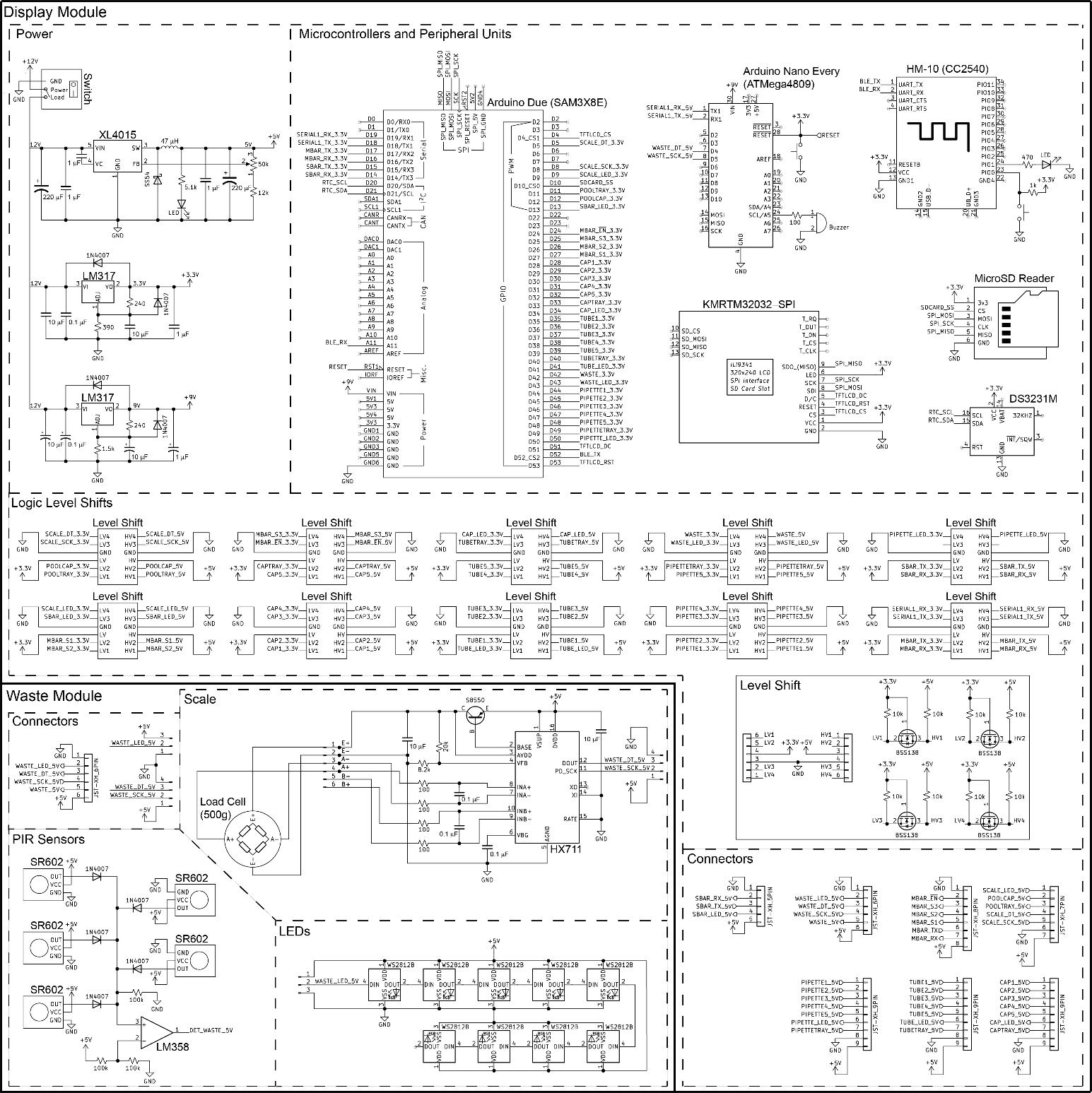

**Fig. S1.** Detailed circuit schematics of Display and Waste Modules of the instructional pooling device.

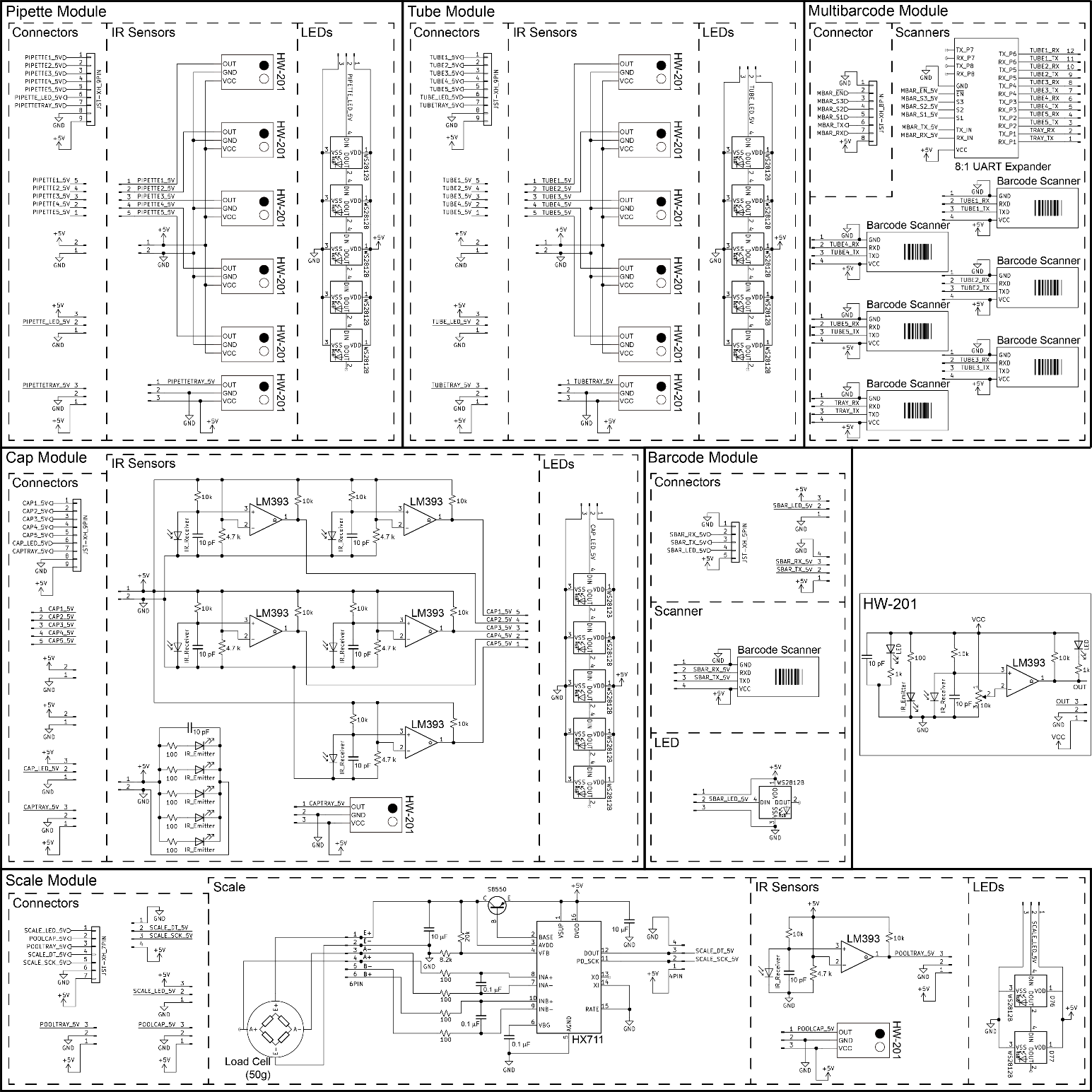

**Fig. S2.** Detailed circuit schematics of Pipette, Tube, Multibarcode, Cap, Barcode, and Scale Modules of the instructional pooling device.

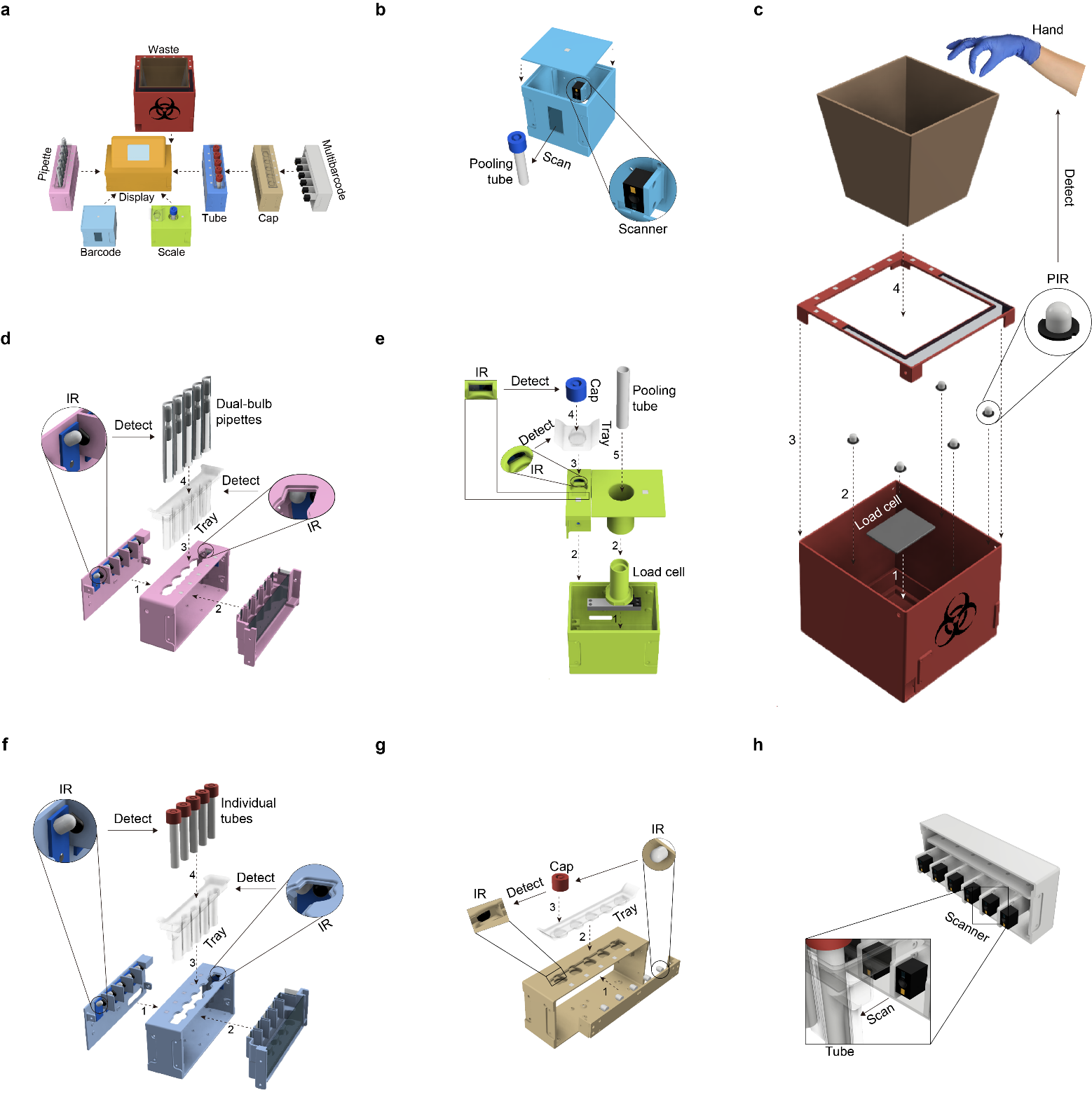

**Fig. S3. Detailed physical design of instructional pooling device for the detection of various objects. a**, Modular type Display Module showing language-agnostic graphics. **b**, Barcode Module with an integrated scanner for identifying barcodes. **c**, Waste Module with PIR sensors and a load cell for detecting waste disposal by a user. **d**, Pipette Module with IR sensors for detecting individually wrapped dual-bulb pipettes and their tray. **e**, Scale Module integrating IR sensors and a load cell for detecting the pooling tube, its cap, and the tray. **f**, Tube Module with IR sensors for detecting individual tubes and their tray. **g**, Cap Module with IR sensors for detecting tube caps and their tray. **h**, Multi-barcode Module with six integrated scanners for scanning barcodes of individual tubes and their tray.

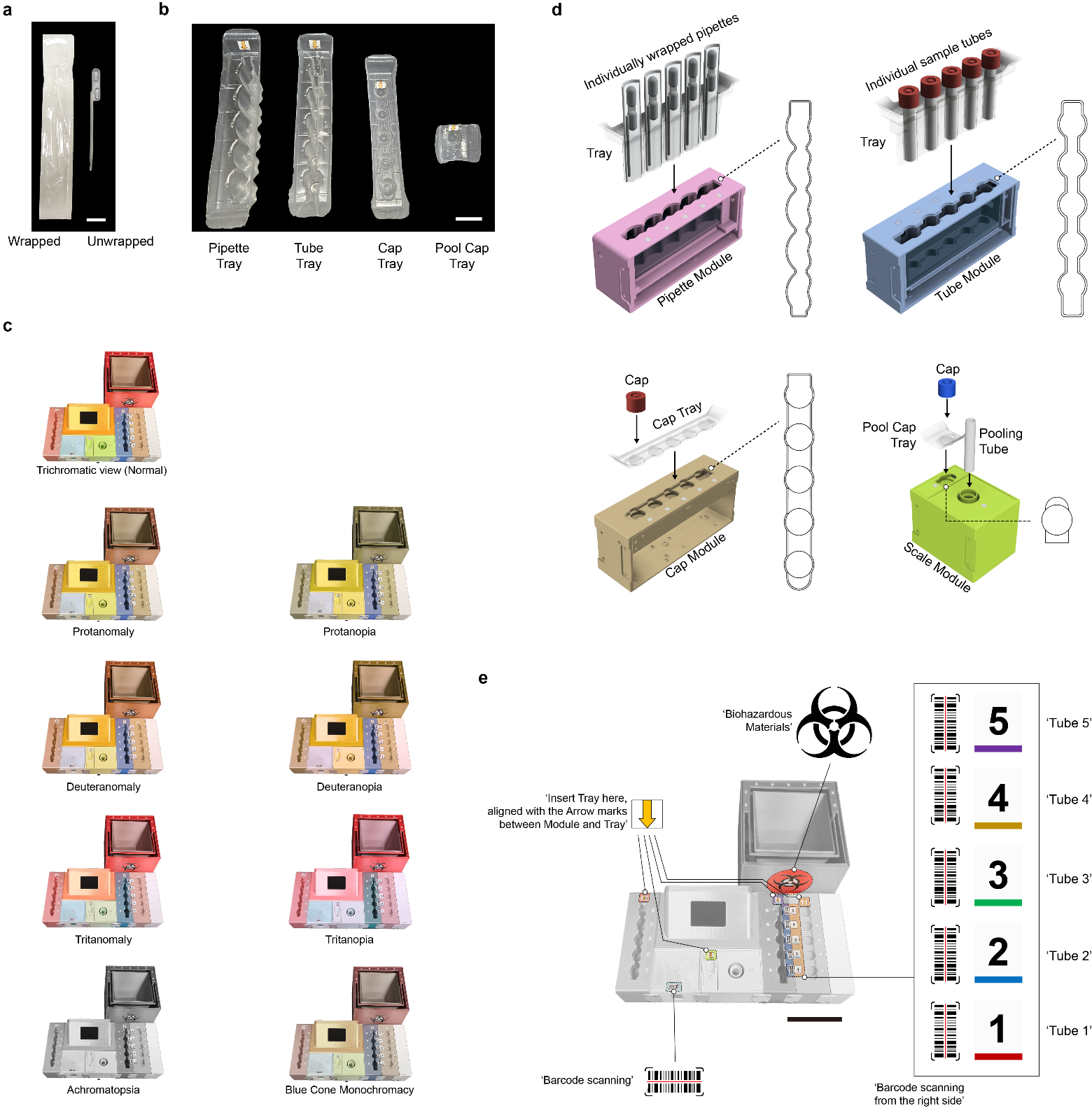

**Fig. S4. Laboratory materials and design components to support the instructional pooling device.** **a**, Individually wrapped and unwrapped 300-μL disposable dual-bulb pipettes. **b**, Device-protective polypropylene trays for pipettes, individual sample tubes, tube caps, and the pooling tube cap. **c**, Simulated device colors across different types of color blindness. Simulation website: <https://www.color-blindness.com/coblis-color-blindness-simulator/> **d**, Device modules with unique shapes to allow their corresponding device-protective trays. **e**, Symbolic indicators on the device.

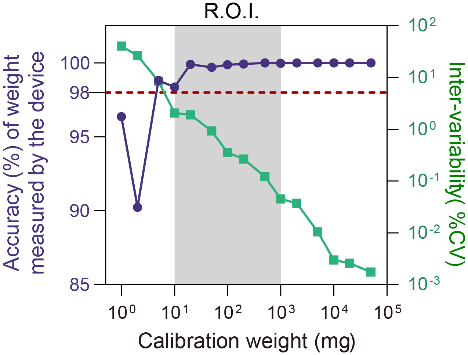

**Fig. S5. Average accuracy and inter-variability of weight measurement (n=20 for each weight) of the device for known mass (1 mg to 50 g).** R.O.I, region of interest, defined as the range of sample volumes capable of being transferred by dual-bulb pipettes. All measured weight values indicated here were converted to mass units (mg).

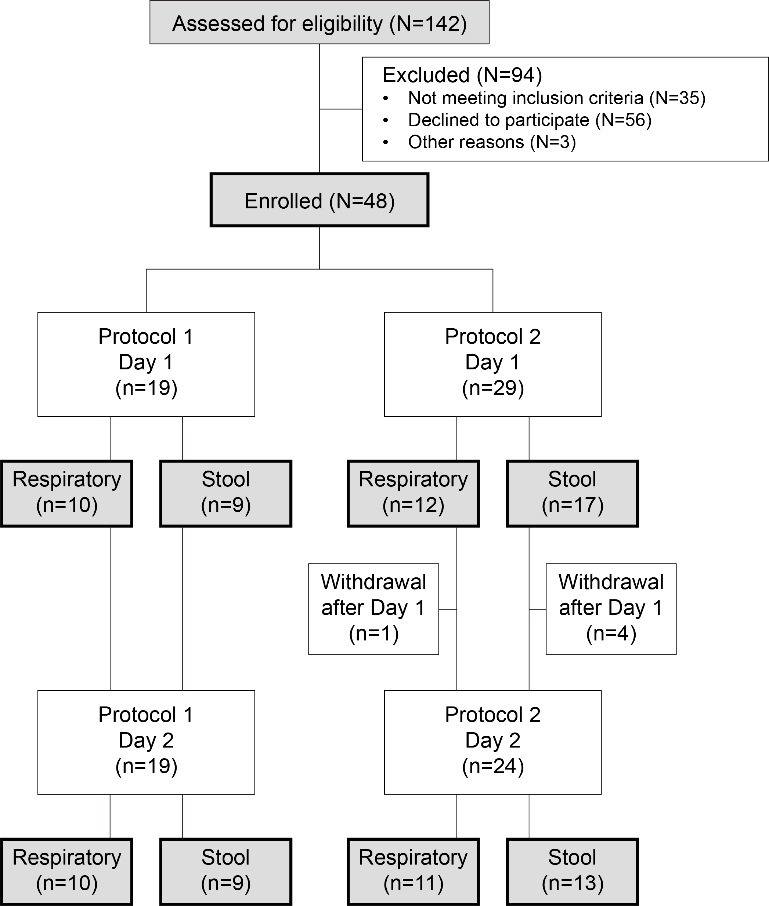

**Fig. S6. A CONSORT flow diagram that shows participant recruitment, eligibility, enrollment, and assignment to the study protocols.** For the exclusion, 35 volunteers were pre-screened by the eligibility evaluation, 56 volunteers declined to participate, and three volunteers were lost to follow up after the evaluation.

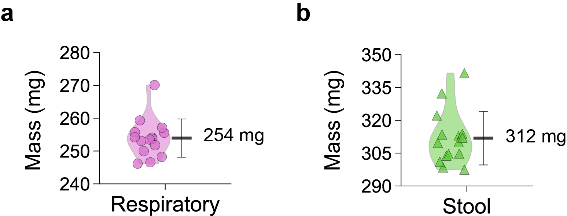

**Fig. S7. Determination of weight for 300 μL of artificial samples. a**, 300 μL of artificial respiratory samples. **b**, 300 μL of artificial stool samples. Horizontal bold lines: mean; vertical lines: standard deviation. The measurements were performed on the laboratory-grade analytical balance.

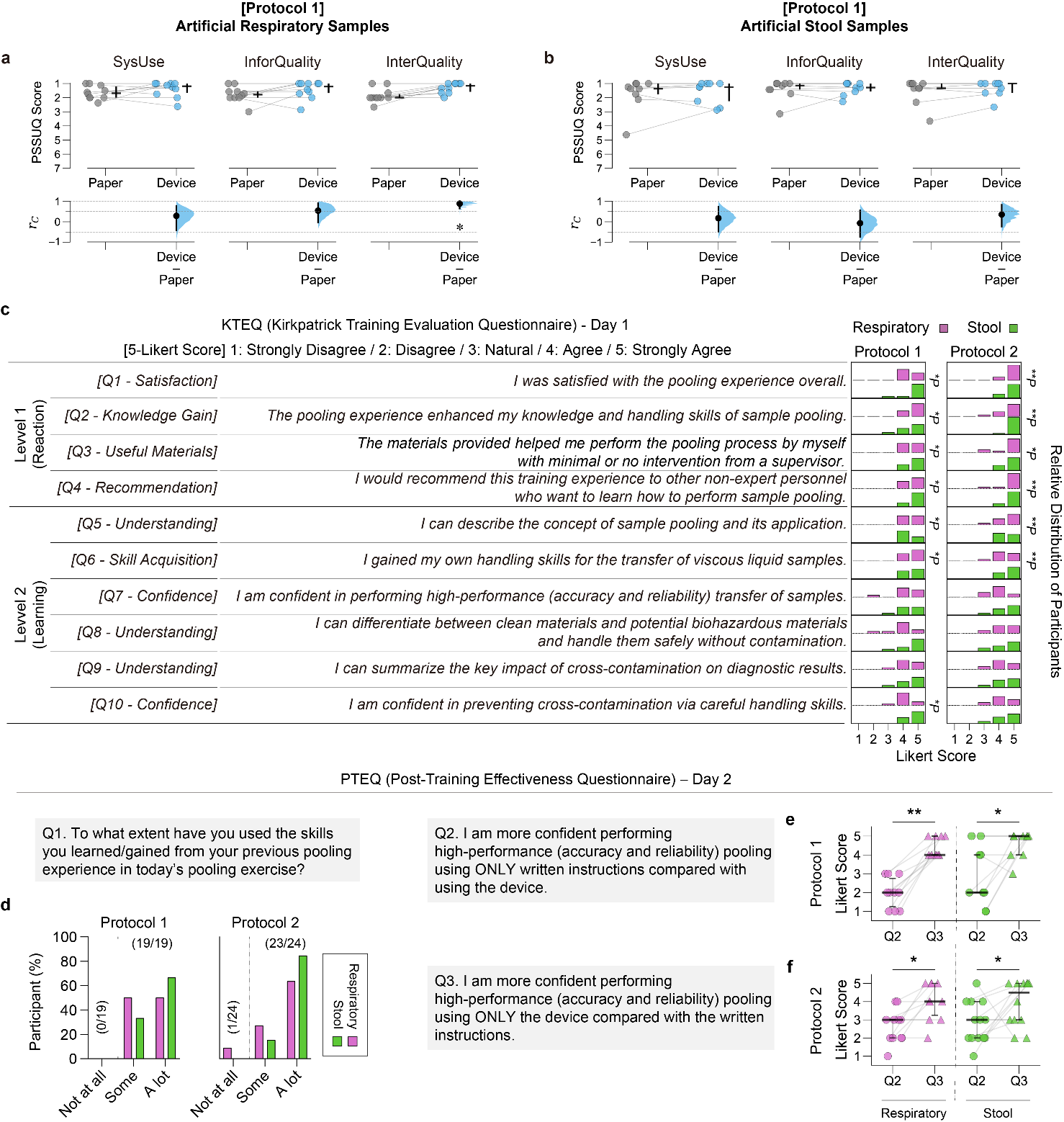

**Fig. S8. Self-reported, post-pooling surveys on usability, training experience and effectiveness.** **a**,**b**, Post-study system usability questionnaire (PSSUQ) scores for system usefulness (SysUse), information quality (InfoQual), and interface quality (InterQual) of Day-1 paper instructions and Day-2 instructional device in Protocol 1 group using artificial respiratory (**a**) and stool samples (**b**). In the upper graphs, horizontal and vertical bold lines represent median and quantile ranges (Q1–Q3), respectively. In the lower graphs, effect sizes *r_C_* (the matched-pairs rank biserial correlation coefficients^50^) were calculated using one-sided Wilcoxon signed-rank tests comparing PSSUQ scores between the paper instructions and the device. The effect sizes were displayed as black dots with 95% confidence intervals (vertical lines) and resampled distributions (blue curves). Statistical significance was extracted from the Wilcoxon tests with the Benjamini-Hochberg correction method with 5% false discovery rate (FDR). **c**, Relative distribution of participants across 5-point Likert scale scores from Kirkpatrick training evaluation questionnaire (KTEQ) surveys. Statistical significance was determined using one-sided binomial tests (with the Benjamini-Hochberg correction with 5% FDR) against the null hypothesis that 75% or fewer participants gave 4+ scores. **d**,**e**,**f**, Day-2 survey results for post-training effectiveness questionnaire (PTEQ) questions #1 (**d**), #2 (**e**), and #3 (**f**). In panels **e** and **f**, statistical significance was determined using one-sided Wilcoxon signed-rank tests (with the Benjamini-Hochberg correction with 5% FDR) comparing participants’ Likert scores between questions #2 and #3. Asterisks in panels **c**, **e**, and **f** indicate significance levels: **P* < 0.05; ***P* < 0.01.

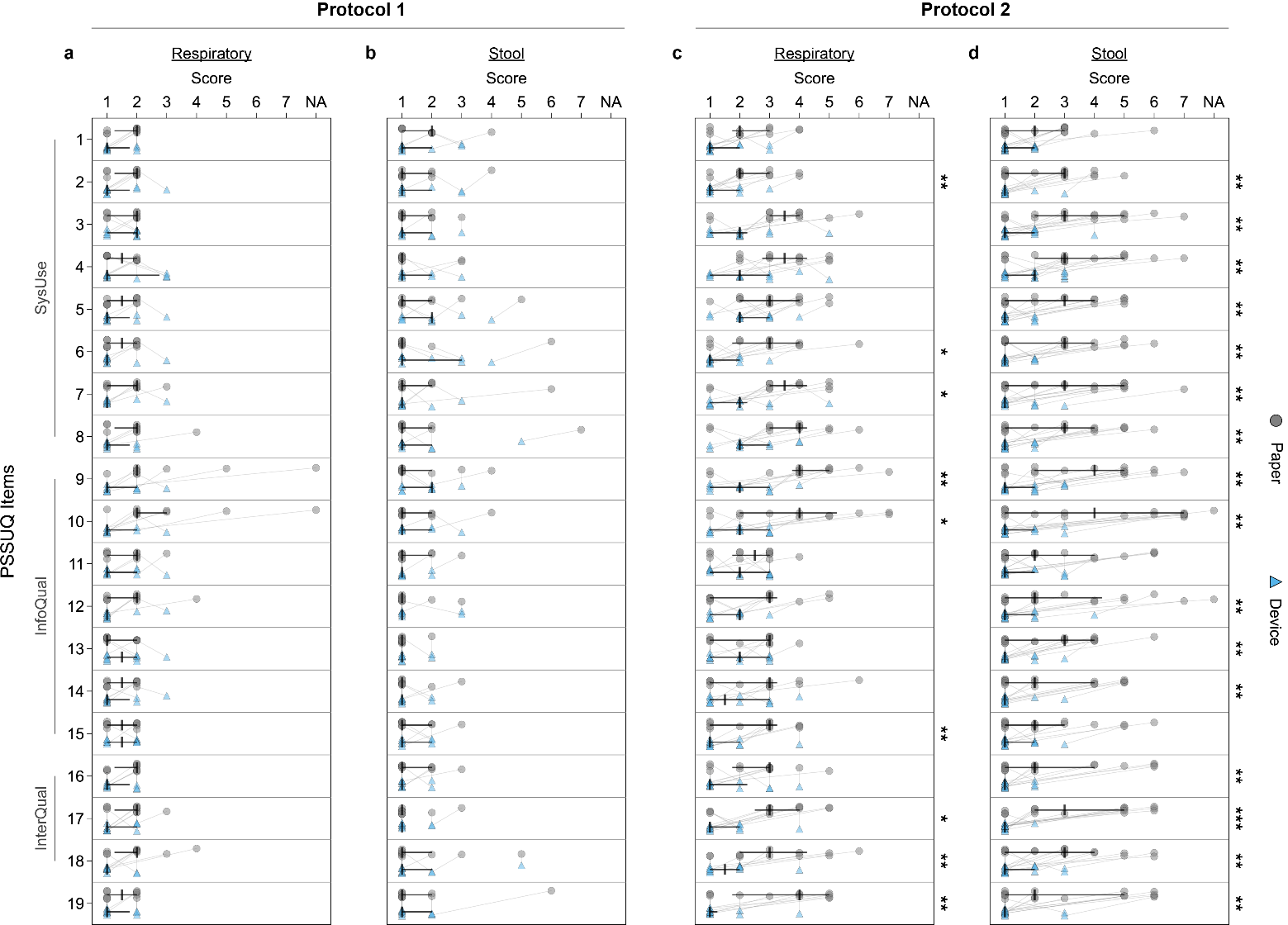

**Fig. S9. Raw PSSUQ score data for 19 questionnaire items. a**,**b**,**c**,**d**, Scores for paper instructions (grey circle) and instructional device (blue triangle) from Protocol 1 (**a**,**b**) and 2 (**c**,**d**) participants using artificial respiratory (**a**,**c**) and stool samples (**b**,**d**). The score follows a 7-point Likert scale, including the ‘not applicable’ (NA) option; lower scores indicate better usability. Vertical and horizontal bold lines represent median and quantile ranges (Q1–Q3), respectively. Statistical significance was extracted from the one-sided Wilcoxon signed-rank tests (with Benjamini-Hochberg correction with 5% false discovery rate) comparing scores for paper instructions and the device. Asterisks indicate significance levels: **P*<0.05; ***P*<0.01; ****P*<0.001; *****P*<0.0001. NA, not applicable; SysUse, system usefulness; InfoQual, information quality; InterQual, interface quality.

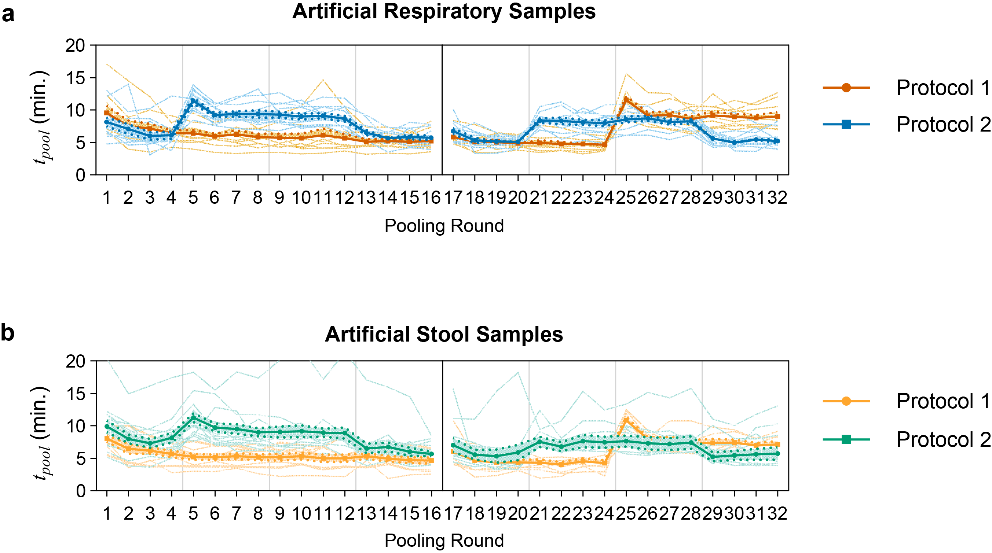

**Fig. S10.** Completion time in each pooling round using artificial respiratory and stool samples for Protocol 1 and 2 groups.

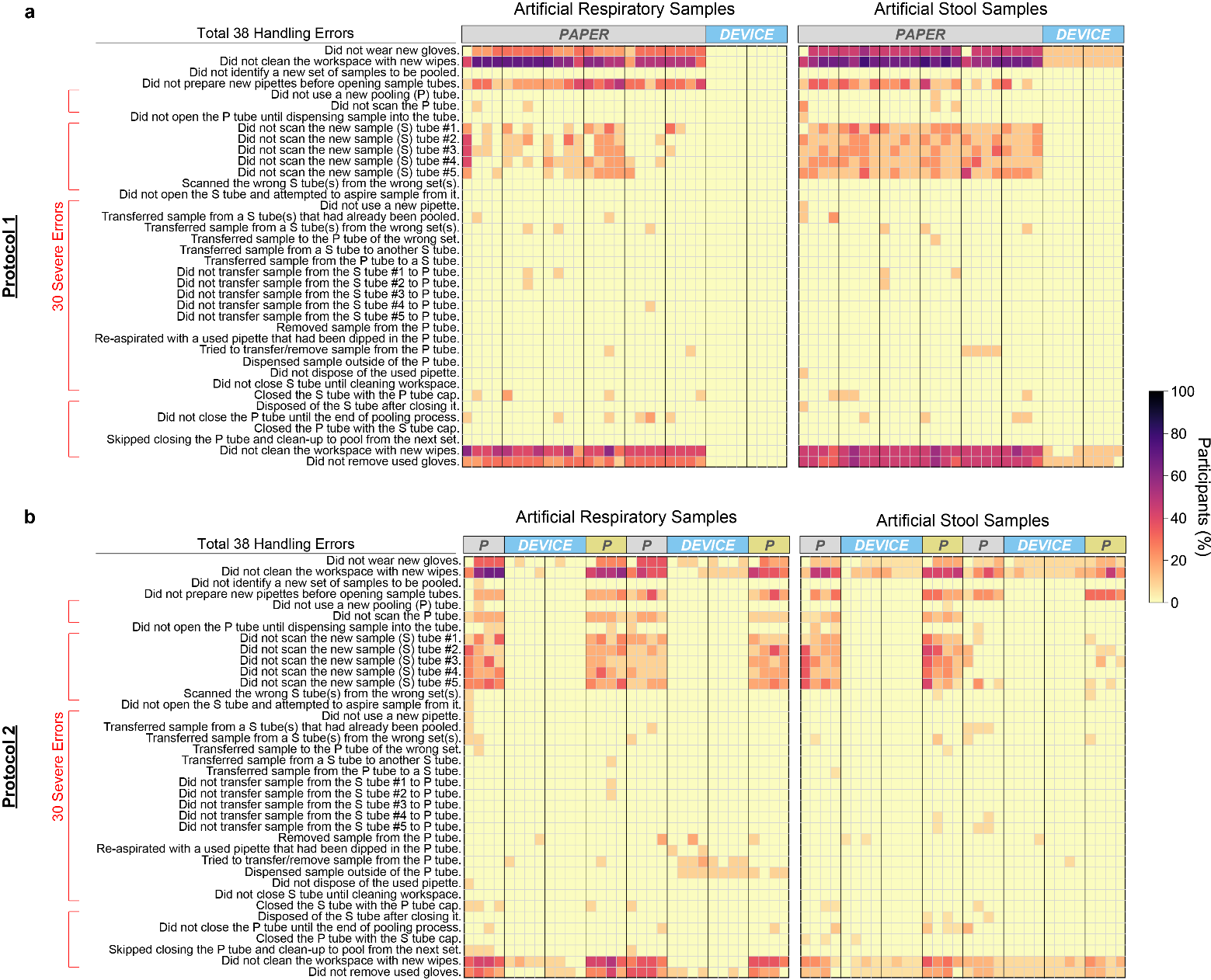

**Fig. S11.** **Uncorrected handling errors of participants during pooling exercises across two training days.** **a**,**b**, Heatmaps displaying the proportion of Protocol 1 (**a**) and Protocol 2 (**b**) participants making handling errors among a total of 38 distinct errors. Data are shown separately for artificial respiratory samples (left panels) and stool samples (right panels). Each row represents a specific handling error, while columns represent pooling rounds (from 1 to 32 across two days per sample type). Color intensity indicates the percentage of participants who made each error (scale bar: 0-100%). Errors are categorized into two groups: severe errors and minor errors (see Supplementary Table 1). Of 38, 30 errors were classified as severe. *P*, paper instructions; *DEVICE*, instructional pooling device.

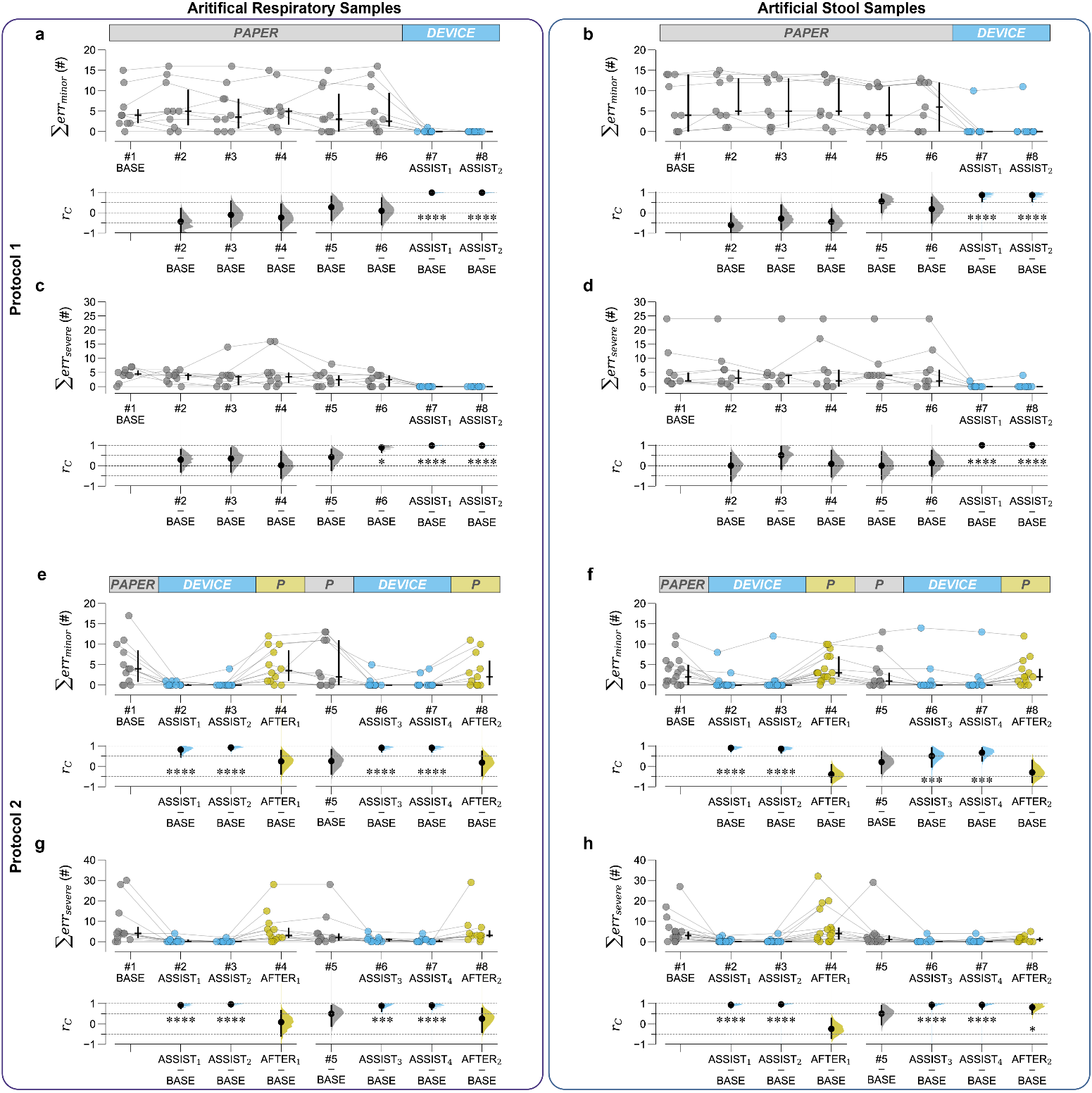

**Fig. S12.** **Assistive effects of instructional pooling device on reducing both minor and severe handling errors in both Protocol 1 and 2 groups for both sample types.** **a**,**b**,**c**,**d**,**e**,**f**,**g**,**h** Gardner-Altman estimation plots comparing the sum of uncorrected minor (**a**,**b**,**e**,**f**) and severe (**c**,**d**,**g**,**h**) errors across pooling sessions in Protocol 1 (**a**,**b**,**c**,**d**) and 2 (**e**,**f**,**g**,**h**) groups. In the upper graphs, horizontal bold lines represent median. Vertical lines represent quartile ranges. Individual data points represent individual participants. In the lower graphs, effect sizes *r_C_* were calculated using one-sided Wilcoxon signed-rank tests comparing the number of uncorrected severe errors of individual participants between the first session (#1, BASE) and each of the other sessions (#2–#8). The effect sizes are displayed as black dots with the 95% C.I. (vertical lines) and resampled distributions of effect sizes (curves), given the observed data. Statistical significance indicated here was extracted from the relevant fixed effects of linear mixed-effect models using dummy coding. Asterisks indicate significance levels: **P*<0.05; ***P*<0.01; ****P*<0.001; *****P*<0.0001.

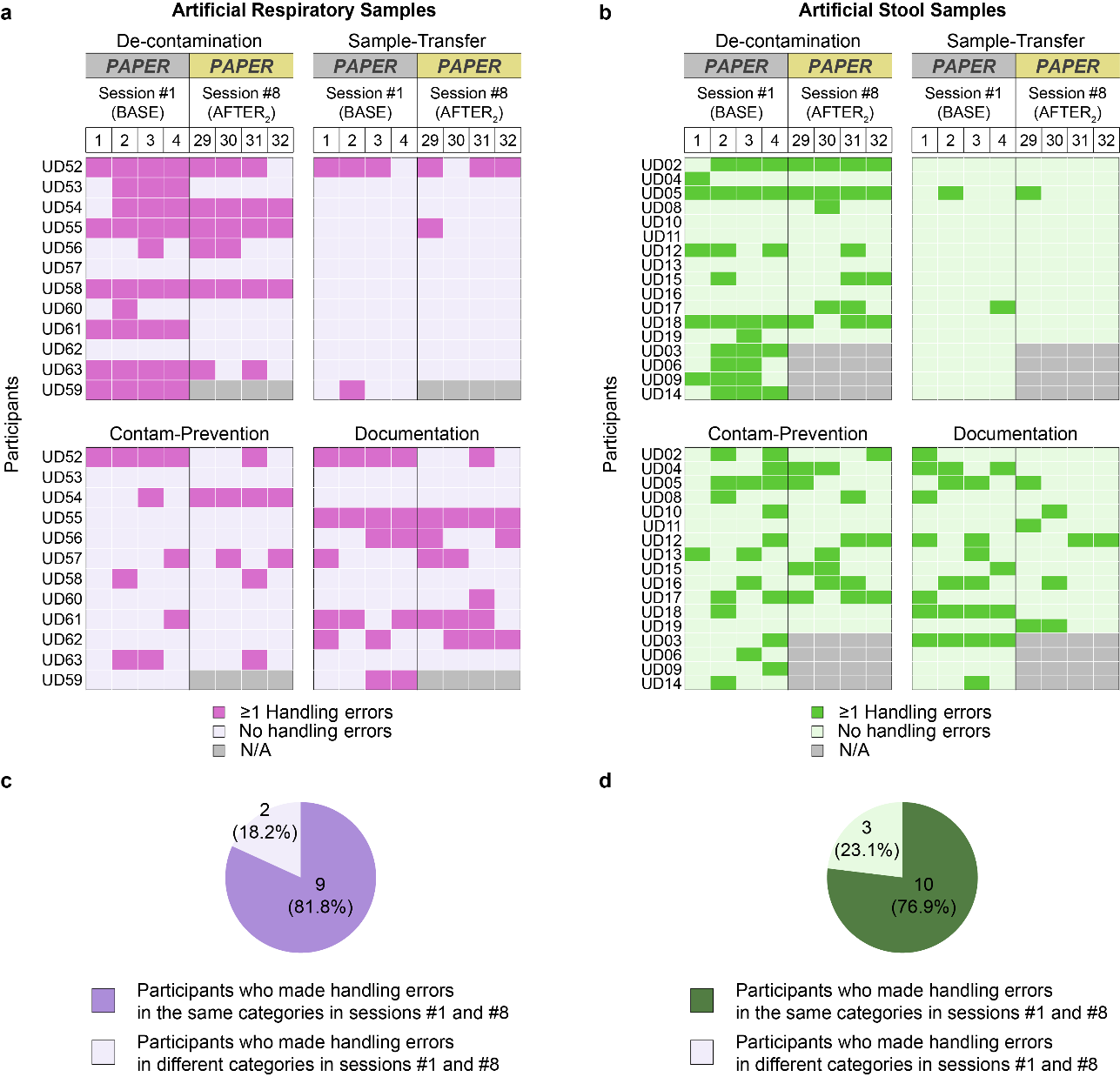

**Fig. S13.** **Assessment of handling error recurrence in the same categories before (session #1; BASE) and after (session #8; AFTER_2_) the use of instructional pooling device.** **a**,**b** Heatmaps displaying handling errors (≥1 errors; dark pink for respiratory samples in the panel **a** and dark green for stool samples in the panel **b**) versus no handling errors (shown in light pink and light green in the panels **a** and **b**, respectively) for individual participants across four categories: De-contamination, Sample-Transfer, Contamination-Prevention, and Documentation (see Supplementary Table 2). Grey cells indicate data not available (N/A) due to participant withdrawal on Day 2. Each column represents a single pooling round in sessions #1 and #8. **c**,**d**, Pie charts showing the proportion of participants who made handling errors in the same versus different categories between sessions #1 and #8 for respiratory samples (**c**, purple) and stool samples (**d**, green). For respiratory samples, 81.8% (9/11) of participants made errors in the same categories. Similarly, for stool samples, 76.9% (10/13) of participants made errors in the same categories.

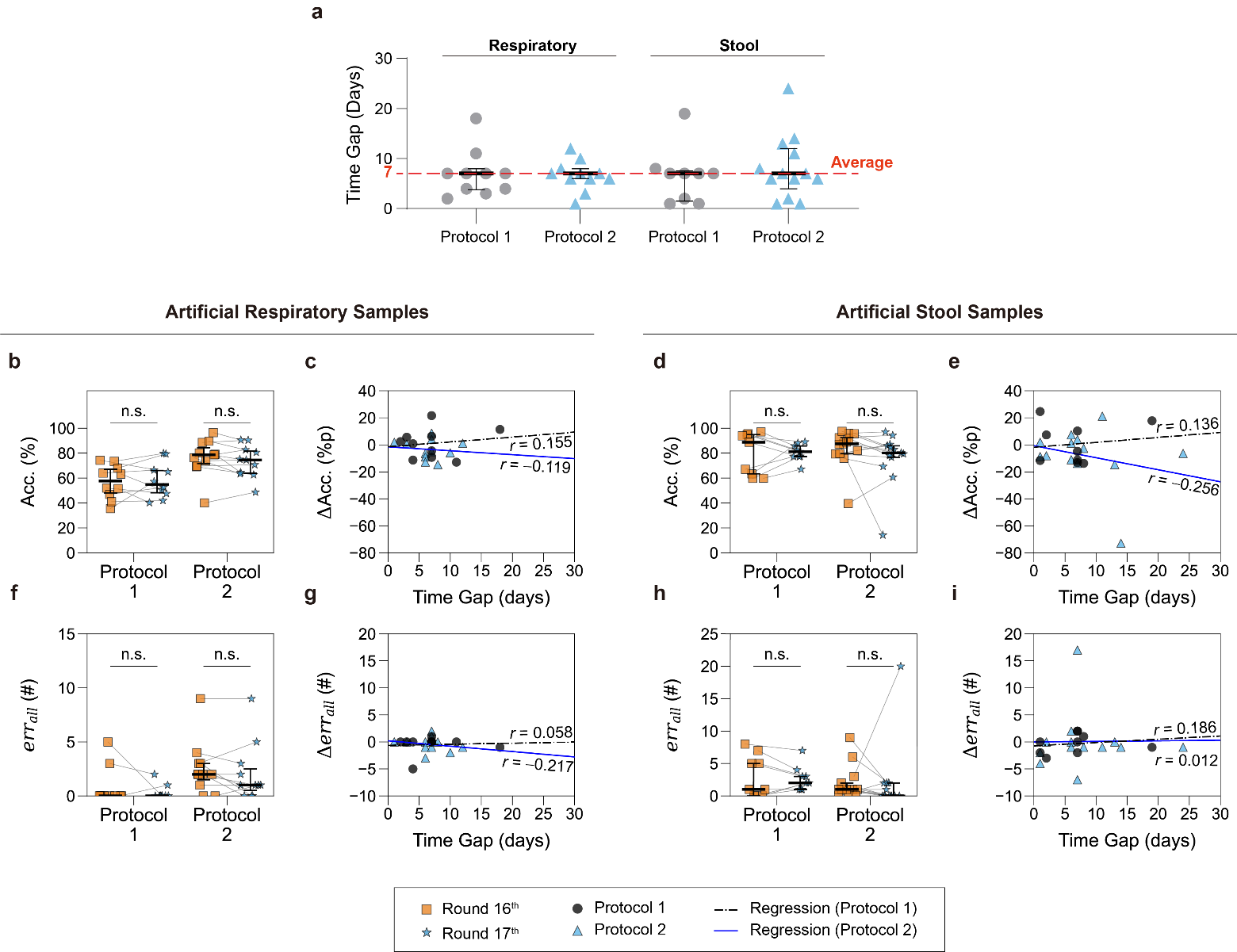

**Fig. S14. Impact of time gaps between experimental Day 1 and Day 2 on participants' handling performance metrics (volume-transfer accuracy and the number of all uncorrected handling errors) comparing the final pooling round on Day 1 (i.e., Round 16^th^) and initial round on Day 2 (i.e., Round 17^th^).** **a**, Distribution of time gaps between Day 1 and Day 2 for Protocol 1 (grey circles) and Protocol 2 (blue triangles) participants using artificial respiratory and stool samples. **b**,**d**,**f**,**h**, Comparison of performance metrics (accuracy, Acc., in **b**,**d**; the number of uncorrected handling errors, *err*_all_, in **f**,**h**) between Round 16^th^ (orange squares) and Round 17^th^ (blue stars) for Protocol 1 and Protocol 2 participants using artificial respiratory samples (**b**,**f**) and stool samples (**d**,**h**). One-sided Wilcoxon signed-rank tests (with the Benjamini-Hochberg correction method with 5% false discovery rate) with were used for the statistical analysis. Null hypothesis for Acc. data: the distribution of differences between paired Acc. values (Round 16^th^ – Round 17^th^) is symmetric around zero or has a negative shift. Null hypothesis for *err*_all_ data: the distribution of differences between paired *err*_all_ counts (Round 16^th^ – Round 17^th^) is symmetric around zero or has a positive shift. n.s.: not significant (*P*>0.05). **c**,**e**,**g**,**i**, Correlation analysis showing ΔAcc. (**c**,**e**) and Δ*err*_all_ (**g**,**i**) against time gaps for Protocol 1 (black circles) and Protocol 2 (blue triangles) participants. ΔAcc. represents differences in accuracy between Rounds 16^th^ and 17^th^. Δ*err*_all_. indicates differences in the number of all uncorrected handling errors between Rounds 16^th^ and 17^th^. n.s., not statistically significant.

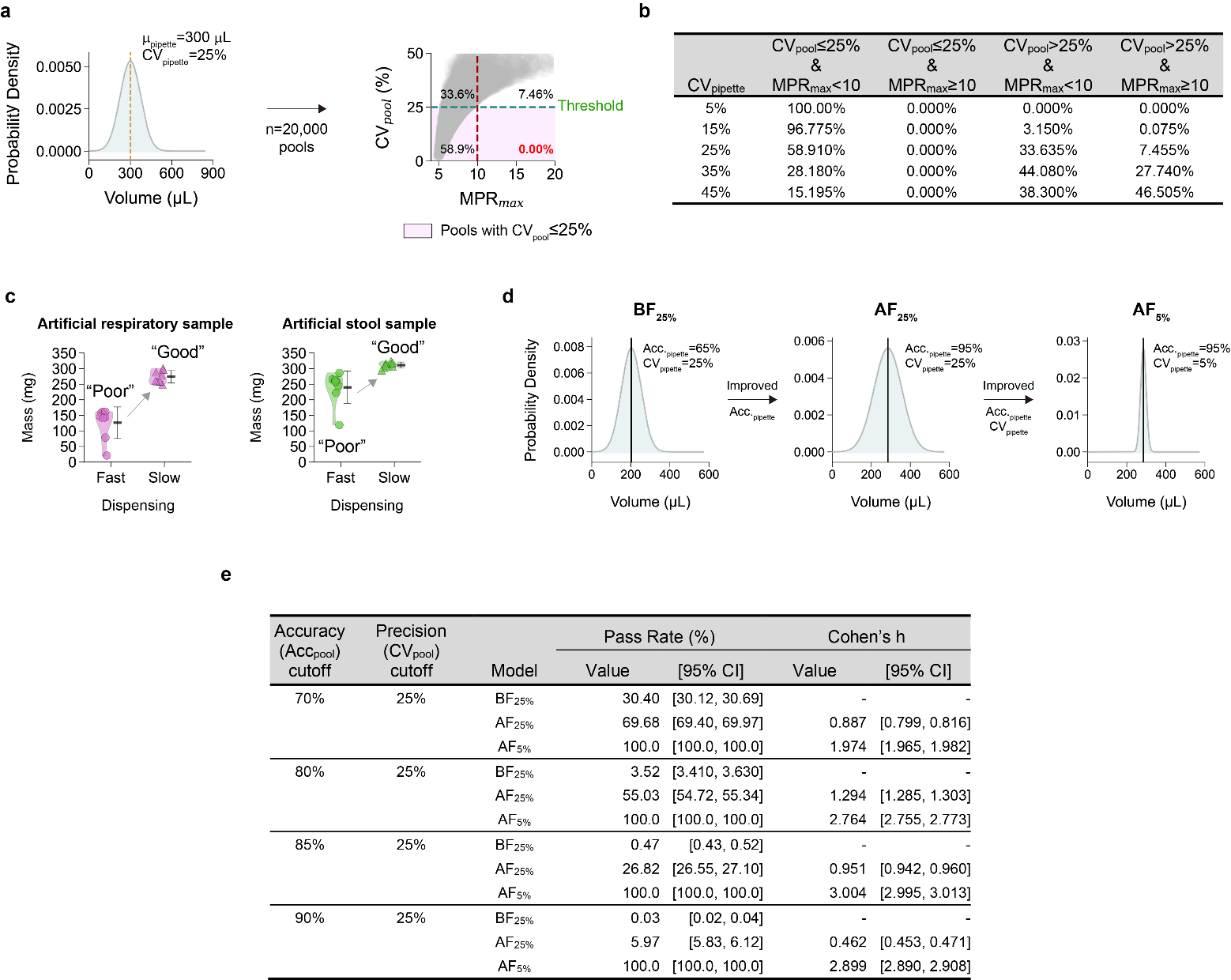

**Fig. S15. Determination of Acc_pool_ and CV_pool_ thresholds through Monte Carlo simulations. a**, An example of simulation of CV_pool_ vs. MPR_max_ from 20,000 simulated pools (via Monte Carlo method) from a normal distribution for pipetting performance of μ_pipette_=300 μL and CV_pipette_=25%. All simulated pools with CV_pool_≤25% (highlighted in purple) show MPR_max_<10. **b**, Proportions of simulated pools (%) meeting different CV_pool_ and MPR_max_ criteria across pipetting precision (CV_pipette_) levels. Each row represents the categorization of 20,000 simulated pools, where each pool was created by sampling 5 components from a truncated normal distribution. The criterion of CV_pool_≤25% ensures pools with MPR_max_<10. **c**, Mass of artificial respiratory and stool samples (n=8) transferred by dual-bulb pipettes with different dispending techniques. When dispensing was slow and gentle, transferred samples were close to the target volume of 300 μL with low variability, compared to those with fast dispensing. **d**, Three distribution models of pipetting performance of untrained personnel before training and improved personnel after training: pre-training baseline (BF_25%_; Acc_pipette_=67.5% & CV_pipette_=25%), post-training with improved accuracy only (AF_25%_; Acc_pipette_=95% & CV_pipette_=25%), and post-training with improved accuracy and precision (AF_5%_, Acc_pipette_=95% & CV_pipette_=25%). **e**, Simulated results (Monte Carlo method; n=20,000) comparing pass rates and effect sizes across different accuracy thresholds for high-quality pools simulated from three models (BF_25%_, AF_25%_, AF_5%_). Pass rates represent the percentage of simulated pools meeting both the accuracy cutoff and precision requirement. Cohen's h measures the effect size of improvements relative to baseline performance, with 95% confidence intervals shown in brackets. A dash (-) indicates the reference group for effect size calculations. The criteria of Acc_pool_≥80% & CV_pool_≤25% can capture the improvements in accuracy and precision, respectively, from the baseline performance.

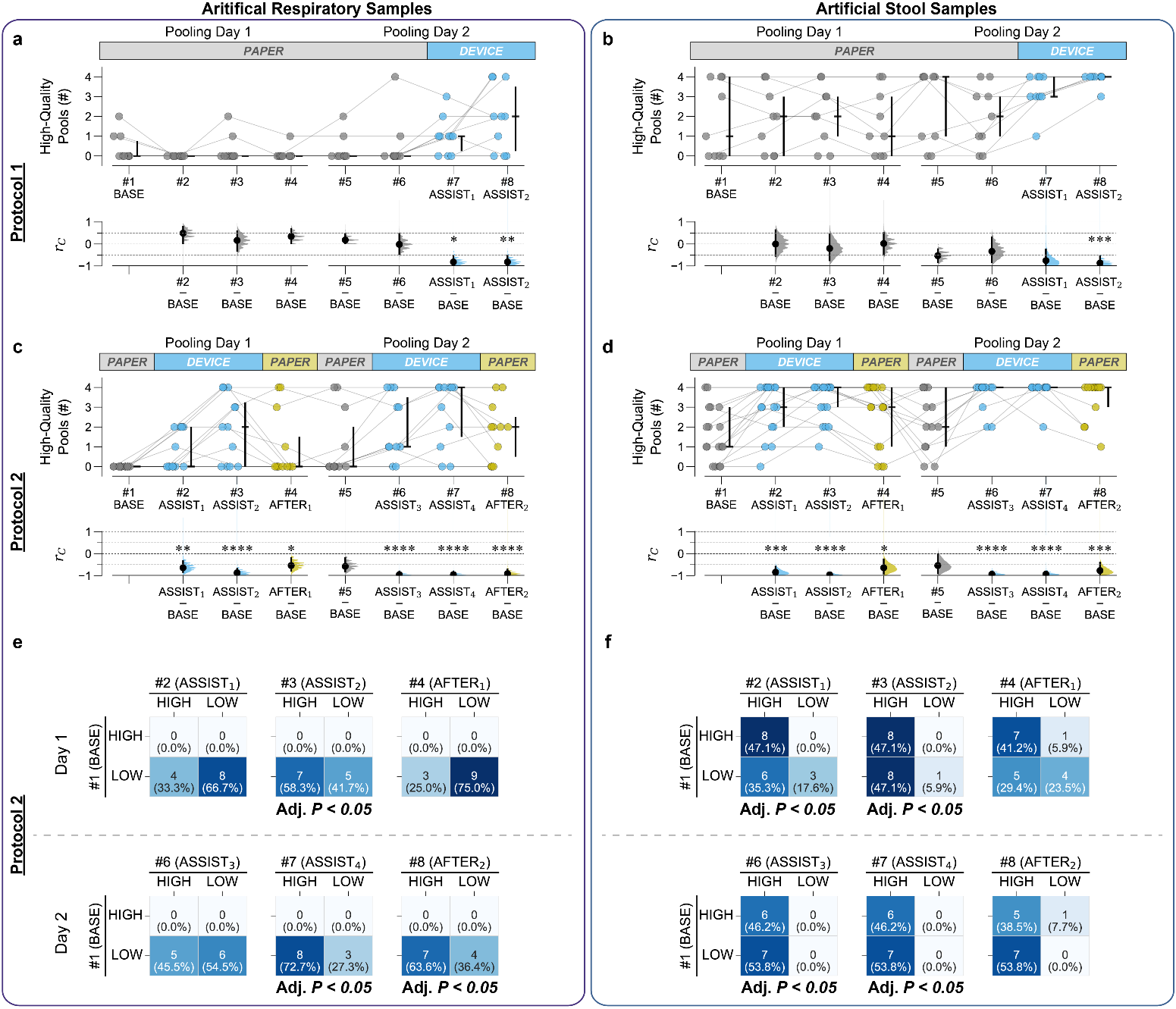

**Fig. S16.** **High-quality pools produced through high-performance volume-transfer, defined as pools with volume-transfer accuracy (Acc_pool_) ≥80% and precision ≤25% CV_pool_. a**,**b**,**c**,**d**, Gardner-Altman estimation plots comparing the number of high-quality pools produced between the first pooling session (#1, BASE) and subsequent sessions (#2–#8) in Protocol 1 (**a**,**b**) and Protocol 2 groups (**c**,**d**). Upper graphs: Data points with median (horizontal bold lines) and quartile ranges (vertical lines) for individual participants. Lower graphs: Effect sizes *r_C_* (black dots) from one-sided Wilcoxon signed-rank tests with 95% CI (vertical lines) and resampled distributions (curves) of effect sizes. Statistical significance levels: **P* < 0.05; ***P* < 0.01; ****P* < 0.001; *****P* < 0.0001 (extracted from results of linear mixed-effect models using dummy coding). **e**,**f**, 2x2 contingency tables for McNemar’s tests comparing the number of high-quality pools of Protocol 2 participants between the first pooling session #1 (BASE) and one of sessions #2–#8 separated by (“HIGH”) and low (“LOW”) performers. Color intensity (light to dark blue) highlights the relative distribution of participants within each individual 2×2 table, with darker blue indicating cells containing higher percentages. Note that each table has its own independent color scale. Cell values: Number of participants in each performance category combination. Cell values: Number of participants in each performance category combination. HIGH, high performers who produced ≥2 high-quality pools during a 4-round session. LOW, low performers who produced <2 high-quality pools during a 4-round session. *P* values for statistical significance were determined by McNemar’s tests with Benjamini-Hochberg correction with 5% false discovery rate.

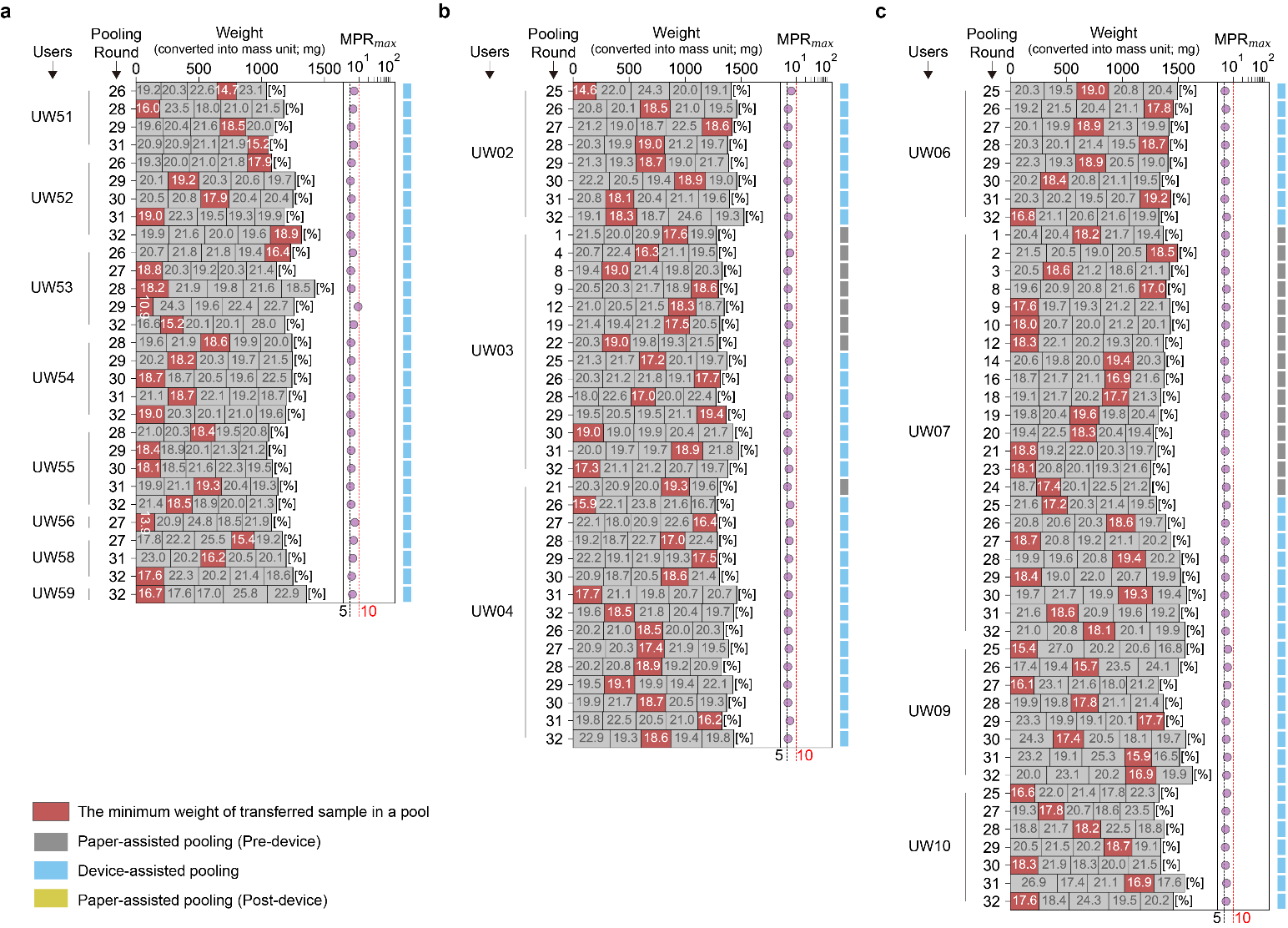

**Fig. S17. High-quality pools through high-performance sample pooling (Acc_pool_≥80%, CV_pool_≤25%, *err*_all_=0) from Protocol 1 participants. a**,**b**,**c**, Weight distributions of high-quality pools produced by Protocol 1 participants using artificial respiratory (**a**) and stool samples (**b**,**c**). Each horizontal bar represents a pool of individual samples with weight percentages. The segments were stacked in the order of volume transfer from different individual tubes. Red highlights the minimum weight sample within each pool. Maximum mass pooling ratio (MPR_max_), defined as the ratio of total pool weight to the minimum sample weight, is shown for each high-quality pool. On the right side of each graph, dark grey, blue, and dark yellow boxes indicate paper-assisted (pre-device), device-assisted, and paper-assisted (post-device) pooling. Weight data were recorded by the customized data logger and the instructional device during paper-assisted and device-assisted pooling, respectively.

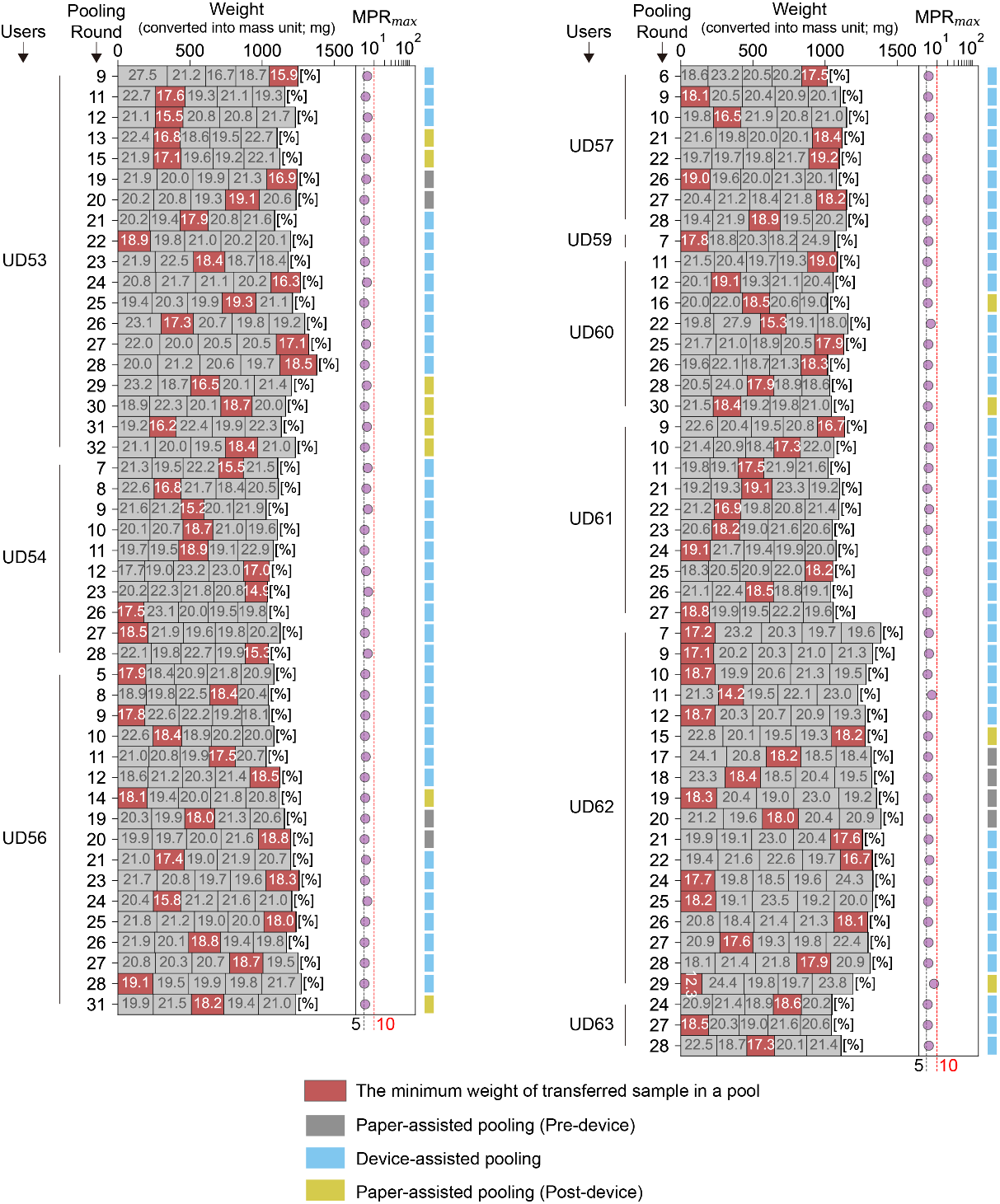

**Fig. S18. High-quality pools through high-performance sample pooling (Acc_pool_≥80%, CV_pool_≤25%, *err*_all_=0) from Protocol 2 participants using artificial respiratory samples.** Stacked bar graphs show weight distributions of high-quality pools produced by Protocol 2 participants using artificial respiratory samples. Each horizontal bar represents a pool of individual samples with weight percentages. The segments were stacked in the order of volume transfer from different individual tubes. Red highlights the minimum weight sample within each pool. Maximum mass pooling ratio (MPR_max_), defined as the ratio of total pool weight to the minimum sample weight, is shown for each high-quality pool. On the right side of each graph, dark grey, blue, and dark yellow boxes indicate paper-assisted (pre-device), device-assisted, and paper-assisted (post-device) pooling. Weight data were recorded by the customized data logger and the instructional device during paper-assisted and device-assisted pooling, respectively.

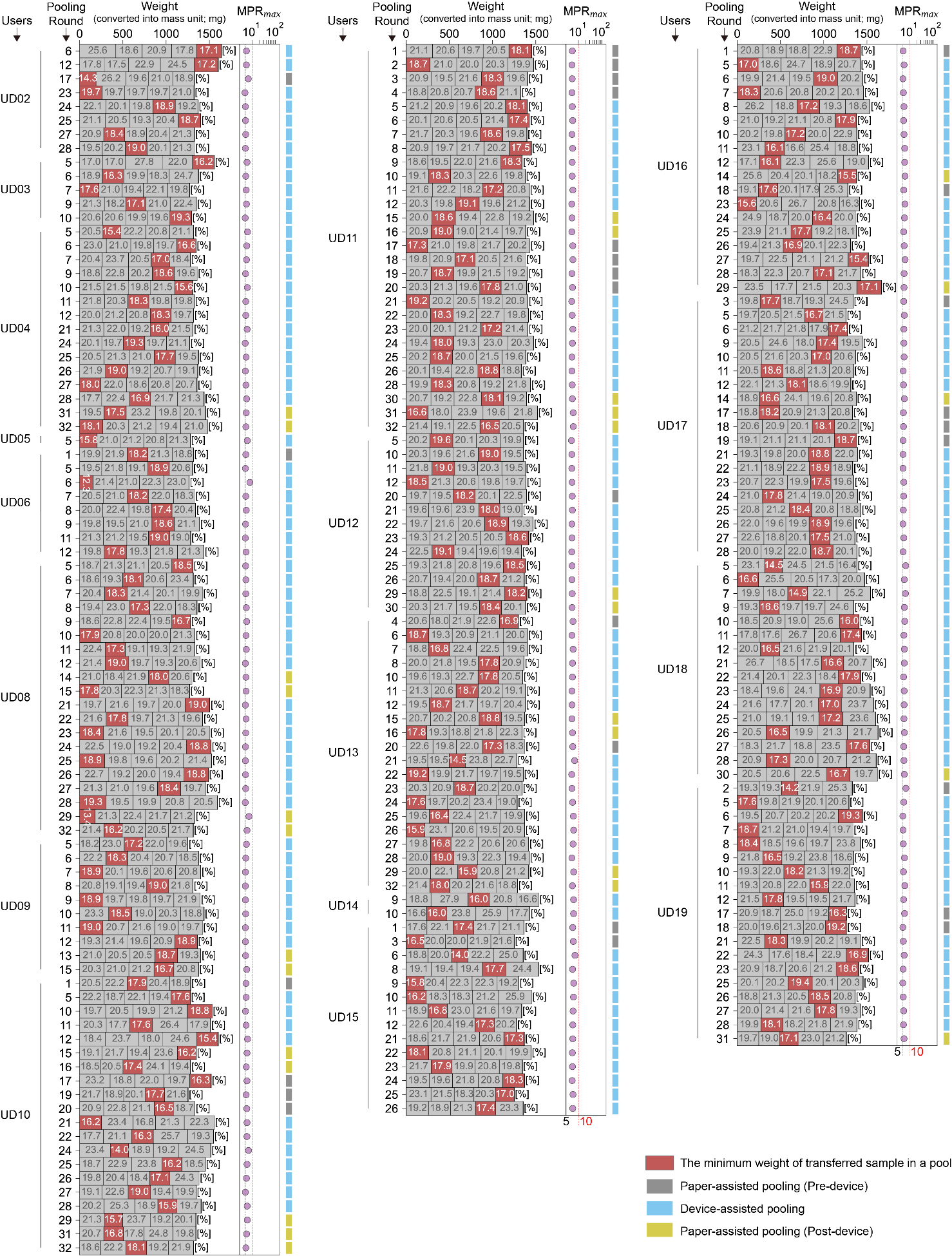

**Fig. S19. High-quality pools through high-performance sample pooling (Acc_pool_≥80%, CV_pool_≤25%, *err*_all_=0) from Protocol 2 participants using artificial stool samples.** Stacked bar graphs show weight distributions of high-quality pools produced by Protocol 2 participants using artificial stool samples. Each horizontal bar represents a pool of individual samples with weight percentages. The segments were stacked in the order of volume transfer from different individual tubes. Red highlights the minimum weight sample within each pool. Maximum mass pooling ratio (MPR_max_), defined as the ratio of total pool weight to the minimum sample weight, is shown for each high-quality pool. On the right side of each graph, dark grey, blue, and dark yellow boxes indicate paper-assisted (pre-device), device-assisted, and paper-assisted (post-device) pooling. Weight data were recorded by the customized data logger and the instructional device during paper-assisted and device-assisted pooling, respectively.

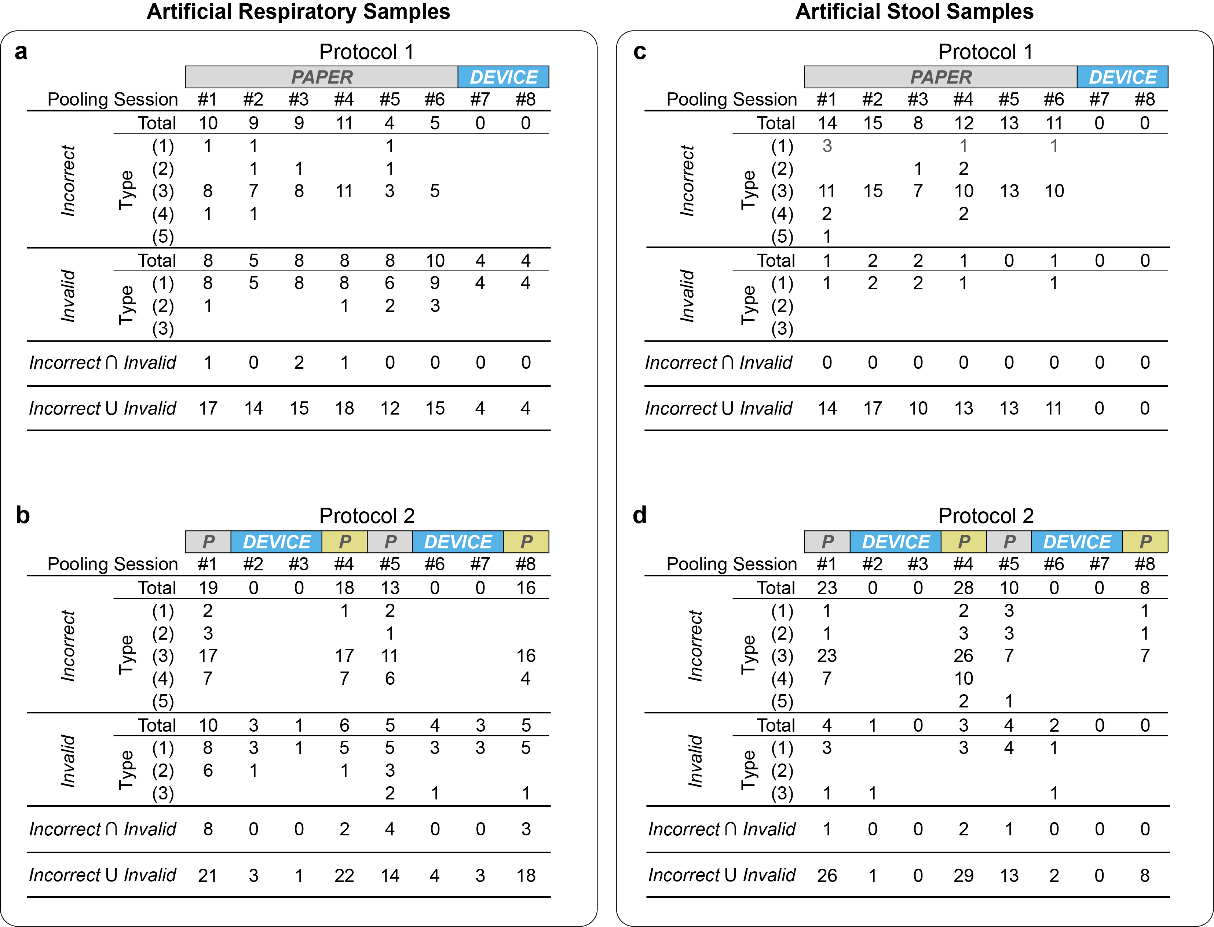

**Fig. S20. The number of *Incorrect* and *Invalid* pools across pooling sessions.** **a**,**b**,**c**,**d**, *Incorrect* and *Invalid* pools produced by Protocol 1 (**a**,**c**) and Protocol 2 (**b**,**d**) participants using artificial respiratory (**a**,**b**) and stool samples (**c**,**d**). *Incorrect* and *Invalid* indicate the total number of pools which meet the requirements for *Incorrect* pools and *Invalid* pools, respectively. Individual pools could be classified under multiple types and simultaneously categorized as both *Incorrect* and *Invalid (*indicated as *Incorrect* ∩ *Invalid*). ‘*Incorrect* U *Invalid*’ indicates the total number of unique pools that are either *Incorrect* or *Invalid* or both, thus can be equal to or lower than the simple sum of *Incorrect* and *Invalid* pools due to overlap (*Incorrect* ∩ *Invalid*).

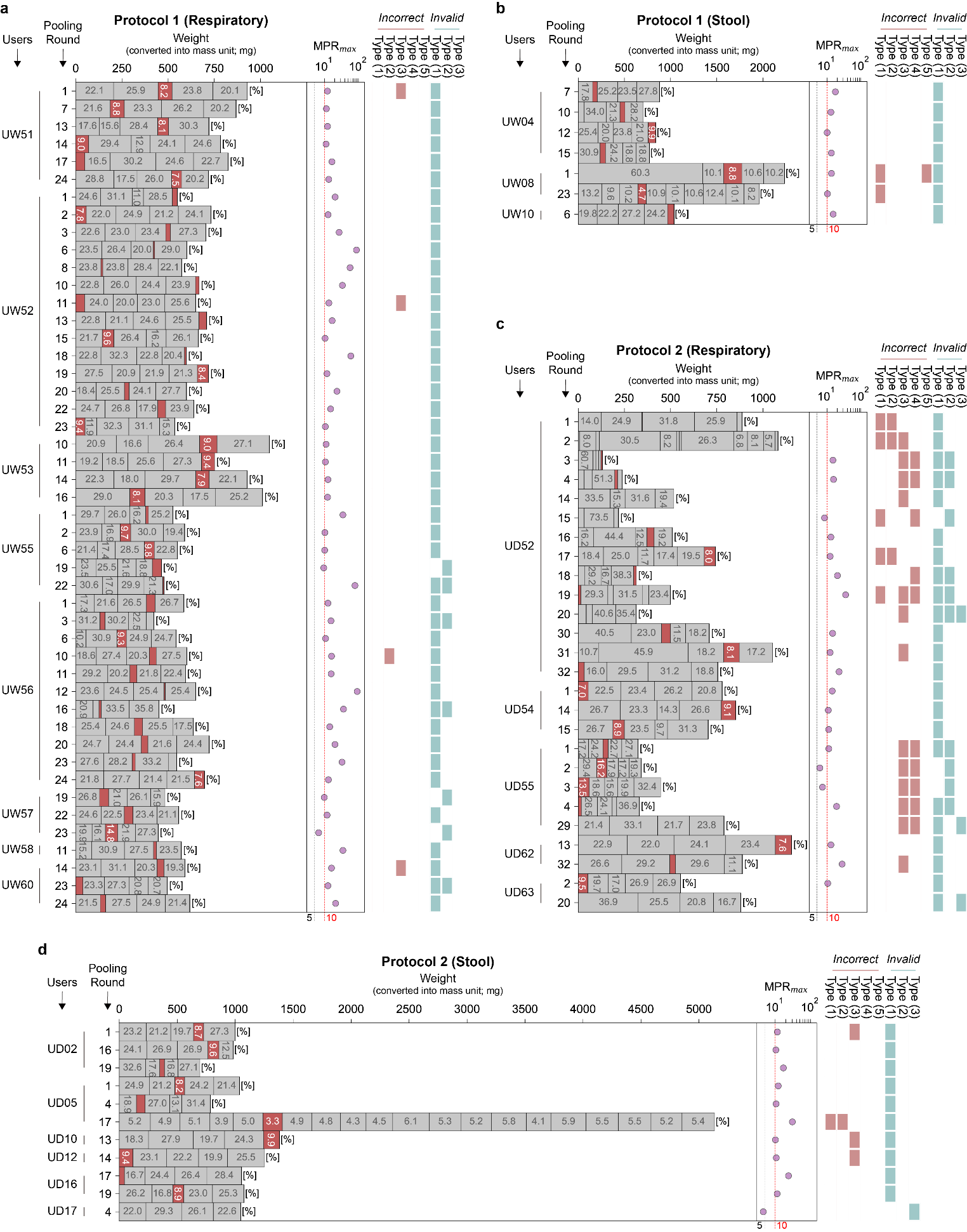

**Fig. S21. *Invalid* pools produced by both Protocol 1 and 2 groups during paper-assisted pooling. a**,**b**,**c**,**d**, Weight distributions of *Invalid* pools produced by Protocol 1 (**a**,**b**) and Protocol 2 (**c**,**d**) groups using artificial respiratory (**a**,**c**) and stool samples (**b**,**d**). Each horizontal bar represents a pool of individual samples with weight percentages. The segments were stacked in the order of volume transfer from different individual tubes. Red highlights the minimum weight sample within each pool. Maximum mass pooling ratio (MPR_max_), defined as the ratio of total pool weight to the minimum sample weight, is shown for each *Invalid* pool. Pink and turquoise boxes indicate type classifications of *Incorrect* (Types 1–5) and *Invalid* (Types 1–3) pools. All weight data were recorded by the customized data logger prepared for the user study.

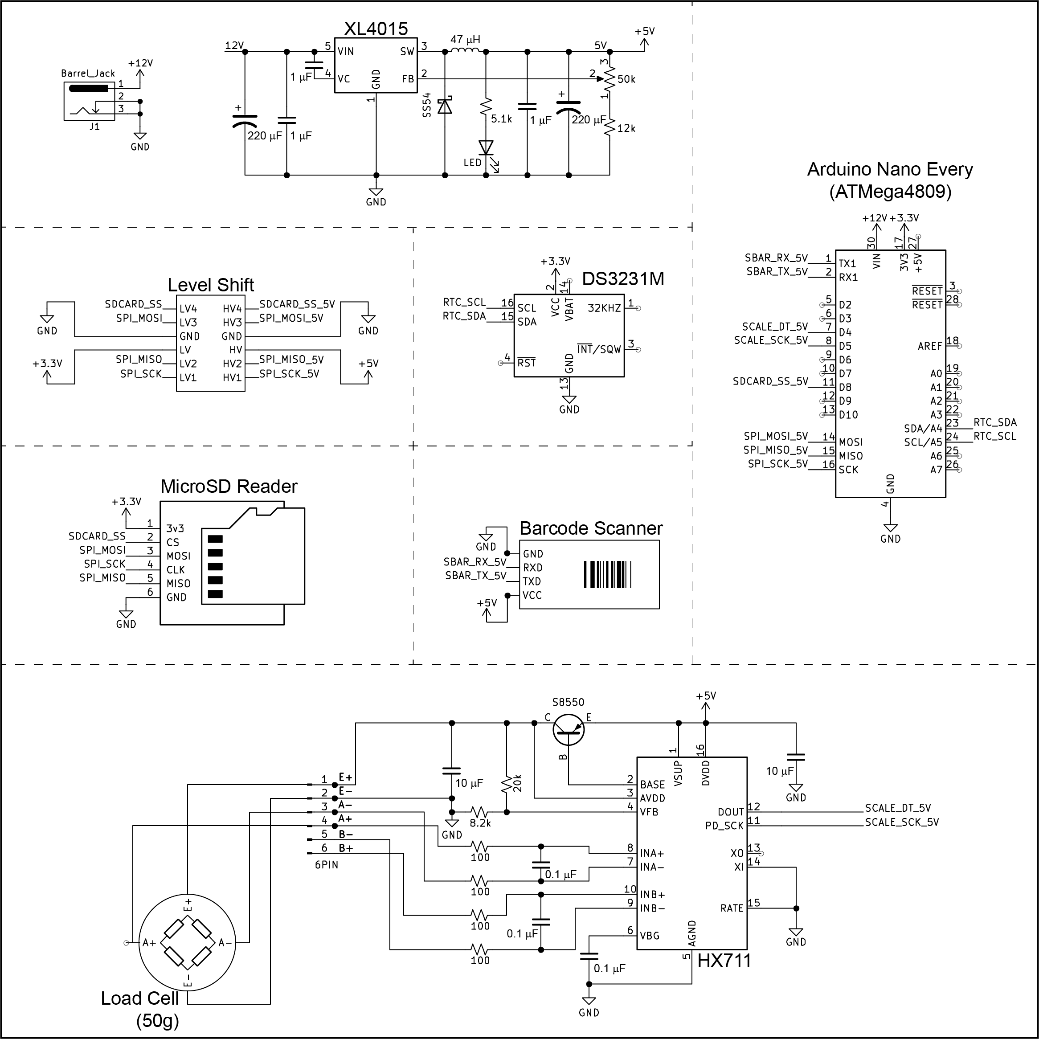

**Fig. S22.** Circuit schematics of customized data logger used in paper-assisted pooling.

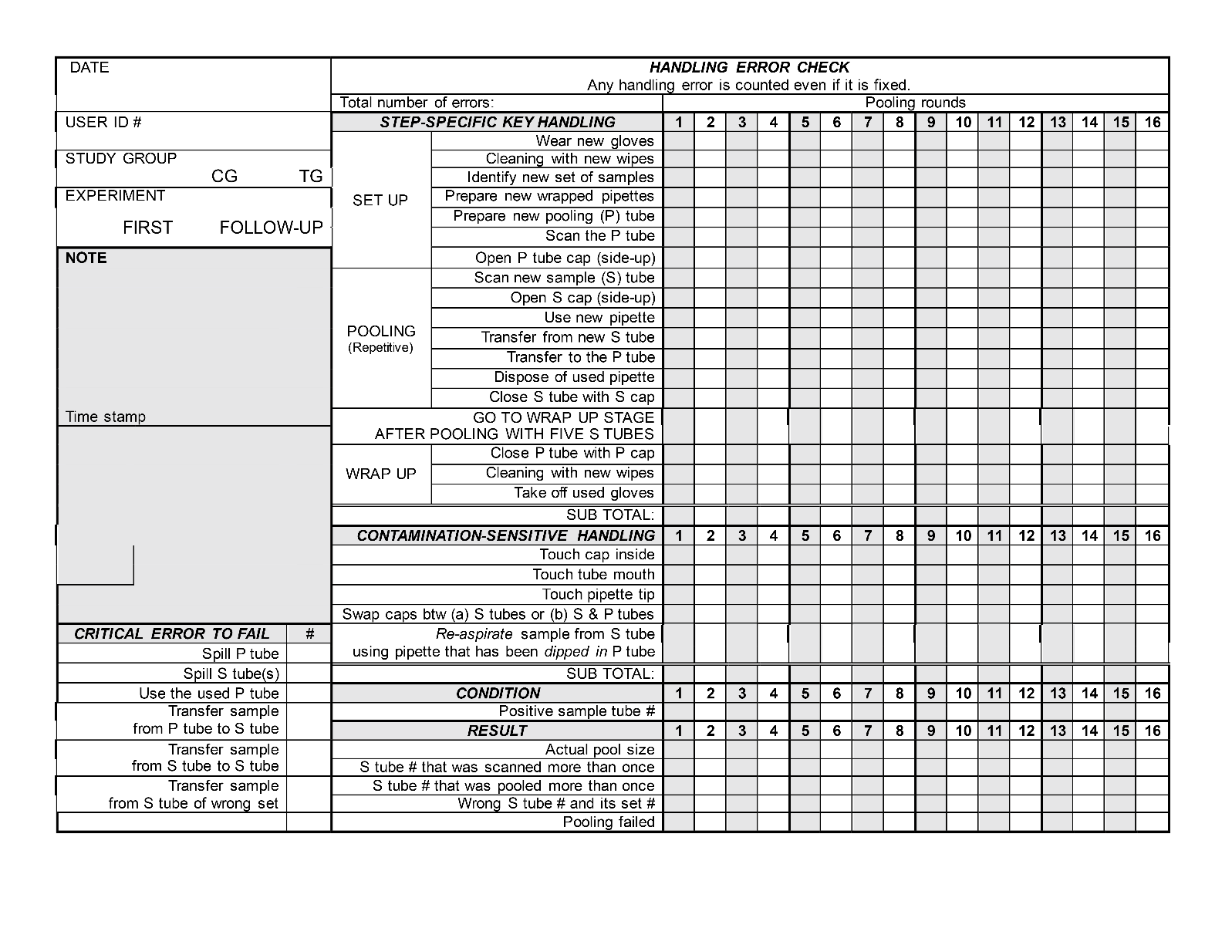

**Fig. S23.** Handling error check sheet used to assess user’s handling errors during the pooling.

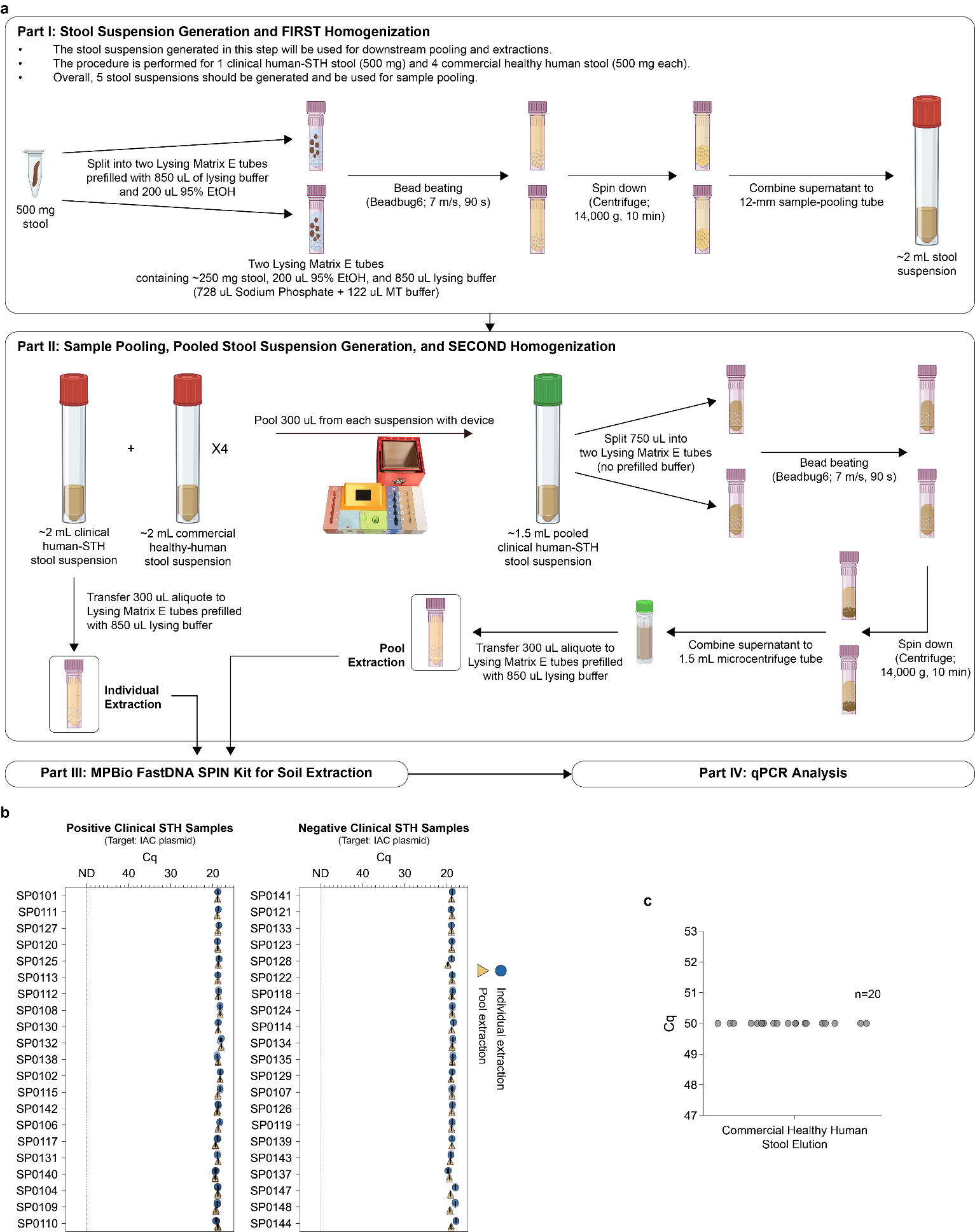

**Fig. S24. Sample processing, extraction control, and limit of blank for 5-sample pooled qPCR testing in the device validation study. a**, Sample processing for 5-sample pooled qPCR testing using clinical human stool samples and commercial healthy human stool samples. **b**, Cq data for internal amplification control (IAC) plasmid spiked into each lysate just before binding the DNA in the lysate to the binding matrix during each individual and pool extraction. **c**, Limit of blank for qPCR analysis (target: Ascaris germline repeat) from human stool elution without *Ascaris lumbricoides* DNA.

**Table S1. Summary demographic and prior training data for participants in the user study.** HS, high school; AP, advanced placement.

|  | **Protocol 1** | | **Protocol 2** | |
| --- | --- | --- | --- | --- |
|  | Respiratory | Stool | Respiratory | Stool |
| **Total Participants** | 10 | 9 | 12 | 17 |
| Day 1 | 10 (100%) | 9 (100%) | 12 (100%) | 17 (100%) |
| Day 2 | 10 (100%) | 9 (100%) | 11 (91.6%) | 13 (76.5%) |
| Withdrew Before Study End | 0 (0.00%) | 0 (0.00%) | 1 (8.33%) | 4 (23.5%) |
| **Age Range** | | | | |
| 18-25 | 7 (70.0%) | 7 (77.8%) | 6 (50.0%) | 10 (58.8%) |
| 26-45 | 2 (20.0%) | 0 (0.00%) | 5 (41.7%) | 3 (17.6%) |
| 46-65 | 1 (10.0%) | 2 (22.2%) | 1 (8.33%) | 4 (23.5%) |
| 66+ | 0 (0.00%) | 0 (0.00%) | 0 (0.00%) | 0 (0.00%) |
| **Sex** | | | | |
| Male | 5 (50.0%) | 4 (44.4%) | 7 (58.3%) | 8 (47.1%) |
| Female | 5 (50.0%) | 5 (55.6%) | 5 (41.7%) | 9 (52.9%) |
| **Highest Education** |  |  |  |  |
| HS degree (or equivalent) | 5 (50.0%) | 3 (33.3%) | 4 (33.3%) | 5 (29.4%) |
| Some college / no degree | 3 (30.0%) | 3 (33.3%) | 2 (16.7%) | 4 (23.5%) |
| Bachelor's degree | 0 (0.00%) | 1 (11.1%) | 5 (41.7%) | 5 (29.4%) |
| Master’s/Doctoral/Professional degree | 2 (20.0%) | 2 (22.2%) | 1 (8.3%) | 3 (17.6%) |
| **Self-Described Level of Wet Lab Experience** | | | | |
| No laboratory experience | 4 (40.0%) | 6 (66.7%) | 3 (25.0%) | 4 (23.5%) |
| HS courses in biology/chemistry laboratory (not AP) | 4 (40.0%) | 2 (22.2%) | 7 (58.3%) | 7 (41.2%) |
| AP HS laboratory courses in biology/chemistry | 1 (10.0%) | 0 (0.00%) | 0 (0.00%) | 0 (0.00%) |
| Community college courses in biology/chemistry | 1 (10.0%) | 1 (11.1%) | 1 (8.33%) | 3 (17.6%) |
| University (BSc program) courses in biology/chemistry | 0 (0.00%) | 0 (0.00%) | 0 (0.00%) | 2 (11.8%) |
| Life Sciences (biology, chemistry, physics, or similar) major in collage | 0 (0.00%) | 0 (0.00%) | 1 (8.30%) | 1 (5.88%) |
| Research-level experience in a wet laboratory (work outside of a classroom) | 0 (0.00%) | 0 (0.00%) | 0 (0.00%) | 0 (0.00%) |
| Expert/Professional experience in a wet laboratory | 0 (0.00%) | 0 (0.00%) | 0 (0.00%) | 0 (0.00%) |
| **Self-Described Level of Relevant Experiences (Multiple Choices)** | | | | |
| I have previously used a pipette. | 7 (70.0%) | 6 (66.7%) | 6 (50.0%) | 10 (58.8%) |
| I have an experience in handling biological or clinical samples in high school. | 4 (40.0%) | 0 (0.00%) | 4 (33.3%) | 8 (47.1%) |
| I am familiar with laboratory biosafety guidelines because I have a basic experience in high school. | 3 (30.0%) | 3 (33.3%) | 1 (8.33%) | 3 (17.6%) |
| I have experience following detailed step-by-step instructions (for work or a hobby). | 0 (0.00%) | 1 (11.1%) | 4 (33.3%) | 6 (35.3%) |
| I am familiar with the concept of sample pooling. | 0 (0.00%) | 0 (0.00%) | 1 (8.33%) | 3 (17.6%) |
| None of these apply to me. | 1 (10.0%) | 4 (44.4%) | 2 (16.7%) | 3 (17.6%) |

**Table S2. Demographic data for participants of Protocol 1 Group using artificial respiratory samples.** HS, high school; AP, advanced placement.

| **Protocol 1 Group (Artificial Respiratory Samples)** | | | | | |
| --- | --- | --- | --- | --- | --- |
| User  Code | Age Range | Sex | Highest Grade of Education Completed | Self-described Level of Wet Laboratory Experience | Relevant Experiences |
| UW51 | 46-65 | Female | Master's/Doctoral/  Professional degree | No laboratory experience | Experience following detailed step-by-step instructions (for work or a hobby) |
| UW52 | 18-25 | Male | Some college/  no degree | No laboratory experience | None |
| UW53 | 26-45 | Male | Master's/Doctoral/  Professional degree | Community college courses  in a biology or chemistry laboratory | Experience following detailed step-by-step instructions (for work or a hobby) |
| UW54 | 18-25 | Male | HS degree  (or equivalent) | HS courses in a biology or chemistry laboratory  (but not AP) | Previously used a pipette |
| UW55 | 26-45 | Female | Some college/  no degree | No laboratory experience | Previously used a pipette |
| UW56 | 18-25 | Female | HS degree  (or equivalent) | AP HS laboratory courses in biology or chemistry | Previously used a pipette |
| UW57 | 18-25 | Female | HS degree  (or equivalent) | No laboratory experience | Previously used a pipette |
| UW58 | 18-25 | Male | Some college/  no degree | HS courses in a biology or chemistry laboratory  (but not AP) | Previously used a pipette;  Experience in handling biological or clinical samples in HS;  Experience following detailed step-by-step instructions (for work or a hobby) |
| UW59 | 18-25 | Female | HS degree  (or equivalent) | HS courses in a biology or chemistry laboratory  (but not AP) | Previously used a pipette;  Experience following detailed step-by-step instructions (for work or a hobby) |
| UW60 | 18-25 | Male | HS degree  (or equivalent) | HS courses in a biology or chemistry laboratory  (but not AP) | Previously used a pipette |

**Table S3. Demographic data for participants of Protocol 1 Group using artificial stool samples.** HS, high school; AP, advanced placement.

| **Protocol 1 Group (Artificial Stool Samples)** | | | | | |
| --- | --- | --- | --- | --- | --- |
| User  Code | Age Range | Sex | Highest Grade of Education Completed | Self-described Level of Wet Laboratory Experience | Relevant Experiences |
| UW02 | 18-25 | Male | Bachelor's degree | No laboratory experience | None |
| UW03 | 18-25 | Female | Some college/  no degree | HS courses in a biology or chemistry laboratory  (but not AP) | Previously used a pipette;  Experience handling biological or clinical samples in HS;  Familiar with laboratory biosafety guidelines from HS |
| UW04 | 46-65 | Female | Some college/  no degree | No laboratory experience | Previously used a pipette |
| UW05 | 18-25 | Female | HS degree  (or equivalent) | No laboratory experience | Previously used a pipette |
| UW06 | 18-25 | Female | Some college/  no degree | Community college courses  in a biology or chemistry laboratory | Experience handling biological or clinical samples in HS |
| UW07 | 18-25 | Female | Master's/Doctoral/  Professional degree | No laboratory experience | None |
| UW08 | 18-25 | Male | HS degree  (or equivalent) | No laboratory experience | None |
| UW09 | 18-25 | Male | HS degree  (or equivalent) | HS courses in a biology or chemistry laboratory  (but not AP) | Experience handling biological or clinical samples in HS |
| UW10 | 46-65 | Male | Master's/Doctoral/  Professional degree | No laboratory experience | None |

**Table S4. Demographic data for participants of Protocol 2 Group using artificial respiratory samples.** Participants who withdrew after the first day are noted as ‘Day 1 only.’ HS, high school; AP, advanced placement.

| **Protocol 2 Group (Artificial Respiratory Samples)** | | | | | |
| --- | --- | --- | --- | --- | --- |
| User  Code | Age Range | Sex | Highest Grade of Education Completed | Self-described Level of Wet Laboratory Experience | Relevant Experiences |
| UD52 | 18-25 | Male | Some college/  no degree | HS courses in a biology or chemistry laboratory  (but not AP) | Previously used a pipette;  Experience in handling biological or clinical samples in HS;  Familiar with laboratory biosafety guidelines from HS |
| UD53 | 26-45 | Male | Bachelor's degree | University (BSc program) courses  in a biology or chemistry laboratory | Familiar with laboratory biosafety guidelines from HS |
| UD54 | 18-25 | Male | HS degree  (or equivalent) | HS courses in a biology or chemistry laboratory  (but not AP) | None |
| UD55 | 18-25 | Male | HS degree  (or equivalent) | Community college courses  in a biology or chemistry laboratory | Familiar with laboratory biosafety guidelines from HS |
| UD56 | 26-45 | Male | Bachelor's degree | HS courses in a biology or chemistry laboratory  (but not AP) | Previously used a pipette;  Experience following detailed step-by-step instructions (for work or a hobby) |
| UD57 | 26-45 | Female | Bachelor's degree | HS courses in a biology or chemistry laboratory  (but not AP) | Experience following detailed step-by-step instructions (for work or a hobby) |
| UD58 | 18-25 | Female | Bachelor's degree | HS courses in a biology or chemistry laboratory  (but not AP) | Previously used a pipette;  Experience following detailed step-by-step instructions (for work or a hobby)  Familiar with the concept of sample pooling |
| UD59  (Day 1 only) | 18-25 | Male | HS degree  (or equivalent) | No laboratory experience | Previously used a pipette |
| UD60 | 26-45 | Female | Master's/Doctoral/  Professional degree | HS courses in a biology or chemistry laboratory  (but not AP) | Previously used a pipette |
| UD61 | 18-25 | Female | Some college/  no degree | No laboratory experience | Familiar with laboratory biosafety guidelines from HS;  Experience following detailed step-by-step instructions (for work or a hobby) |
| UD62 | 26-45 | Male | Bachelor's degree | HS courses in a biology or chemistry laboratory  (but not AP) | Previously used a pipette |
| UD63 | 46-65 | Female | HS degree  (or equivalent) | No laboratory experience | None |

**Table S5. Demographic data for participants of Protocol 2 Group using artificial stool samples.** Participants who withdrew after the first day are noted as ‘Day 1 only.’ HS, high school; AP, advanced placement.

| **Protocol 2 Group (Artificial Stool Samples)** | | | | | |
| --- | --- | --- | --- | --- | --- |
| User  Code | Age Range | Sex | Highest Grade of Education Completed | Self-described Level of Wet Laboratory Experience | Relevant Experiences |
| UD02 | 46-65 | Male | Bachelor's degree | No laboratory experience | Previously used a pipette;  Experience handling biological or clinical samples in HS;  Familiar with laboratory biosafety guidelines from HS;  Experience following detailed step-by-step instructions (for work or a hobby);  Familiar with the concept of sample pooling |
| UD03 (Day 1 only) | 18-25 | Female | Bachelor's degree | HS courses in a biology or chemistry laboratory  (but not AP) | Previously used a pipette;  Experience handling biological or clinical samples in HS, familiar with laboratory biosafety guidelines from HS, familiar with the concept of sample pooling |
| UD04 | 46-65 | Male | Master's/Doctoral/  Professional degree | HS courses in a biology or chemistry laboratory  (but not AP) | Previously used a pipette |
| UD05 | 18-25 | Female | Bachelor's degree | University (BSc program) courses  in a biology or chemistry laboratory | Previously used a pipette |
| UD06 (Day 1 only) | 18-25 | Male | Some college/  no degree | University (BSc program) courses  in a biology or chemistry laboratory | Familiar with laboratory biosafety guidelines from HS |
| UD08 | 18-25 | Female | HS degree  (or equivalent) | HS courses in a biology or chemistry laboratory  (but not AP) | Previously used a pipette |
| UD09 (Day 1 only) | 18-25 | Male | HS degree  (or equivalent) | Community college courses  in a biology or chemistry laboratory | Previously used a pipette, familiar with laboratory biosafety guidelines from HS;  Experience following detailed step-by-step instructions (for work or a hobby) |
| UD10 | 18-25 | Female | HS degree  (or equivalent) | Community college courses  in a biology or chemistry laboratory | Previously used a pipette;  Familiar with laboratory biosafety guidelines from HS;  Experience following detailed step-by-step instructions (for work or a hobby) |
| UD11 | 18-25 | Female | HS degree  (or equivalent) | Community college courses  in a biology or chemistry laboratory | Previously used a pipette;  Experience in handling biological or clinical samples in HS;  Experience following detailed step-by-step instructions (for work or a hobby) |
| UD12 | 18-25 | Female | Some college/  no degree | No laboratory experience | Previously used a pipette |
| UD13 | 26-45 | Female | Bachelor's degree | HS courses in a biology or chemistry laboratory  (but not AP) | Experience following detailed step-by-step instructions (for work or a hobby) |
| UD14 (Day 1 only) | 18-25 | Male | Some college/  no degree | HS courses in a biology or chemistry laboratory  (but not AP) | Previously used a pipette;  Experience following detailed step-by-step instructions (for work or a hobby) |
| UD15 | 18-25 | Male | Some college/  no degree | HS courses in a biology or chemistry laboratory  (but not AP) | Experience following detailed step-by-step instructions (for work or a hobby) |
| UD16 | 46-65 | Female | Master's/Doctoral/  Professional degree | AP HS laboratory courses in biology or chemistry;  University (BSc program) courses  in a biology or chemistry laboratory;  Life Sciences (biology, chemistry, physics or similar) major in college | Previously used a pipette;  Experience in handling biological or clinical samples in HS;  Familiar with laboratory biosafety guidelines from HS;  Experience following detailed step-by-step instructions (for work or a hobby);  Familiar with the concept of sample pooling |
| UD17 | 26-45 | Male | HS degree  (or equivalent) | No laboratory experience | None |
| UD18 | 46-65 | Female | Bachelor's degree | HS courses in a biology or chemistry laboratory  (but not AP) | None |
| UD19 | 26-45 | Male | Master's/Doctoral/  Professional degree | No laboratory experience | None |

**Table S6.** **A modified post-study system usefulness questionnaire (PSSUQ) to evaluate the usability of written paper instructions.** The score follows a 7-point Likert scale, including the option of ‘Not Applicable.’ NA, not applicable.

| These questions are about the usability of the written instructions. Please indicate how strongly you agree/disagree with each of the following statements. 1 = strongly agree ... 7 = strongly disagree | | | | | | | | |
| --- | --- | --- | --- | --- | --- | --- | --- | --- |
| 1 | Overall, I am satisfied with how easy it is to perform sample pooling when using the written instructions. | | | | | | | |
| 1 | | 2 | 3 | 4 | 5 | 6 | 7 | NA |
| 2 | It was simple to perform sample pooling when using the written instructions. | | | | | | | |
| 1 | | 2 | 3 | 4 | 5 | 6 | 7 | NA |
| 3 | I could perform high-accuracy sample pooling when using the written instructions. | | | | | | | |
| 1 | | 2 | 3 | 4 | 5 | 6 | 7 | NA |
| 4 | I was able to complete sample pooling quickly, with minimal mistakes, when using the written instructions. | | | | | | | |
| 1 | | 2 | 3 | 4 | 5 | 6 | 7 | NA |
| 5 | I was able to perform sample pooling efficiently (I was productive with minimal wasted effort) when using the written instructions. | | | | | | | |
| 1 | | 2 | 3 | 4 | 5 | 6 | 7 | NA |
| 6 | I felt comfortable performing sample pooling when using the written instructions. | | | | | | | |
| 1 | | 2 | 3 | 4 | 5 | 6 | 7 | NA |
| 7 | It was easy to learn how to achieve high-performance sample pooling when using the written instructions. | | | | | | | |
| 1 | | 2 | 3 | 4 | 5 | 6 | 7 | NA |
| 8 | I believe I could become productive quickly to achieve high-performance sample pooling when using the written instructions. | | | | | | | |
| 1 | | 2 | 3 | 4 | 5 | 6 | 7 | NA |
| 9 | The written instructions gave guidance that clearly told me how to fix problems. | | | | | | | |
| 1 | | 2 | 3 | 4 | 5 | 6 | 7 | NA |
| 10 | Whenever I made a mistake, I could recover easily and quickly by following the written instructions. | | | | | | | |
| 1 | | 2 | 3 | 4 | 5 | 6 | 7 | NA |
| 11 | The information (such as descriptive texts or graphical messages) provided was clear and intuitive. | | | | | | | |
| 1 | | 2 | 3 | 4 | 5 | 6 | 7 | NA |
| 12 | It was easy to find the information I needed from the written instructions. | | | | | | | |
| 1 | | 2 | 3 | 4 | 5 | 6 | 7 | NA |
| 13 | The information (such as descriptive texts or graphical messages) provided in the written instructions was easy to understand. | | | | | | | |
| 1 | | 2 | 3 | 4 | 5 | 6 | 7 | NA |
| 14 | The information (such as descriptive texts or graphical messages) in the written instructions was effective in helping me complete sample pooling. | | | | | | | |
| 1 | | 2 | 3 | 4 | 5 | 6 | 7 | NA |
| 15 | The organization of information in the written instructions was clear. | | | | | | | |
| 1 | | 2 | 3 | 4 | 5 | 6 | 7 | NA |
| 16 | The interface of the written instructions was pleasant. NOTE: The interface of the written instructions includes, but is not limited to: text, language, visual formatting, instruction structure and organization, and user-centric perspectives. | | | | | | | |
| 1 | | 2 | 3 | 4 | 5 | 6 | 7 | NA |
| 17 | I liked reading and following the written instructions to perform sample pooling. | | | | | | | |
| 1 | | 2 | 3 | 4 | 5 | 6 | 7 | NA |
| 18 | The written instructions had all the functions and capabilities I expect it to have. | | | | | | | |
| 1 | | 2 | 3 | 4 | 5 | 6 | 7 | NA |
| 19 | Overall, I am satisfied with the written instructions that guided me in completing high-performance sample pooling. | | | | | | | |
| 1 | | 2 | 3 | 4 | 5 | 6 | 7 | NA |

**Table S7. A modified post-study system usefulness questionnaire (PSSUQ) to evaluate the usability of instructional pooling device.** NA, Not Applicable.

| These questions are about the usability of the written instructions. Please indicate how strongly you agree/disagree with each of the following statements. 1 = strongly agree ... 7 = strongly disagree | | | | | | | | |
| --- | --- | --- | --- | --- | --- | --- | --- | --- |
| 1 | Overall, I am satisfied with how easy it is to perform sample pooling when using the device. | | | | | | | |
| 1 | | 2 | 3 | 4 | 5 | 6 | 7 | NA |
| 2 | It was simple to perform sample pooling when using the device. | | | | | | | |
| 1 | | 2 | 3 | 4 | 5 | 6 | 7 | NA |
| 3 | I could perform high-accuracy sample pooling when using the device. | | | | | | | |
| 1 | | 2 | 3 | 4 | 5 | 6 | 7 | NA |
| 4 | I was able to complete sample pooling quickly, with minimal mistakes, when using the device. | | | | | | | |
| 1 | | 2 | 3 | 4 | 5 | 6 | 7 | NA |
| 5 | I was able to perform sample pooling efficiently (I was productive with minimal wasted effort) when using the device. | | | | | | | |
| 1 | | 2 | 3 | 4 | 5 | 6 | 7 | NA |
| 6 | I felt comfortable performing sample pooling when using the device. | | | | | | | |
| 1 | | 2 | 3 | 4 | 5 | 6 | 7 | NA |
| 7 | It was easy to learn how to achieve high-performance sample pooling when using the device. | | | | | | | |
| 1 | | 2 | 3 | 4 | 5 | 6 | 7 | NA |
| 8 | I believe I could become productive quickly to achieve high-performance sample pooling when using the device. | | | | | | | |
| 1 | | 2 | 3 | 4 | 5 | 6 | 7 | NA |
| 9 | The device gave guidance that clearly told me how to fix problems. | | | | | | | |
| 1 | | 2 | 3 | 4 | 5 | 6 | 7 | NA |
| 10 | Whenever I made a mistake, I could recover easily and quickly by following the device. | | | | | | | |
| 1 | | 2 | 3 | 4 | 5 | 6 | 7 | NA |
| 11 | The information (such as descriptive texts or graphical messages) provided was clear and intuitive. | | | | | | | |
| 1 | | 2 | 3 | 4 | 5 | 6 | 7 | NA |
| 12 | It was easy to find the information I needed from the device. | | | | | | | |
| 1 | | 2 | 3 | 4 | 5 | 6 | 7 | NA |
| 13 | The information (such as descriptive texts or graphical messages) provided by the device was easy to understand. | | | | | | | |
| 1 | | 2 | 3 | 4 | 5 | 6 | 7 | NA |
| 14 | The information (such as descriptive texts or graphical messages) from the device was effective in helping me complete sample pooling. | | | | | | | |
| 1 | | 2 | 3 | 4 | 5 | 6 | 7 | NA |
| 15 | The organization of information on the device was clear. | | | | | | | |
| 1 | | 2 | 3 | 4 | 5 | 6 | 7 | NA |
| 16 | The interface of the device was pleasant. NOTE: The interface of the device includes, but is not limited to: language-agnostic graphics, blinking LEDs, color-coded device architecture, audible beeps, instruction structure and organization, and user-centric perspectives. | | | | | | | |
| 1 | | 2 | 3 | 4 | 5 | 6 | 7 | NA |
| 17 | I liked interacting with the device to perform sample pooling. | | | | | | | |
| 1 | | 2 | 3 | 4 | 5 | 6 | 7 | NA |
| 18 | The device had all the functions and capabilities I expect it to have. | | | | | | | |
| 1 | | 2 | 3 | 4 | 5 | 6 | 7 | NA |
| 19 | Overall, I am satisfied with the device that guides me in completing high-performance sample pooling. | | | | | | | |
| 1 | | 2 | 3 | 4 | 5 | 6 | 7 | NA |

**Table S8. Statistical analysis results for PSSUQ surveys.** One-sided Wilcoxon signed-rank test was used for the comparison of paired PSSUQ scores between paper instructions and instructional device from individual participants. Statistic and *P* values were extracted from the one-sided Wilcoxon signed-rank tests. Matched-pairs rank biserial correlation coefficients were calculated for effect sizes (*r*_C_) using the statistic and ranks from the Wilcoxon tests. Significance levels: **P* < 0.05; ***P* < 0.01; ****P* < 0.001; *****P* < 0.0001. Adj. *P* value, Benjamini-Hochberg adjusted *P* value (false-discovery rate: 5%); SysUse, system usefulness; InfoQual, information quality; InterQual, interface quality; *r*_C_, matched-pairs rank biserial correlation coefficients; CI, confidence interval.

| **Protocol 1 Group** | | | | | | | |
| --- | --- | --- | --- | --- | --- | --- | --- |
| Sample Type | Category | Statistic (*T*) | *P* value | Adj. *P* value | Effect size (*r*_C_) | 95% CI Lower | 95% CI Upper |
| Respiratory  (N=10) | SysUse | 35.5 | 0.207 | 0.207 | 0.291 | −0.455 | 0.818 |
|  | InfoQual | 42.5 | 0.0618 | 0.0928 | 0.545 | −0.0727 | 0.945 |
|  | InterQual | 52.0 | **0.00600 | *0.0180 | 0.891 | 0.618 | 1.000 |
| Stool  (N=9) | SysUse | 26.5 | 0.316 | 0.474 | 0.178 | −0.533 | 0.778 |
|  | InfoQual | 21.0 | 0.571 | 0.571 | −0.0667 | −0.778 | 0.600 |
|  | InterQual | 30.5 | 0.167 | 0.474 | 0.356 | −0.267 | 0.867 |
| **Protocol 2 Group** | | | | | | | |
| Sample Type | Category | Statistic (*T*) | *P* value | Adj. *P* value | Effect size (*r*_C_) | 95% CI Lower | 95% CI Upper |
| Respiratory  (N=12) | SysUse | 72.5 | ******0.00426 | *0.0103 | 0.859 | 0.500 | 1.000 |
|  | InfoQual | 69.5 | ******0.00836 | *0.0103 | 0.782 | 0.321 | 1.000 |
|  | InterQual | 68.0 | *****0.0103 | *0.0103 | 0.756 | 0.295 | 0.974 |
| Stool  (N=17) | SysUse | 140.5 | **0.00122 | **0.00122 | 0.837 | 0.523 | 0.993 |
|  | InfoQual | 141.5 | **0.00104 | **0.00122 | 0.850 | 0.549 | 0.993 |
|  | InterQual | 148.0 | ***0.000873 | **0.00122 | 0.863 | 0.595 | 0.980 |

**Table S9. Statistical analysis results for KTEQ and PTEQ surveys.** One-sided Binomial tests (threshold probability: 75%) and Wilcoxon signed-rank tests were used for statistical analysis. Significance levels: **P* < 0.05; ***P* < 0.01; ****P* < 0.001; *****P* < 0.0001. Adj. *P* value, Benjamini-Hochberg adjusted *P* value (false-discovery rate: 5%).

| **Kirkpatrick Training Evaluation Questionnaire (KTEQ) on Day 1** | | | | | | | |
| --- | --- | --- | --- | --- | --- | --- | --- |
| Protocol | Items | Total responses | Responses to  Likert Score 4 or 5 | Percentage | *P* value | Adj. *P* value | Test |
| 1 | Q1 | 19 | 18 | 94.7% | *0.0310 | *0.0443 | One-sided Binomial tests  (Threshold: 75%) |
|  | Q2 |  | 18 | 94.7% | *0.0310 | *0.0443 |  |
|  | Q3 |  | 19 | 100% | **0.00423 | *0.0106 |  |
|  | Q4 |  | 19 | 100% | **0.00423 | *0.0106 |  |
|  | Q5 |  | 19 | 100% | **0.00423 | *0.0106 |  |
|  | Q6 |  | 19 | 100% | **0.00423 | *0.0106 |  |
|  | Q7 |  | 17 | 89.5% | 0.111 | 0.124 |  |
|  | Q8 |  | 16 | 84.2% | 0.263 | 0.263 |  |
|  | Q9 |  | 17 | 89.5% | 0.111 | 0.124 |  |
|  | Q10 |  | 18 | 94.7% | *0.0310 | *0.0443 |  |
| 2 | Q1 | 29 | 29 | 100% | ***0.000238 | **0.00238 |  |
|  | Q2 |  | 28 | 96.6% | **0.00254 | **0.00508 |  |
|  | Q3 |  | 27 | 93.1% | *0.0133 | *0.0221 |  |
|  | Q4 |  | 28 | 96.6% | **0.00254 | **0.00508 |  |
|  | Q5 |  | 28 | 96.6% | **0.00254 | **0.00508 |  |
|  | Q6 |  | 28 | 96.6% | **0.00254 | **0.00508 |  |
|  | Q7 |  | 24 | 82.8% | 0.232 | 0.232 |  |
|  | Q8 |  | 24 | 82.8% | 0.232 | 0.232 |  |
|  | Q9 |  | 25 | 86.2% | 0.115 | 0.144 |  |
|  | Q10 |  | 25 | 86.2% | 0.115 | 0.144 |  |
| **Post-Training Effectiveness Questionnaire (PTEQ) on Day 2** | | | | | | | |
| Protocol | Items | Total responses | Responses to  ‘Some’ or ‘A lot’ | Percentage | *P* value | Adj. *P* value | Test |
| 1 | Q1 | 19 | 19 | 100% | **0.00423 | **0.00846 | One-sided Binomial tests  (Threshold: 75%) |
| 2 | Q1 | 24 | 23 | 95.8% | **0.00903 | **0.00903 |  |
| Protocol | Sample Type | Reference | Test | Statistic (T) | *P* value | Adj. *P* value | Test |
| 1 | Respiratory | Q2 | Q3 | 0.0 | ***0.000977 | **0.00391 | One-sided  Wilcoxon  signed-rank test |
|  | Stool | Q2 | Q3 | 1.0 | *0.0226 | *0.0324 |  |
| 2 | Respiratory | Q2 | Q3 | 0.0 | *0.0132 | *0.0207 |  |
|  | Stool | Q2 | Q3 | 7.0 | *0.0321 | *0.0269 |  |

**Table S10. Severity level of user handling errors.**

| Severity level | Definition |
| --- | --- |
| Minor | (1) Handling errors may pose minimal risk to biosafety, which is unlikely to cause harm to personnel from biohazardous materials. (2) Handling errors that have a potential impact on the pooling process but are unlikely to lead to significant degradation of pooling performance or process failure. These errors are relatively easy to correct and do not require immediate attention. |
| Severe | (1) Handling errors that pose a significant risk to biosafety. These actions can lead to severe consequences, including serious harm to personnel from clinical samples. Immediate corrective action is required to prevent catastrophic outcomes and ensure the highest level of safety. (2) Handling errors that significantly affect the process and can cause false diagnostic results. These errors can make it difficult to track samples or lead to cross-contamination. Immediate attention and corrective action are required to prevent further complications and ensure the integrity of the pooling process. (3) Handling errors that can cause the complete failure of the pooling process. Failure of pooling occurs if the pooling size is not 5, the generated pool consists of samples from incorrect pooling set(s), the volume of the pool is insufficient for downstream testing, or no new pool is generated. These errors are severe and can lead to catastrophic outcomes if not addressed immediately. Outcomes may include, but are not limited to, unreliable downstream processes and invalid diagnostic results. These errors demand urgent corrective action to prevent incorrect results, safety hazards, or irreparable damage to the process. |

**Table S11. Statistical analysis results for all uncorrected handling errors (∑*err*_all_) across pooling sessions for Protocol 1 group.** Matched-pairs rank biserial correlation coefficients (*r*_C_) were calculated for effect sizes using the statistic and ranks from one-sided Wilcoxon signed rank tests. Two mixed linear model regressions with maximum likelihood (ML) estimation were performed to analyze assistive effects and training effects, separately. The handling error data were transformed using Yeo-Johnson before the analyses. Dummy coding was used for the regressions. Reference categories were Protocol 1 for the ‘Group’ variable and Session #1 for the ‘Session’ variable. Significance levels: **P* < 0.05; ***P* < 0.01; ****P* < 0.001; *****P* < 0.0001. Adj. *P* value, Benjamini-Hochberg adjusted *P* value (false-discovery rate: 5%); SE, standard error; CI, confidence interval.

| **Protocol 1 Group** | | | | | | | |
| --- | --- | --- | --- | --- | --- | --- | --- |
| **Effect sizes (*r*_C_)** | | | | | | | |
| Sample Type | Reference | Test | Statistic (*T*) | Adj. *P* value | Effect size | 95% CI Lower | 95% CI Upper |
| Respiratory  (N=10) | Session #1 (BASE) | Session #2 | 24.0 | 0.677 | −0.127 | −0.945 | 0.564 |
|  |  | Session #3 | 37.5 | 0.213 | 0.364 | −0.309 | 0.945 |
|  |  | Session #4 | 23.0 | 0.677 | −0.164 | −0.800 | 0.582 |
|  |  | Session #5 | 41.0 | 0.192 | 0.491 | −0.200 | 0.891 |
|  |  | Session #6 | 39.5 | 0.192 | 0.436 | −0.236 | 0.909 |
|  |  | Session #7 | 54.5 | *0.0102 | 0.982 | 0.891 | 1.000 |
|  |  | Session #8 | 54.5 | *0.0102 | 0.982 | 0.891 | 1.000 |
| Stool  (N=10) | Session #1 (BASE) | Session #2 | 12.0 | 0.895 | −0.467 | −1.000 | 0.244 |
|  |  | Session #3 | 28.5 | 0.356 | 0.267 | −0.467 | 0.933 |
|  |  | Session #4 | 16.0 | 0.895 | −0.289 | −0.844 | 0.489 |
|  |  | Session #5 | 32.0 | 0.302 | 0.422 | −0.289 | 0.933 |
|  |  | Session #6 | 28.0 | 0.356 | 0.244 | −0.467 | 0.778 |
|  |  | Session #7 | 45.0 | **0.00684 | 1.000 | 1.000 | 1.000 |
|  |  | Session #8 | 45.0 | **0.00684 | 1.000 | 1.000 | 1.000 |
| **Mixed Linear Model Regression Results (Analysis for Assistive Effects)** | | | | | | | |
| Sample Type | Reference | Fixed Effects | Coefficient *β* (SE) | 95% CI Lower | 95% CI Upper | Statistic (z) | *P* value |
| Respiratory | Within Group (Protocol 1) | | | | | | |
|  | - | Intercept | 1.62 (0.169) | 1.29 | 1.95 | 9.62 | ****6.80e-22 |
|  | Session #1 (BASE) | Session #2 | −0.0338 (1.38) | −0.420 | 0.352 | −0.171 | 0.864 |
|  |  | Session #3 | −1.23 (1.38) | −3.93 | 1.46 | −0.896 | 0.370 |
|  |  | Session #6 | −0.212 (1.38) | −0.598 | 0.175 | −1.07 | 0.283 |
|  |  | Session #7 | −10.8 (1.38) | −13.5 | −8.12 | −7.86 | ****3.84e-15 |
|  | Between Groups (Protocol 2 x Session) | | | | | | |
|  | Protocol 1 | Session #1 | 0.0134 (0.190) | −0.359 | 0.386 | 0.705 | 0.944 |
|  |  | Session #2 | −8.36 (1.86) | −12.0 | −4.71 | −4.49 | ****7.24e-06 |
|  |  | Session #3 | −8.85 (1.86) | −12.5 | −5.20 | −4.75 | ****2.05e-06 |
|  |  | Session #6 | −5.63 (1.89) | −9.32 | −1.94 | −2.99 | **0.00282 |
|  |  | Session #7 | 2.67 (1.89) | −1.02 | 6.37 | 1.42 | 0.156 |
|  | Random Effects | | Variance |  |  |  |  |
|  | Between-Participant (Intercept) | | 0.103 |  |  |  |  |
|  | Within-Participant (Residual) | | 0.194 |  |  |  |  |
| Note: N=108 (Protocol 1: 10 participants x 5 sessions; Protocol 2: 11 participants x 5 sessions + 1 participant x 3 sessions). Log-likelihood: −75.1 | | | | | | | |
| Stool | Within Group (Protocol 1) | | | | | | |
|  | - | Intercept | 1.28 (0.142) | 1.01 | 1.56 | 9.04 | ****1.52e-19 |
|  | Session #1 (BASE) | Session #2 | 0.061 (1.17) | −0.363 | 0.389 | 0.363 | 0.717 |
|  |  | Session #3 | −0.257 (1.17) | −2.55 | 2.03 | −0.220 | 0.826 |
|  |  | Session #6 | −0.103 (1.17) | −0.430 | 0.225 | −0.613 | 0.540 |
|  |  | Session #7 | −8.23 (1.17) | −10.5 | −5.94 | −7.04 | ****1.87e-12 |
|  | Between Groups (Protocol 2 x Session) | | | | | | |
|  | Protocol 1 | Session #1 | −0.0729 (0.150) | −0.367 | 0.222 | −0.486 | 0.627 |
|  |  | Session #2 | −6.92 (1.44) | −9.75 | −4.09 | −4.79 | ****1.68e-06 |
|  |  | Session #3 | −6.92 (1.44) | −9.75 | −4.09 | −4.79 | ****1.68e-06 |
|  |  | Session #6 | −5.43 (1.49) | −8.35 | −2.52 | −3.65 | ***0.000261 |
|  |  | Session #7 | 2.07 (1.49) | −0.852 | 4.98 | 1.39 | 0.165 |
|  | Random Effects | | Variance |  |  |  |  |
|  | Between-Participant (Intercept) | | 0.0853 |  |  |  |  |
|  | Within-Participant (Residual) | | 0.126 |  |  |  |  |
| Note: N=122 (Protocol 1: 9 participants x 5 sessions; Protocol 2: 13 participants x 5 sessions + 4 participants x 3 sessions). Log-likelihood: −61.4 | | | | | | | |
| **Mixed Linear Model Regression Results (Analysis for Training Effects)** | | | | | | | |
| Sample Type | Reference | Fixed Effects | Coefficient *β* (SE) | 95% CI Lower | 95% CI Upper | Statistic (z) | *P* value |
| Respiratory | Within Group (Protocol 1) | | | | | | |
|  | - | Intercept | 2.41 (0.337) | 1.75 | 3.07 | 7.15 | ****8.57e-13 |
|  | Session #1 (BASE) | Session #4 | 0.00832 (1.42) | −2.77 | 2.78 | 0.00587 | 0.995 |
|  |  | Session #8 | −8.53 (1.42) | −11.3 | −5.75 | −6.02 | ****1.73e-09 |
|  | Between Groups (Protocol 2 x Session) | | | | | | |
|  | Protocol 1 | Session #1 | 0.123 (0.390) | −0.640 | 0.887 | 0.317 | 0.752 |
|  |  | Session #4 | −0.790 (1.92) | −4.55 | 2.97 | −0.0412 | 0.680 |
|  |  | Session #8 | 7.11 (1.92) | 3.31 | 10.9 | 3.67 | ***0.000246 |
|  | Random Effects | | Variance |  |  |  |  |
|  | Between-Participant (Intercept) | | 0.351 |  |  |  |  |
|  | Within-Participant (Residual) | | 0.814 |  |  |  |  |
| Note: N=65 (Protocol 1: 10 participants x 3 sessions; Protocol 2: 11 participants x 3 sessions + 1 participant x 2 sessions). Log-likelihood: −92.7 | | | | | | | |
| Stool | Within Group (Protocol 1) | | | | | | |
|  | - | Intercept | 2.17 (0.266) | 1.65 | 2.69 | 8.16 | ****3.34e-16 |
|  | Session #1 (BASE) | Session #4 | −0.09 (1.11) | −2.26 | 2.08 | −0.081 | 0.935 |
|  |  | Session #8 | −6.44 (1.11) | −8.61 | −4.27 | −5.81 | ****6.31e-09 |
|  | Between Groups (Protocol 2 x Session) | | | | | | |
|  | Protocol 1 | Session #1 | −0.336 (0.286) | −0.896 | 0.223 | −1.18 | 0.239 |
|  |  | Session #4 | 0.954 (1.37) | −1.73 | 3.64 | 0.696 | 0.487 |
|  |  | Session #8 | 5.34 (1.41) | 2.57 | 8.11 | 3.78 | ***0.000159 |
|  | Random Effects | | Variance |  |  |  |  |
|  | Between-Participant (Intercept) | | 0.261 |  |  |  |  |
|  | Within-Participant (Residual) | | 0.449 |  |  |  |  |
| Note: N=74 (Protocol 1: 9 participants x 3 sessions; Protocol 2: 13 participants x 3 sessions + 4 participants x 2 sessions). Log-likelihood: −85.8 | | | | | | | |

**Table S12. Statistical analysis results for average volume-transfer accuracy (Avg. Acc_pool_) across pooling sessions for Protocol 1 group.** Hedges’ g values were calculated for effect sizes. As a reference, statistical results from one-sided paired Student *t* tests are reported. Two mixed linear model regressions with maximum likelihood (ML) estimation were performed to analyze assistive effects and training effects, separately. The accuracy data were logit-transformed before the analyses. Dummy coding was used for the regressions. Reference categories were Protocol 1 for the ‘Group’ variable and Session #1 for the ‘Session’ variable. Significance levels: **P* < 0.05; ***P* < 0.01; ****P* < 0.001; *****P* < 0.0001. Adj. *P* value, Benjamini-Hochberg adjusted *P* value (false-discovery rate: 5%); SE, standard error; CI, confidence interval.

| **Protocol 1 Group** | | | | | | | |
| --- | --- | --- | --- | --- | --- | --- | --- |
| **Effect sizes (Hedges’ g)** | | | | | | | |
| Sample Type | Reference | Test | Statistic (*t*) | Adj. *P* value | Effect size | 95% CI Lower | 95% CI Upper |
| Respiratory  (N=10) | Session #1 (BASE) | Session #2 | 2.52 | 0.984 | −0.344 | −0.622 | −0.0537 |
|  |  | Session #3 | −0.0315 | 0.811 | 0.00460 | −0.287 | 0.319 |
|  |  | Session #4 | 0.354 | 0.811 | −0.0512 | −0.307 | 0.323 |
|  |  | Session #5 | 0.528 | 0.811 | −0.0938 | −0.654 | 0.218 |
|  |  | Session #6 | 0.479 | 0.811 | −0.0774 | −0.555 | 0.222 |
|  |  | Session #7 | −4.90 | **0.00282 | 1.51 | 0.943 | 2.55 |
|  |  | Session #8 | −4.45 | **0.00282 | 1.78 | 1.09 | 2.69 |
| Stool  (N=9) | Session #1 (BASE) | Session #2 | 0.283 | 0.709 | −0.0447 | −0.391 | 0.246 |
|  |  | Session #3 | −0.00639 | 0.697 | 0.00161 | −0.497 | 0.548 |
|  |  | Session #4 | 1.20 | 0.868 | −0.187 | −0.490 | 0.120 |
|  |  | Session #5 | −1.08 | 0.364 | 0.225 | −0.188 | 0.826 |
|  |  | Session #6 | −0.202 | 0.697 | 0.0398 | −0.406 | 0.482 |
|  |  | Session #7 | −3.68 | *0.0109 | 1.02 | 0.469 | 1.83 |
|  |  | Session #8 | −4.93 | **0.00405 | 1.70 | 0.973 | 2.86 |
| **Mixed Linear Model Regression Results (Analysis for Assistive Effects)** | | | | | | | |
| Sample Type | Reference | Fixed Effects | Coefficient *β* (SE) | 95% CI Lower | 95% CI Upper | Statistic (z) | *P* value |
| Respiratory | Within Group (Protocol 1) | | | | | | |
|  | - | Intercept | 0.441 (0.160) | 0.127 | 0.754 | 2.75 | **0.00593 |
|  | Session #1 (BASE) | Session #2 | −0.143 (0.207) | −0.550 | 0.263 | −0.690 | 0.490 |
|  |  | Session #3 | −0.00305 (0.207) | −0.409 | 0.403 | −0.0147 | 0.988 |
|  |  | Session #6 | −0.0147 (0.207) | −0.421 | 0.392 | −0.0707 | 0.944 |
|  |  | Session #7 | 0.593 (0.207) | 0.187 | 1.000 | 2.86 | **0.00423 |
|  | Between Groups (Protocol 2 x Session) | | | | | | |
|  | Protocol 1 | Session #1 | −0.300 (0.200) | −0.691 | 0.0914 | −1.50 | 0.133 |
|  |  | Session #2 | 0.922 (0.281) | 0.372 | 1.47 | 3.29 | **0.00102 |
|  |  | Session #3 | 1.23 (0.281) | 0.675 | 1.78 | 4.36 | ****0.0000128 |
|  |  | Session #6 | 1.30 (0.284) | 0.747 | 1.86 | 4.59 | ****4.42e-06 |
|  |  | Session #7 | 0.949 (0.284) | 0.393 | 1.51 | 3.34 | ***0.000827 |
|  | Random Effects | | Variance |  |  |  |  |
|  | Between-Participant (Intercept) | | 0.0442 |  |  |  |  |
|  | Within-Participant (Residual) | | 0.215 |  |  |  |  |
| Note: N=108 (Protocol 1: 10 participants x 5 sessions; Protocol 2: 11 participants x 5 sessions + 1 participant x 3 sessions). Log-likelihood: −76.5 | | | | | | | |
| Stool | Within Group (Protocol 1) | | | | | | |
|  | - | Intercept | 1.29 (0.128) | 1.04 | 1.55 | 10.1 | ****6.07e-24 |
|  | Session #1 (BASE) | Session #2 | −0.0345 (0.162) | −0.352 | 0.283 | −0.213 | 0.831 |
|  |  | Session #3 | 0.0205 (0.162) | −0.297 | 0.338 | 0.127 | 0.899 |
|  |  | Session #6 | −0.0178 (0.162) | −0.335 | 0.299 | −0.110 | 0.912 |
|  |  | Session #7 | 0.498 (0.162) | 0.181 | 0.815 | 3.08 | **0.00209 |
|  | Between Groups (Protocol 2 x Session) | | | | | | |
|  | Protocol 1 | Session #1 | −0.040 (0.145) | −0.323 | 0.243 | −0.277 | 0.782 |
|  |  | Session #2 | 0.415 (0.200) | 0.0232 | 0.808 | 2.08 | *0.0379 |
|  |  | Session #3 | 0.443 (0.200) | 0.0506 | 0.835 | 2.21 | *0.0269 |
|  |  | Session #6 | 0.696 (0.206) | 0.293 | 1.100 | 3.38 | ***0.000725 |
|  |  | Session #7 | 0.323 (0.206) | −0.0811 | 0.727 | 1.57 | 0.117 |
|  | Random Effects | | Variance |  |  |  |  |
|  | Between-Participant (Intercept) | | 0.0427 |  |  |  |  |
|  | Within-Participant (Residual) | | 0.118 |  |  |  |  |
| Note: N=122 (Protocol 1: 9 participants x 5 sessions; Protocol 2: 13 participants x 5 sessions + 4 participants x 3 sessions). Log-likelihood: −53.3 | | | | | | | |
| **Mixed Linear Model Regression Results (Analysis for Training Effects)** | | | | | | | |
| Sample Type | Reference | Fixed Effects | Coefficient *β* (SE) | 95% CI Lower | 95% CI Upper | Statistic (z) | *P* value |
| Respiratory | Within Group (Protocol 1) | | | | | | |
|  | - | Intercept | 0.427 (0.187) | 0.0600 | 0.793 | 2.28 | *0.0226 |
|  | Session #1 (BASE) | Session #4 | −0.0239 (0.254) | −0.521 | 0.473 | −0.0946 | 0.925 |
|  |  | Session #8 | 1.10 (0.254) | 0.605 | 1.60 | 4.35 | ****0.000014 |
|  | Between Groups (Protocol 2 x Session) | | | | | | |
|  | Protocol 1 | Session #1 | −0.286 (0.244) | −0.764 | 0.192 | −1.17 | 0.241 |
|  |  | Session #4 | 0.971 (0.343) | 0.298 | 1.64 | 2.83 | **0.00469 |
|  |  | Session #8 | 0.112 (0.347) | −0.568 | 0.792 | 0.324 | 0.746 |
|  | Random Effects | | Variance |  |  |  |  |
|  | Between-Participant (Intercept) | | 0.0283 |  |  |  |  |
|  | Within-Participant (Residual) | | 0.321 |  |  |  |  |
| Note: N=65 (Protocol 1: 10 participants x 3 sessions; Protocol 2: 11 participants x 3 sessions + 1 participant x 2 sessions). Log-likelihood: −57.7 | | | | | | | |
| Stool | Within Group (Protocol 1) | | | | | | |
|  | - | Intercept | 1.30 (0.141) | 1.02 | 1.58 | 9.21 | ****3.21e-20 |
|  | Session #1 (BASE) | Session #4 | −0.103 (0.190) | −0.476 | 0.270 | −0.542 | 0.588 |
|  |  | Session #8 | 0.833 (0.190) | 0.460 | 1.21 | 4.38 | ****0.0000120 |
|  | Between Groups (Protocol 2 x Session) | | | | | | |
|  | Protocol 1 | Session #1 | −0.047 (0.169) | −0.378 | 0.284 | −0.277 | 0.782 |
|  |  | Session #4 | 0.368 (0.235) | −0.093 | 0.830 | 1.57 | 0.117 |
|  |  | Session #8 | −0.247 (0.242) | −0.722 | 0.227 | −1.02 | 0.308 |
|  | Random Effects | | Variance |  |  |  |  |
|  | Between-Participant (Intercept) | | 0.0180 |  |  |  |  |
|  | Within-Participant (Residual) | | 0.163 |  |  |  |  |
| Note: N=74 (Protocol 1: 9 participants x 3 sessions; Protocol 2: 13 participants x 3 sessions + 4 participants x 2 sessions). Log-likelihood: −41.2 | | | | | | | |

**Table S13. Linear mixed model analyses (with continuous pooling round variable) for completion time in the Protocol 1 group.** Completion time data were standardized. Significance levels: **P* < 0.05; ***P* < 0.01; ****P* < 0.001; *****P* < 0.0001. SE, standard error; CI, confidence interval.

| **Completion Time ~ Device + Pooling Round + Device × Pooling Round + (1\|Participant)** | | | | | | |
| --- | --- | --- | --- | --- | --- | --- |
| Sample Type | Fixed Effects | Coefficient *β* (SE) | 95% CI Lower | 95% CI Upper | Statistic (z) | *P* value |
| Respiratory | Intercept | 0.302 (0.186) | −0.0621 | 0.666 | 1.63 | 0.104 |
|  | Device | 3.47 (0.688) | 2.12 | 4.82 | 5.04 | ****4.54e-07 |
|  | Pooling Round | −0.0525 (0.00458) | −0.0615 | −0.0435 | −11.5 | ****1.80e-30 |
|  | Device x Pooling Round | −0.0425 (0.0244) | −0.0903 | 0.00523 | −1.75 | 0.0809 |
|  | Combined *β* |  |  |  |  |  |
|  | Pooling Round x (1 + Device) | −0.0950 (0.0239) | −0.142 | −0.0481 | −3.97 | ****7.21e-05 |
|  | Random Effects | Variance |  |  |  |  |
|  | Between-Participant (Intercept) | 0.302 |  |  |  |  |
|  | Within-Participant (Residual) | 0.241 |  |  |  |  |
| Note: N=320 (10 participants x 32 rounds). Log-likelihood: −253 | | | | | | |
| Stool | Intercept | 0.254 (0.202) | −0.141 | 0.650 | 1.26 | 0.208 |
|  | Device | 5.83 (0.787) | 4.28 | 7.37 | 7.40 | ****1.33e-13 |
|  | Pooling Round | −0.0463 (0.00524) | −0.0566 | −0.0360 | −8.84 | ****9.42e-19 |
|  | Device x Pooling Round | −0.133 (0.0279) | −0.188 | −0.0783 | −4.76 | ****1.89e-06 |
|  | Combined *β* |  |  |  |  |  |
|  | Pooling Round x (1 + Device) | −0.179 (0.0274) | −0.233 | −0.126 | −6.54 | ****6.14e-11 |
|  | Random Effects | Variance |  |  |  |  |
|  | Between-Participant (Intercept) | 0.317 |  |  |  |  |
|  | Within-Participant (Residual) | 0.284 |  |  |  |  |
| Note: N=288 (9 participants x 32 rounds). Log-likelihood: −251 | | | | | | |

**Table S14. Linear mixed model analyses (with continuous pooling round variable) for volume-transfer accuracy (Acc_pool_), controlling for completion time in the Protocol 1 group.** Accuracy data were logit-transformed, and completion time data were standardized. Significance levels: **P* < 0.05; ***P* < 0.01; ****P* < 0.001; *****P* < 0.0001. SE, standard error; CI, confidence interval.

| **Accuracy ~ Device + Pooling Round + Completion Time + Device × Pooling Round + Device × Completion Time + (1\|Participant)** | | | | | | |
| --- | --- | --- | --- | --- | --- | --- |
| Sample Type | Fixed Effects | Coefficient *β* (SE) | 95% CI Lower | 95% CI Upper | Statistic (z) | *P* value |
| Respiratory | Intercept | 0.403 (0.160) | 0.0901 | 0.717 | 2.52 | *0.0116 |
|  | Device | −1.87 (0.693) | −3.23 | −0.514 | −2.70 | **0.00691 |
|  | Pooling Round | −0.00365 (0.00517) | −0.0138 | 0.00648 | −0.706 | 0.480 |
|  | Completion Time | −0.0875 (0.0580) | −0.201 | 0.0262 | −1.51 | 0.131 |
|  | Device x Pooling Round | 0.111 (0.0234) | 0.0648 | 0.157 | 4.73 | ****2.28e-06 |
|  | Device x Completion Time | −0.162 (0.0854) | −0.330 | 0.00494 | −1.90 | 0.0572 |
|  | Combined *β* |  |  |  |  |  |
|  | Pooling Round x (1 + Device) | 0.107 (0.0231) | 0.0618 | 0.152 | 4.63 | ****3.58e-06 |
|  | Completion Time x (1 + Device) | −0.250 (0.0794) | −0.406 | −0.0944 | −3.15 | **0.00164 |
|  | Random Effects | Variance |  |  |  |  |
|  | Between-Participant (Intercept) | 0.217 |  |  |  |  |
|  | Within-Participant (Residual) | 0.200 |  |  |  |  |
| Note: N=320 (10 participants x 32 rounds). Log-likelihood: −226 | | | | | | |
| Stool | Intercept | 1.27 (0.153) | 0.970 | 1.57 | 8.31 | ****9.80e-17 |
|  | Device | −1.29 (0.634) | −2.53 | −0.0474 | −2.03 | *0.0419 |
|  | Pooling Round | 0.00481 (0.00404) | −0.00310 | 0.0127 | 1.19 | 0.234 |
|  | Completion Time | 0.0924 (0.0448) | 0.00458 | 0.180 | 2.06 | *0.0392 |
|  | Device x Pooling Round | 0.0670 (0.0212) | 0.0254 | 0.109 | 3.16 | **0.00160 |
|  | Device x Completion Time | −0.124 (0.0666) | −0.254 | 0.00695 | −1.86 | 0.0635 |
|  | Combined *β* |  |  |  |  |  |
|  | Pooling Round x (1 + Device) | 0.0718 (0.0211) | 0.0305 | 0.113 | 3.41 | ***0.000655 |
|  | Completion Time x (1 + Device) | −0.0311 (0.0599) | −0.149 | 0.0864 | −0.519 | 0.603 |
|  | Random Effects | Variance |  |  |  |  |
|  | Between-Participant (Intercept) | 0.217 |  |  |  |  |
|  | Within-Participant (Residual) | 0.200 |  |  |  |  |
| Note: N=288 (9 participants x 32 rounds). Log-likelihood: −251 | | | | | | |

**Table S15. Linear mixed model analyses (with continuous pooling round variable) for all handling (*err*_all_), minor (*err*_minor_), and severe errors (*err*_severe_), controlling for completion time in the Protocol 1 group.** Handling error data were Yeo-Johnson transformed, and completion time data were standardized. Significance levels: **P* < 0.05; ***P* < 0.01; ****P* < 0.001; *****P* < 0.0001. SE, standard error; CI, confidence interval.

| **AllErrors ~ Device + Pooling Round + Completion Time + Device × Pooling Round + Device × Completion Time + (1\|Participant)** | | | | | | |
| --- | --- | --- | --- | --- | --- | --- |
| Sample Type | Fixed Effects | Coefficient *β* (SE) | 95% CI Lower | 95% CI Upper | Statistic (z) | *P* value |
| Respiratory  Note: N=320  (10 participants x 32 rounds).  Log-likelihood: −25.4 | Intercept | 0.685 (0.0808) | 0.527 | 0.843 | 8.48 | ****2.26e-17 |
|  | Device | −0.776 (0.367) | −1.49 | −0.0574 | −2.12 | *0.0343 |
|  | Pooling Round | −0.00317 (0.00272) | −0.00850 | 0.00215 | −1.17 | 0.243 |
|  | Completion Time | 0.0414 (0.0302) | −0.0177 | 0.100 | 1.37 | 0.170 |
|  | Device x Pooling Round | 0.00436 (0.0124) | −0.0199 | 0.0287 | 0.352 | 0.725 |
|  | Device x Completion Time | 0.0193 (0.0452) | −0.0693 | 0.108 | 0.427 | 0.670 |
|  | Combined *β* |  |  |  |  |  |
|  | Pooling Round x (1 + Device) | 0.00119 (0.0122) | −0.0228 | 0.0251 | 0.0969 | 0.923 |
|  | Completion Time x (1 + Device) | 0.0607 (0.0418) | −0.0213 | 0.143 | 1.45 | 0.147 |
|  | Random Effects | Variance |  |  |  |  |
|  | Between-Participant (Intercept) | 0.0544 |  |  |  |  |
|  | Within-Participant (Residual) | 0.0561 |  |  |  |  |
| Stool | Intercept | 0.657 (0.0958) | 0.469 | 0.845 | 6.86 | ****7.03e-12 |
|  | Device | −1.23 (0.375) | −1.97 | −0.496 | −3.28 | **0.00103 |
| Note: N=288  (9 participants x 32 rounds).  Log-likelihood: 10.1 | Pooling Round | −0.00342 (0.00239) | −0.00812 | 0.00127 | −1.43 | 0.153 |
|  | Completion Time | −0.0144 (0.0268) | −0.0670 | 0.0382 | −0.536 | 0.592 |
|  | Device x Pooling Round | 0.0232 (0.0125) | −0.00138 | 0.0478 | 1.85 | 0.0643 |
|  | Device x Completion Time | 0.118 (0.0394) | 0.0413 | 0.196 | 3.01 | **0.00262 |
|  | Combined *β* |  |  |  |  |  |
|  | Pooling Round x (1 + Device) | 0.0198 (0.0125) | −0.00464 | 0.0442 | 1.59 | 0.112 |
|  | Completion Time x (1 + Device) | 0.104 (0.0355) | 0.0344 | 0.174 | 2.93 | **0.00340 |
|  | Random Effects | Variance |  |  |  |  |
|  | Between-Participant (Intercept) | 0.0746 |  |  |  |  |
|  | Within-Participant (Residual) | 0.0433 |  |  |  |  |
| **MinorErrors ~ Device + Pooling Round + Completion Time + Device × Pooling Round + Device × Completion Time + (1\|Participant)** | | | | | | |
| Sample Type | Fixed Effects | Coefficient *β* (SE) | 95% CI Lower | 95% CI Upper | Statistic (z) | *P* value |
| Respiratory | Intercept | 0.364 (0.0544) | 0.258 | 0.471 | 6.70 | ****2.05e-11 |
|  | Device | −0.392 (0.254) | −0.890 | 0.106 | −1.54 | 0.123 |
| Note: N=320  (10 participants x 32 rounds).  Log-likelihood: 90.3 | Pooling Round | −0.00353 (0.00188) | −0.00721 | 0.000161 | −1.87 | 0.0609 |
|  | Completion Time | −0.0245 (0.0209) | −0.0654 | 0.0163 | −1.18 | 0.239 |
|  | Device x Pooling Round | 0.00334 (0.00859) | −0.0135 | 0.0202 | 0.389 | 0.697 |
|  | Device x Completion Time | 0.0605 (0.0313) | −0.000781 | 0.122 | 1.93 | 0.0530 |
|  | Combined *β* |  |  |  |  |  |
|  | Pooling Round x (1 + Device) | −0.000185 (0.00846) | −0.0168 | 0.0164 | −0.0219 | 0.983 |
|  | Completion Time x (1 + Device) | 0.0360 (0.0289) | −0.0207 | 0.0927 | 1.24 | 0.214 |
|  | Random Effects | Variance |  |  |  |  |
|  | Between-Participant (Intercept) | 0.0244 |  |  |  |  |
|  | Within-Participant (Residual) | 0.0269 |  |  |  |  |
| Stool | Intercept | 0.416 (0.0667) | 0.285 | 0.546 | 6.24 | ****4.49e-10 |
|  | Device | −0.881 (0.258) | −1.39 | −0.375 | −3.41 | ***0.000643 |
| Note: N=288  (9 participants x 32 rounds).  Log-likelihood: 115 | Pooling Round | −0.00417 (0.00165) | −0.00739 | −0.000937 | −2.53 | *0.0114 |
|  | Completion Time | −0.0492 (0.0185) | −0.0854 | −0.0131 | −2.67 | **0.00765 |
|  | Device x Pooling Round | 0.0198 (0.00863) | 0.00285 | 0.0367 | 2.29 | *0.0220 |
|  | Device x Completion Time | 0.136 (0.0271) | 0.0828 | 0.189 | 5.02 | ****5.22e-07 |
|  | Combined *β* |  |  |  |  |  |
|  | Pooling Round x (1 + Device) | 0.0156 (0.00857) | −0.00120 | 0.0324 | 1.82 | 0.0687 |
|  | Completion Time x (1 + Device) | 0.0867 (0.0245) | 0.0388 | 0.135 | 3.54 | ***0.000394 |
|  | Random Effects | Variance |  |  |  |  |
|  | Between-Participant (Intercept) | 0.0361 |  |  |  |  |
|  | Within-Participant (Residual) | 0.0205 |  |  |  |  |
| **SevereErrors ~ Device + Pooling Round + Completion Time + Device × Pooling Round + Device × Completion Time + (1\|Participant)** | | | | | | |
| Sample Type | Fixed Effects | Coefficient *β* (SE) | 95% CI Lower | 95% CI Upper | Statistic (z) | *P* value |
| Respiratory | Intercept | 0.306 (0.0432) | 0.221 | 0.391 | 7.08 | ****1.40e-12 |
|  | Device | −0.467 (0.243) | −0.943 | 0.00946 | −1.92 | 0.0547 |
| Note: N=320  (10 participants x 32 rounds).  Log-likelihood: 106 | Pooling Round | −0.00353 (0.00179) | −0.00704 | −0.000010 | −1.97 | *0.0494 |
|  | Completion Time | 0.0207 (0.0198) | −0.0181 | 0.0594 | 1.05 | 0.296 |
|  | Device x Pooling Round | 0.00757 (0.00822) | −0.00853 | 0.0237 | 0.921 | 0.357 |
|  | Device x Completion Time | 0.0219 (0.0299) | −0.0367 | 0.0806 | 0.733 | 0.464 |
|  | Combined *β* |  |  |  |  |  |
|  | Pooling Round x (1 + Device) | 0.00405 (0.00810) | −0.0118 | 0.0199 | 0.500 | 0.617 |
|  | Completion Time x (1 + Device) | 0.0426 (0.0276) | −0.0116 | 0.0967 | 1.54 | 0.123 |
|  | Random Effects | Variance |  |  |  |  |
|  | Between-Participant (Intercept) | 0.0139 |  |  |  |  |
|  | Within-Participant (Residual) | 0.0247 |  |  |  |  |
| Stool | Intercept | 0.216 (0.0503) | 0.163 | 0.360 | 5.19 | ****2.07e-07 |
|  | Device | −0.631 (0.237) | −1.10 | −0.167 | −2.66 | **0.00774 |
| Note: N=288  (9 participants x 32 rounds).  Log-likelihood: 141 | Pooling Round | −0.00133 (0.00151) | −0.00430 | 0.00164 | −0.879 | 0.379 |
|  | Completion Time | −0.00866 (0.0170) | −0.0420 | 0.0247 | −0.509 | 0.611 |
|  | Device x Pooling Round | 0.0138 (0.00793) | −0.00175 | 0.0293 | 1.74 | 0.0820 |
|  | Device x Completion Time | 0.0561 (0.0249) | 0.00733 | 0.105 | 2.25 | *0.0242 |
|  | Combined *β* |  |  |  |  |  |
|  | Pooling Round x (1 + Device) | 0.0125 (0.00788) | −0.00298 | 0.0279 | 1.58 | 0.114 |
|  | Completion Time x (1 + Device) | 0.0475 (0.0225) | 0.00346 | 0.0915 | 2.11 | *0.0345 |
|  | Random Effects | Variance |  |  |  |  |
|  | Between-Participant (Intercept) | 0.0195 |  |  |  |  |
|  | Within-Participant (Residual) | 0.0173 |  |  |  |  |

**Table S16. Linear mixed model analyses (with dummy-coded pooling sessions and group effects) for all uncorrected handling errors (∑*err*_all_), controlling for completion time in the Protocol 1 group.** Two mixed linear model regressions with maximum likelihood (ML) estimation were performed to analyze assistive effects and training effects, separately, with completion time as a covariate. Before the analyses, the handling error data were Yeo-Johnson transformed, and completion time data were standardized. Dummy coding was used for the regressions. Reference categories were Protocol 1 for the ‘Group’ variable and Session #1 for the ‘Session’ variable. Significance levels: **P* < 0.05; ***P* < 0.01; ****P* < 0.001; *****P* < 0.0001. SE, standard error; CI, confidence interval.

| **Protocol 1 Group** | | | | | | | |
| --- | --- | --- | --- | --- | --- | --- | --- |
| **Mixed Linear Model Regression Results (Analysis for Assistive Effects)** | | | | | | | |
| Sample Type | Reference | Fixed Effects | Coefficient *β* (SE) | 95% CI Lower | 95% CI Upper | Statistic (z) | *P* value |
| Respiratory | Within Group (Protocol 1) | | | | | | |
|  | - | Intercept | 1.62 (0.168) | 1.29 | 1.95 | 9.65 | ****4.75e-22 |
|  | Session #1 (BASE) | Time | 0.0206 (0.0832) | −0.143 | 0.184 | 0.247 | 0.805 |
|  |  | Session #2 | −0.134 (1.44) | −2.95 | −2.68 | −0.934 | 0.925 |
|  |  | Session #3 | −1.11 (1.46) | −3.80 | −1.75 | −0.760 | 0.447 |
|  |  | Session #6 | −1.29 (1.57) | −4.37 | −1.79 | −0.821 | 0.411 |
|  |  | Session #7 | −10.9 (1.47) | −13.8 | −8.06 | −7.45 | ****9.14e-14 |
|  | Between Groups (Protocol 2 x Session) | | | | | | |
|  | Protocol 1 | Session #1 | 0.0202 (0.192) | −0.356 | 0.397 | 0.105 | 0.916 |
|  |  | Session #2 | −8.66 (2.22) | −13.0 | −4.31 | −3.90 | ****9.55e-05 |
|  |  | Session #3 | −9.11 (2.15) | −13.3 | −4.90 | −4.24 | ****2.20e-05 |
|  |  | Session #6 | −5.90 (2.19) | −10.2 | −1.61 | −2.70 | **0.00698 |
|  |  | Session #7 | 2.70 (1.89) | −1.01 | 6.40 | 1.43 | 0.154 |
|  | Random Effects | | Variance |  |  |  |  |
|  | Between-Participant (Intercept) | | 0.0998 |  |  |  |  |
|  | Within-Participant (Residual) | | 0.195 |  |  |  |  |
| Note: N=108 (Protocol 1: 10 participants x 5 sessions; Protocol 2: 11 participants x 5 sessions + 1 participant x 3 sessions). Log-likelihood: −75.1 | | | | | | | |
| Stool | Within Group (Protocol 1) | | | | | | |
|  | - | Intercept | 1.28 (0.144) | 0.997 | 1.56 | 8.90 | ****5.61e-19 |
|  | Session #1 (BASE) | Time | −0.0201 (0.0608) | −0.139 | 0.0990 | −0.331 | 0.741 |
|  |  | Session #2 | 0.353 (1.19) | −1.97 | −2.68 | 0.297 | 0.766 |
|  |  | Session #3 | −0.333 (1.19) | −2.66 | −2.00 | −0.281 | 0.779 |
|  |  | Session #6 | −0.836 (1.22) | −3.23 | −1.56 | −0.685 | 0.494 |
|  |  | Session #7 | −8.13 (1.20) | −10.5 | −5.78 | −6.76 | ****1.34e-11 |
|  | Between Groups (Protocol 2 x Session) | | | | | | |
|  | Protocol 1 | Session #1 | −0.0618 (0.154) | −0.363 | 0.239 | −0.402 | 0.687 |
|  |  | Session #2 | −6.77 (1.51) | −9.73 | −3.80 | −4.48 | ****7.61e-06 |
|  |  | Session #3 | −6.80 (1.48) | −9.71 | −3.90 | −4.59 | ****4.42e-06 |
|  |  | Session #6 | −5.33 (1.51) | −8.30 | −2.37 | −3.53 | ***0.000420 |
|  |  | Session #7 | 1.94 (1.53) | −1.07 | 4.94 | 1.26 | 0.206 |
|  | Random Effects | | Variance |  |  |  |  |
|  | Between-Participant (Intercept) | | 0.0872 |  |  |  |  |
|  | Within-Participant (Residual) | | 0.125 |  |  |  |  |
| Note: N=122 (Protocol 1: 9 participants x 5 sessions; Protocol 2: 13 participants x 5 sessions + 4 participants x 3 sessions). Log-likelihood: −61.3 | | | | | | | |
| **Mixed Linear Model Regression Results (Analysis for Training Effects)** | | | | | | | |
| Sample Type | Reference | Fixed Effects | Coefficient *β* (SE) | 95% CI Lower | 95% CI Upper | Statistic (z) | *P* value |
| Respiratory | Within Group (Protocol 1) | | | | | | |
|  | - | Intercept | 2.49 (0.347) | 1.81 | 3.17 | 7.17 | ****7.29e-13 |
|  | Session #1 (BASE) | Time | −0.180 (0.171) | −0.515 | 0.155 | −1.05 | 0.293 |
|  |  | Session #4 | −0.797 (1.59) | −3.91 | 2.31 | −0.502 | 0.615 |
|  |  | Session #8 | −8.11 (1.45) | −10.9 | −5.28 | −5.62 | ****1.96e-08 |
|  | Between Groups (Protocol 2 x Session) | | | | | | |
|  | Protocol 1 | Session #1 | 0.0578 (0.388) | −0.702 | 0.818 | 0.149 | 0.881 |
|  |  | Session #4 | −0.270 (1.95) | −4.08 | 3.54 | −0.139 | 0.889 |
|  |  | Session #8 | 6.21 (2.09) | 2.12 | 10.3 | 2.98 | **0.00291 |
|  | Random Effects | | Variance |  |  |  |  |
|  | Between-Participant (Intercept) | | 0.396 |  |  |  |  |
|  | Within-Participant (Residual) | | 0.784 |  |  |  |  |
| Note: N=65 (Protocol 1: 10 participants x 3 sessions; Protocol 2: 11 participants x 3 sessions + 1 participant x 2 sessions). Log-likelihood: −92.2 | | | | | | | |
| Stool | Within Group (Protocol 1) | | | | | | |
|  | - | Intercept | 2.18 (0.261) | 1.67 | 2.69 | 8.36 | ****6.18e-17 |
|  | Session #1 (BASE) | Time | −0.280 (0.125) | −0.525 | −0.0346 | −2.24 | *0.0253 |
|  |  | Session #4 | −0.810 (1.09) | −2.95 | 1.33 | −0.742 | 0.458 |
|  |  | Session #8 | −6.16 (1.05) | −8.22 | −4.10 | −5.85 | ****4.77e-09 |
|  | Between Groups (Protocol 2 x Session) | | | | | | |
|  | Protocol 1 | Session #1 | −0.157 (0.281) | −0.709 | 0.395 | −0.558 | 0.577 |
|  |  | Session #4 | 0.815 (1.29) | −1.72 | 3.35 | 0.630 | 0.528 |
|  |  | Session #8 | 4.06 (1.45) | 1.21 | 6.90 | 2.79 | **0.00521 |
|  | Random Effects | | Variance |  |  |  |  |
|  | Between-Participant (Intercept) | | 0.314 |  |  |  |  |
|  | Within-Participant (Residual) | | 0.398 |  |  |  |  |
| Note: N=74 (Protocol 1: 9 participants x 3 sessions; Protocol 2: 13 participants x 3 sessions + 4 participants x 2 sessions). Log-likelihood: −83.3 | | | | | | | |

**Table S17. Linear mixed model analyses (with dummy-coded pooling sessions and group effects) for average volume-transfer accuracy (Avg. Acc_pool_), controlling for completion time in the Protocol 1 group.** Two mixed linear model regressions with maximum likelihood (ML) estimation were performed to analyze assistive effects and training effects, separately, with completion time as a covariate. Before the analyses, the accuracy data were logit-transformed, and completion time data were standardized. Dummy coding was used for the regressions. Reference categories were Protocol 1 for the ‘Group’ variable and Session #1 for the ‘Session’ variable. Significance levels: **P* < 0.05; ***P* < 0.01; ****P* < 0.001; *****P* < 0.0001. SE, standard error; CI, confidence interval.

| **Protocol 1 Group** | | | | | | | |
| --- | --- | --- | --- | --- | --- | --- | --- |
| **Mixed Linear Model Regression Results (Analysis for Assistive Effects)** | | | | | | | |
| Sample Type | Reference | Fixed Effects | Coefficient *β* (SE) | 95% CI Lower | 95% CI Upper | Statistic (z) | *P* value |
| Respiratory | Within Group (Protocol 1) | | | | | | |
|  | - | Intercept | 0.450 (0.162) | 0.133 | 0.766 | 2.78 | **0.00538 |
|  | Session #1 (BASE) | Time | 0.108 (0.0856) | −0.0602 | 0.275 | 1.26 | 0.209 |
|  |  | Session #2 | −0.0670 (0.212) | −0.483 | 0.349 | −0.316 | 0.752 |
|  |  | Session #3 | 0.0879 (0.216) | −0.335 | 0.511 | 0.407 | 0.684 |
|  |  | Session #6 | 0.125 (0.232) | −0.329 | 0.580 | 0.541 | 0.589 |
|  |  | Session #7 | 0.499 (0.217) | 0.0746 | 0.924 | 2.30 | *0.0212 |
|  | Between Groups (Protocol 2 x Session) | | | | | | |
|  | Protocol 1 | Session #1 | −0.267 (0.198) | −0.655 | 0.120 | −1.35 | 0.176 |
|  |  | Session #2 | 0.700 (0.327) | 0.0588 | 1.34 | 2.14 | *0.0324 |
|  |  | Session #3 | 1.03 (0.317) | 0.408 | 1.65 | 3.25 | **0.00117 |
|  |  | Session #6 | 1.10 (0.324) | 0.461 | 1.73 | 3.38 | ***0.000717 |
|  |  | Session #7 | 0.965 (0.279) | 0.418 | 1.51 | 3.46 | ***0.000539 |
|  | Random Effects | | Variance |  |  |  |  |
|  | Between-Participant (Intercept) | | 0.0583 |  |  |  |  |
|  | Within-Participant (Residual) | | 0.207 |  |  |  |  |
| Note: N=108 (Protocol 1: 10 participants x 5 sessions; Protocol 2: 11 participants x 5 sessions + 1 participant x 3 sessions). Log-likelihood: −75.7 | | | | | | | |
| Stool | Within Group (Protocol 1) | | | | | | |
|  | - | Intercept | 1.32 (0.0962) | 1.07 | 1.57 | 10.3 | ***8.25e-25 |
|  | Session #1 (BASE) | Time | 0.0674 (0.0529) | −0.0363 | 0.171 | 1.27 | 0.203 |
|  |  | Session #2 | −0.0020 (0.164) | −0.322 | 0.322 | −0.00119 | 0.999 |
|  |  | Session #3 | 0.0573 (0.164) | −0.265 | 0.380 | 0.349 | 0.727 |
|  |  | Session #6 | 0.0398 (0.168) | −0.290 | 0.777 | 0.237 | 0.813 |
|  |  | Session #7 | 0.451 (0.166) | 0.126 | 0.212 | 2.72 | **0.00655 |
|  | Between Groups (Protocol 2 x Session) | | | | | | |
|  | Protocol 1 | Session #1 | −0.0770 (0.147) | −0.366 | 0.212 | −0.522 | 0.601 |
|  |  | Session #2 | 0.343 (0.208) | −0.0648 | 0.751 | 1.65 | 0.0992 |
|  |  | Session #3 | 0.388 (0.205) | −0.0131 | 0.790 | 1.90 | 0.0579 |
|  |  | Session #6 | 0.651 (0.209) | 0.241 | 1.06 | 3.11 | **0.00188 |
|  |  | Session #7 | 0.384 (0.212) | −0.0308 | 0.799 | 1.81 | 0.0696 |
|  | Random Effects | | Variance |  |  |  |  |
|  | Between-Participant (Intercept) | | 0.0369 |  |  |  |  |
|  | Within-Participant (Residual) | | 0.118 |  |  |  |  |
| Note: N=122 (Protocol 1: 9 participants x 5 sessions; Protocol 2: 13 participants x 5 sessions + 4 participants x 3 sessions). Log-likelihood: −52.5 | | | | | | | |
| **Mixed Linear Model Regression Results (Analysis for Training Effects)** | | | | | | | |
| Sample Type | Reference | Fixed Effects | Coefficient *β* (SE) | 95% CI Lower | 95% CI Upper | Statistic (z) | *P* value |
| Respiratory | Within Group (Protocol 1) | | | | | | |
|  | - | Intercept | 0.309 (0.187) | −0.0567 | 0.675 | 1.66 | 0.097 |
|  | Session #1 (BASE) | Time | 0.246 (0.0984) | 0.0527 | 0.438 | 2.50 | *0.0126 |
|  |  | Session #4 | 0.289 (0.268) | −0.235 | 0.814 | 1.08 | 0.280 |
|  |  | Session #8 | 0.941 (0.245) | 0.461 | 1.42 | 3.84 | ***0.000123 |
|  | Between Groups (Protocol 2 x Session) | | | | | | |
|  | Protocol 1 | Session #1 | −0.196 (0.231) | −0.648 | 0.257 | −0.848 | 0.396 |
|  |  | Session #4 | 0.769 (0.330) | 0.122 | 1.42 | 2.33 | *0.0199 |
|  |  | Session #8 | 0.459 (0.352) | −0.232 | 1.15 | 1.30 | 0.193 |
|  | Random Effects | | Variance |  |  |  |  |
|  | Between-Participant (Intercept) | | 0.0481 |  |  |  |  |
|  | Within-Participant (Residual) | | 0.279 |  |  |  |  |
| Note: N=65 (Protocol 1: 10 participants x 3 sessions; Protocol 2: 11 participants x 3 sessions + 1 participant x 2 sessions). Log-likelihood: −54.7 | | | | | | | |
| Stool | Within Group (Protocol 1) | | | | | | |
|  | - | Intercept | 1.30 (0.134) | 1.04 | 1.56 | 9.67 | ****3.75e-22 |
|  | Session #1 (BASE) | Time | 0.153 (0.0549) | 0.0457 | 0.261 | 2.79 | **0.00524 |
|  |  | Session #4 | 0.00934 (0.189) | −0.362 | 0.380 | 0.0494 | 0.961 |
|  |  | Session #8 | 0.789 (0.186) | 0.425 | 1.15 | 4.25 | ****2.11e-05 |
|  | Between Groups (Protocol 2 x Session) | | | | | | |
|  | Protocol 1 | Session #1 | −0.151 (0.167) | −0.479 | 0.178 | −0.899 | 0.368 |
|  |  | Session #4 | 0.390 (0.229) | −0.0583 | 0.839 | 1.71 | 0.0881 |
|  |  | Session #8 | −0.0456 (0.246) | −0.527 | 0.436 | −0.186 | 0.853 |
|  | Random Effects | | Variance |  |  |  |  |
|  | Between-Participant (Intercept) | | 0.00861 |  |  |  |  |
|  | Within-Participant (Residual) | | 0.154 |  |  |  |  |
| Note: N=74 (Protocol 1: 9 participants x 3 sessions; Protocol 2: 13 participants x 3 sessions + 4 participants x 2 sessions). Log-likelihood: −37.6 | | | | | | | |

**Table S18. Categories of user handling errors: De-contamination, Sample-Transfer, Contamination-Prevention, and Documentation.**

| Categories | User handling errors |
| --- | --- |
| De-contamination | Did not wear new gloves |
|  | Did not clean the workspace with new wipes (before starting the pooling) |
|  | Did not clean the workspace with new wipes (after finishing the pooling) |
|  | Did not remove used gloves |
| Sample-Transfer | Did not identify new set of samples to be pooled |
|  | Did not use a new pooling tube |
|  | Transferred sample from a sample tube(s) that had already been pooled |
|  | Transferred sample from a sample tube(s) from the wrong set(s) |
|  | Transferred sample to the pooling tube of the wrong set |
|  | Transferred sample from a sample tube to another sample tube |
|  | Transferred sample from the pooling tube to a sample tube |
|  | Did not transfer sample from the sample tube #1 to pooling tube |
|  | Did not transfer sample from the sample tube #2 to pooling tube |
|  | Did not transfer sample from the sample tube #3 to pooling tube |
|  | Did not transfer sample from the sample tube #4 to pooling tube |
|  | Did not transfer sample from the sample tube #5 to pooling tube |
|  | Removed sample from the pooling tube |
|  | Tried to transfer/remove sample from the pooling tube |
|  | Dispensed sample outside of the pooling tube |
|  | Disposed of the sample tube after closing it |
| Contamination-Prevention | Did not prepare new pipettes before opening sample tubes |
|  | Did not open the pooling tube until dispensing sample into the tube |
|  | Did not open the sample tube and attempted to aspirate sample from it |
|  | Did not use a new pipette |
|  | Re-aspirated with a used pipette that had been dipped in the pooling tube |
|  | Did not dispose of the used pipette |
|  | Did not close the sample tube until cleaning workspace |
|  | Closed the sample tube with the pooling tube cap |
|  | Did not close the pooling tube until the end of pooling process |
|  | Closed the pooling tube with the sample tube cap |
| Documentation | Did not scan the pooling tube |
|  | Did not scan new sample tube #1 |
|  | Did not scan new sample tube #2 |
|  | Did not scan new sample tube #3 |
|  | Did not scan new sample tube #4 |
|  | Did not scan new sample tube #5 |
|  | Scanned the wrong sample tube(s) from the wrong set(s) |

**Table S19. Statistical analysis results for uncorrected minor handling errors (∑*err*_minor_) across pooling sessions for Protocol 1 group.** Matched-pairs rank biserial correlation coefficients (*r*_C_) were calculated for effect sizes using the statistic and ranks from one-sided Wilcoxon signed rank tests. Two mixed linear model regressions with maximum likelihood (ML) estimation were performed to analyze assistive effects and training effects, separately. The handling error data were transformed using Yeo-Johnson before the analyses. Dummy coding was used for the regressions. Reference categories were Protocol 1 for the ‘Group’ variable and Session #1 for the ‘Session’ variable. Significance levels: **P* < 0.05; ***P* < 0.01; ****P* < 0.001; *****P* < 0.0001. Adj. *P* value, Benjamini-Hochberg adjusted *P* value (false-discovery rate: 5%); SE, standard error; CI, confidence interval.

| **Protocol 1 Group** | | | | | | | |
| --- | --- | --- | --- | --- | --- | --- | --- |
| **Effect sizes (*r*_C_)** | | | | | | | |
| Sample Type | Reference | Test | Statistic (*T*) | Adj. *P* value | Effect size | 95% CI Lower | 95% CI Upper |
| Respiratory  (N=10) | Session #1 (BASE) | Session #2 | 15.5 | 0.891 | −0.436 | −1.000 | 0.236 |
|  |  | Session #3 | 24.5 | 0.869 | −0.109 | −0.728 | 0.600 |
|  |  | Session #4 | 21.0 | 0.872 | −0.236 | −0.891 | 0.455 |
|  |  | Session #5 | 35.0 | 0.515 | 0.273 | −0.364 | 0.836 |
|  |  | Session #6 | 30.0 | 0.698 | 0.091 | −0.600 | 0.764 |
|  |  | Session #7 | 54.5 | *0.0101 | 0.982 | 0.891 | 1.000 |
|  |  | Session #8 | 54.5 | *0.0101 | 0.982 | 0.891 | 1.000 |
| Stool  (N=9) | Session #1 (BASE) | Session #2 | 9.00 | 0.949 | −0.600 | −0.978 | 0.000 |
|  |  | Session #3 | 16.0 | 0.949 | −0.289 | −0.867 | 0.400 |
|  |  | Session #4 | 12.5 | 0.949 | −0.444 | −0.833 | 0.222 |
|  |  | Session #5 | 35.0 | 0.156 | 0.556 | −0.0222 | 0.933 |
|  |  | Session #6 | 26.5 | 0.552 | 0.178 | −0.533 | 0.778 |
|  |  | Session #7 | 42.0 | *0.0354 | 0.867 | 0.533 | 0.978 |
|  |  | Session #8 | 42.0 | *0.0354 | 0.867 | 0.533 | 0.978 |
| **Mixed Linear Model Regression Results (Analysis for Assistive Effects)** | | | | | | | |
| Sample Type | Reference | Fixed Effects | Coefficient *β* (SE) | 95% CI Lower | 95% CI Upper | Statistic (z) | *P* value |
| Respiratory | Within Group (Protocol 1) | | | | | | |
|  | - | Intercept | 0.860 (0.115) | 0.634 | 1.09 | 7.47 | ****8.19e-14 |
|  | Session #1 (BASE) | Session #2 | −0.371 (0.952) | −2.24 | 1.49 | −0.390 | 0.697 |
|  |  | Session #3 | −0.889 (0.952) | −2.75 | 0.978 | −0.933 | 0.350 |
|  |  | Session #6 | −0.748 (0.952) | −2.61 | 1.12 | −0.785 | 0.432 |
|  |  | Session #7 | −5.76 (0.952) | −7.63 | −3.90 | −6.05 | ****1.42e-09 |
|  | Between Groups (Protocol 2 x Session) | | | | | | |
|  | Protocol 1 | Session #1 | −0.0897 (0.131) | −0.347 | 0.168 | −0.682 | 0.495 |
|  |  | Session #2 | −4.04 (1.29) | −6.57 | −1.51 | −3.13 | **0.00172 |
|  |  | Session #3 | −3.92 (1.29) | −6.45 | −1.39 | −3.04 | **0.00237 |
|  |  | Session #6 | −3.79 (1.30) | −6.34 | −1.23 | −2.90 | **0.00368 |
|  |  | Session #7 | 1.41 (1.30) | −1.14 | 3.97 | 1.08 | 0.279 |
|  | Random Effects | | Variance |  |  |  |  |
|  | Between-Participant (Intercept) | | 0.0448 |  |  |  |  |
|  | Within-Participant (Residual) | | 0.0929 |  |  |  |  |
| Note: N=108 (Protocol 1: 10 participants x 5 sessions; Protocol 2: 11 participants x 5 sessions + 1 participant x 3 sessions). Log-likelihood: −34.9 | | | | | | | |
| Stool | Within Group (Protocol 1) | | | | | | |
|  | - | Intercept | 0.670 (0.115) | 0.445 | 0.895 | 5.83 | ****5.62e-09 |
|  | Session #1 (BASE) | Session #2 | 1.61 (0.937) | −0.226 | 3.45 | 1.72 | 0.0856 |
|  |  | Session #3 | 0.873 (0.937) | −0.964 | 2.71 | 0.931 | 0.352 |
|  |  | Session #6 | 0.0404 (0.937) | −1.80 | 1.88 | 0.0431 | 0.966 |
|  |  | Session #7 | −4.16 (0.937) | −5.99 | −2.32 | −4.43 | ****9.28e-06 |
|  | Between Groups (Protocol 2 x Session) | | | | | | |
|  | Protocol 1 | Session #1 | −0.0594 (0.121) | −0.296 | 0.177 | −0.493 | 0.622 |
|  |  | Session #2 | −4.86 (1.16) | −7.13 | −2.59 | −4.19 | ****2.78e-05 |
|  |  | Session #3 | −4.23 (1.16) | −6.50 | −1.95 | −3.65 | ***0.000267 |
|  |  | Session #6 | −2.75 (1.19) | −5.09 | −0.412 | −2.31 | *0.0212 |
|  |  | Session #7 | 1.34 (1.19) | −1.00 | 3.68 | 1.12 | 0.262 |
|  | Random Effects | | Variance |  |  |  |  |
|  | Between-Participant (Intercept) | | 0.0588 |  |  |  |  |
|  | Within-Participant (Residual) | | 0.0811 |  |  |  |  |
| Note: N=122 (Protocol 1: 9 participants x 5 sessions; Protocol 2: 13 participants x 5 sessions + 4 participants x 3 sessions). Log-likelihood: −35.0 | | | | | | | |
| **Mixed Linear Model Regression Results (Analysis for Training Effects)** | | | | | | | |
| Sample Type | Reference | Fixed Effects | Coefficient *β* (SE) | 95% CI Lower | 95% CI Upper | Statistic (z) | *P* value |
| Respiratory | Within Group (Protocol 1) | | | | | | |
|  | - | Intercept | 1.44 (0.242) | 0.962 | 1.91 | 5.94 | ****2.92e-09 |
|  | Session #1 (BASE) | Session #4 | 0.118 (0.975) | −1.79 | 2.03 | 0.121 | 0.904 |
|  |  | Session #8 | −5.19 (0.975) | −7.10 | −3.27 | −5.32 | ****1.05e-07 |
|  | Between Groups (Protocol 2 x Session) | | | | | | |
|  | Protocol 1 | Session #1 | −0.0788 (0.277) | −0.606 | 0.448 | −0.293 | 0.769 |
|  |  | Session #4 | −0.166 (1.32) | −2.76 | 2.42 | −0.126 | 0.900 |
|  |  | Session #8 | 3.96 (1.34) | 1.34 | 6.58 | 2.96 | **0.00305 |
|  | Random Effects | | Variance |  |  |  |  |
|  | Between-Participant (Intercept) | | 0.222 |  |  |  |  |
|  | Within-Participant (Residual) | | 0.386 |  |  |  |  |
| Note: N=65 (Protocol 1: 10 participants x 3 sessions; Protocol 2: 11 participants x 3 sessions + 1 participant x 2 sessions). Log-likelihood: −69.7 | | | | | | | |
| Stool | Within Group (Protocol 1) | | | | | | |
|  | - | Intercept | 1.41 (0.248) | 0.926 | 1.90 | 5.69 | ****1.28e-08 |
|  | Session #1 (BASE) | Session #4 | 0.937 (1.04) | −1.10 | 2.98 | 0.901 | 0.367 |
|  |  | Session #8 | −4.16 (1.04) | −6.20 | −2.13 | −4.00 | ****6.24e-05 |
|  | Between Groups (Protocol 2 x Session) | | | | | | |
|  | Protocol 1 | Session #1 | −0.377 (0.268) | −0.902 | 0.148 | −1.41 | 0.160 |
|  |  | Session #4 | 0.128 (1.29) | −2.39 | 2.65 | 0.0997 | 0.921 |
|  |  | Session #8 | 4.57 (1.33) | 1.97 | 7.17 | 3.44 | ***0.000573 |
|  | Random Effects | | Variance |  |  |  |  |
|  | Between-Participant (Intercept) | | 0.224 |  |  |  |  |
|  | Within-Participant (Residual) | | 0.395 |  |  |  |  |
| Note: N=74 (Protocol 1: 9 participants x 3 sessions; Protocol 2: 13 participants x 3 sessions + 4 participants x 2 sessions). Log-likelihood: −81.0 | | | | | | | |

**Table S20. Statistical analysis results for uncorrected severe handling errors (∑*err*_severe_) across pooling sessions for Protocol 1 group.** Matched-pairs rank biserial correlation coefficients (*r*_C_) were calculated for effect sizes using the statistic and ranks from one-sided Wilcoxon signed rank tests. Two mixed linear model regressions with maximum likelihood (ML) estimation were performed to analyze assistive effects and training effects, separately. The handling error data were transformed using Yeo-Johnson before the analyses. Dummy coding was used for the regressions. Reference categories were Protocol 1 for the ‘Group’ variable and Session #1 for the ‘Session’ variable. Significance levels: **P* < 0.05; ***P* < 0.01; ****P* < 0.001; *****P* < 0.0001. Adj. *P* value, Benjamini-Hochberg adjusted *P* value (false-discovery rate: 5%); SE, standard error; CI, confidence interval.

| **Protocol 1 Group** | | | | | | | |
| --- | --- | --- | --- | --- | --- | --- | --- |
| **Effect sizes (*r*_C_)** | | | | | | | |
| Sample Type | Reference | Test | Statistic (*T*) | Adj. *P* value | Effect size | 95% CI Lower | 95% CI Upper |
| Respiratory  (N=10) | Session #1 (BASE) | Session #2 | 35.5 | 0.236 | 0.291 | −0.255 | 0.818 |
|  |  | Session #3 | 37.0 | 0.231 | 0.345 | −0.345 | 0.891 |
|  |  | Session #4 | 28.0 | 0.480 | 0.0182 | −0.636 | 0.727 |
|  |  | Session #5 | 39.0 | 0.207 | 0.418 | −0.255 | 0.836 |
|  |  | Session #6 | 52.0 | *0.0140 | 0.891 | 0.618 | 1.000 |
|  |  | Session #7 | 54.5 | *0.0100 | 0.982 | 0.891 | 1.000 |
|  |  | Session #8 | 54.5 | *0.0100 | 0.982 | 0.891 | 1.000 |
| Stool  (N=9) | Session #1 (BASE) | Session #2 | 22.5 | 0.500 | 0.000 | −0.778 | 0.667 |
|  |  | Session #3 | 34.0 | 0.198 | 0.511 | −0.200 | 0.978 |
|  |  | Session #4 | 24.5 | 0.500 | 0.0889 | −0.578 | 0.778 |
|  |  | Session #5 | 22.5 | 0.500 | 0.000 | −0.689 | 0.711 |
|  |  | Session #6 | 25.5 | 0.500 | 0.133 | −0.533 | 0.778 |
|  |  | Session #7 | 45.0 | **0.00684 | 1.000 | 1.000 | 1.000 |
|  |  | Session #8 | 45.0 | **0.00684 | 1.000 | 1.000 | 1.000 |
| **Mixed Linear Model Regression Results (Analysis for Assistive Effects)** | | | | | | | |
| Sample Type | Reference | Fixed Effects | Coefficient *β* (SE) | 95% CI Lower | 95% CI Upper | Statistic (z) | *P* value |
| Respiratory | Within Group (Protocol 1) | | | | | | |
|  | - | Intercept | 0.978 (0.122) | 0.739 | 1.22 | 8.03 | ****9.81e-15 |
|  | Session #1 (BASE) | Session #2 | −0.432 (1.16) | −2.70 | 1.83 | −0.374 | 0.709 |
|  |  | Session #3 | −1.56 (1.16) | −3.83 | 0.700 | −1.35 | 0.176 |
|  |  | Session #6 | −2.45 (1.16) | −4.71 | −0.187 | −2.12 | *0.0338 |
|  |  | Session #7 | −6.75 (1.16) | −9.01 | −4.48 | −5.84 | ****5.15e-09 |
|  | Between Groups (Protocol 2 x Session) | | | | | | |
|  | Protocol 1 | Session #1 | −0.0440 (0.159) | −0.356 | 0.268 | −0.267 | 0.782 |
|  |  | Session #2 | −4.47 (1.56) | −7.53 | −1.40 | −2.86 | **0.00427 |
|  |  | Session #3 | −4.48 (1.56) | −7.54 | −1.41 | −2.86 | **0.00419 |
|  |  | Session #6 | −1.32 (1.58) | −4.42 | 1.78 | −0.835 | 0.404 |
|  |  | Session #7 | 1.89 (1.58) | −1.21 | 4.99 | 1.19 | 0.233 |
|  | Random Effects | | Variance |  |  |  |  |
|  | Between-Participant (Intercept) | | 0.0110 |  |  |  |  |
|  | Within-Participant (Residual) | | 0.137 |  |  |  |  |
| Note: N=108 (Protocol 1: 10 participants x 5 sessions; Protocol 2: 11 participants x 5 sessions + 1 participant x 3 sessions). Log-likelihood: −49.1 | | | | | | | |
| Stool | Within Group (Protocol 1) | | | | | | |
|  | - | Intercept | 0.781 (0.0959) | 0.593 | 0.969 | 8.14 | ****4.09e-14 |
|  | Session #1 (BASE) | Session #2 | −0.422 (0.832) | −2.05 | 1.21 | −0.507 | 0.612 |
|  |  | Session #3 | −0.977 (0.832) | −2.61 | 0.653 | −1.17 | 0.240 |
|  |  | Session #6 | −1.31 (0.832) | −2.94 | 0.316 | −1.58 | 0.114 |
|  |  | Session #7 | −5.07 (0.832) | −6.70 | −3.44 | −6.09 | ****1.10e-09 |
|  | Between Groups (Protocol 2 x Session) | | | | | | |
|  | Protocol 1 | Session #1 | −0.0710 (0.107) | −0.280 | 0.138 | −0.666 | 0.505 |
|  |  | Session #2 | −3.46 (1.03) | −5.48 | −1.45 | −3.37 | ***0.000759 |
|  |  | Session #3 | −3.40 (1.03) | −5.41 | −1.38 | −3.30 | ***0.000961 |
|  |  | Session #6 | −2.80 (1.06) | −4.87 | −0.724 | −2.64 | **0.00821 |
|  |  | Session #7 | 1.24 (1.06) | −0.836 | 3.31 | 1.17 | 0.242 |
|  | Random Effects | | Variance |  |  |  |  |
|  | Between-Participant (Intercept) | | 0.0273 |  |  |  |  |
|  | Within-Participant (Residual) | | 0.0638 |  |  |  |  |
| Note: N=122 (Protocol 1: 9 participants x 5 sessions; Protocol 2: 13 participants x 5 sessions + 4 participants x 3 sessions). Log-likelihood: −16.9 | | | | | | | |
| **Mixed Linear Model Regression Results (Analysis for Training Effects)** | | | | | | | |
| Sample Type | Reference | Fixed Effects | Coefficient *β* (SE) | 95% CI Lower | 95% CI Upper | Statistic (z) | *P* value |
| Respiratory | Within Group (Protocol 1) | | | | | | |
|  | - | Intercept | 1.35 (0.215) | 0.930 | 1.77 | 6.29 | ****3.28e-10 |
|  | Session #1 (BASE) | Session #4 | −0.434 (0.944) | −2.28 | 1.42 | −0.460 | 0.645 |
|  |  | Session #8 | −4.73 (0.944) | −6.58 | −2.88 | −5.02 | ****5.27e-07 |
|  | Between Groups (Protocol 2 x Session) | | | | | | |
|  | Protocol 1 | Session #1 | 0.0430 (0.259) | −0.465 | 0.551 | 0.166 | 0.868 |
|  |  | Session #4 | 0.0881 (1.28) | −2.42 | 2.59 | 0.0690 | 0.945 |
|  |  | Session #8 | 4.33 (1.29) | 1.79 | 6.86 | 3.35 | ***0.000812 |
|  | Random Effects | | Variance |  |  |  |  |
|  | Between-Participant (Intercept) | | 0.109 |  |  |  |  |
|  | Within-Participant (Residual) | | 0.362 |  |  |  |  |
| Note: N=65 (Protocol 1: 10 participants x 3 sessions; Protocol 2: 11 participants x 3 sessions + 1 participant x 2 sessions). Log-likelihood: −64.9 | | | | | | | |
| Stool | Within Group (Protocol 1) | | | | | | |
|  | - | Intercept | 1.21 (0.196) | 0.823 | 1.59 | 6.17 | ****6.83e-10 |
|  | Session #1 (BASE) | Session #4 | −0.839 (0.867) | −2.54 | 0.859 | −0.968 | 0.333 |
|  |  | Session #8 | −3.60 (0.867) | −5.29 | −1.90 | −4.15 | ****3.34e-05 |
|  | Between Groups (Protocol 2 x Session) | | | | | | |
|  | Protocol 1 | Session #1 | −0.157 (0.222) | −0.592 | 0.278 | −0.706 | 0.480 |
|  |  | Session #4 | 0.919 (1.07) | −1.18 | 3.02 | 0.858 | 0.391 |
|  |  | Session #8 | 1.99 (1.10) | −0.172 | 4.16 | 1.80 | 0.0713 |
|  | Random Effects | | Variance |  |  |  |  |
|  | Between-Participant (Intercept) | | 0.0869 |  |  |  |  |
|  | Within-Participant (Residual) | | 0.274 |  |  |  |  |
| Note: N=74 (Protocol 1: 9 participants x 3 sessions; Protocol 2: 13 participants x 3 sessions + 4 participants x 2 sessions). Log-likelihood: −64.3 | | | | | | | |

**Table S21. Statistical analysis results for all uncorrected handling errors (∑*err*_all_) across pooling sessions for Protocol 2 group.** Matched-pairs rank biserial correlation coefficients (*r*_C_) were calculated for effect sizes using the statistic and ranks from one-sided Wilcoxon signed-rank tests. Two mixed linear model regressions with maximum likelihood (ML) estimation were performed to analyze assistive effects and training effects, separately. The handling error data were transformed using Yeo-Johnson before the analyses. Dummy coding was used for the regressions. Reference categories were Protocol 2 for the ‘Group’ variable and Session #1 for the ‘Session’ variable. Significance levels: **P* < 0.05; ***P* < 0.01; ****P* < 0.001; *****P* < 0.0001. Adj. *P* value, Benjamini-Hochberg adjusted *P* value (false-discovery rate: 5%); SE, standard error; CI, confidence interval.

| **Protocol 2 Group** | | | | | | | |
| --- | --- | --- | --- | --- | --- | --- | --- |
| **Effect sizes (*r*_C_)** | | | | | | | |
| Sample Type | Reference | Test | Statistic (*T*) | Adj. *P* value | Effect size | 95% CI Lower | 95% CI Upper |
| Respiratory | Session #1 (BASE) | Session #2 | 78.0 | ***0.000854 | 1.000 | 1.000 | 1.000 |
| (N=12) |  | Session #3 | 78.0 | ***0.000854 | 1.000 | 1.000 | 1.000 |
|  |  | Session #4 | 42.5 | 0.391 | 0.0897 | −0.577 | 0.667 |
| (N=11) |  | Session #5 | 51.5 | 0.0694 | 0.561 | −0.0758 | 0.970 |
|  |  | Session #6 | 66.0 | **0.00114 | 1.000 | 1.000 | 1.000 |
|  |  | Session #7 | 65.5 | **0.00331 | 0.984 | 0.909 | 1.000 |
|  |  | Session #8 | 37.0 | 0.391 | 0.121 | −0.576 | 1.000 |
| Stool | Session #1 (BASE) | Session #2 | 151.5 | **0.00131 | 0.980 | 0.902 | 1.000 |
| (N=17) |  | Session #3 | 152.5 | ***0.000656 | 0.993 | 0.961 | 1.000 |
|  |  | Session #4 | 45.5 | 0.929 | −0.405 | −0.830 | 0.131 |
| (N=13) |  | Session #5 | 66.5 | 0.0823 | 0.462 | −0.143 | 0.901 |
|  |  | Session #6 | 88.0 | **0.00256 | 0.934 | 0.769 | 1.000 |
|  |  | Session #7 | 89.0 | ***0.000854 | 0.956 | 0.780 | 1.000 |
|  |  | Session #8 | 68.0 | 0.0797 | 0.495 | −0.110 | 0.912 |
| **Mixed Linear Model Regression Results (Analysis for Assistive Effects)** | | | | | | | |
| Sample Type | Reference | Fixed Effects | Coefficient *β* (SE) | 95% CI Lower | 95% CI Upper | Statistic (z) | *P* value |
| Respiratory | Within Group (Protocol 2) | | | | | | |
|  | - | Intercept | 1.64 (0.157) | 1.33 | 1.94 | 10.4 | ****2.53e-25 |
|  | Session #1 (BASE) | Session #2 | −8.60 (1.26) | −11.1 | −6.13 | −6.84 | ****7.79e-12 |
|  |  | Session #3 | −10.1 (1.26) | −12.5 | −7.62 | −8.03 | ****1.01e-15 |
|  |  | Session #6 | −7.11 (1.29) | −9.63 | −4.58 | −5.52 | ****3.42e-08 |
|  |  | Session #7 | −8.14 (1.29) | −10.7 | −5.62 | −6.32 | ****2.56e-10 |
|  | Between Groups (Protocol 1 x Session) | | | | | | |
|  | Protocol 2 | Session #1 | −0.0134 (0.190) | −0.386 | 0.359 | −0.705 | 0.944 |
|  |  | Session #2 | 8.36 (1.86) | 4.71 | 12.0 | 4.49 | ****7.24e-06 |
|  |  | Session #3 | 8.85 (1.86) | 5.20 | 12.5 | 4.75 | ****2.05e-06 |
|  |  | Session #6 | 5.63 (1.89) | 1.94 | 9.32 | 2.99 | **0.00282 |
|  |  | Session #7 | −2.67 (1.89) | −6.37 | 1.02 | −1.42 | 0.156 |
|  | Random Effects | | Variance |  |  |  |  |
|  | Between-Participant (Intercept) | | 0.103 |  |  |  |  |
|  | Within-Participant (Residual) | | 0.194 |  |  |  |  |
| Note: N=108 (Protocol 1: 10 participants x 5 sessions; Protocol 2: 11 participants x 5 sessions + 1 participant x 3 sessions). Log-likelihood: −75.1 | | | | | | | |
| Stool | Within Group (Protocol 2) | | | | | | |
|  | - | Intercept | 1.21 (0.111) | 0.993 | 1.43 | 10.9 | ****1.57e-27 |
|  | Session #1 (BASE) | Session #2 | −6.49 (0.850) | −8.16 | −4.83 | −7.64 | ****2.17e-14 |
|  |  | Session #3 | −7.18 (0.850) | −8.84 | −5.51 | −8.44 | ****3.16e-17 |
|  |  | Session #6 | −6.15 (0.922) | −7.96 | −4.34 | −6.67 | ****2.55e-11 |
|  |  | Session #7 | −6.16 (0.922) | −7.97 | −4.36 | −6.69 | ****2.29e-11 |
|  | Between Groups (Protocol 1 x Session) | | | | | | |
|  | Protocol 2 | Session #1 | 0.0729 (0.150) | −0.222 | 0.367 | 0.486 | 0.627 |
|  |  | Session #2 | 6.92 (1.44) | 4.09 | 9.75 | 4.79 | ****1.68e-06 |
|  |  | Session #3 | 6.92 (1.44) | 4.09 | 9.75 | 4.79 | ****1.68e-06 |
|  |  | Session #6 | 5.43 (1.49) | 2.52 | 8.35 | 3.65 | ***0.000261 |
|  |  | Session #7 | −2.07 (1.49) | −4.98 | 0.852 | −1.39 | 0.165 |
|  | Random Effects | | Variance |  |  |  |  |
|  | Between-Participant (Intercept) | | 0.0853 |  |  |  |  |
|  | Within-Participant (Residual) | | 0.126 |  |  |  |  |
| Note: N=122 (Protocol 1: 9 participants x 5 sessions; Protocol 2: 13 participants x 5 sessions + 4 participants x 3 sessions). Log-likelihood: −61.4 | | | | | | | |
| **Mixed Linear Model Regression Results (Analysis for Training Effects)** | | | | | | | |
| Sample Type | Reference | Fixed Effects | Coefficient *β* (SE) | 95% CI Lower | 95% CI Upper | Statistic (z) | *P* value |
| Respiratory | Within Group (Protocol 2) | | | | | | |
|  | - | Intercept | 2.53 (0.312) | 1.92 | 3.14 | 8.12 | ****4.65e-16 |
|  | Session #1 (BASE) | Session #4 | −0.781 (1.29) | −3.32 | 1.75 | −0.604 | 0.546 |
|  |  | Session #8 | −1.41 (1.33) | −4.01 | 1.18 | −1.07 | 0.286 |
|  | Between Groups (Protocol 1 x Session) | | | | | | |
|  | Protocol 2 | Session #1 | −0.123 (0.390) | −0.887 | 0.640 | −0.317 | 0.752 |
|  |  | Session #4 | 0.790 (1.92) | −2.97 | 4.55 | 0.0412 | 0.680 |
|  |  | Session #8 | −7.11 (1.92) | −10.9 | −3.31 | −3.67 | ***0.000246 |
|  | Random Effects | | Variance |  |  |  |  |
|  | Between-Participant (Intercept) | | 0.351 |  |  |  |  |
|  | Within-Participant (Residual) | | 0.814 |  |  |  |  |
| Note: N=65 (Protocol 1: 10 participants x 3 sessions; Protocol 2: 11 participants x 3 sessions + 1 participant x 2 sessions). Log-likelihood: −92.7 | | | | | | | |
| Stool | Within Group (Protocol 2) | | | | | | |
|  | - | Intercept | 1.83 (0.204) | 1.43 | 2.23 | 8.96 | ****3.26e-19 |
|  | Session #1 (BASE) | Session #4 | 0.246 (0.230) | −0.204 | 0.697 | 1.07 | 0.284 |
|  |  | Session #8 | −0.313 (0.250) | −0.803 | −0.178 | −1.25 | 0.211 |
|  | Between Groups (Protocol 1 x Session) | | | | | | |
|  | Protocol 2 | Session #1 | 0.336 (0.286) | −0.223 | 0.896 | 1.18 | 0.239 |
|  |  | Session #4 | −0.954 (1.37) | −3.64 | 1.73 | −0.696 | 0.487 |
|  |  | Session #8 | −5.34 (1.41) | −2.57 | −8.11 | −3.78 | ***0.000159 |
|  | Random Effects | | Variance |  |  |  |  |
|  | Between-Participant (Intercept) | | 0.261 |  |  |  |  |
|  | Within-Participant (Residual) | | 0.449 |  |  |  |  |
| Note: N=74 (Protocol 1: 9 participants x 3 sessions; Protocol 2: 13 participants x 3 sessions + 4 participants x 2 sessions). Log-likelihood: −85.8 | | | | | | | |

**Table S22. Statistical analysis results for average volume-transfer accuracy (Avg. Acc_pool_) across pooling sessions for Protocol 2 group.** Hedges’ g values were calculated for effect sizes. As a reference, statistical results from one-sided paired Student *t* tests are reported. Two mixed linear model regressions with maximum likelihood (ML) estimation were performed to analyze assistive effects and training effects, separately. The accuracy data were logit-transformed before the analyses. Dummy coding was used for the regressions. Reference categories were Protocol 2 for the ‘Group’ variable and Session #1 for the ‘Session’ variable. Significance levels: **P* < 0.05; ***P* < 0.01; ****P* < 0.001; *****P* < 0.0001. Adj. *P* value, Benjamini-Hochberg adjusted *P* value (false-discovery rate: 5%); SE, standard error; CI, confidence interval.

| **Protocol 2 Group** | | | | | | | |
| --- | --- | --- | --- | --- | --- | --- | --- |
| **Effect sizes (Hedges’ g)** | | | | | | | |
| Sample Type | Reference | Test | Statistic (*t*) | Adj. *P* value | Effect size | 95% CI Lower | 95% CI Upper |
| Respiratory | Session #1 (BASE) | Session #2 | −5.59 | ***0.000114 | 1.62 | 1.32 | 1.94 |
| (N=12) |  | Session #3 | −7.68 | ****0.00000838 | 2.39 | 1.76 | 3.32 |
|  |  | Session #4 | −5.10 | ***0.000202 | 1.54 | 0.848 | 2.39 |
| (N=11) |  | Session #5 | −4.70 | ***0.000422 | 1.11 | 0.450 | 1.82 |
|  |  | Session #6 | −9.55 | ****0.00000282 | 2.22 | 1.65 | 2.93 |
|  |  | Session #7 | −12.2 | ****0.00000085 | 2.55 | 1.99 | 3.79 |
|  |  | Session #8 | −9.78 | ****0.00000283 | 2.24 | 1.55 | 3.10 |
| Stool | Session #1 (BASE) | Session #2 | −5.72 | ****0. 0000551 | 0.942 | 0.473 | 1.49 |
| (N=17) |  | Session #3 | −6.46 | ****0. 0000276 | 1.40 | 0.748 | 2.09 |
|  |  | Session #4 | −3.89 | ***0. 000864 | 0.779 | 0.317 | 1.38 |
| (N=13) |  | Session #5 | −1.78 | 0. 0501 | 0.439 | −0.0291 | 1.06 |
|  |  | Session #6 | −4.27 | ***0. 000864 | 1.63 | 0.798 | 2.36 |
|  |  | Session #7 | −4.95 | ***0. 000394 | 1.98 | 1.04 | 2.81 |
|  |  | Session #8 | −4.10 | ***0. 000864 | 1.40 | 0.691 | 2.06 |
| **Mixed Linear Model Regression Results (Analysis for Assistive Effects)** | | | | | | | |
| Sample Type | Reference | Fixed Effects | Coefficient *β* (SE) | 95% CI Lower | 95% CI Upper | Statistic (z) | *P* value |
| Respiratory | Within Group (Protocol 2) | | | | | | |
|  | - | Intercept | 0.141 (0.147) | −0.147 | 0.429 | 0.957 | 0.339 |
|  | Session #1 (BASE) | Session #2 | 0.779 (0.189) | 0.408 | 1.15 | 4.12 | ****0.0000383 |
|  |  | Session #3 | 1.22 (0.189) | 0.851 | 1.59 | 6.46 | ****1.07e-10 |
|  |  | Session #6 | 1.29 (0.194) | 0.909 | 1.67 | 6.65 | ****3.02e-11 |
|  |  | Session #7 | 1.54 (0.194) | 1.16 | 1.92 | 7.95 | ****1.79e-15 |
|  | Between Groups (Protocol 1 x Session) | | | | | | |
|  | Protocol 2 | Session #1 | 0.300 (0.200) | −0.0914 | 0.691 | 1.50 | 0.133 |
|  |  | Session #2 | −0.922 (0.281) | −1.47 | −0.372 | −3.29 | **0.00102 |
|  |  | Session #3 | −1.23 (0.281) | −1.78 | −0.675 | −4.36 | ****0.0000128 |
|  |  | Session #6 | −1.30 (0.284) | −1.86 | −0.747 | −4.59 | ****4.42e-06 |
|  |  | Session #7 | −0.949 (0.284) | −1.51 | −0.393 | −3.34 | ***0.000827 |
|  | Random Effects | | Variance |  |  |  |  |
|  | Between-Participant (Intercept) | | 0.0442 |  |  |  |  |
|  | Within-Participant (Residual) | | 0.215 |  |  |  |  |
| Note: N=108 (Protocol 1: 10 participants x 5 sessions; Protocol 2: 11 participants x 5 sessions + 1 participant x 3 sessions). Log-likelihood: −76.5 | | | | | | | |
| Stool | Within Group (Protocol 2) | | | | | | |
|  | - | Intercept | 1.25 (0.097) | 1.06 | 1.45 | 12.9 | ****3.97e-38 |
|  | Session #1 (BASE) | Session #2 | 0.381 (0.118) | 0.150 | 0.612 | 3.24 | **0.00121 |
|  |  | Session #3 | 0.463 (0.118) | 0.233 | 0.694 | 3.94 | ****0.0000830 |
|  |  | Session #6 | 0.679 (0.128) | 0.429 | 0.929 | 5.32 | ****1.03e-07 |
|  |  | Session #7 | 0.821 (0.128) | 0.571 | 1.07 | 6.43 | ****1.24e-10 |
|  | Between Groups (Protocol 1 x Session) | | | | | | |
|  | Protocol 2 | Session #1 | 0.040 (0.145) | −0.243 | 0.323 | 0.277 | 0.782 |
|  |  | Session #2 | −0.415 (0.200) | −0.808 | −0.0232 | −2.08 | *0.0379 |
|  |  | Session #3 | −0.443 (0.200) | −0.835 | −0.0506 | −2.21 | *0.0269 |
|  |  | Session #6 | −0.696 (0.206) | −1.100 | −0.293 | −3.38 | ***0.000725 |
|  |  | Session #7 | −0.323 (0.206) | −0.727 | 0.0811 | −1.57 | 0.117 |
|  | Random Effects | | Variance |  |  |  |  |
|  | Between-Participant (Intercept) | | 0.0427 |  |  |  |  |
|  | Within-Participant (Residual) | | 0.118 |  |  |  |  |
| Note: N=122 (Protocol 1: 9 participants x 5 sessions; Protocol 2: 13 participants x 5 sessions + 4 participants x 3 sessions). Log-likelihood: −53.3 | | | | | | | |
| **Mixed Linear Model Regression Results (Analysis for Training Effects)** | | | | | | | |
| Sample Type | Reference | Fixed Effects | Coefficient *β* (SE) | 95% CI Lower | 95% CI Upper | Statistic (z) | *P* value |
| Respiratory | Within Group (Protocol 2) | | | | | | |
|  | - | Intercept | 0.141 (0.171) | −0.194 | 0.475 | 0.824 | 0.410 |
|  | Session #1 (BASE) | Session #4 | 0.947 (0.231) | 0.493 | 1.40 | 4.09 | ****0.0000431 |
|  |  | Session #8 | 1.21 (0.237) | 0.750 | 1.68 | 5.13 | ****2.98e-07 |
|  | Between Groups (Protocol 1 x Session) | | | | | | |
|  | Protocol 2 | Session #1 | 0.286 (0.244) | −0.192 | 0.764 | 1.17 | 0.241 |
|  |  | Session #4 | −0.971 (0.343) | −1.64 | −0.298 | −2.83 | **0.00469 |
|  |  | Session #8 | −0.112 (0.347) | −0.792 | 0.568 | −0.324 | 0.746 |
|  | Random Effects | | Variance |  |  |  |  |
|  | Between-Participant (Intercept) | | 0.0283 |  |  |  |  |
|  | Within-Participant (Residual) | | 0.321 |  |  |  |  |
| Note: N=65 (Protocol 1: 10 participants x 3 sessions; Protocol 2: 11 participants x 3 sessions + 1 participant x 2 sessions). Log-likelihood: −57.7 | | | | | | | |
| Stool | Within Group (Protocol 2) | | | | | | |
|  | - | Intercept | 1.25 (0.103) | 1.05 | 1.46 | 12.2 | ****5.19e-34 |
|  | Session #1 (BASE) | Session #4 | 0.265 (0.138) | −0.00617 | 0.537 | 1.92 | 0.0554 |
|  |  | Session #8 | 0.586 (0.150) | 0.293 | 0.879 | 3.91 | ****0.0000907 |
|  | Between Groups (Protocol 1 x Session) | | | | | | |
|  | Protocol 2 | Session #1 | 0.047 (0.169) | −0.284 | 0.378 | 0.277 | 0.782 |
|  |  | Session #4 | −0.368 (0.235) | −0.830 | 0.093 | −1.57 | 0.117 |
|  |  | Session #8 | 0.247 (0.242) | −0.227 | 0.722 | 1.02 | 0.308 |
|  | Random Effects | | Variance |  |  |  |  |
|  | Between-Participant (Intercept) | | 0.0180 |  |  |  |  |
|  | Within-Participant (Residual) | | 0.163 |  |  |  |  |
| Note: N=74 (Protocol 1: 9 participants x 3 sessions; Protocol 2: 13 participants x 3 sessions + 4 participants x 2 sessions). Log-likelihood: −41.2 | | | | | | | |

**Table S23. Statistical analysis results for uncorrected minor handling errors (∑*err*_minor_) across pooling sessions for Protocol 2 group.** Matched-pairs rank biserial correlation coefficients (*r*_C_) were calculated for effect sizes using the statistic and ranks from one-sided Wilcoxon signed rank tests. Two mixed linear model regressions with maximum likelihood (ML) estimation were performed to analyze assistive effects and training effects, separately. The handling error data were transformed using Yeo-Johnson before the analyses. Dummy coding was used for the regressions. Reference categories were Protocol 2 for the ‘Group’ variable and Session #1 for the ‘Session’ variable. Significance levels: **P* < 0.05; ***P* < 0.01; ****P* < 0.001; *****P* < 0.0001. Adj. *P* value, Benjamini-Hochberg adjusted *P* value (false-discovery rate: 5%); SE, standard error; CI, confidence interval.

| **Protocol 2 Group** | | | | | | | |
| --- | --- | --- | --- | --- | --- | --- | --- |
| **Effect sizes (*r*_C_)** | | | | | | | |
| Sample Type | Reference | Test | Statistic (*T*) | Adj. *P* value | Effect size | 95% CI Lower | 95% CI Upper |
| Respiratory | Session #1 (BASE) | Session #2 | 71.0 | *0.0104 | 0.821 | 0.436 | 0.987 |
| (N=12) |  | Session #3 | 75.0 | **0.00878 | 0.923 | 0.731 | 1.000 |
|  |  | Session #4 | 48.5 | 0.265 | 0.244 | −0.397 | 0.808 |
| (N=11) |  | Session #5 | 41.5 | 0.265 | 0.258 | −0.394 | 0.848 |
|  |  | Session #6 | 63.0 | **0.00878 | 0.909 | 0.682 | 1.000 |
|  |  | Session #7 | 63.0 | **0.00878 | 0.909 | 0.682 | 1.000 |
|  |  | Session #8 | 39.0 | 0.296 | 0.182 | −0.500 | 0.773 |
| Stool | Session #1 (BASE) | Session #2 | 145.5 | **0.00293 | 0.902 | 0.706 | 0.980 |
| (N=17) |  | Session #3 | 142.5 | **0.00293 | 0.863 | 0.641 | 0.980 |
|  |  | Session #4 | 46.5 | 0.925 | −0.392 | −0.830 | 0.118 |
| (N=13) |  | Session #5 | 55.0 | 0.351 | 0.209 | −0.385 | 0.747 |
|  |  | Session #6 | 68.5 | 0.0916 | 0.505 | −0.0549 | 0.934 |
|  |  | Session #7 | 76.0 | 0.0366 | 0.670 | 0.231 | 0.934 |
|  |  | Session #8 | 32.0 | 0.925 | −0.297 | −0.824 | 0.330 |
| **Mixed Linear Model Regression Results (Analysis for Assistive Effects)** | | | | | | | |
| Sample Type | Reference | Fixed Effects | Coefficient *β* (SE) | 95% CI Lower | 95% CI Upper | Statistic (z) | *P* value |
| Respiratory | Within Group (Protocol 2) | | | | | | |
|  | - | Intercept | 0.770 (0.107) | 0.560 | 0.980 | 7.19 | ****6.58e-13 |
|  | Session #1 (BASE) | Session #2 | −4.41 (0.869) | −6.12 | −2.71 | −5.08 | ****3.85e-07 |
|  |  | Session #3 | −4.81 (0.869) | −6.51 | −3.10 | −5.53 | ****3.18e-08 |
|  |  | Session #6 | −4.53 (0.891) | −6.28 | −2.79 | −5.09 | ****3.57e-07 |
|  |  | Session #7 | −4.35 (0.891) | −6.10 | −2.61 | −4.89 | ****1.03e-06 |
|  | Between Groups (Protocol 1 x Session) | | | | | | |
|  | Protocol 2 | Session #1 | 0.0897 (0.131) | −0.168 | 0.347 | 0.682 | 0.495 |
|  |  | Session #2 | 4.04 (1.29) | 1.51 | 6.57 | 3.13 | **0.00172 |
|  |  | Session #3 | 3.92 (1.29) | 1.39 | 6.45 | 3.04 | **0.00237 |
|  |  | Session #6 | 3.79 (1.30) | 1.23 | 6.34 | 2.90 | **0.00368 |
|  |  | Session #7 | −1.41 (1.30) | −3.97 | 1.14 | −1.08 | 0.279 |
|  | Random Effects | | Variance |  |  |  |  |
|  | Between-Participant (Intercept) | | 0.0448 |  |  |  |  |
|  | Within-Participant (Residual) | | 0.0929 |  |  |  |  |
| Note: N=108 (Protocol 1: 10 participants x 5 sessions; Protocol 2: 11 participants x 5 sessions + 1 participant x 3 sessions). Log-likelihood: −34.9 | | | | | | | |
| Stool | Within Group (Protocol 2) | | | | | | |
|  | - | Intercept | 0.611 (0.0909) | 0.433 | 0.788 | 6.73 | ****1.67e-11 |
|  | Session #1 (BASE) | Session #2 | −3.25 (0.682) | −4.58 | −1.91 | −4.76 | ****1.94e-06 |
|  |  | Session #3 | −3.35 (0.682) | −4.69 | −2.02 | −4.92 | ****8.83e-07 |
|  |  | Session #6 | −2.71 (0.740) | −4.16 | −1.26 | −3.67 | ***0.000246 |
|  |  | Session #7 | −2.82 (0.740) | −4.27 | −1.36 | −3.81 | ***0.000140 |
|  | Between Groups (Protocol 1 x Session) | | | | | | |
|  | Protocol 2 | Session #1 | 0.0594 (0.121) | −0.177 | 0.296 | 0.493 | 0.622 |
|  |  | Session #2 | 4.86 (1.16) | 2.59 | 7.13 | 4.19 | ****2.78e-05 |
|  |  | Session #3 | 4.23 (1.16) | 1.95 | 6.50 | 3.65 | ***0.000267 |
|  |  | Session #6 | 2.75 (1.19) | 0.412 | 5.09 | 2.31 | *0.0212 |
|  |  | Session #7 | −1.34 (1.19) | −3.68 | 1.00 | −1.12 | 0.262 |
|  | Random Effects | | Variance |  |  |  |  |
|  | Between-Participant (Intercept) | | 0.0588 |  |  |  |  |
|  | Within-Participant (Residual) | | 0.0811 |  |  |  |  |
| Note: N=122 (Protocol 1: 9 participants x 5 sessions; Protocol 2: 13 participants x 5 sessions + 4 participants x 3 sessions). Log-likelihood: −35.0 | | | | | | | |
| **Mixed Linear Model Regression Results (Analysis for Training Effects)** | | | | | | | |
| Sample Type | Reference | Fixed Effects | Coefficient *β* (SE) | 95% CI Lower | 95% CI Upper | Statistic (z) | *P* value |
| Respiratory | Within Group (Protocol 2) | | | | | | |
|  | - | Intercept | 1.36 (0.225) | 0.917 | 1.80 | 6.03 | ****1.63e-09 |
|  | Session #1 (BASE) | Session #4 | −0.0483 (0.890) | −1.79 | 1.70 | −0.0543 | 0.957 |
|  |  | Session #8 | −1.23 (0.914) | −3.02 | 0.564 | −1.34 | 0.179 |
|  | Between Groups (Protocol 1 x Session) | | | | | | |
|  | Protocol 2 | Session #1 | 0.0788 (0.269) | −0.448 | 0.606 | 0.293 | 0.769 |
|  |  | Session #4 | 0.166 (1.32) | −2.42 | 2.76 | 0.126 | 0.900 |
|  |  | Session #8 | −3.96 (1.34) | −6.58 | −1.34 | −2.93 | **0.00305 |
|  | Random Effects | | Variance |  |  |  |  |
|  | Between-Participant (Intercept) | | 0.222 |  |  |  |  |
|  | Within-Participant (Residual) | | 0.386 |  |  |  |  |
| Note: N=65 (Protocol 1: 10 participants x 3 sessions; Protocol 2: 11 participants x 3 sessions + 1 participant x 2 sessions). Log-likelihood: −69.7 | | | | | | | |
| Stool | Within Group (Protocol 2) | | | | | | |
|  | - | Intercept | 1.04 (0.191) | 0.662 | 1.41 | 5.43 | ****5.60e-08 |
|  | Session #1 (BASE) | Session #4 | 1.07 (0.757) | −0.418 | 2.55 | 1.41 | 0.159 |
|  |  | Session #8 | 0.409 (0.825) | −1.21 | 2.03 | 0.496 | 0.620 |
|  | Between Groups (Protocol 1 x Session) | | | | | | |
|  | Protocol 2 | Session #1 | 0.377 (0.268) | −0.148 | 0.902 | 1.41 | 0.160 |
|  |  | Session #4 | −0.128 (1.29) | −2.65 | 2.39 | −0.0997 | 0.921 |
|  |  | Session #8 | −4.57 (1.33) | −7.17 | −1.97 | −3.44 | ***0.000573 |
|  | Random Effects | | Variance |  |  |  |  |
|  | Between-Participant (Intercept) | | 0.224 |  |  |  |  |
|  | Within-Participant (Residual) | | 0.395 |  |  |  |  |
| Note: N=74 (Protocol 1: 9 participants x 3 sessions; Protocol 2: 13 participants x 3 sessions + 4 participants x 2 sessions). Log-likelihood: −81.0 | | | | | | | |

**Table S24. Statistical analysis results for uncorrected severe handling errors (∑*err*_severe_) across pooling sessions for Protocol 2 group.** Matched-pairs rank biserial correlation coefficients (*r*_C_) were calculated for effect sizes using the statistic and ranks from one-sided Wilcoxon signed rank tests. Two mixed linear model regressions with maximum likelihood (ML) estimation were performed to analyze assistive effects and training effects, separately. The handling error data were transformed using Yeo-Johnson before the analyses. Dummy coding was used for the regressions. Reference categories were Protocol 2 for the ‘Group’ variable and Session #1 for the ‘Session’ variable. Significance levels: **P* < 0.05; ***P* < 0.01; ****P* < 0.001; *****P* < 0.0001. Adj. *P* value, Benjamini-Hochberg adjusted *P* value (false-discovery rate: 5%); SE, standard error; CI, confidence interval.

| **Protocol 2 Group** | | | | | | | |
| --- | --- | --- | --- | --- | --- | --- | --- |
| **Effect sizes (*r*_C_)** | | | | | | | |
| Sample Type | Reference | Test | Statistic (*T*) | Adj. *P* value | Effect size | 95% CI Lower | 95% CI Upper |
| Respiratory | Session #1 (BASE) | Session #2 | 75.0 | **0.00748 | 0.923 | 0.731 | 1.000 |
| (N=12) |  | Session #3 | 76.5 | **0.00748 | 0.962 | 0.808 | 1.000 |
|  |  | Session #4 | 42.5 | 0.391 | 0.0897 | −0.629 | 0.679 |
| (N=11) |  | Session #5 | 49.5 | 0.0988 | 0.500 | −0.136 | 0.924 |
|  |  | Session #6 | 62.5 | **0.00748 | 0.894 | 0.576 | 1.000 |
|  |  | Session #7 | 63.0 | **0.00748 | 0.909 | 0.682 | 1.000 |
|  |  | Session #8 | 41.5 | 0.261 | 0.258 | −0.439 | 0.803 |
| Stool | Session #1 (BASE) | Session #2 | 148.0 | **0.00120 | 0.935 | 0.765 | 0.993 |
| (N=17) |  | Session #3 | 150.0 | **0.00120 | 0.961 | 0.863 | 1.000 |
|  |  | Session #4 | 58.0 | 0.810 | −0.242 | −0.739 | 0.307 |
| (N=13) |  | Session #5 | 68.5 | 0.0622 | 0.505 | −0.0659 | 0.934 |
|  |  | Session #6 | 88.0 | **0.00255 | 0.934 | 0.703 | 1.000 |
|  |  | Session #7 | 88.0 | **0.00255 | 0.934 | 0.703 | 1.000 |
|  |  | Session #8 | 82.5 | **0.00594 | 0.813 | 0.439 | 1.000 |
| **Mixed Linear Model Regression Results (Analysis for Assistive Effects)** | | | | | | | |
| Sample Type | Reference | Fixed Effects | Coefficient *β* (SE) | 95% CI Lower | 95% CI Upper | Statistic (z) | *P* value |
| Respiratory | Within Group (Protocol 2) | | | | | | |
|  | - | Intercept | 0.934 (0.111) | 0.716 | 1.15 | 8.41 | ****4.01e-17 |
|  | Session #1 (BASE) | Session #2 | −4.90 (1.05) | −6.97 | −2.83 | −4.65 | ****3.36e-06 |
|  |  | Session #3 | −6.04 (1.05) | −8.11 | −3.98 | −5.73 | ****9.97e-09 |
|  |  | Session #6 | −3.77 (1.08) | −5.89 | −1.65 | −3.49 | ***0.000491 |
|  |  | Session #7 | −4.86 (1.08) | −6.98 | −2.74 | −4.49 | ****7.12e-06 |
|  | Between Groups (Protocol 1 x Session) | | | | | | |
|  | Protocol 2 | Session #1 | 0.0440 (0.159) | −0.268 | 0.356 | 0.267 | 0.782 |
|  |  | Session #2 | 4.47 (1.56) | 1.40 | 7.53 | 2.86 | **0.00427 |
|  |  | Session #3 | 4.48 (1.56) | 1.41 | 7.54 | 2.86 | **0.00419 |
|  |  | Session #6 | 1.32 (1.58) | −1.78 | 4.42 | 0.835 | 0.404 |
|  |  | Session #7 | −1.89 (1.58) | −4.99 | 1.21 | −1.19 | 0.233 |
|  | Random Effects | | Variance |  |  |  |  |
|  | Between-Participant (Intercept) | | 0.0110 |  |  |  |  |
|  | Within-Participant (Residual) | | 0.137 |  |  |  |  |
| Note: N=108 (Protocol 1: 10 participants x 5 sessions; Protocol 2: 11 participants x 5 sessions + 1 participant x 3 sessions). Log-likelihood: −49.1 | | | | | | | |
| Stool | Within Group (Protocol 2) | | | | | | |
|  | - | Intercept | 0.710 (0.0732) | 0.566 | 0.853 | 9.70 | ****3.14e-22 |
|  | Session #1 (BASE) | Session #2 | −3.89 (0.605) | −5.07 | −2.70 | −6.42 | ****1.36e-10 |
|  |  | Session #3 | −4.37 (0.605) | −5.56 | −3.19 | −7.23 | ****4.97e-13 |
|  |  | Session #6 | −4.11 (0.655) | −5.40 | −2.83 | −6.27 | ****3.51e-10 |
|  |  | Session #7 | −3.83 (0.655) | −5.11 | −2.54 | −5.84 | ****5.18e-09 |
|  | Between Groups (Protocol 1 x Session) | | | | | | |
|  | Protocol 2 | Session #1 | 0.0710 (0.107) | −0.138 | 0.280 | 0.666 | 0.505 |
|  |  | Session #2 | 3.46 (1.03) | 1.45 | 5.48 | 3.37 | ***0.000759 |
|  |  | Session #3 | 3.40 (1.03) | 1.38 | 5.41 | 3.30 | ***0.000961 |
|  |  | Session #6 | 2.80 (1.06) | 0.724 | 4.87 | 2.64 | **0.00821 |
|  |  | Session #7 | −1.24 (1.06) | −3.31 | 0.836 | −1.17 | 0.242 |
|  | Random Effects | | Variance |  |  |  |  |
|  | Between-Participant (Intercept) | | 0.0273 |  |  |  |  |
|  | Within-Participant (Residual) | | 0.0638 |  |  |  |  |
| Note: N=122 (Protocol 1: 9 participants x 5 sessions; Protocol 2: 13 participants x 5 sessions + 4 participants x 3 sessions). Log-likelihood: −16.9 | | | | | | | |
| **Mixed Linear Model Regression Results (Analysis for Training Effects)** | | | | | | | |
| Sample Type | Reference | Fixed Effects | Coefficient *β* (SE) | 95% CI Lower | 95% CI Upper | Statistic (z) | *P* value |
| Respiratory | Within Group (Protocol 2) | | | | | | |
|  | - | Intercept | 1.39 (0.198) | 1.01 | 1.78 | 7.04 | ****1.94e-12 |
|  | Session #1 (BASE) | Session #4 | −0.346 (0.861) | −2.03 | 1.34 | −0.402 | 0.688 |
|  |  | Session #8 | −0.405 (0.883) | −2.14 | 1.33 | −0.459 | 0.646 |
|  | Between Groups (Protocol 1 x Session) | | | | | | |
|  | Protocol 2 | Session #1 | −0.0430 (0.259) | −0.551 | 0.465 | −0.166 | 0.868 |
|  |  | Session #4 | −0.0881 (1.28) | −2.59 | 2.42 | −0.0690 | 0.945 |
|  |  | Session #8 | −4.33 (1.29) | −6.86 | −1.79 | −3.35 | ***0.000812 |
|  | Random Effects | | Variance |  |  |  |  |
|  | Between-Participant (Intercept) | | 0.109 |  |  |  |  |
|  | Within-Participant (Residual) | | 0.362 |  |  |  |  |
| Note: N=65 (Protocol 1: 10 participants x 3 sessions; Protocol 2: 11 participants x 3 sessions + 1 participant x 2 sessions). Log-likelihood: −64.9 | | | | | | | |
| Stool | Within Group (Protocol 2) | | | | | | |
|  | - | Intercept | 1.05 (0.146) | 0.764 | 1.34 | 7.20 | ****5.99e-13 |
|  | Session #1 (BASE) | Session #4 | 0.0802 (0.631) | −1.16 | 1.32 | 0.127 | 0.899 |
|  |  | Session #8 | −1.60 (0.684) | −2.94 | −0.263 | −2.34 | *0.0190 |
|  | Between Groups (Protocol 1 x Session) | | | | | | |
|  | Protocol 2 | Session #1 | 0.157 (0.222) | −0.278 | 0.592 | 0.706 | 0.480 |
|  |  | Session #4 | −0.919 (1.07) | −3.02 | 1.18 | −0.858 | 0.391 |
|  |  | Session #8 | −1.99 (1.10) | −4.16 | 0.172 | −1.80 | 0.0713 |
|  | Random Effects | | Variance |  |  |  |  |
|  | Between-Participant (Intercept) | | 0.0869 |  |  |  |  |
|  | Within-Participant (Residual) | | 0.274 |  |  |  |  |
| Note: N=74 (Protocol 1: 9 participants x 3 sessions; Protocol 2: 13 participants x 3 sessions + 4 participants x 2 sessions). Log-likelihood: −64.3 | | | | | | | |

**Table S25. Linear mixed model analyses (with continuous pooling round variable) for completion time in the Protocol 2 group.** Completion time data were standardized. Significance levels: **P* < 0.05; ***P* < 0.01; ****P* < 0.001; *****P* < 0.0001. SE, standard error; CI, confidence interval.

| **Completion Time ~ Device + Pooling Round + Device × Pooling Round + (1\|Participant)** | | | | | | |
| --- | --- | --- | --- | --- | --- | --- |
| Sample Type | Fixed Effects | Coefficient *β* (SE) | 95% CI Lower | 95% CI Upper | Statistic (z) | *P* value |
| Respiratory | Intercept | −0.241 (0.126) | −0.489 | 0.00634 | −1.91 | 0.0562 |
|  | Device | 1.44 (0.134) | 1.17 | 1.70 | 10.7 | ****1,16e-26 |
|  | Pooling Round | −0.0253 (0.00465) | −0.0344 | −0.0162 | −5.45 | ****5.17e-08 |
|  | Device x Pooling Round | −0.00664 (0.00726) | −0.0209 | 0.00759 | −0.914 | 0.360 |
|  | Combined *β* |  |  |  |  |  |
|  | Pooling Round x (1 + Device) | −0.0319 (0.00564) | −0.0430 | −0.0209 | −5.67 | ****1.45e-08 |
|  | Random Effects | Variance |  |  |  |  |
|  | Between-Participant (Intercept) | 0.0984 |  |  |  |  |
|  | Within-Participant (Residual) | 0.397 |  |  |  |  |
| Note: N=368 (11 participants x 32 rounds + 1 participants x 16 rounds). Log-likelihood: −375 | | | | | | |
| Stool | Intercept | 0.132 (0.184) | −0.230 | 0.493 | 0.713 | 0.476 |
|  | Device | 0.752 (0.0972) | 0.562 | 0.943 | 7.74 | ****9.67e-15 |
|  | Pooling Round | −0.0324 (0.00352) | −0.0393 | −0.0255 | −9.19 | ****3.87e-20 |
|  | Device x Pooling Round | −0.00535 (0.00544) | −0.0160 | 0.00532 | −0.982 | 0.326 |
|  | Combined *β* |  |  |  |  |  |
|  | Pooling Round x (1 + Device) | −0.0377 (0.00431) | −0.0462 | −0.0293 | −8.76 | ****2.04e-18 |
|  | Random Effects | Variance |  |  |  |  |
|  | Between-Participant (Intercept) | 0.510 |  |  |  |  |
|  | Within-Participant (Residual) | 0.287 |  |  |  |  |
| Note: N=480 (13 participants x 32 rounds + 4 participants x 16 rounds). Log-likelihood: −425 | | | | | | |

**Table S26. Linear mixed model analyses (with continuous pooling round variable) for volume-transfer accuracy (Acc_pool_), controlling for completion time in the Protocol 2 group.** Accuracy data were logit-transformed, and completion time data were standardized. Significance levels: **P* < 0.05; ***P* < 0.01; ****P* < 0.001; *****P* < 0.0001. SE, standard error; CI, confidence interval.

| **Accuracy ~ Device + Pooling Round + Completion Time + Device × Pooling Round + Device × Completion Time + (1\|Participant)** | | | | | | |
| --- | --- | --- | --- | --- | --- | --- |
| Sample Type | Fixed Effects | Coefficient *β* (SE) | 95% CI Lower | 95% CI Upper | Statistic (z) | *P* value |
| Respiratory | Intercept | 0.228 (0.179) | −0.122 | 0.578 | 1.28 | 0.202 |
|  | Device | 0.743 (0.135) | 0.478 | 1.01 | 5.49 | ****4.00e-08 |
|  | Pooling Round | 0.0479 (0.00411) | 0.0399 | 0.0560 | 11.7 | ****1.77e-31 |
|  | Completion Time | 0.181 (0.0571) | 0.0692 | 0.293 | 3.17 | **0.00151 |
|  | Device x Pooling Round | −0.0202 (0.00648) | −0.0329 | −0.00751 | −3.12 | **0.00181 |
|  | Device x Completion Time | −0.240 (0.0831) | −0.403 | −0.0775 | −2.89 | **0.00382 |
|  | Combined *β* |  |  |  |  |  |
|  | Pooling Round x (1 + Device) | 0.0277 (0.00506) | 0.0178 | 0.0376 | 5.48 | ****4.21e-08 |
|  | Completion Time x (1 + Device) | −0.0593 (0.0636) | −0.184 | 0.0654 | −0.932 | 0.351 |
|  | Random Effects | Variance |  |  |  |  |
|  | Between-Participant (Intercept) | 0.318 |  |  |  |  |
|  | Within-Participant (Residual) | 0.268 |  |  |  |  |
| Note: N=368 (11 participants x 32 rounds + 1 participants x 16 rounds). Log-likelihood: −314 | | | | | | |
| Stool | Intercept | 1.20 (0.0730) | 1.06 | 1.35 | 16.5 | ****3.54e-61 |
|  | Device | 0.243 (0.0800) | 0.0864 | 0.400 | 3.04 | **0.00236 |
|  | Pooling Round | 0.0236 (0.00288) | 0.0179 | 0.0292 | 8.18 | ****2.76e-16 |
|  | Completion Time | 0.149 (0.0369) | 0.0763 | 0.221 | 4.02 | ****5.71e-05 |
|  | Device x Pooling Round | −0.000085 (0.00430) | −0.00850 | 0.00833 | −0.0199 | 0.984 |
|  | Device x Completion Time | −0.0438 (0.0416) | −0.125 | 0.0378 | −1.05 | 0.293 |
|  | Combined *β* |  |  |  |  |  |
|  | Pooling Round x (1 + Device) | 0.0235 (0.00353) | 0.0166 | 0.0304 | 6.66 | ****2.75e-11 |
|  | Completion Time x (1 + Device) | 0.105 (0.0399) | 0.0266 | 0.183 | 2.63 | **0.00863 |
|  | Random Effects | Variance |  |  |  |  |
|  | Between-Participant (Intercept) | 0.0513 |  |  |  |  |
|  | Within-Participant (Residual) | 0.162 |  |  |  |  |
| Note: N=480 (13 participants x 32 rounds + 4 participants x 16 rounds). Log-likelihood: −279 | | | | | | |

**Table S27. Linear mixed model analyses (with continuous pooling round variable) for all handling (*err*_all_), minor (*err*_minor_), and severe errors (*err*_severe_), controlling for completion time in the Protocol 2 group.** Handling error data were Yeo-Johnson transformed, and completion time data were standardized. Significance levels: **P* < 0.05; ***P* < 0.01; ****P* < 0.001; *****P* < 0.0001. SE, standard error; CI, confidence interval.

| **AllErrors ~ Device + Pooling Round + Completion Time + Device × Pooling Round + Device × Completion Time + (1\|Participant)** | | | | | | |
| --- | --- | --- | --- | --- | --- | --- |
| Sample Type | Fixed Effects | Coefficient *β* (SE) | 95% CI Lower | 95% CI Upper | Statistic (z) | *P* value |
| Respiratory  Note: N=368  (11 participants x 32 rounds + 1 participants x 16 rounds).  Log-likelihood: −83.7 | Intercept | 0.639 (0.0750) | 0.492 | 0.786 | 8.52 | ****1.59e-17 |
|  | Device | −0.509 (0.0721) | −0.651 | −0.368 | −7.06 | ****1.66e-12 |
|  | Pooling Round | −0.00408 (0.00219) | −0.00837 | 0.000211 | −1.86 | 0.0624 |
|  | Completion Time | −0.0396 (0.0304) | −0.0992 | 0.0200 | −1.30 | 0.193 |
|  | Device x Pooling Round | 0.00777 (0.00345) | 0.00101 | 0.0145 | 2.25 | *0.0243 |
|  | Device x Completion Time | −0.0359 (0.0443) | −0.123 | 0.0509 | −0.811 | 0.417 |
|  | Combined *β* |  |  |  |  |  |
|  | Pooling Round x (1 + Device) | 0.00369 (0.00270) | −0.00159 | 0.00898 | 1.37 | 0.171 |
|  | Completion Time x (1 + Device) | −0.0755 (0.0339) | −0.142 | −0.00906 | −2.23 | *0.0259 |
|  | Random Effects | Variance |  |  |  |  |
|  | Between-Participant (Intercept) | 0.0491 |  |  |  |  |
|  | Within-Participant (Residual) | 0.0763 |  |  |  |  |
| Stool | Intercept | 0.541 (0.0551) | 0.433 | 0.649 | 9.82 | ****8.93e-23 |
|  | Device | −0.437 (0.0487) | −0.533 | −0.342 | −8.98 | ****2.76e-19 |
| Note: N=480  (13 participants x 32 rounds + 4 participants x 16 rounds).  Log-likelihood: −46.6 | Pooling Round | −0.00695 (0.00176) | −0.0104 | −0.00350 | −3.95 | ****7.77e-05 |
|  | Completion Time | −0.0605 (0.0231) | −0.106 | −0.0153 | −2.62 | **0.00869 |
|  | Device x Pooling Round | 0.00730 (0.00260) | 0.00220 | 0.0124 | 2.80 | **0.00505 |
|  | Device x Completion Time | 0.0231 (0.0252) | −0.0263 | 0.0726 | 0.918 | 0.359 |
|  | Combined *β* |  |  |  |  |  |
|  | Pooling Round x (1 + Device) | 0.000348 (0.00216) | −0.00464 | 0.0442 | 0.161 | 0.872 |
|  | Completion Time x (1 + Device) | −0.0374 (0.0250) | 0.0344 | 0.174 | −1.49 | 0.135 |
|  | Random Effects | Variance |  |  |  |  |
|  | Between-Participant (Intercept) | 0.0372 |  |  |  |  |
|  | Within-Participant (Residual) | 0.0594 |  |  |  |  |
| **MinorErrors ~ Device + Pooling Round + Completion Time + Device × Pooling Round + Device × Completion Time + (1\|Participant)** | | | | | | |
| Sample Type | Fixed Effects | Coefficient *β* (SE) | 95% CI Lower | 95% CI Upper | Statistic (z) | *P* value |
| Respiratory | Intercept | 0.322 (0.0470) | 0.230 | 0.414 | 6.85 | ****7.38e-12 |
|  | Device | −0.227 (0.0422) | −0.310 | −0.144 | −5.38 | ****7.57e-08 |
| Note: N=368  (11 participants x 32 rounds + 1 participants x 16 rounds).  Log-likelihood: 109 | Pooling Round | −0.00404 (0.00128) | −0.00655 | −0.00152 | −3.15 | **0.00164 |
|  | Completion Time | −0.0436 (0.0178) | −0.0785 | −0.00864 | −2.45 | *0.0145 |
|  | Device x Pooling Round | 0.00413 (0.00202) | 0.000165 | 0.00809 | 2.04 | *0.0412 |
|  | Device x Completion Time | −0.0296 (0.0259) | −0.0804 | 0.0212 | −1.14 | 0.253 |
|  | Combined *β* |  |  |  |  |  |
|  | Pooling Round x (1 + Device) | 9.08e-05 (0.00158) | −0.00300 | 0.00319 | 0.0575 | 0.954 |
|  | Completion Time x (1 + Device) | −0.0732 (0.0199) | −0.112 | −0.0343 | −3.69 | ***0.000227 |
|  | Random Effects | Variance |  |  |  |  |
|  | Between-Participant (Intercept) | 0.0202 |  |  |  |  |
|  | Within-Participant (Residual) | 0.0262 |  |  |  |  |
| Stool | Intercept | 0.243 (0.0379) | 0.169 | 0.317 | 6.41 | ****1.47e-10 |
|  | Device | −0.189 (0.0328) | −0.253 | −0.125 | −5.77 | ****8.11e-09 |
| Note: N=480  (13 participants x 32 rounds + 4 participants x 16 rounds).  Log-likelihood: 140 | Pooling Round | −0.00124 (0.00118) | −0.00356 | 0.00108 | −1.05 | 0.294 |
|  | Completion Time | −0.0590 (0.0155) | −0.0895 | −0.0286 | −3.80 | ***0.000144 |
|  | Device x Pooling Round | 0.00215 (0.00175) | −0.00128 | 0.00559 | 1.23 | 0.219 |
|  | Device x Completion Time | 0.0399 (0.0170) | 0.00660 | 0.0732 | 2.35 | *0.0188 |
|  | Combined *β* |  |  |  |  |  |
|  | Pooling Round x (1 + Device) | 0.000913 (0.00145) | −0.00193 | 0.00376 | 0.628 | 0.530 |
|  | Completion Time x (1 + Device) | −0.0191 (0.0168) | −0.0521 | 0.0139 | −1.14 | 0.256 |
|  | Random Effects | Variance |  |  |  |  |
|  | Between-Participant (Intercept) | 0.0179 |  |  |  |  |
|  | Within-Participant (Residual) | 0.0269 |  |  |  |  |
| **SevereErrors ~ Device + Pooling Round + Completion Time + Device × Pooling Round + Device × Completion Time + (1\|Participant)** | | | | | | |
| Sample Type | Fixed Effects | Coefficient *β* (SE) | 95% CI Lower | 95% CI Upper | Statistic (z) | *P* value |
| Respiratory | Intercept | 0.267 (0.0384) | 0.192 | 0.342 | 6.96 | ****3.45e-12 |
|  | Device | −0.225 (0.0465) | −0.316 | −0.134 | −4.83 | ****1.34e-06 |
| Note: N=368  (11 participants x 32 rounds + 1 participants x 16 rounds).  Log-likelihood: 78.3 | Pooling Round | −0.000437 (0.00141) | −0.00321 | 0.00233 | −0.309 | 0.757 |
|  | Completion Time | −0.0213 (0.0196) | −0.0596 | 0.0171 | −1.08 | 0.278 |
|  | Device x Pooling Round | 0.00226 (0.00223) | −0.00212 | 0.00663 | 1.01 | 0.312 |
|  | Device x Completion Time | 0.00646 (0.0286) | −0.0497 | 0.0626 | 0.226 | 0.821 |
|  | Combined *β* |  |  |  |  |  |
|  | Pooling Round x (1 + Device) | 0.00182 (0.00174) | −0.00159 | 0.00523 | 1.05 | 0.296 |
|  | Completion Time x (1 + Device) | −0.0148 (0.0219) | −0.0576 | 0.0280 | −0.677 | 0.499 |
|  | Random Effects | Variance |  |  |  |  |
|  | Between-Participant (Intercept) | 0.0100 |  |  |  |  |
|  | Within-Participant (Residual) | 0.0318 |  |  |  |  |
| Stool | Intercept | 0.241 (0.0274) | 0.187 | 0.295 | 8.78 | ****1.62e-18 |
|  | Device | −0.185 (0.0279) | −0.240 | −0.130 | −6.62 | ****3.49e-11 |
| Note: N=480  (13 participants x 32 rounds + 4 participants x 16 rounds).  Log-likelihood: 219 | Pooling Round | −0.00521 (0.00101) | −0.00718 | −0.00324 | −5.18 | ****2.26e-07 |
|  | Completion Time | −0.0212 (0.0131) | −0.0468 | 0.00440 | −1.62 | 0.105 |
|  | Device x Pooling Round | 0.00436 (0.00149) | 0.00143 | 0.00728 | 2.91 | **0.00357 |
|  | Device x Completion Time | −0.00693 (0.0145) | −0.0353 | 0.0214 | −0.478 | 0.632 |
|  | Combined *β* |  |  |  |  |  |
|  | Pooling Round x (1 + Device) | −0.000856 (0.00123) | −0.00328 | 0.00156 | −0.693 | 0.488 |
|  | Completion Time x (1 + Device) | −0.0281 (0.0142) | −0.0559 | −0.000378 | −1.99 | *0.0470 |
|  | Random Effects | Variance |  |  |  |  |
|  | Between-Participant (Intercept) | 0.00806 |  |  |  |  |
|  | Within-Participant (Residual) | 0.0196 |  |  |  |  |

**Table S28. Linear mixed model analyses of all uncorrected handling errors (∑*err*_all_) controlling for completion time across pooling sessions for Protocol 2 group.** Two mixed linear model regressions with maximum likelihood (ML) estimation were performed to analyze assistive effects and training effects, separately, with completion time as a covariate. Before the analyses, the handling error data were Yeo-Johnson transformed, and completion time data were standardized. Dummy coding was used for the regressions. Reference categories were Protocol 2 for the ‘Group’ variable and Session #1 for the ‘Session’ variable. Significance levels: **P* < 0.05; ***P* < 0.01; ****P* < 0.001; *****P* < 0.0001. SE, standard error; CI, confidence interval.

| **Protocol 2 Group** | | | | | | | |
| --- | --- | --- | --- | --- | --- | --- | --- |
| **Mixed Linear Model Regression Results (Analysis for Assistive Effects)** | | | | | | | |
| Sample Type | Reference | Fixed Effects | Coefficient *β* (SE) | 95% CI Lower | 95% CI Upper | Statistic (z) | *P* value |
| Respiratory | Within Group (Protocol 2) | | | | | | |
|  | - | Intercept | 1.64 (0.160) | 1.33 | 1.96 | 10.3 | ****9.00e-25 |
|  | Session #1 (BASE) | Time | 0.0206 (0.0832) | −0.143 | 0.184 | 0.247 | 0.805 |
|  |  | Session #2 | −8.79 (1.49) | −11.7 | −5.88 | −5.92 | ****3.22e-09 |
|  |  | Session #3 | −10.2 (1.38) | −12.9 | −7.52 | −7.40 | ****1.35e-13 |
|  |  | Session #6 | −7.19 (1.34) | −9.81 | −4.58 | −5.38 | ****7.28e-08 |
|  |  | Session #7 | −8.25 (1.35) | −10.9 | −5.59 | −6.10 | ****1.09e-09 |
|  | Between Groups (Protocol 1 x Session) | | | | | | |
|  | Protocol 2 | Session #1 | −0.0202 (0.192) | −0.397 | 0.356 | −0.105 | 0.916 |
|  |  | Session #2 | 8.66 (2.22) | 4.31 | 13.0 | 3.90 | ****9.55e-05 |
|  |  | Session #3 | 9.11 (2.15) | 4.90 | 13.3 | 4.24 | ****2.20e-05 |
|  |  | Session #6 | 5.90 (2.19) | 1.61 | 10.2 | 2.70 | **0.00698 |
|  |  | Session #7 | −2.70 (1.89) | −6.40 | 1.01 | −1.43 | 0.154 |
|  | Random Effects | | Variance |  |  |  |  |
|  | Between-Participant (Intercept) | | 0.0998 |  |  |  |  |
|  | Within-Participant (Residual) | | 0.195 |  |  |  |  |
| Note: N=108 (Protocol 1: 10 participants x 5 sessions; Protocol 2: 11 participants x 5 sessions + 1 participant x 3 sessions). Log-likelihood: −75.1 | | | | | | | |
| Stool | Within Group (Protocol 2) | | | | | | |
|  | - | Intercept | 1.22 (0.113) | 0.996 | 1.44 | 10.8 | ****3.77e-27 |
|  | Session #1 (BASE) | Time | −0.0201 (0.0608) | −0.139 | 0.0990 | −0.331 | 0.741 |
|  |  | Session #2 | −6.42 (0.881) | −8.14 | −4.69 | −7.28 | ****3.39e-13 |
|  |  | Session #3 | −7.14 (0.856) | −8.81 | −5.46 | −8.34 | ****7.32e-17 |
|  |  | Session #6 | −6.17 (0.923) | −7.98 | −4.36 | −6.69 | ****2.54e-11 |
|  |  | Session #7 | −6.19 (0.925) | −8.01 | −4.38 | −6.70 | ****2.10e-11 |
|  | Between Groups (Protocol 1 x Session) | | | | | | |
|  | Protocol 2 | Session #1 | 0.0618 (0.154) | −0.239 | 0.363 | 0.402 | 0.687 |
|  |  | Session #2 | 6.77 (1.51) | 3.80 | 9.73 | 4.48 | ****7.61e-06 |
|  |  | Session #3 | 6.80 (1.48) | 3.90 | 9.71 | 4.59 | ****4.42e-06 |
|  |  | Session #6 | 5.33 (1.51) | 2.37 | 8.30 | 3.53 | ***0.000420 |
|  |  | Session #7 | −1.94 (1.53) | −4.94 | 1.07 | −1.26 | 0.206 |
|  | Random Effects | | Variance |  |  |  |  |
|  | Between-Participant (Intercept) | | 0.0872 |  |  |  |  |
|  | Within-Participant (Residual) | | 0.125 |  |  |  |  |
| Note: N=122 (Protocol 1: 9 participants x 5 sessions; Protocol 2: 13 participants x 5 sessions + 4 participants x 3 sessions). Log-likelihood: −61.3 | | | | | | | |
| **Mixed Linear Model Regression Results (Analysis for Training Effects)** | | | | | | | |
| Sample Type | Reference | Fixed Effects | Coefficient *β* (SE) | 95% CI Lower | 95% CI Upper | Statistic (z) | *P* value |
| Respiratory | Within Group (Protocol 2) | | | | | | |
|  | - | Intercept | 2.55 (0.314) | 1.93 | 3.17 | 8.12 | ****4.82e-16 |
|  | Session #1 (BASE) | Time | −0.180 (0.171) | −0.515 | 0.155 | −1.05 | 0.293 |
|  |  | Session #4 | −1.07 (1.30) | −3.61 | 1.48 | −0.823 | 0.411 |
|  |  | Session #8 | −1.90 (1.38) | −4.61 | 0.806 | −1.38 | 0.169 |
|  | Between Groups (Protocol 1 x Session) | | | | | | |
|  | Protocol 2 | Session #1 | −0.0578 (0.388) | −0.818 | 0.702 | −0.149 | 0.881 |
|  |  | Session #4 | 0.270 (1.95) | −3.54 | 4.08 | 0.139 | 0.889 |
|  |  | Session #8 | −6.21 (2.09) | −10.3 | −2.12 | −2.98 | **0.00291 |
|  | Random Effects | | Variance |  |  |  |  |
|  | Between-Participant (Intercept) | | 0.396 |  |  |  |  |
|  | Within-Participant (Residual) | | 0.784 |  |  |  |  |
| Note: N=65 (Protocol 1: 10 participants x 3 sessions; Protocol 2: 11 participants x 3 sessions + 1 participant x 2 sessions). Log-likelihood: −92.2 | | | | | | | |
| Stool | Within Group (Protocol 2) | | | | | | |
|  | - | Intercept | 2.03 (0.222) | 1.59 | 2.46 | 9.11 | ****8.21e-20 |
|  | Session #1 (BASE) | Time | −0.280 (0.125) | −0.525 | −0.0346 | −2.24 | *0.0253 |
|  |  | Session #4 | 0.00477 (0.851) | −1.66 | 1.67 | 0.00560 | 0.996 |
|  |  | Session #8 | −2.10 (0.942) | −3.95 | −0.256 | −2.23 | *0.0257 |
|  | Between Groups (Protocol 1 x Session) | | | | | | |
|  | Protocol 2 | Session #1 | 0.157 (0.281) | −0.395 | 0.709 | 0.558 | 0.577 |
|  |  | Session #4 | −0.815 (1.29) | −3.35 | 1.72 | −0.630 | 0.528 |
|  |  | Session #8 | −4.06 (1.45) | −6.90 | −1.21 | −2.79 | **0.00521 |
|  | Random Effects | | Variance |  |  |  |  |
|  | Between-Participant (Intercept) | | 0.314 |  |  |  |  |
|  | Within-Participant (Residual) | | 0.398 |  |  |  |  |
| Note: N=74 (Protocol 1: 9 participants x 3 sessions; Protocol 2: 13 participants x 3 sessions + 4 participants x 2 sessions). Log-likelihood: −83.3 | | | | | | | |

**Table S29. Linear mixed model analyses of average volume-transfer accuracy (Avg. Acc_pool_) controlling for completion time across pooling sessions for Protocol 2 group.** Two mixed linear model regressions with maximum likelihood (ML) estimation were performed to analyze assistive effects and training effects, separately, with completion time as a covariate. Before the analyses, the accuracy data were logit-transformed, and completion time data were standardized. Dummy coding was used for the regressions. Reference categories were Protocol 2 for the ‘Group’ variable and Session #1 for the ‘Session’ variable. Significance levels: **P* < 0.05; ***P* < 0.01; ****P* < 0.001; *****P* < 0.0001. SE, standard error; CI, confidence interval.

| **Protocol 2 Group** | | | | | | | |
| --- | --- | --- | --- | --- | --- | --- | --- |
| **Mixed Linear Model Regression Results (Analysis for Assistive Effects)** | | | | | | | |
| Sample Type | Reference | Fixed Effects | Coefficient *β* (SE) | 95% CI Lower | 95% CI Upper | Statistic (z) | *P* value |
| Respiratory | Within Group (Protocol 2) | | | | | | |
|  | - | Intercept | 0.182 (0.152) | −0.116 | 0.481 | 1.20 | 0.231 |
|  | Session #1 (BASE) | Time | 0.108 (0.0856) | −0.0602 | 0.275 | 1.26 | 0.209 |
|  |  | Session #2 | 0.633 (0.219) | 0.204 | 1.06 | 2.89 | **0.00384 |
|  |  | Session #3 | 1.12 (0.204) | 0.717 | 1.52 | 5.48 | ****4.33e-08 |
|  |  | Session #6 | 1.22 (0.198) | 0.833 | 1.61 | 6.18 | ****6.59e-10 |
|  |  | Session #7 | 1.46 (0.200) | 1.07 | 1.86 | 7.31 | ****2.59e-13 |
|  | Between Groups (Protocol 1 x Session) | | | | | | |
|  | Protocol 2 | Session #1 | 0.267 (0.198) | −0.120 | 0.655 | 1.35 | 0.176 |
|  |  | Session #2 | −0.700 (0.327) | −1.34 | −0.0588 | −2.14 | *0.0324 |
|  |  | Session #3 | −1.03 (0.317) | −1.65 | −0.408 | −3.25 | **0.00117 |
|  |  | Session #6 | −1.10 (0.324) | −1.73 | −0.461 | −3.38 | ***0.000717 |
|  |  | Session #7 | −0.965 (0.279) | −1.51 | −0.418 | −3.46 | ***0.000539 |
|  | Random Effects | | Variance |  |  |  |  |
|  | Between-Participant (Intercept) | | 0.0583 |  |  |  |  |
|  | Within-Participant (Residual) | | 0.207 |  |  |  |  |
| Note: N=108 (Protocol 1: 10 participants x 5 sessions; Protocol 2: 11 participants x 5 sessions + 1 participant x 3 sessions). Log-likelihood: −75.7 | | | | | | | |
| Stool | Within Group (Protocol 2) | | | | | | |
|  | - | Intercept | 1.24 (0.0962) | 1.05 | 1.43 | 12.9 | ****5.97e-38 |
|  | Session #1 (BASE) | Time | 0.0674 (0.0529) | −0.0363 | 0.171 | 1.27 | 0.203 |
|  |  | Session #2 | 0.343 (0.122) | 0.105 | 0.581 | 2.82 | **0.00478 |
|  |  | Session #3 | 0.446 (0.119) | 0.213 | 0.678 | 3.76 | ***0.000172 |
|  |  | Session #6 | 0.691 (0.128) | 0.440 | 0.941 | 5.40 | ****6.65e-08 |
|  |  | Session #7 | 0.835 (0.128) | 0.584 | 1.09 | 6.52 | ****6.87e-11 |
|  | Between Groups (Protocol 1 x Session) | | | | | | |
|  | Protocol 2 | Session #1 | 0.0770 (0.147) | −0.212 | 0.366 | 0.522 | 0.601 |
|  |  | Session #2 | −0.343 (0.208) | −0.751 | 0.0648 | −1.65 | 0.0992 |
|  |  | Session #3 | −0.388 (0.205) | −0.790 | 0.0131 | −1.90 | 0.0579 |
|  |  | Session #6 | −0.651 (0.209) | −1.06 | −0.241 | −3.11 | **0.00188 |
|  |  | Session #7 | −0.384 (0.212) | −0.799 | 0.0308 | −1.81 | 0.0696 |
|  | Random Effects | | Variance |  |  |  |  |
|  | Between-Participant (Intercept) | | 0.0369 |  |  |  |  |
|  | Within-Participant (Residual) | | 0.118 |  |  |  |  |
| Note: N=122 (Protocol 1: 9 participants x 5 sessions; Protocol 2: 13 participants x 5 sessions + 4 participants x 3 sessions). Log-likelihood: −52.5 | | | | | | | |
| **Mixed Linear Model Regression Results (Analysis for Training Effects)** | | | | | | | |
| Sample Type | Reference | Fixed Effects | Coefficient *β* (SE) | 95% CI Lower | 95% CI Upper | Statistic (z) | *P* value |
| Respiratory | Within Group (Protocol 2) | | | | | | |
|  | - | Intercept | 0.114 (0.166) | −0.211 | 0.438 | 0.686 | 0.493 |
|  | Session #1 (BASE) | Time | 0.246 (0.0984) | 0.0527 | 0.438 | 2.50 | *0.0126 |
|  |  | Session #4 | 1.06 (0.220) | 0.626 | 1.49 | 4.80 | ****1.57e-06 |
|  |  | Session #8 | 1.40 (0.233) | 0.943 | 1.86 | 6.00 | ****1.94e-09 |
|  | Between Groups (Protocol 1 x Session) | | | | | | |
|  | Protocol 2 | Session #1 | 0.196 (0.231) | −0.257 | 0.648 | 0.848 | 0.396 |
|  |  | Session #4 | −0.769 (0.330) | −1.42 | −0.122 | −2.33 | *0.0199 |
|  |  | Session #8 | −0.459 (0.352) | −1.15 | 0.232 | −1.30 | 0.193 |
|  | Random Effects | | Variance |  |  |  |  |
|  | Between-Participant (Intercept) | | 0.0481 |  |  |  |  |
|  | Within-Participant (Residual) | | 0.279 |  |  |  |  |
| Note: N=65 (Protocol 1: 10 participants x 3 sessions; Protocol 2: 11 participants x 3 sessions + 1 participant x 2 sessions). Log-likelihood: −54.7 | | | | | | | |
| Stool | Within Group (Protocol 2) | | | | | | |
|  | - | Intercept | 1.15 (0.105) | 0.943 | 1.35 | 11.0 | ****6.61e-28 |
|  | Session #1 (BASE) | Time | 0.153 (0.0549) | 0.0457 | 0.261 | 2.79 | **0.00524 |
|  |  | Session #4 | 0.399 (0.143) | 0.120 | 0.680 | 2.80 | **0.00517 |
|  |  | Session #8 | 0.744 (0.155) | 0.439 | 1.05 | 4.79 | ****1.68e-06 |
|  | Between Groups (Protocol 1 x Session) | | | | | | |
|  | Protocol 2 | Session #1 | 0.151 (0.167) | −0.178 | 0.479 | 0.899 | 0.368 |
|  |  | Session #4 | −0.390 (0.229) | −0.839 | 0.0583 | −1.71 | 0.0881 |
|  |  | Session #8 | 0.0456 (0.246) | −0.436 | 0.527 | 0.186 | 0.853 |
|  | Random Effects | | Variance |  |  |  |  |
|  | Between-Participant (Intercept) | | 0.00861 |  |  |  |  |
|  | Within-Participant (Residual) | | 0.154 |  |  |  |  |
| Note: N=74 (Protocol 1: 9 participants x 3 sessions; Protocol 2: 13 participants x 3 sessions + 4 participants x 2 sessions). Log-likelihood: −37.6 | | | | | | | |

**Table S30. Statistical analysis results for the number of high-quality pools across pooling sessions for Protocol 1 group.** Matched-pairs rank biserial correlation coefficients (*r*_C_) were calculated for effect sizes using the statistic and ranks from one-sided Wilcoxon signed rank tests. Two mixed linear model regressions with maximum likelihood (ML) estimation were performed to analyze assistive effects and training effects, separately. The number of pools data were transformed using Yeo-Johnson before the analyses. Dummy coding was used for the regressions. Reference categories were Protocol 1 for the ‘Group’ variable and Session #1 for the ‘Session’ variable. Significance levels: **P* < 0.05; ***P* < 0.01; ****P* < 0.001; *****P* < 0.0001. Adj. *P* value, Benjamini-Hochberg adjusted *P* value (false-discovery rate: 5%); SE, standard error; CI, confidence interval.

| **Protocol 1 Group** | | | | | | | |
| --- | --- | --- | --- | --- | --- | --- | --- |
| **Effect sizes (*r*_C_)** | | | | | | | |
| Sample Type | Reference | Test | Statistic (*T*) | Adj. *P* value | Effect size | 95% CI Lower | 95% CI Upper |
| Respiratory | Session #1 (BASE) | Session #2 | 41.0 | 0.924 | 0.491 | 0.000 | 0.818 |
| (N=10) |  | Session #3 | 32.0 | 0.924 | 0.164 | −0.345 | 0.618 |
|  |  | Session #4 | 37.0 | 0.924 | 0.345 | 0.000 | 0.727 |
|  |  | Session #5 | 32.5 | 0.924 | 0.182 | 0.000 | 0.491 |
|  |  | Session #6 | 27.0 | 0.924 | −0.0182 | −0.491 | 0.491 |
|  |  | Session #7 | 5.00 | *0.0364 | −0.818 | −0.982 | −0.491 |
|  |  | Session #8 | 5.00 | *0.0364 | −0.818 | −0.982 | −0.491 |
| Stool  (N=9) | Session #1 (BASE) | Session #2 | 22.5 | 0.500 | 0.000 | −0.667 | 0.667 |
|  |  | Session #3 | 18.0 | 0.294 | −0.200 | −0.778 | 0.489 |
|  |  | Session #4 | 23.0 | 0.525 | 0.0222 | −0.533 | 0.533 |
|  |  | Session #5 | 10.5 | 0.0709 | −0.533 | −0.867 | −0.200 |
|  |  | Session #6 | 15.0 | 0.181 | −0.333 | −0.780 | 0.333 |
|  |  | Session #7 | 5.50 | *0.0215 | −0.756 | −1.000 | −0.200 |
|  |  | Session #8 | 3.00 | **0.00994 | −0.867 | −0.978 | −0.533 |
| **Mixed Linear Model Regression Results (Analysis for Assistive Effects)** | | | | | | | |
| Sample Type | Reference | Fixed Effects | Coefficient *β* (SE) | 95% CI Lower | 95% CI Upper | Statistic (z) | *P* value |
| Respiratory | Within Group (Protocol 1) | | | | | | |
|  | - | Intercept | 0.163 (0.0728) | 0.0199 | 0.305 | 2.23 | *0.0255 |
|  | Session #1 (BASE) | Session #2 | −1.07 (0.661) | −2.37 | 0.222 | −1.62 | 0.104 |
|  |  | Session #3 | −0.326 (0.661) | −1.62 | 0.970 | −0.493 | 0.622 |
|  |  | Session #6 | −0.257 (0.661) | −1.55 | 1.04 | −0.389 | 0.697 |
|  |  | Session #7 | 1.44 (0.661) | 0.149 | 2.74 | 2.18 | *0.0289 |
|  | Between Groups (Protocol 2 x Session) | | | | | | |
|  | Protocol 1 | Session #1 | −0.163 (0.0911) | −0.341 | 0.0160 | −1.78 | 0.0744 |
|  |  | Session #2 | 2.75 (0.895) | 0.998 | 4.51 | 3.07 | **0.00211 |
|  |  | Session #3 | 3.31 (0.895) | 1.55 | 5.06 | 3.69 | ***0.000223 |
|  |  | Session #6 | 3.61 (0.905) | 1.83 | 5.38 | 3.98 | ****0.0000683 |
|  |  | Session #7 | 2.31 (0.905) | 0.538 | 4.09 | 2.55 | *0.0107 |
|  | Random Effects | | Variance |  |  |  |  |
|  | Between-Participant (Intercept) | | 0.00875 |  |  |  |  |
|  | Within-Participant (Residual) | | 0.0448 |  |  |  |  |
| Note: N=108 (Protocol 1: 10 participants x 5 sessions; Protocol 2: 11 participants x 5 sessions + 1 participant x 3 sessions). Log-likelihood: 8.39 | | | | | | | |
| Stool | Within Group (Protocol 1) | | | | | | |
|  | - | Intercept | 2.90 (0.668) | 1.59 | 4.21 | 4.34 | ****0.0000144 |
|  | Session #1 (BASE) | Session #2 | −0.840 (5.77) | −12.1 | 10.5 | −0.146 | 0.884 |
|  |  | Session #3 | −0.840 (5.77) | −12.1 | 10.5 | −0.146 | 0.884 |
|  |  | Session #6 | 1.49 (5.77) | −9.82 | 12.8 | 0.258 | 0.796 |
|  |  | Session #7 | 10.6 (5.77) | −0.749 | 21.9 | 1.83 | 0.0672 |
|  | Between Groups (Protocol 2 x Session) | | | | | | |
|  | Protocol 1 | Session #1 | −0.488 (0.739) | −1.935 | 0.960 | −0.660 | 0.509 |
|  |  | Session #2 | 16.3 (7.13) | 2.28 | 30.2 | 2.28 | *0.0227 |
|  |  | Session #3 | 20.4 (7.13) | 6.43 | 34.4 | 2.86 | **0.00422 |
|  |  | Session #6 | 22.1 (7.35) | 7.75 | 36.5 | 3.02 | **0.00257 |
|  |  | Session #7 | 16.2 (7.35) | 1.82 | 30.6 | 2.21 | *0.0272 |
|  | Random Effects | | Variance |  |  |  |  |
|  | Between-Participant (Intercept) | | 1.37 |  |  |  |  |
|  | Within-Participant (Residual) | | 3.07 |  |  |  |  |
| Note: N=122 (Protocol 1: 9 participants x 5 sessions; Protocol 2: 13 participants x 5 sessions + 4 participants x 3 sessions). Log-likelihood: −253 | | | | | | | |
| **Mixed Linear Model Regression Results (Analysis for Training Effects)** | | | | | | | |
| Sample Type | Reference | Fixed Effects | Coefficient *β* (SE) | 95% CI Lower | 95% CI Upper | Statistic (z) | *P* value |
| Respiratory | Within Group (Protocol 1) | | | | | | |
|  | - | Intercept | 0.132 (0.0615) | 0.0118 | 0.253 | 2.15 | *0.0314 |
|  | Session #1 (BASE) | Session #4 | −0.321 (0.304) | −0.917 | 0.276 | −1.05 | 0.292 |
|  |  | Session #8 | 0.796 (0.304) | 0.200 | 1.39 | 2.62 | **0.00890 |
|  | Between Groups (Protocol 2 x Session) | | | | | | |
|  | Protocol 1 | Session #1 | −0.132 (0.0832) | −0.295 | 0.0306 | −1.59 | 0.111 |
|  |  | Session #4 | 0.925 (0.412) | 0.117 | 1.73 | 2.24 | *0.0249 |
|  |  | Session #8 | 0.505 (0.416) | −0.311 | 1.32 | 1.21 | 0.226 |
|  | Random Effects | | Variance |  |  |  |  |
|  | Between-Participant (Intercept) | | 4.40e-11 |  |  |  |  |
|  | Within-Participant (Residual) | | 0.0376 |  |  |  |  |
| Note: N=65 (Protocol 1: 10 participants x 3 sessions; Protocol 2: 11 participants x 3 sessions + 1 participant x 2 sessions). Log-likelihood: 14.4 | | | | | | | |
| Stool | Within Group (Protocol 1) | | | | | | |
|  | - | Intercept | 2.08 (0.534) | 1.03 | 3.13 | 3.89 | ***0.000100 |
|  | Session #1 (BASE) | Session #4 | −0.558 (2.61) | −5.68 | 4.56 | −0.214 | 0.831 |
|  |  | Session #8 | 8.78 (2.61) | 3.66 | 13.9 | 3.36 | ***0.000772 |
|  | Between Groups (Protocol 2 x Session) | | | | | | |
|  | Protocol 1 | Session #1 | −0.211 (0.654) | −1.49 | 1.07 | −0.323 | 0.746 |
|  |  | Session #4 | 4.81 (3.23) | −1.52 | 11.1 | 1.49 | 0.136 |
|  |  | Session #8 | −1.49 (3.32) | −7.99 | 5.01 | −0.449 | 0.653 |
|  | Random Effects | | Variance |  |  |  |  |
|  | Between-Participant (Intercept) | | 0.0785 |  |  |  |  |
|  | Within-Participant (Residual) | | 2.49 |  |  |  |  |
| Note: N=74 (Protocol 1: 9 participants x 3 sessions; Protocol 2: 13 participants x 3 sessions + 4 participants x 2 sessions). Log-likelihood: −140 | | | | | | | |

**Table S31. Statistical analysis results for the number of high-quality pools across pooling sessions for Protocol 2 group.** Matched-pairs rank biserial correlation coefficients (*r*_C_) were calculated for effect sizes using the statistic and ranks from one-sided Wilcoxon signed rank tests. Two mixed linear model regressions with maximum likelihood (ML) estimation were performed to analyze assistive effects and training effects, separately. The number of pools data were transformed using Yeo-Johnson before the analyses. Dummy coding was used for the regressions. Reference categories were Protocol 2 for the ‘Group’ variable and Session #1 for the ‘Session’ variable. Significance levels: **P* < 0.05; ***P* < 0.01; ****P* < 0.001; *****P* < 0.0001. Adj. *P* value, Benjamini-Hochberg adjusted *P* value (false-discovery rate: 5%); SE, standard error; CI, confidence interval.

| **Protocol 2 Group** | | | | | | | |
| --- | --- | --- | --- | --- | --- | --- | --- |
| **Effect sizes (*r*_C_)** | | | | | | | |
| Sample Type | Reference | Test | Statistic (*T*) | Adj. *P* value | Effect size | 95% CI Lower | 95% CI Upper |
| Respiratory | Session #1 (BASE) | Session #2 | 14.0 | *0.0309 | −0.641 | −0.872 | −0.295 |
| (N=12) |  | Session #3 | 5.00 | **0.00637 | −0.872 | −0.987 | −0.641 |
|  |  | Session #4 | 18.0 | *0.0442 | −0.538 | −0.808 | −0.154 |
| (N=11) |  | Session #5 | 14.0 | *0.0442 | −0.576 | −0.848 | −0.167 |
|  |  | Session #6 | 1.50 | **0.00637 | −0.955 | −1.000 | −0.773 |
|  |  | Session #7 | 1.50 | **0.00637 | −0.955 | −1.000 | −0.773 |
|  |  | Session #8 | 3.00 | **0.00637 | −0.909 | −1.000 | −0.682 |
| Stool | Session #1 (BASE) | Session #2 | 12.0 | **0.00248 | −0.843 | −0.980 | −0.555 |
| (N=17) |  | Session #3 | 3.00 | **0.00161 | −0.961 | −1.000 | −0.863 |
|  |  | Session #4 | 27.5 | *0.0108 | −0.640 | −0.935 | −0.190 |
| (N=13) |  | Session #5 | 21.0 | *0.0402 | −0.538 | −0.934 | 0.000 |
|  |  | Session #6 | 3.00 | **0.00248 | −0.934 | −1.000 | −0.769 |
|  |  | Session #7 | 3.00 | **0.00248 | −0.934 | −1.000 | −0.769 |
|  |  | Session #8 | 10.5 | **0.00972 | −0.769 | −0.967 | −0.385 |
| **Mixed Linear Model Regression Results (Analysis for Assistive Effects)** | | | | | | | |
| Sample Type | Reference | Fixed Effects | Coefficient *β* (SE) | 95% CI Lower | 95% CI Upper | Statistic (z) | *P* value |
| Respiratory | Within Group (Protocol 2) | | | | | | |
|  | - | Intercept | −1.93e-16 (0.0668) | −0.131 | 0.131 | −2.89e-15 | 1.000 |
|  | Session #1 (BASE) | Session #2 | 1.68 (0.604) | 0.495 | 2.86 | 2.78 | **0.00544 |
|  |  | Session #3 | 2.98 (0.604) | 1.80 | 4.16 | 4.94 | ****8.01e-07 |
|  |  | Session #6 | 3.35 (0.618) | 2.14 | 4.56 | 5.41 | ****6.14e-08 |
|  |  | Session #7 | 3.76 (0.618) | 2.55 | 4.97 | 6.08 | ****1.23e-09 |
|  | Between Groups (Protocol 1 x Session) | | | | | | |
|  | Protocol 2 | Session #1 | 1.63 (0.0911) | −0.0160 | 0.341 | 1.78 | 0.0744 |
|  |  | Session #2 | −2.75 (7.13) | −4.51 | −0.998 | −3.07 | **0.00211 |
|  |  | Session #3 | −3.31 (7.13) | −5.06 | −1.55 | −3.69 | ***0.000223 |
|  |  | Session #6 | −3.61 (7.35) | −5.38 | −1.83 | −3.98 | ****0.0000683 |
|  |  | Session #7 | −2.31 (7.35) | −4.09 | −0.538 | −2.55 | *0.0107 |
|  | Random Effects | | Variance |  |  |  |  |
|  | Between-Participant (Intercept) | | 0.00875 |  |  |  |  |
|  | Within-Participant (Residual) | | 0.0448 |  |  |  |  |
| Note: N=108 (Protocol 1: 10 participants x 5 sessions; Protocol 2: 11 participants x 5 sessions + 1 participant x 3 sessions). Log-likelihood: 8.39 | | | | | | | |
| Stool | Within Group (Protocol 2) | | | | | | |
|  | - | Intercept | 2.41 (0.511) | 1.41 | 3.41 | 4.71 | ****2.45e-06 |
|  | Session #1 (BASE) | Session #2 | 15.4 (4.20) | 7.19 | 23.6 | 3.67 | ***0.000239 |
|  |  | Session #3 | 19.6 (4.20) | 11.3 | 27.8 | 4.66 | ****3.11e-06 |
|  |  | Session #6 | 23.6 (4.55) | 14.7 | 32.6 | 5.20 | ****2.02e-07 |
|  |  | Session #7 | 26.8 (4.55) | 17.9 | 35.7 | 5.89 | ****3.93e-09 |
|  | Between Groups (Protocol 1 x Session) | | | | | | |
|  | Protocol 2 | Session #1 | 0.488 (0.739) | −0.960 | 1.935 | 0.660 | 0.509 |
|  |  | Session #2 | −16.3 (7.13) | −30.2 | −2.28 | −2.28 | *0.0227 |
|  |  | Session #3 | −20.4 (7.13) | −34.4 | −6.43 | −2.86 | **0.00422 |
|  |  | Session #6 | −22.1 (7.35) | −36.5 | −7.75 | −3.02 | **0.00257 |
|  |  | Session #7 | −16.2 (7.35) | −30.6 | −1.82 | −2.21 | *0.0272 |
|  | Random Effects | | Variance |  |  |  |  |
|  | Between-Participant (Intercept) | | 1.37 |  |  |  |  |
|  | Within-Participant (Residual) | | 3.07 |  |  |  |  |
| Note: N=122 (Protocol 1: 9 participants x 5 sessions; Protocol 2: 13 participants x 5 sessions + 4 participants x 3 sessions). Log-likelihood: −253 | | | | | | | |
| **Mixed Linear Model Regression Results (Analysis for Training Effects)** | | | | | | | |
| Sample Type | Reference | Fixed Effects | Coefficient *β* (SE) | 95% CI Lower | 95% CI Upper | Statistic (z) | *P* value |
| Respiratory | Within Group (Protocol 2) | | | | | | |
|  | - | Intercept | 2.24e-16 (0.0560) | −0.110 | 0.110 | 4.01e-15 | 1.000 |
|  | Session #1 (BASE) | Session #4 | 0.604 (0.278) | 0.0594 | 1.15 | 2.17 | *0.0297 |
|  |  | Session #8 | 1.30 (0.284) | 0.744 | 1.86 | 4.58 | ****4.70e-06 |
|  | Between Groups (Protocol 1 x Session) | | | | | | |
|  | Protocol 2 | Session #1 | 0.132 (0.0832) | −0.0306 | 0.295 | 1.59 | 0.111 |
|  |  | Session #4 | −0.925 (0.412) | −1.73 | −0.117 | −2.24 | *0.0249 |
|  |  | Session #8 | −0.505 (0.416) | −1.32 | 0.311 | −1.21 | 0.226 |
|  | Random Effects | | Variance |  |  |  |  |
|  | Between-Participant (Intercept) | | 3.60e-11 |  |  |  |  |
|  | Within-Participant (Residual) | | 0.0376 |  |  |  |  |
| Note: N=65 (Protocol 1: 10 participants x 3 sessions; Protocol 2: 11 participants x 3 sessions + 1 participant x 2 sessions). Log-likelihood: 14.4 | | | | | | | |
| Stool | Within Group (Protocol 2) | | | | | | |
|  | - | Intercept | 1.87 (0.389) | 1.11 | 2.63 | 4.80 | ****1.56e-06 |
|  | Session #1 (BASE) | Session #4 | 4.25 (1.90) | 0.531 | 7.98 | 2.24 | *0.0251 |
|  |  | Session #8 | 7.29 (2.05) | 3.28 | 11.3 | 3.56 | ***0.000368 |
|  | Between Groups (Protocol 1 x Session) | | | | | | |
|  | Protocol 2 | Session #1 | 0.211 (0.654) | −1.07 | 1.49 | 0.323 | 0.746 |
|  |  | Session #4 | −4.81 (3.23) | −11.1 | 1.52 | −1.49 | 0.136 |
|  |  | Session #8 | 1.49 (3.32) | −5.01 | 7.99 | 0.449 | 0.653 |
|  | Random Effects | | Variance |  |  |  |  |
|  | Between-Participant (Intercept) | | 0.0785 |  |  |  |  |
|  | Within-Participant (Residual) | | 2.49 |  |  |  |  |
| Note: N=74 (Protocol 1: 9 participants x 3 sessions; Protocol 2: 13 participants x 3 sessions + 4 participants x 2 sessions). Log-likelihood: −140 | | | | | | | |

**Table S32.** **Statistical analysis results for the number of Protocol 2 participants generating high-quality pools across pooling sessions.** McNemar's exact test was used to compare the proportion of participants who generated ≥2 high-quality pools through high-performance volume transfer between the first pooling session (#1, BASE) and subsequent sessions. Adjusted *P* values indicate *P* values adjusted for multiple comparisons using the Benjamini-Hochberg procedure with 5% false-discovery rate. Significance levels: **P* < 0.05; ***P* < 0.01; ****P* < 0.001; *****P* < 0.0001.

| **Protocol 2 (Artificial Respiratory Samples)** | | | |
| --- | --- | --- | --- |
| Comparison | Test statistic | *P* value | Adjusted *P* value |
| #1 (BASE) vs #2 (ASSIST_1_) | 0.00 | 0.125 | 0.150 |
| #1 (BASE) vs #3 (ASSIST_2_) | 0.00 | *0.0156 | *0.0313 |
| #1 (BASE) vs #4 (AFTER_1_) | 0.00 | 0.250 | 0.250 |
| #1 (BASE) vs #6 (ASSIST_3_) | 0.00 | 0.0625 | 0.0938 |
| #1 (BASE) vs #7 (ASSIST_4_) | 0.00 | **0.00781 | *0.0313 |
| #1 (BASE) vs #8 (AFTER_2_) | 0.00 | *0.0156 | *0.0313 |
| **Protocol 2 (Artificial Stool Samples)** | | | |
| Comparison | Test statistic | *P* value | Adjusted *P* value |
| #1 (BASE) vs #2 (ASSIST_1_) | 0.00 | *0.0313 | *0.0469 |
| #1 (BASE) vs #3 (ASSIST_2_) | 0.00 | **0.00781 | *0.0313 |
| #1 (BASE) vs #4 (AFTER_1_) | 1.00 | 0.219 | 0.219 |
| #1 (BASE) vs #6 (ASSIST_3_) | 0.00 | *0.0156 | *0.0313 |
| #1 (BASE) vs #7 (ASSIST_4_) | 0.00 | *0.0156 | *0.0313 |
| #1 (BASE) vs #8 (AFTER_2_) | 1.00 | 0.0703 | 0.0844 |

**Table S33. Statistical analysis results for the number of high-performed pooling rounds across pooling sessions for Protocol 1 group.** Matched-pairs rank biserial correlation coefficients (*r*_C_) were calculated for effect sizes using the statistic and ranks from one-sided Wilcoxon signed rank tests. Two mixed linear model regressions with maximum likelihood (ML) estimation were performed to analyze assistive effects and training effects, separately. The number of rounds data were transformed using Yeo-Johnson before the analyses. Dummy coding was used for the regressions. Reference categories were Protocol 1 for the ‘Group’ variable and Session #1 for the ‘Session’ variable. Significance levels: **P* < 0.05; ***P* < 0.01; ****P* < 0.001; *****P* < 0.0001. Adj. *P* value, Benjamini-Hochberg adjusted *P* value (false-discovery rate: 5%); SE, standard error; CI, confidence interval.

| **Protocol 1 Group** | | | | | | | |
| --- | --- | --- | --- | --- | --- | --- | --- |
| **Effect sizes (*r*_C_)** | | | | | | | |
| Sample Type | Reference | Test | Statistic (*T*) | Adj. *P* value | Effect size | 95% CI Lower | 95% CI Upper |
| Respiratory  (N=10) | Session #1 (BASE) | Session #2 | 27.5 | 0.500 | 0.000 | 0.000 | 0.000 |
|  |  | Session #3 | 27.5 | 0.500 | 0.000 | 0.000 | 0.000 |
|  |  | Session #4 | 27.5 | 0.500 | 0.000 | 0.000 | 0.000 |
|  |  | Session #5 | 27.5 | 0.500 | 0.000 | 0.000 | 0.000 |
|  |  | Session #6 | 27.5 | 0.500 | 0.000 | 0.000 | 0.000 |
|  |  | Session #7 | 3.00 | *0.0207 | −0.891 | −1.000 | −0.618 |
|  |  | Session #8 | 3.00 | *0.0207 | −0.891 | −1.000 | −0.618 |
| Stool  (N=9) | Session #1 (BASE) | Session #2 | 31.0 | 0.856 | 0.378 | 0.000 | 0.778 |
|  |  | Session #3 | 22.5 | 0.856 | 0.000 | 0.000 | 0.000 |
|  |  | Session #4 | 31.0 | 0.856 | 0.378 | 0.000 | 0.778 |
|  |  | Session #5 | 27.0 | 0.856 | 0.200 | 0.000 | 0.533 |
|  |  | Session #6 | 22.5 | 0.856 | 0.000 | −0.533 | 0.533 |
|  |  | Session #7 | 0.500 | *0.0151 | −0.978 | −1.000 | −0.867 |
|  |  | Session #8 | 0.500 | *0.0151 | −0.978 | −1.000 | −0.867 |
| **Mixed Linear Model Regression Results (Analysis for Assistive Effects)** | | | | | | | |
| Sample Type | Reference | Fixed Effects | Coefficient *β* (SE) | 95% CI Lower | 95% CI Upper | Statistic (z) | *P* value |
| Respiratory | Within Group (Protocol 1) | | | | | | |
|  | - | Intercept | 0.00415 (0.0468) | −0.0875 | 0.0958 | 0.0888 | 0.929 |
|  | Session #1 (BASE) | Session #2 | 4.90e-16 (0.416) | −0.0815 | 0.0958 | 1.18e-15 | 1.000 |
|  |  | Session #3 | 6.24e-16 (0.416) | −0.0815 | 0.0958 | 1.50e-15 | 1.000 |
|  |  | Session #6 | 1.14e-15 (0.416) | −0.0815 | 0.0958 | 2.74e-15 | 1.000 |
|  |  | Session #7 | 2.00 (0.416) | 1.18 | 2.81 | 4.80 | ****1.61e-06 |
|  | Between Groups (Protocol 2 x Session) | | | | | | |
|  | Protocol 1 | Session #1 | −0.00415 (0.0573) | −0.117 | 0.108 | −0.0724 | 0.942 |
|  |  | Session #2 | 1.20 (0.563) | 0.0974 | 2.30 | 2.13 | *0.0329 |
|  |  | Session #3 | 1.96 (0.563) | 0.860 | 3.07 | 3.49 | ***0.000490 |
|  |  | Session #6 | 2.51 (0.569) | 1.39 | 3.63 | 4.41 | ****0.0000105 |
|  |  | Session #7 | 0.471 (0.569) | −0.645 | 1.59 | 0.828 | 0.408 |
|  | Random Effects | | Variance |  |  |  |  |
|  | Between-Participant (Intercept) | | 0.00453 |  |  |  |  |
|  | Within-Participant (Residual) | | 0.0177 |  |  |  |  |
| Note: N=108 (Protocol 1: 10 participants x 5 sessions; Protocol 2: 11 participants x 5 sessions + 1 participant x 3 sessions). Log-likelihood: 57.4 | | | | | | | |
| Stool | Within Group (Protocol 1) | | | | | | |
|  | - | Intercept | 0.404 (0.204) | 0.00447 | 0.803 | 1.98 | *0.0475 |
|  | Session #1 (BASE) | Session #2 | −1.28 (1.78) | −4.78 | 2.22 | −0.718 | 0.473 |
|  |  | Session #3 | 1.08e-14 (1.78) | −3.50 | 3.50 | 6.06e-15 | 1.000 |
|  |  | Session #6 | 0.167 (1.78) | −3.33 | −3.67 | 0.0938 | 0.925 |
|  |  | Session #7 | 10.3 (1.78) | 6.84 | 13.8 | 5.79 | ****6.91e-09 |
|  | Between Groups (Protocol 2 x Session) | | | | | | |
|  | Protocol 1 | Session #1 | 0.129 (0.228) | −0.318 | 0.577 | 0.566 | 0.572 |
|  |  | Session #2 | 8.70 (2.207) | 4.37 | 13.0 | 3.94 | ****0.0000813 |
|  |  | Session #3 | 9.24 (2.207) | 4.91 | 13.6 | 4.19 | ****0.0000285 |
|  |  | Session #6 | 8.88 (2.273) | 4.43 | 13.3 | 3.91 | ****0.0000927 |
|  |  | Session #7 | −0.807 (2.273) | −5.26 | 3.65 | −0.355 | 0.722 |
|  | Random Effects | | Variance |  |  |  |  |
|  | Between-Participant (Intercept) | | 0.113 |  |  |  |  |
|  | Within-Participant (Residual) | | 0.294 |  |  |  |  |
| Note: N=122 (Protocol 1: 9 participants x 5 sessions; Protocol 2: 13 participants x 5 sessions + 4 participants x 3 sessions). Log-likelihood: −109 | | | | | | | |
| **Mixed Linear Model Regression Results (Analysis for Training Effects)** | | | | | | | |
| Sample Type | Reference | Fixed Effects | Coefficient *β* (SE) | 95% CI Lower | 95% CI Upper | Statistic (z) | *P* value |
| Respiratory | Within Group (Protocol 1) | | | | | | |
|  | - | Intercept | 0.00265 (0.0274) | −0.0511 | 0.0564 | 0.0966 | 0.923 |
|  | Session #1 (BASE) | Session #4 | −3.25e-17 (0.117) | −0.229 | 0.229 | −2.79e-16 | 1.000 |
|  |  | Session #8 | 0.659 (0.117) | 0.430 | 0.888 | 5.65 | ****1.60e-08 |
|  | Between Groups (Protocol 2 x Session) | | | | | | |
|  | Protocol 1 | Session #1 | −0.00265 (0.0321) | −0.0655 | 0.0602 | −0.0825 | 0.934 |
|  |  | Session #4 | 0.300 (0.158) | −0.00973 | 0.609 | 1.90 | 0.0577 |
|  |  | Session #8 | −0.331 (0.160) | −0.644 | −0.0181 | −2.07 | *0.0382 |
|  | Random Effects | | Variance |  |  |  |  |
|  | Between-Participant (Intercept) | | 0.00218 |  |  |  |  |
|  | Within-Participant (Residual) | | 0.0055 |  |  |  |  |
| Note: N=65 (Protocol 1: 10 participants x 3 sessions; Protocol 2: 11 participants x 3 sessions + 1 participant x 2 sessions). Log-likelihood: 69.9 | | | | | | | |
| Stool | Within Group (Protocol 1) | | | | | | |
|  | - | Intercept | 0.188 (0.116) | −0.0387 | 0.415 | 1.63 | 0.104 |
|  | Session #1 (BASE) | Session #4 | −0.362 (0.546) | −1.43 | 0.708 | −0.662 | 0.508 |
|  |  | Session #8 | 2.49 (0.546) | 1.43 | 3.56 | 4.57 | ****4.88e-06 |
|  | Between Groups (Protocol 2 x Session) | | | | | | |
|  | Protocol 1 | Session #1 | 0.131 (0.138) | −0.140 | 0.401 | 0.947 | 0.344 |
|  |  | Session #4 | 0.651 (0.675) | −0.672 | 1.97 | 0.965 | 0.335 |
|  |  | Session #8 | −1.84 (0.695) | −3.20 | −0.478 | −2.65 | **0.00810 |
|  | Random Effects | | Variance |  |  |  |  |
|  | Between-Participant (Intercept) | | 0.0131 |  |  |  |  |
|  | Within-Participant (Residual) | | 0.109 |  |  |  |  |
| Note: N=74 (Protocol 1: 9 participants x 3 sessions; Protocol 2: 13 participants x 3 sessions + 4 participants x 2 sessions). Log-likelihood: −26.5 | | | | | | | |

**Table S34. Statistical analysis results for the number of high-performed pooling rounds across pooling sessions for Protocol 2 group.** Matched-pairs rank biserial correlation coefficients (*r*_C_) were calculated for effect sizes using the statistic and ranks from one-sided Wilcoxon signed rank tests. Two mixed linear model regressions with maximum likelihood (ML) estimation were performed to analyze assistive effects and training effects, separately. The number of rounds data were transformed using Yeo-Johnson before the analyses. Dummy coding was used for the regressions. Reference categories were Protocol 2 for the ‘Group’ variable and Session #1 for the ‘Session’ variable. Significance levels: **P* < 0.05; ***P* < 0.01; ****P* < 0.001; *****P* < 0.0001. Adj. *P* value, Benjamini-Hochberg adjusted *P* value (false-discovery rate: 5%); SE, standard error; CI, confidence interval.

| **Protocol 2 Group** | | | | | | | |
| --- | --- | --- | --- | --- | --- | --- | --- |
| **Effect sizes (*r*_C_)** | | | | | | | |
| Sample Type | Reference | Test | Statistic (*T*) | Adj. *P* value | Effect size | 95% CI Lower | 95% CI Upper |
| Respiratory | Session #1 (BASE) | Session #2 | 14.0 | *0.0390 | −0.641 | −0.872 | −0.295 |
| (N=12) |  | Session #3 | 7.50 | *0.0146 | −0.808 | −0.962 | −0.538 |
|  |  | Session #4 | 18.0 | 0.0513 | −0.538 | −0.808 | −0.154 |
| (N=11) |  | Session #5 | 18.0 | 0.0817 | −0.455 | −0.773 | 0.000 |
|  |  | Session #6 | 1.50 | *0.0125 | −0.955 | −1.000 | −0.773 |
|  |  | Session #7 | 3.00 | *0.0125 | −0.909 | −1.000 | −0.682 |
|  |  | Session #8 | 14.0 | 0.0513 | −0.576 | −0.848 | −0.167 |
| Stool | Session #1 (BASE) | Session #2 | 5.00 | **0.00116 | −0.935 | −0.993 | −0.765 |
| (N=17) |  | Session #3 | 1.50 | **0.00116 | −0.980 | −1.000 | −0.902 |
|  |  | Session #4 | 62.5 | 0.250 | −0.183 | −0.634 | 0.307 |
| (N=13) |  | Session #5 | 26.0 | 0.115 | −0.429 | −0.835 | 0.0659 |
|  |  | Session #6 | 1.50 | **0.00231 | −0.967 | −1.000 | −0.835 |
|  |  | Session #7 | 4.50 | **0.00334 | −0.901 | −1.000 | −0.615 |
|  |  | Session #8 | 29.0 | 0.142 | −0.363 | −0.835 | 0.209 |
| **Mixed Linear Model Regression Results (Analysis for Assistive Effects)** | | | | | | | |
| Sample Type | Reference | Fixed Effects | Coefficient *β* (SE) | 95% CI Lower | 95% CI Upper | Statistic (z) | *P* value |
| Respiratory | Within Group (Protocol 2) | | | | | | |
|  | - | Intercept | 5.77e-16 (0.0431) | −0,0844 | 0.0844 | 1.34e-14 | 1.000 |
|  | Session #1 (BASE) | Session #2 | 1.20 (0.380) | 0.457 | 1.95 | 3.16 | **0.00156 |
|  |  | Session #3 | 1.96 (0.380) | 1.22 | 2.71 | 5.17 | ****2.33e-07 |
|  |  | Session #6 | 2.51 (0.389) | 1.75 | 3.27 | 6.45 | ****1.11e-10 |
|  |  | Session #7 | 2.47 (0.389) | 1.70 | 3.23 | 6.34 | ****2.27e-10 |
|  | Between Groups (Protocol 1 x Session) | | | | | | |
|  | Protocol 2 | Session #1 | 0.00415 (0.0573) | −0.108 | 0.117 | 0.0724 | 0.942 |
|  |  | Session #2 | −1.20 (0.563) | −2.30 | −0.0974 | −2.13 | *0.0329 |
|  |  | Session #3 | −1.96 (0.563) | −3.07 | −0.860 | −3.49 | ***0.000490 |
|  |  | Session #6 | −2.51 (0.569) | −3.63 | −1.39 | −4.41 | ****0.0000105 |
|  |  | Session #7 | −0.471 (0.569) | −1.59 | 0.645 | −0.828 | 0.408 |
|  | Random Effects | | Variance |  |  |  |  |
|  | Between-Participant (Intercept) | | 0.00453 |  |  |  |  |
|  | Within-Participant (Residual) | | 0.0177 |  |  |  |  |
| Note: N=108 (Protocol 1: 10 participants x 5 sessions; Protocol 2: 11 participants x 5 sessions + 1 participant x 3 sessions). Log-likelihood: 57.4 | | | | | | | |
| Stool | Within Group (Protocol 2) | | | | | | |
|  | - | Intercept | 0.533 (0.155) | 0.230 | 0.836 | 3.45 | ***0.000570 |
|  | Session #1 (BASE) | Session #2 | 7.42 (1.30) | 4.87 | 9.96 | 5.71 | ****1.12e-08 |
|  |  | Session #3 | 9.24 (1.30) | 6.69 | 11.8 | 7.11 | ****1.14e-12 |
|  |  | Session #6 | 9.05 (1.41) | 6.29 | 11.8 | 6.43 | ****1.27e-10 |
|  |  | Session #7 | 9.53 (1.41) | 6.77 | 12.3 | 6.77 | ****1.28e-11 |
|  | Between Groups (Protocol 1 x Session) | | | | | | |
|  | Protocol 2 | Session #1 | −0.129 (0.228) | −0.577 | 0.318 | −0.566 | 0.572 |
|  |  | Session #2 | −8.70 (2.21) | −13.0 | −4.37 | −3.94 | ****0.0000813 |
|  |  | Session #3 | −9.24 (2.21) | −13.6 | −4.91 | −4.19 | ****0.0000285 |
|  |  | Session #6 | −8.88 (2.27) | −13.3 | −4.43 | −3.91 | ****0.0000927 |
|  |  | Session #7 | 0.807 (2.27) | −3.65 | 5.262 | 0.355 | 0.722 |
|  | Random Effects | | Variance |  |  |  |  |
|  | Between-Participant (Intercept) | | 0.113 |  |  |  |  |
|  | Within-Participant (Residual) | | 0.294 |  |  |  |  |
| Note: N=122 (Protocol 1: 9 participants x 5 sessions; Protocol 2: 13 participants x 5 sessions + 4 participants x 3 sessions). Log-likelihood: −109 | | | | | | | |
| **Mixed Linear Model Regression Results (Analysis for Training Effects)** | | | | | | | |
| Sample Type | Reference | Fixed Effects | Coefficient *β* (SE) | 95% CI Lower | 95% CI Upper | Statistic (z) | *P* value |
| Respiratory | Within Group (Protocol 2) | | | | | | |
|  | - | Intercept | −6.59e-17 (0.0253) | −0.0497 | 0.0497 | −2.60e-15 | 1.00 |
|  | Session #1 (BASE) | Session #4 | 0.300 (0.106) | 0.0911 | 0.508 | 2.82 | **0.00487 |
|  |  | Session #8 | 0.328 (0.109) | 0.114 | 0.542 | 3.00 | **0.00268 |
|  | Between Groups (Protocol 1 x Session) | | | | | | |
|  | Protocol 2 | Session #1 | 0.00265 (0.0321) | −0.0602 | 0.0655 | 0.0825 | 0.934 |
|  |  | Session #4 | −0.300 (0.158) | −0.609 | 0.00973 | −1.90 | 0.0577 |
|  |  | Session #8 | 0.331 (0.160) | 0.0181 | 0.644 | 2.07 | *0.0382 |
|  | Random Effects | | Variance |  |  |  |  |
|  | Between-Participant (Intercept) | | 0.00218 |  |  |  |  |
|  | Within-Participant (Residual) | | 0.0055 |  |  |  |  |
| Note: N=65 (Protocol 1: 10 participants x 3 sessions; Protocol 2: 11 participants x 3 sessions + 1 participant x 2 sessions). Log-likelihood: 69.9 | | | | | | | |
| Stool | Within Group (Protocol 2) | | | | | | |
|  | - | Intercept | 0.319 (0.0847) | 0.153 | 0.485 | 3.76 | ***0.000169 |
|  | Session #1 (BASE) | Session #4 | 0.290 (0.397) | −0.489 | 1.07 | 0.730 | 0.466 |
|  |  | Session #8 | 0.654 (0.430) | −0.189 | 1.50 | 1.52 | 0.128 |
|  | Between Groups (Protocol 1 x Session) | | | | | | |
|  | Protocol 2 | Session #1 | −0.131 (0.138) | −0.401 | 0.140 | −0.947 | 0.344 |
|  |  | Session #4 | −0.651 (0.675) | −1.97 | 0.672 | −0.965 | 0.335 |
|  |  | Session #8 | 1.84 (0.695) | 0.478 | 3.20 | 2.65 | **0.00810 |
|  | Random Effects | | Variance |  |  |  |  |
|  | Between-Participant (Intercept) | | 0.0131 |  |  |  |  |
|  | Within-Participant (Residual) | | 0.109 |  |  |  |  |
| Note: N=74 (Protocol 1: 9 participants x 3 sessions; Protocol 2: 13 participants x 3 sessions + 4 participants x 2 sessions). Log-likelihood: −26.5 | | | | | | | |

**Table S35. Statistical analysis results for the number of Protocol 2 participants achieving high-performance pooling across pooling sessions.** McNemar's exact test was used to compare the proportion of participants who achieved high-performance pooing for ≥2 pooling rounds between the first pooling session (#1, BASE) and subsequent sessions. Adjusted *P* values indicate *P* values adjusted for multiple comparisons using the Benjamini-Hochberg procedure with 5% false-discovery rate. Significance levels: **P* < 0.05; ***P* < 0.01; ****P* < 0.001; *****P* < 0.0001.

| **Protocol 2 (Artificial Respiratory Samples)** | | | |
| --- | --- | --- | --- |
| Comparison | Test statistic | *P* value | Adjusted *P* value |
| #1 (BASE) vs #2 (ASSIST_1_) | 0.00 | 0.500 | 0.750 |
| #1 (BASE) vs #3 (ASSIST_2_) | 0.00 | *0.0156 | *0.0469 |
| #1 (BASE) vs #4 (AFTER_1_) | 0.00 | 1.00 | 1.00 |
| #1 (BASE) vs #6 (ASSIST_3_) | 0.00 | 0.0625 | 0.125 |
| #1 (BASE) vs #7 (ASSIST_4_) | 0.00 | **0.00781 | *0.0469 |
| #1 (BASE) vs #8 (AFTER_2_) | 0.00 | 1.00 | 1.00 |
| **Protocol 2 (Artificial Stool Samples)** | | | |
| Comparison | Test statistic | *P* value | Adjusted *P* value |
| #1 (BASE) vs #2 (ASSIST_1_) | 0.00 | **0.00195 | **0.00293 |
| #1 (BASE) vs #3 (ASSIST_2_) | 0.00 | ***0.000244 | **0.00146 |
| #1 (BASE) vs #4 (AFTER_1_) | 1.00 | 0.375 | 0.375 |
| #1 (BASE) vs #6 (ASSIST_3_) | 0.00 | **0.00195 | **0.00293 |
| #1 (BASE) vs #7 (ASSIST_4_) | 0.00 | **0.00195 | **0.00293 |
| #1 (BASE) vs #8 (AFTER_2_) | 1.00 | 0.219 | 0.2625 |

**Table S36. Statistical analysis results for the rates of *Incorrect* pools between paper-assisted and device-assisted pooling among Protocol 2 participants (N=29; Respiratory: 12, Stool: 17).** One-sided Fisher’s Exact test was used. Null hypothesis is that the proportion of *Incorrect* pools is the same regardless of whether paper instructions or instructional pooling device is used. Paper, written paper instructions; Device, instructional pooling device; *Incorrect*, *Incorrect* pools; *Correct*, *Correct* pools; *Invalid*, *Invalid* pools. Significance levels: **P* < 0.05; ***P* < 0.01; ****P* < 0.001; *****P* < 0.0001.

| Instruction tools | *Incorrect* | *Correct* + *Invalid* | Odds Ratio | *P* value |
| --- | --- | --- | --- | --- |
| Paper | 135 | 289 | Infinity | ****6.64e-47 |
| Device | 0 | 424 |  |  |

**Table S37. Statistical analysis results for the weight of transferred liquid across homogenized stool suspension groups.** Statistical analysis was performed on the weight of transferred liquid across six homogenized stool suspension groups. The Shapiro-Wilk test assessed data normality for each group. Due to non-normal distribution in the Ascaris positive group, the Kruskal-Wallis test was used to evaluate differences between groups. Groups consisted of homogenized stool suspensions from clinical Ascaris-positive samples (Ascaris positive), clinical Ascaris-negative samples (Ascaris negative), and four unique commercial healthy human samples (Negative 1, Negative 2, Negative 3, and Negative 4). Significance levels: **P* < 0.05; ***P* < 0.01; ****P* < 0.001; *****P* < 0.0001.

| **Shapiro-Wilk Normality Test** | | | | | | | |
| --- | --- | --- | --- | --- | --- | --- | --- |
| Group | Sample size (N) | Mean ± SD | Skewness | Kurtosis | W-stat | *P* value | Normality |
| Ascaris positive | 21 | 291 ± 12.4 | 0.742 | 2.18 | 0.872 | *0.0103 | No |
| Ascaris negative | 21 | 293 ± 17.8 | 0.0162 | 2.08 | 0.974 | 0.809 | Yes |
| Negative 1 | 42 | 290 ± 17.6 | 0.0702 | 2.22 | 0.983 | 0.786 | Yes |
| Negative 2 | 42 | 292 ± 14.2 | 0.230 | 1.96 | 0.951 | 0.0706 | Yes |
| Negative 3 | 42 | 296 ± 16.5 | 0.0906 | 2.00 | 0.958 | 0.123 | Yes |
| Negative 4 | 42 | 288 ± 13.3 | 0.222 | 2.10 | 0.963 | 0.183 | Yes |
| **Kruskal-Wallis Test** | | | | | | | |
| H-statistic | 5.93 |  |  |  |  |  |  |
| *P* value | 0.314 |  |  |  |  |  |  |

**Table S38. Bill of materials for the instructional sample-pooling device.** Some components (such as jumper wires, solder wires, adhesives, screws, etc.), are not included. Bare spool weight of 3D printing filament was estimated 245 g. Weight of used 3D printing filament (without support) was calculated by PrusaSlicer 2.9.2 software. Costs are presented in US dollars. Since some products are discontinued (as of 2025), alternative products are indicated in the table. ASIN, Amazon Standard Identification Number.

| Model Number | Description | Quantity | Unit | Unit Cost | Source | ASIN |
| --- | --- | --- | --- | --- | --- | --- |
| Display Module | | | | | | |
| A000062 | Arduino Due | 1 | EA | $ 35.75 | Amazon US | B00A6C3JN2 |
| ABX00028 | Arduino Nano Every | 1 | EA | $ 9.67 | Amazon US | B07YQ4Q3Y5 |
| KMRTM32032-SPI | 3.2” TFT LCD Display, ILI9341, SPI | 1 | EA | $ 15.99 | Amazon US | B0B1M9S9V6 |
| ANMBEST_MD272 | MicroSD Card Reader | 1 | EA | $ 0.899 | Amazon US | B08CMLG4D6 |
| SDSQUA4-032G-GN6MT | SanDisk MicroSD Card | 1 | EA | $ 6.46 | Amazon US | B08J4HJ98L |
| DS3231 AT24C32 | DS3231 RTC Module | 1 | EA | $ 2.598 | Amazon US | B00LX3V7F0 |
| CYT1036 | Active Buzzer | 1 | EA | $ 0.698 | Amazon US | B01N7NHSY6 |
| ML-HM-10 | Bluetooth 4.0 BLE Module | 1 | EA | $ 10.99 | Amazon US | B06WGZB2N4 |
| DC5A75W | HW-64 DC-DC Step-Down Converter | 1 | EA | $ 8.98 | Amazon US | B06XT6KMPX |
| ALT-1210 | 12V 10A Power Supply | 1 | EA | $ 20.99 | Amazon US | B07MXXXBV8 |
| AZ18051707 | DC Power Jack | 1 | EA | $ 0.2163 | Amazon US | B07D4BK8W4 |
| IN4007 | 1N4007 Rectifier Diode | 4 | EA | $ 0.04792 | Amazon US | B07Q6J9TNW |
| LM317T | LM317 Voltage Regulator IC | 2 | EA | $ 0.2663 | Amazon US | B0CBLN2X7H |
| 705883356441 | Logic Level Converter 3.3V-5V | 10 | EA | $ 0.769 | Amazon US | B07LG646VS |
| BJ-RS-25VALUES-1000PCS | 1/4W Carbon Film Resistor | 4 | EA | $ 0.00909 | Amazon US | B08FD1XVL6 |
| 603935399364 | JST-XHP 2.54 mm Connector | 34 | EA | $ 0.01423 | Amazon US | B07CTH46S7 |
| 8541770554 | 2.54 mm Male-Female Pin Connector | 7 | EA | $ 0.115625 | Amazon US | B09X32766Z |
| a18061500ux0235 | 2x8 cm Double Sided Solderable Board | 1 | EA | $ 0.849 | Amazon US | B07FK3NLG2 |
| a18061500ux0227 | 4x6 cm Double Sided Solderable Board | 3 | EA | $ 0.999 | Amazon US | B07FK48ZMN |
| a18061500ux0210 | 6x8 cm Double Sided Solderable Board | 1 | EA | $ 1.367 | Amazon US | B07FK46JPR |
| a18061500ux0234 | 7x9 cm Double Sided Solderable Board | 2 | EA | $ 1.499 | Amazon US | B07FK49T55 |
| KCD1-8-101 | 12V On/Off Switch | 1 | EA | $ 4.995 | Amazon US | B0CF4L549N |
| 191314011674 | HATCHBOX 1.75 mm Silk Gold PLA | 335.94 | g | $ 0.034423 | Amazon US | B07HXRNM1M |
|  | 100 nF Ceramic Capacitor | 2 | EA | $ 0.01615 | Amazon US | B07PRC5JJY |
|  | 1 μF Ceramic Capacitor | 2 | EA | $ 0.01615 | Amazon US | B07PRC5JJY |
|  | 10 μF Ceramic Capacitor | 4 | EA | $ 0.01615 | Amazon US | B07PRC5JJY |
| Sub-Total | | | | $ 147.88 |  |  |
| Tube Module | | | | | | |
| HW-201 | IR Infrared Sensor Module | 6 | EA | $ 0.4995 | Amazon US | B08DR1W3BK |
| WS2812-02 | WS2812B 5050SMD RGB LED Socket | 5 | EA | $ 0.0666 | Amazon US | B097379J1V |
| 603935399364 | JST-XHP 2.54 mm Connector | 24 | EA | $ 0.01423 | Amazon US | B07CTH46S7 |
| a18061500ux0235 | 2x8 cm Double Sided Solderable Board | 2 | EA | $ 0.849 | Amazon US | B07FK3NLG2 |
| a18061500ux0231 | 3x7 cm Double Sided Solderable Board | 1 | EA | $ 0.859 | Amazon US | B07FK5N39S |
| 3D PLA-1KG1.75-SHNY-LBLU | HATCHBOX 1.75 mm Metallic Finish Light Blue PLA | 199.58 | g | $ 0.035748 | Amazon US | B09T8RPQYK |
| Sub-Total | | | | $ 13.36 |  |  |
| Cap Module | | | | | | |
| LM393P | LM393 Voltage Dual Differential Comparator | 3 | EA | $ 0.1198 | Amazon US | B0DCBPJLZT |
| BJ-RS-25VALUES-1000PCS | 1/4W Carbon Film Resistor | 20 | EA | $ 0.00909 | Amazon US | B08FD1XVL6 |
| HW-201 | IR Infrared Sensor Module | 1 | EA | $ 0.4995 | Amazon US | B08DR1W3BK |
| WS2812-02 | WS2812B 5050SMD RGB LED Socket | 5 | EA | $ 0.0666 | Amazon US | B097379J1V |
| 603935399364 | JST-XHP 2.54 mm Connector | 18 | EA | $ 0.01423 | Amazon US | B07CTH46S7 |
| a18061500ux0235 | 2x8 cm Double Sided Solderable Board | 4 | EA | $ 0.849 | Amazon US | B07FK3NLG2 |
| a18061500ux0231 | 3x7 cm Double Sided Solderable Board | 1 | EA | $ 0.859 | Amazon US | B07FK5N39S |
| 849344040439 | HATCHBOX 1.75 mm Matte Kraft PLA  (Alternative option: HATCHBOX 1.75 mm Wood PLA) | 128.54 | g | $ 0.034423 | Amazon US | B01092XXD4 |
|  | 940 nm Infrared LED Pair (Emitter and Receiver) | 5 | EA | $ 0.2198 | Amazon US | B01MFCFLA7 |
|  | 10 μF Ceramic Capacitor | 6 | EA | $ 0.01615 | Amazon US | B07PRC5JJY |
| Sub-Total | | | | $ 11.51 |  |  |
| Pipette Module | | | | | | |
| HW-201 | IR Infrared Sensor Module | 6 | EA | $ 0.4995 | Amazon US | B08DR1W3BK |
| WS2812-02 | WS2812B 5050SMD RGB LED Socket | 5 | EA | $ 0.0666 | Amazon US | B097379J1V |
| 603935399364 | JST-XHP 2.54 mm Connector | 24 | EA | $ 0.01423 | Amazon US | B07CTH46S7 |
| a18061500ux0235 | 2x8 cm Double Sided Solderable Board | 2 | EA | $ 0.849 | Amazon US | B07FK3NLG2 |
| a18061500ux0231 | 3x7 cm Double Sided Solderable Board | 1 | EA | $ 0.859 | Amazon US | B07FK5N39S |
| 3D PLA-1KG1.75-SHNY-PNK | HATCHBOX 1.75 mm Metallic Finish Pink PLA | 197.28 | g | $ 0.035748 | Amazon US | B09T8YMZZD |
| Sub-Total | | | | $ 13.28 |  |  |
| Scale Module | | | | | | |
| HW-201 | IR Infrared Sensor Module | 1 | EA | $ 0.4995 | Amazon US | B08DR1W3BK |
| WS2812-02 | WS2812B 5050SMD RGB LED Socket | 2 | EA | $ 0.0666 | Amazon US | B097379J1V |
| Stemedu-ST0728X5 | HX711 Amplifier Module | 1 | EA | $ 1.898 | Amazon US | B07MTYT95R |
| 603935399364 | JST-XHP 2.54 mm Connector | 10 | EA | $ 0.01423 | Amazon US | B07CTH46S7 |
| a18061500ux0227 | 4x6 cm Double Sided Solderable Board | 3 | EA | $ 0.999 | Amazon US | B07FK48ZMN |
| 3D PLA-1KG1.75-LMGRN | HATCHBOX 1.75 mm Pastel Green PLA  (Alternative option: HATCHBOX 1.75 mm Lime Green PLA) | 153 | g | $ 0.033099 | Amazon US | B09WW5HJCV |
|  | 50g Load Cell | 1 | EA | $ 17.99 | Amazon US | B07TBJBFRK |
| Sub-Total | | | | $ 28.72 |  |  |
| Barcode Module | | | | | | |
| WS2812-02 | WS2812B 5050SMD RGB LED Socket | 5 | EA | $ 0.0666 | Amazon US | B097379J1V |
| 681413425088 | Waveshare Barcode Scanner Module | 1 | EA | $ 42.23 | Amazon US | B07P3GD3XV |
| 8541770554 | 2.54 mm Male-Female Pin Connector | 1 | EA | $ 0.115625 | Amazon US | B09X32766Z |
| 603935399364 | JST-XHP 2.54 mm Connector | 6 | EA | $ 0.01423 | Amazon US | B07CTH46S7 |
| 191314011711 | HATCHBOX 1.75 mm Metallic Finish Mint PLA | 105.06 | g | $ 0.034423 | Amazon US | B07HXR222M |
| 686091797090 | MBSS Mixed Solderable Breadboard Proto Board | 1 | EA | $ 0.81145 | Amazon US | B082PX1GKT |
| Sub-Total | | | | $ 47.19 |  |  |
| Multibarcode Module | | | | | | |
| 681413425088 | Waveshare Barcode Scanner Module | 6 | EA | $ 42.23 | Amazon US | B07P3GD3XV |
| COM-102 | Atlas Scientific 8:1 Serial Port Expander | 1 | EA | $ 19.99 | Amazon US | B07KJNZWST |
| 8541770554 | 2.54 mm Male-Female Pin Connector | 4 | EA | $ 0.115625 | Amazon US | B09X32766Z |
| 603935399364 | JST-XHP 2.54 mm Connector | 2 | EA | $ 0.01423 | Amazon US | B07CTH46S7 |
| 3D PLA-1KG1.75-SHNY-WHT | HATCHBOX 1.75 mm Metallic Finish White PLA | 163.20 | g | $ 0.035748 | Amazon US | B09T9HCJ4P |
| a18061500ux0234 | 7x9 cm Double Sided Solderable Board | 1 | EA | $ 1.499 | Amazon US | B07FK49T55 |
| Sub-Total | | | | $ 281.19 |  |  |
| Waste Module | | | | | | |
| WS2812-02 | WS2812B 5050SMD RGB LED Socket | 9 | EA | $ 0.0666 | Amazon US | B097379J1V |
| DH-MS | 500g Load Cell | 1 | EA | $ 6.99 | Amazon US | B07YRCQ6L3 |
| LM358P | LM358P Operational Amplifier IC | 1 | EA | $ 0.1398 | Amazon US | B07WQWPLSP |
| IN4007 | 1N4007 Rectifier Diode | 4 | EA | $ 0.04792 | Amazon US | B07Q6J9TNW |
| 8541770554 | 2.54 mm Male-Female Pin Connector | 1 | EA | $ 0.115625 | Amazon US | B09X32766Z |
| 603935399364 | JST-XHP 2.54 mm Connector | 24 | EA | $ 0.01423 | Amazon US | B07CTH46S7 |
| DKMTm | 54x33 mm Double Sided Solderable Board | 5 | EA | $ 1.665 | Amazon US | B09ZPHDN3V |
| Stemedu-ST0728X5 | HX711 Amplifier Module | 1 | EA | $ 1.898 | Amazon US | B07MTYT95R |
| 3D PLA-1KG1.75-SHNY-RED | HATCHBOX 1.75 mm Metallic Finish Red PLA | 566.89 | g | $ 0.035748 | Amazon US | B09T8PB3WK |
| 191314011667 | HATCHBOX 1.75 mm Silk Brown PLA | 223.10 | g | $ 0.034423 | Amazon US | B07HXDPCMQ |
|  | SR602 PIR Sensor | 5 | EA | $ 0.994 | Amazon US | B0D319TSY3 |
| Sub-Total | | | | $ 51.52 |  |  |
| **Total Device Cost** | | | | $ 594.66 |  |  |

**Method S1. Materials for device fabrication**

We used the following materials to fabricate our instructional pooling device and disposable trays for pooling. 1.75-mm polylactic acid (PLA) 3D printing filaments with different colors (i.e., metallic shiny filaments with white, red, pink, and mint colors; basic filament with pastel green color; silky filaments with gold and light blue colors; matte filaments with the kraft color) were purchased from HATCHBOX 3D (Rowland Heights, CA, USA). 1.75-mm polyamide 6 (PA6) with carbon fiber (CF) 3D printing filaments were purchased from Polymaker (Shanghai, China). Arduino Due and Nano Every were purchased from Arduino LLC (Boston, MA). Barcode scanner modules (a CMOS image sensor with a 640x480 pixel resolution; field-of-view: 21.5°–28°; a reading distance of 60–110 mm) supporting Code-128 and QR formats were purchased from Waveshare Electronics (Shenzen, China). 8:1 Serial Port Expander was purchased from Atlas Scientific (Long Island City, NY). DS3231 real-time clock (RTC) was purchased from Shenzhen HitLetgo Technology Co., Ltd. (Shenzhen, China). RGB LEDs (5050 SMD) with WS2812B integrated circuits were purchased from BTF-LIGHTING Technology Co., Ltd. (Shenzhen, China). HM-10 Bluetooth 4.0 BLE module was purchased from DSD TECH^®^ (Shenzhen, China). Micro SDHC UHS-I 32GB cards (model #: SDSQUA4-032G-GN6MT) were purchased from SanDisk Corporation (Milpitas, CA, USA). The primer coater was purchased from Polar Coatings (Winsford, UK). Acrylic paint was purchased from Magicfly (Charlotte, NC). Silicon caulks were purchased from The Gorilla Glue Company (Cincinnati, OH) and Henkel (Düsseldorf, Germany). Epoxy resin was purchased from East Coast Resin (Staten Island, NY, USA). Individually wrapped 300-uL dual-bulb transfer pipettes were purchased from Shenzhen Scilab-bio Technology Industrial Co., Ltd (Shenzhen, China). 12-mm sample-collection tubes (tubes – Cat# 22040238; caps – Cat# 22040244; cap inserts – Cat# 22045361 & Cat# 22045422) were purchased from Simport Scientific Inc. (QC, Canada). 1/8″ acrylic plates (Cat# 8536K131) were purchased from McMaster-Carr (Chicago, IL, USA). 500-µm polypropylene film (Cat# 46170) was purchased from Spartech (Maryland Heights, MO, USA). Other inexpensive components from unspecified manufacturers were purchased through Amazon.com, including HW-201 infrared (IR) object detection sensors (940 nm IR; sensing distance: 20–300 mm), HX711 modules, LM317 voltage regulators, LM393 dual differential comparators, SR602 passive-IR (PIR) sensors, 3.2-inch SPI TFT LCD with ILI9341 IC driver, 940nm IR LED pairs of emitters and receivers, 50-g and 500-g strain gauge load cells, micro SDHC card reader module (3.3V logic), HW-64 DC-DC buck converter with XL4015 chip, 1N4007 rectifier diodes, an active piezoelectric buzzer, 3.3V-5V bi-directional logic level shifters, 2.1mm DC power barrel jack, resistors, capacitors, electronic wires, double-sided solderable circuit boards, Dupont connectors, JST-SM 2.54-mm connectors, JST-XH 2.54-mm connectors, waterproof toggle switches for 12V, 12V power adapter, and nylon cable gland with IP68 waterproof.

**Method S2. Fabrication of instructional pooling device and trays**

The instructional pooling device consisted of eight colored modules **(Supplementary Fig. S3)** manufactured using an Original Prusa i3 MK3S+ 3D printer (Prusa Research a.s.; Prague 7, Czech Republic). For the user study version, we used PLA filaments in different colors for simple fabrication. For the device validation study using clinical STH samples, we chose black Nylon/PA filaments to ensure chemical resistance, as the device needed decontamination with solutions like 20% bleach, 70% ethanol, and 10% povidone-iodine after handling biohazardous solutions potentially containing *Ascaris lumbricoides* eggs in a biosafety cabinet. To maintain consistent color coding on the device for device validation study, we applied white primer to the black Nylon/PA modules, followed by colored acrylic paints. We then coated the surface with epoxy resin to seal any gaps where Ascaris eggs might lodge and to create smooth, water- and chemical-resistant surfaces. We used 1/8” acrylic plates and 500-µm polypropylene sheets for the object detection/scanning windows. These plates and sheets were cut using an Epilog Engrave Zing 16, 30-Watt, laser cutting machine (Epilog Laser; Golden, CO, USA) with the following settings: 40% power, 50% speed, 2500 Hz frequency, and 500 DPI in vector mode. Then, the plates and sheets were secured to the modules with acrylic adhesive. These transparent windows allowed IR or PIR sensors to detect objects and barcode scanners to read tube barcodes while protecting against liquids and chemicals. Finally, we sealed all gaps and holes between modules, plastic parts, and components using silicone caulks and epoxy to ensure complete protection from biohazardous solutions, chemicals, and water.

To produce translucent device-protective trays, we used a vacuum former (Formech 300XQ, Formech; Middleton, WI, USA) to shape 500-µm polypropylene sheets over 3D-printed molds. The molds were created using a Polyjet Connex 260 printer (Stratasys Ltd.; Eden Prairie, MN, USA). The vacuum forming process was conducted at ~180°C for 50 seconds.

The eight device modules **(Fig. 2*A* and Supplementary Fig. S3)** were designed to allow human-device interactions through graphic display, object detection, barcode scanning, and weight measurement. The Display Module was equipped with a 3.2-inch TFT LCD screen covered by a transparent acrylic window to display language-agnostic graphics. To control the other device-modules and peripheral devices, the Display Module inside was also equipped with key components of the system, including two microcontroller boards, power switch, voltage regulators, 3.3V-5V level shifters, and peripheral modules of microSD card, Bluetooth communication, RTC, and piezoelectric buzzer **(Supplementary Fig. S2)**.

Three device modules (i.e., Tube, Cap, and Pipette) were designed to monitor laboratory materials (e.g., sample-collection tubes, tube caps, wrapped pipettes, etc.) loaded in thermoformed translucent trays designed for protecting the device interior from potential liquid contamination **(Supplementary Figs. S2–3)**. The Tube and Pipette Modules shared identical IR sensor configurations, each utilizing six pairs of 940 nm IR emitters and receivers protected by nested 3D printed casings with transparent acrylic windows. For detecting individual tubes or pipettes, we installed five IR emitter-receiver pairs along the left side of each module. Each pair was oriented at ~45° angles relative to each other, enabling detection of tubes or pipettes placed in the tray at approximately 25 mm distance. Each receiver incorporated a custom slotted black mask to ensure precise detection of target objects while filtering peripheral reflections. For tray detection, we configured the sixth IR emitter-receiver pair in parallel orientation to detect the tray's lateral surface. The Cap Module had a different sensor arrangement from the Tube and Pipette Modules. Each cap slot featured transparent acrylic windows on opposite sides, with IR emitter-receiver pairs mounted in direct opposition – the emitter on one side sending its beam across the slot to its corresponding receiver on the other side. This arrangement enables detection when a cap in the translucent tray interrupts the IR beam passing through the slot. As with the Tube and Pipette Modules, we installed the sixth sensor pair in parallel orientation to detect the presence of the cap tray.

The Scale Module was designed to monitor the pooling tube loaded in the device throughout the pooling process **(Supplementary Figs. S2–3)**. The Scale Module incorporated a 50-g strain gauge load cell (resolution: 1 mg) with a 24-bit ADC module to serve two functions: (1) detecting the presence of a pooling tube when loaded on the weighing scale and (2) measuring real-time weight changes during sample dispensing. For cap detection after tube opening, the module featured a dual IR sensor system: one IR emitter paired with two IR receivers, all protected by transparent acrylic windows. The first IR receiver was positioned alongside the emitter on one side of the cap slot, while the second receiver was mounted on the opposite side. The translucent cap tray had a black mask applied to one side of the tray facing the single IR receiver to block the IR transmission. This configuration enabled three-state detection: when the slot is empty, IR light passes through transparent acrylic windows from the emitter to the opposite receiver; when the tray is inserted, the black mask blocks the IR transmission to the opposite receiver; and when a cap is present in the tray, the IR light reflects off the cap surface to the adjacent receiver, confirming cap placement.

The Waste Module was designed to detect the disposal of waste generated during the pooling process **(Supplementary Figs. S1 and S3)**. The Waste Module was equipped with five passive-IR sensors to detect user’s hand and 500-g strain gauge load cell (resolution: 10 mg) to detect the weight changes in the secondary waste bin inside the Waste Module. For easier device maintenance in the biosafety cabinet, the load cell was not installed in the device for device validation study with clinical STH samples.

The other two device modules incorporated barcode scanners to scan and record barcodes of objects **(Supplementary Figs. S2–3)**. The Barcode Module was equipped with one barcode scanner for scanning QR codes on user cards and barcodes of pooling tubes. The Multi-barcode Module incorporated five barcode scanners to scan barcodes of individual sample-collection tubes. Each scanner was positioned to align with a corresponding tube slot in the protective tube tray. To allow each scanner to read the tube barcode at the side, we installed a transparent acrylic window along the right side of the Tube Module that enabled scanning through both the window and the translucent protective tray.

**Method S3. System architecture of instructional pooling device**

The instructional device system **(Supplementary Figs. S1–2)** was implemented using a dual-microcontroller architecture operating at 9 – 12VDC. The main unit utilizes the 32-bit ARM® Cortex®-M3 processor, 84 MHz, (AT91SAM3X8E; Microchip Technology Inc.), interfaced with an 8-bit AVR® processor, 16 MHz, (ATmega4809; Microchip Technology Inc.) via UART communication (baud rate: 115,200 bps). The power management system was designed with independent voltage regulation circuits connected in parallel to the input power (≥9V, ≥3A). The primary power rail uses a DC-DC buck converter (XL4015E1, 180 kHz switching frequency) to generate 5V with up to 5A output capability, supplying power to high-current peripherals operating at 5V (e.g., object detection sensors, LEDs, and barcode scanners). Two secondary rails using linear voltage regulators (LM317) provide: (1) 8V for the microcontroller boards, and (2) 3.3V for both low-power peripherals and logic-level shifters between 5V logic I/O of sensors and 3.3V logic I/O of the primary 32-bit microcontroller.

The system architecture **(Fig. 2*B* and Supplementary Movie S2)** was designed to perform five primary functions: (1) detection of laboratory materials and operator’s hands for user guidance; (2) real-time monitoring for user error detection and feedback for error correction; (3) sample tracking through 1D/2D barcode readers; (4) precise weight measurement of pooled liquid samples using a load cell (range: 1 mg – 50 g, resolution: 1 mg); and (5) wireless data transmission to a mobile device via Bluetooth 4.0 BLE technology.

For the user guidance, the primary MCU (AT91SAM3X8E) was programmed to manage peripheral device interactions (e.g., display, LEDs, IR sensors, PIR sensors, load cells etc.) and implement polling loops **(Fig. 2*F*)** in each handling step. At the beginning of each pooling loop, the MCU retrieves the customized language-agnostic RGB565 images (320x240 pixels) **(Fig. 2*E*)** from the 32GB SD card via serial peripheral interface (SPI) communication and rendered them on the LCD screen with the ILI9341 driver. To achieve faster responsive visual feedback, the MCU utilizes its internal direct memory access (DMA) controller (channels 0 and 1) for efficient 16-bit SPI data transfer between memory and the display, utilizing polling-based synchronization. While signaling with a pink/purple LED (RGB values: [9,0,18]) corresponding to a certain step, the system monitors object presence/absence to capture handling actions during the loop: individual sample-collection tubes and their tray (Tube Module), tube caps and their tray (Cap Module), individually wrapped dual-bulb pipettes and their tray (Pipette Module), a pooling tube and the tray for its cap (Scale Module), or the user’s hands or disposed waste (Waste Module). After successful proper handling action completion, we configured the primary MCU to signal progression to the next handling step by changing the LED color to green (RGB values: [2,20,2]).

For real-time error detection during the guidance, the primary MCU implements interrupt-driven control loops to monitor all other modules through edge-triggered interrupts from IR proximity sensors. Upon detecting a handling error, the system performs a two-step validation process: interrupt-based signal change detection (rising edge ≥2.7V; falling edge ≤1V), followed by confirmatory validation through repeated sensor readings (10 consecutive readings at 5 ms intervals). When multiple interrupts are triggered, the primary MCU validates the interrupts with a hierarchical priority structure: the validation for device-protective trays take precedence, followed by object presence/absence validation in descending priority (tubes > caps > pipettes > pooling tube > pooling tube cap). Upon confirming a user error, the system provides both visual feedback (a warning sign on the display) and auditory alerts (piezoelectric buzzer, 1,050 Hz for 125 ms duration, 1-second intervals), with error correction guidance through error-specific instructional graphic.

For sample tracking, the system creates a data recording file when the user scans a user card with a QR code with a certain barcode identifier: "USER" for unique individuals (i.e., participants in the user study) or "EP" for unique samples (i.e., clinical STH samples in the clinical sample pooling validation study). The data file is used for storing tube barcodes scanned as well as weight data during the process. All barcode scanners initiate scanning (with infinite scanning time) upon receiving a specific command (0x7E 00 08 01 00 02 01 AB CD) from the primary MCU via UART communication at 9,600 bps. When the tube tray with individual tubes is inserted in the device, the system scans the barcode of the tube tray and identifies two key parameters: the pool size (2, 3, 4, or 5 samples) and the desired volume per sample (100 µL, 150 µL, 200 µL, 250 µL, 300 µL, or 400 µL). After scanning, the system scans the individual tubes sequentially. When scanning, the system uses unique tube barcode indicators to differentiate objects: "SP" for individual sample tubes and "PL" for pooling tubes. To maintain data integrity, the primary MCU validates each scanned barcode and only records those matching the correct object type.

For reliable and precise weight measurement of an object, the system reads three consecutive 24-bit digital signals from the HX711 analog-to-digital converter (ADC) and converted them into weight values at 100 ms (or 200 ms) intervals. These weight values are then processed to calculate their mean and coefficient of variation (CV), which helps minimize signal noise and determine weight stabilization. CV below 1% indicates minimal weight fluctuation, at which point the average weight is recorded as the actual measurement.

The weight measurement for each pooled sample begins after the system tares the pooling tube. During the pooling operation, the system monitors user interaction or sample dispensing by detecting when the absolute average weight (converted to mass units, mg) exceeds 2 mg with a CV ≥ 5%. After dispensing, the system confirms a successful sample transfer when three conditions are met: the CV drops below 1%, the standard deviation is less than 5 mg, and the average weight exceeds 2 mg. Once these criteria are satisfied, the system records the stabilized weight of the pooled sample on the data file, while displaying the relative accuracy of the transferred volume on the screen.

For wireless data transmission, the primary MCU interfaces with a Bluetooth module via two-wire serial communication (I2C). The data transfer process is initiated when the user scans a card containing the "BLE" QR code, followed by scanning their user card. Upon detecting the user card, the MCU locates the associated data file, retrieves its contents, and transmits the data to the user's device through the Bluetooth connection.

**Method S4. Expected handling movement on the instructional pooling device**

Based on the device pooling instructions **(Supplementary Note S1)**, we analyzed the expected handling movements across device modules **(Fig. 1*G*)** and quantified their frequencies **(Fig. 1*H*)**. For each instructional step, we calculated movement frequency values by considering the number of possible pathways between device modules. When only one pathway existed in a step, we assigned a full count (value of 1); for steps with two possible pathways, we assigned a partial count (value of 0.5) to each possible pathway. The cumulative frequency values for all module-to-module transitions were calculated by summing these assigned values across all instructional steps, providing a quantitative measure of expected handling movements throughout the complete pooling procedure.

**Method S5. Characterization of strain gauge-based weight measurement system of instructional pooling device**

A certified F1-grade calibration weight kit (Goetland; Shanghai, China) was used to characterize the Scale Module's weighing accuracy. For low-mass calibration weights (1 mg and 2 mg), we performed 20 measurement trials to determine the limit of blank (LoB) and limit of detection (LOD). For higher-mass weights (5 mg – 50 g), 10 trials were conducted. In each trial, after taring the scale, weight measurements were recorded at 100-ms intervals. Data collection began prior to weight placement to establish baseline (blank) measurements and continued for 15 seconds after loading to ensure weight stabilization. From these time-series measurements, we analyzed both the blank signal (pre-loading baseline) and the loaded weight values, calculating their respective means for each trial. Using the compiled mean values, we calculated LoB and LoD as follows^3^:

LoB = μ_avg,blank_ + 1.645 × σ_std,blank_

where μ_avg,blank_ is the average of all 170 blank mean values (combined across all trials and calibration weights) and σ_std,blank_ is their standard deviation.

LoD = LoB + 1.645 × σ_std,1mg_

where σ_std,1mg_ is the standard deviation of the 20 mean values obtained from 1-mg weight measurements. For each calibration weight, measurement accuracy and inter-trial variability were calculated as:

Accuracy (%) = (1 - |known weight - μ_avg,weight_| / known weight) × 100

Inter-trial variability (%CV_inter_) = (σ_std,weight_ / μ_avg,weight_) × 100

where μ_avg,weight_ is the average of mean values across trials for each calibration weight, and σ_std,weight_ is their standard deviation. For each trial, the signal-to-noise ratio (SNR) was calculated in decibels using:

SNR = 10log10(μ_mean,weight_^2^ / σ_std,noise_^2^)

where μ_mean,weight_ is the mean of the stabilized weight measurements after loading the calibration weight, and σ_std,noise_ is the standard deviation of blank measurements before weight loading in each trial. The final SNR for each calibration weight was determined by averaging the SNR values across all corresponding trials (20 trials for 1 – 2 mg weights; 10 trials for 5 mg – 50 g weights).

**Method S6. Design of user study**

The user study was designed for evaluating two primary hypotheses: (1) whether an instructional pooling device could assist laboratory-inexperienced personnel in improving their performance when handling challenging sample types, and (2) whether the device could effectively train these personnel to maintain high performance levels in sample pooling without the assistance from the device. The study implemented two distinct training protocols (Protocol 1 and 2), each comprising 8 pooling sessions distributed across two days (4 sessions per day). Each session consisted of 4 pooling rounds, totaling 16 rounds per day. Sessions were conducted either with paper-based instructions or with the instructional pooling device, depending on the protocol.

Protocol 1 **(Fig. 3*A*)** was structured to examine two key aspects of the training process: the effectiveness of conventional paper-based instructions in training participants, and the subsequent impact of introducing an instructional pooling device. Day 1 protocol was designed with four paper-assisted pooling sessions (16 rounds) to investigate whether the use of paper instructions alone could improve pooling performance. The Day 2 protocol was structured in two phases. The first phase consisting of two paper-assisted sessions (8 rounds) was designed to reacquaint participants with the pooling procedure and establish a baseline performance level for comparison. The second phase with two device-assisted sessions was designed to evaluate the potential performance impact of the instructional pooling device relative to the baseline performance with the paper instructions.

To systematically evaluate Protocol 1, we tested three null hypotheses: (1) repeated practice with paper instructions does not significantly improve sample-handling competency across the 16 rounds on Day 1; (2) the continued practice on Day 2's first 8 rounds shows no significant performance improvement compared to Day 1, despite potential carryover effects; and (3) the introduction of the instructional pooling device in the final 8 rounds of Day 2 does not significantly enhance sample-handling competency compared to performance with paper instructions.

Protocol 2 **(Fig. 3*B*)** was designed to investigate the device's effectiveness in skill acquisition during device use and performance maintenance after device removal. The protocol structure was identical for both Day 1 and Day 2: four consecutive sessions consisting of one paper-assisted session (session #1 or #5), followed by two device-assisted sessions (sessions #2-3 or #6-7), and concluding with one paper-assisted session (session #4 or #8). The initial paper-assisted session was designed to establish a baseline performance level for Day 1, and to reacquaint participants with the pooling procedure on Day 2. The two device-assisted sessions were designed to evaluate the immediate assistive effects of the instructional pooling device compared to the baseline performance. The final paper-assisted session was designed to evaluate whether participants could maintain performance quality after device removal.

To systematically evaluate Protocol 2, we tested three null hypotheses: (1) the instructional pooling device does not significantly improve sample-handling competency (assistive effects; skill acquisition) across 8 device-assisted rounds compared to the initial 4 paper-assisted rounds (session #1); (2) there are no significant training effects (skill retention/application) on sample-handling competency when comparing the initial paper-assisted rounds (session #1) with the final paper-assisted rounds (session #4 or #8); and (3) there is no significant difference in sample-handling competency between Day 1's final paper-assisted rounds (session #4) and Day 2's initial paper-assisted rounds (session #5).

**Method S7. Participants for the user study**

We enrolled a total of 48 participants recruited from the Pasadena, CA, USA area. As part of the recruitment process, we prescreened participants and excluded those who had research-level laboratory experience to ensure the population group was representative of personnel inexperienced in wet laboratories. We included participants with some prior laboratory experience through high school, community college, or university freshmen courses in biology or chemistry, according to self-reported demographic surveys conducted after individual participants completed the pooling exercise on Day 1 (see “Post-experimental procedures” below). Of total 48 participants, 37.5% were Caltech-affiliated undergraduate students/employees, while 62.5% were non-Caltech members. 37 participants (77.08%) had no laboratory experience or only limited experience through non-AP high school biology or chemistry courses; we classified these individuals as “laboratory-inexperienced personnel” for this study. There were equal number of males and females, with 62.5% of participants aged 18-25.

Participants were assigned to one of four groups at the time of recruitment: Protocol 1 – respiratory, Protocol 1 – stool, Protocol 2 – respiratory, and Protocol 2 – stool as they were enrolled. While initial assignment occurred in the order of enrollment, we subsequently balanced sex distribution across groups within the same protocols to minimize potential sex-related confounding effects. However, five participants in Protocol 2 (four from stool; one from respiratory) withdrew before the second experiment day (Day 2), necessitating additional recruitment for these groups. The final distribution of participants across the four groups was: Protocol 1 – respiratory (10 on both days), Protocol 1 – stool (9 on both days), Protocol 2 – respiratory (12 on Day 1, 11 on Day2), and Protocol 2 – stool (17 on Day 1, 13 on Day 2). Raw demographic information and its summary are described in **Supplementary Fig. S6 and Tables S1–5**.

The study was conducted one participant at a time over two non-consecutive days (a median of 7 days apart, IQR: 4–8 days, range: 1–24 days, *n*=43; **Supplementary Fig. S14a**). Compensation (total up to $90) was given to individual participants as remuneration for time of participation as well as an incentive to participate, once they completed the whole training program with 16 pooling rounds on each experiment day. The user study was reviewed by the California Institute of Technology Institutional Review Board (IRB) and determined meet the criteria for exemption pursuant to (45 C.F.R. § 46.104(d)(2)(i),(ii)): Research that only includes interactions involving educational tests (e.g., cognitive, diagnostic, aptitude, achievement tests), survey procedures, interview procedures or observation of public behavior (IRB protocol #23-1366).

**Method S8. Preparation of customized data logger**

To record quantitative data during paper-assisted pooling, we prepared a customized data logger (with grey color) using Arduino Nano Every, one barcode scanner, one weighing scale, one RTC module, and one micro SDHC 32GB card **(Supplementary Fig. S22)**. The scanner was set up for scanning a barcode if the ambient brightness was changed, allowing individual participants to scan a barcode only when they manually activated the scanner. When scanned, the barcode was transmitted to MCU (ATMega4809) via UART communication and recorded on the SD card via SPI communication. The weighing scale was prepared using a strain gauge load cell (range: 1 mg – 50 g, resolution: 1 mg) connected to a HX711 ADC module. This scale had the same performance as the scale of Scale module of instructional device. However, the data logger did not display any measured weight values to participants. When the user scanned a user card, the weight measurement began (100 ms interval), and the weight data (converted to mass units, mg) was transmitted to the MCU and recorded on the SD card in real time. Whenever a tube barcode was scanned, the MCU tared the weighing scale while recording the tube barcode on the SD card. This algorithm could allow to (a) detect any human actions regarding volume-transfer (e.g., loading the pooling tube, touching the tube, accidently removing the pooled sample from the pooling tube etc.), (b) calculate the pooled volume of sample, (c) detect whether participants scanned tube barcodes or not, and (d) track the pairs of individual and pooling tubes.

**Method S9. Preparation of sample-collection tubes**

To simulate actual diagnostic sample-collection tubes, we labeled artificial respiratory and stool samples using printed vinyl stickers (1.25 x 1.125 mm) covering 80% of the tube's side surfaces, starting 4 mm from the bottom. This 4-mm window at the bottom allowed participants to verify the presence of liquid samples and confirm successful transfer to the corresponding pooling tubes. We attached 4 x 1.4 mm 1D barcodes onto the label stickers of each tube. To eliminate any potential bias or preconceptions, we applied these labels and barcodes to both individual and pooling tubes, ensuring participants could not distinguish between them based on external appearance or labeling characteristics. We assigned barcodes SP0001 to SP0080 for individual tubes and PL0001 to PL0016 for pooling tubes in each pooling exercise. To help participants differentiate between individual and pooling tubes, we used color-coded cap inserts: red for individual tubes and green for pooling tubes. The labels and barcodes mimicked those typically found on diagnostic specimen tubes that include tracking identifiers such as collection dates and patient ID numbers.

We arranged 80 individual tubes in two tube racks (Thermo Scientific, Cat# 1480943), each capable of holding 72 tubes (6 rows x 12 columns). In each row, we placed five individual tubes in sequential barcode order (e.g., SP0001 to SP0005 in the first row). The first rack contained 60 tubes across 12 rows, while the second rack held the remaining 20 tubes across 4 rows. We placed 16 pooling tubes in sequential barcode order (e.g., PL0001 to PL0006 in the first row) across three rows (6/6/4 tubes) in the second rack, three rows away from the last five individual tubes. This arrangement allowed participants to naturally pool samples from five individual tubes in a row into a corresponding pooling tube from the second rack.

Participants were free to rearrange tubes for easier pooling. For example, participant A moved the pooling tubes from the second rack to the sixth empty position in each row, facilitating easy identification of individual and pooling tube pairs. Participant B pushed five individual tubes by one position within a row after each pooling round to differentiate between pooled and unpooled individual tubes.

**Method S10. Preparation of artificial samples**

We used the following materials to prepare artificial samples. POLYOX™ Water-Soluble Resins (WSR) Coagulant for polyethylene oxide (PEO) with molecular weight of 5 MDa was purchased from Dow (Midland, MI, USA). Bacto™ Yeast Extract (Cat# 212750; Lot# 2853522), microcrystalline cellulose (Cat# 382310010; Lot# A0442673), oleic acid (Cat# A195-500; Lot# 107295), potassium chloride (Cat# P217-500; Lot# 016778), and sodium chloride (Cat# S5886-500G; Lot# SLBL7642V) were purchased from Thermo Fisher Scientific Inc. (Waltham, MA, USA). Mucin from porcine stomach (Cat# M2378-100G; Lot# 0000258728; Source: SLCR5756) and calcium chloride dihydrate (Cat# C7902-500G; Lot# 041M00751V) were purchased from Sigma Aldrich, Inc. (St. Louis, MO, USA). Psyllium husk was purchased from Yerba Prima, Inc. (Ashland, OR, USA). Miso paste (containing protein 10.0 g, fat 5.6 g, carbohydrates 29.4 g, sodium 4.6 g, sodium chloride equivalent 11.7 g; per 100 g) was purchased from HIKARI Miso Co., LTD. (Nagano, Japan). Fluorescein sodium (Cat# 46960; Lot & Filling code: 456103/1 41606064) was purchased from Fluka Chemie GmbH (Buchs, Switzerland). De-ionized water from Milli-Q^®^ Reference water purification system was used.

Artificial respiratory samples were to mimic a type of highly viscous clinical liquid samples. The samples were prepared using PEO (5 MDa) 1.25% solution, mucin (powder) from porcine stomach, and de-ionized water. First, we prepared 20% porcine mucin in de-ionized water and stirred thoroughly at 60°C until homogenized. The homogenized 20% mucin solution was heated at 95°C for 10 minutes. Then, we added 2.5 mL of heat-treated 20% mucin and 47.5 mL of PEO (5 MDa) 1.25% in 50-mL tubes. The 50-mL tubes with the mixture were rotated by a tube rotator (Fisherbrand™ Mini Tube Rotator) at the minimum speed for ≥20 minutes and vortexed by a vortexer (Ohaus Analog Vortex Mixer) at the maximum speed. Once homogenized, artificial respiratory samples were ready for the user study.

Artificial stool samples were created to mimic clinical liquid stool samples that contain a lot of particles, gels, and fibers. The samples were prepared based on a modified version of an established protocol^4^. To make 150 g synthetic feces, we thoroughly mixed dry powders of yeast extract (11.25 g), cellulose (3.75 g), psyllium husk (6.5625 g), sodium chloride (0.75 g), potassium chloride (0.75 g), and calcium chloride dihydrate (0.375 g) in a 250 mL beaker. Next, we added 7.5 g of oleic acid oil and mixed it with the dry powders thoroughly, followed by adding miso paste (6.5625 g). The mixture was stirred thoroughly until homogenized well. Then, we poured 112.5 g of PEO (5 MDa) 1.25% solution into the beaker and stirred the mixture thoroughly until it was entirely homogenized. The homogenized mixture for synthetic feces was left standing at room temperature for 1.5 hours in the beaker covered with aluminum foil. After 1.5 hours, we diluted the synthetic feces with de-ionized water in a 1:2 ratio (w/w; one part synthetic feces to two parts water) in 50-mL tubes. The 50-mL tubes with diluted synthetic feces were vortexed thoroughly. This dilution step was to mimic the homogenized human stool suspension (stool 500 mg in 1 mL 95% ethanol) used for the clinical STH stool pooling^28^ (see the section “5-sample pooling and DNA extraction” below). Once homogenized, artificial stool samples were ready for the user study.

When artificial samples were made, we transferred 2 mL of the samples to each of 80 sample-collection tubes (12-mm) for 16 pooling rounds (5 tubes each). The volume transfer was performed by using normal P1000 pipette tips (respiratory samples) or pre-cut P1000 pipette tips (stool samples). All artificial samples were prepared within 12 hours before the start of pooling exercise of individual participants.

**Method S11. Determination of mass for 300 µL of artificial samples**

To assess volume-transfer accuracy, we measured the mass of 300 µL aliquots of artificial respiratory and stool samples using an Eppendorf P1000 micropipette and a laboratory-grade analytical balance (Denver Instrument PI-225D). For each sample type, twenty independent 300 µL aliquots were transferred into pre-tared 12-mm sample-collection tubes. The expected mass for a 300 µL volume was calculated by averaging the twenty measurements for each sample type **(Supplementary Fig. S7)**.

**Method S12. Setting for pooling exercises (user study)**

All pooling exercises were conducted using non-biohazardous materials in a conference room or similar space equipped with a desk and electricity, ensuring participant safety. The following materials were set up on the desk prior to each participant's arrival: general protective latex gloves (small, medium, large), protective eyeglasses, wiping towels (Kimberly-Clark; KCC01500), a 70% ethanol spray bottle, a secondary container for generated wastes, a pack of 100 individually wrapped 300-uL dual-bulb pipettes, two tube racks containing 80 individual sample tubes and 16 pooling tubes, and user cards with participant-specific barcodes. For the pooling of artificial stool samples, we prepared 300-uL dual-bulb pipettes with pre-cut by ~10 mm before the study.

For the volume-transfer practice (see “Initial study phase: orientation and practice on Day 1”) and paper-assisted pooling, the customized data logger was added for real-time data collection. The setup mirrored the architecture of our instructional device, with the pack of dual-bulb pipettes in the left zone, the data logger with secondary container behind it in the middle zone, and two tube racks in the right zone. This arrangement was intended to create a controlled environment that minimized any potential bias or preconceptions regarding experimental settings between paper-assisted and device-assisted pooling methods. Despite this setup, participants were allowed to rearrange materials as needed during pooling to accommodate individual preferences for natural handling pathways. For example, some participants preferred to put tube racks in the left zone and the dual-bulb pipettes and the ethanol spray in the right zone.

For paper-assisted pooling, written paper instructions **(Supplementary Note S3)** were designed and provided for all participants to maximize understandability and usability, incorporating step-by-step guidance with action diagrams that combined graphics and texts^1,2^. The procedural flow closely mirrored that of the instructional device to ensure a fair comparison between the paper instructions and the device instructions, preventing potential bias that could arise from significant differences in workflow. While maintaining the structural similarity, the paper instructions intentionally used different graphics from those on the instructional device. This choice was made to ensure participants had no prior exposure to the device's language-agnostic graphics and to avoid another potential source of bias and maintain the authenticity of participants' first interaction with the device instructions.

For device-assisted pooling, the data logger and the second container were replaced with the instructional device from the middle zone. This setup was reversed for Protocol 2 users when they returned to paper-assisted pooling for the last 4 rounds on either Day 1 or Day 2.

This approach to experimental design aimed to minimize potential biases between paper-assisted and device-assisted pooling while maintaining a consistent experimental environment across all phases of the study, thereby supporting the validity and reliability of the comparisons.

**Method S13. Initial study phase: orientation and practice on Day 1**

Prior to performing the actual pooling exercise on Day 1, each individual participant was given a short orientation (~20 minutes long) with a quick information sheet provided **(Supplementary Note S7)**. We explained to individuals step-by-step about the motivation and purpose of the user study, the value and principles of sample pooling, specific learning/training objectives from the pooling exercise, the mechanism of dual-bulb pipettes, possible causes and effects of cross-contamination, and key handling skills to avoid cross-contamination.

After the orientation, participants were also given a short practice of volume-transfer of artificial samples using dual-bulb pipettes. The purpose of the practice was to help participants learn how the dual-bulb pipettes work and get familiar with transferring samples. We set up the data logger and provided participants with 10~20 dual-bulb pipettes, a 50-mL tube containing artificial samples, and an empty 12-mm sample-collection tube without any labels, barcodes, or cap inserts. We scanned a user card to begin the weight measurement of data logger, opened the sample-collection tube, and placed the tube on the scale. Each participant wore gloves and transferred artificial liquid samples from a 50-mL tube to a 12-mm sample-collection tube. During the practice, we explained to each participant verbally about the mechanism of pipettes, such as “The liquid sample goes through and fills the entire channel, and the excess amount of sample is collected in the bottom bulb of pipette,” and “Filling the entire channel is to ensure the volume-transfer of the exact 300 uL of sample.” Individual participants performed the practice until they felt confidence in using the pipettes and expressed their confidence verbally, such as “I got confidence and am ready to start the actual pooling.” Once the practice was finished, the participants took off the gloves and disposed of them into the second container to be ready for the actual pooling exercise.

**Method S14. Pooling exercises**

Protocol 1 users performed 16 consecutive rounds of paper-assisted pooling on Day 1 and resumed 8 rounds with the paper instructions and 8 rounds with the device on Day 2 **(Fig. 3*A*)**. Protocol 2 users completed a total of 16 rounds each Day 1 and Day 2: 4 rounds of paper-assisted pooling, followed by 8 rounds of device-assisted pooling, and finishing with 4 rounds of paper-assisted pooling **(Fig. 3*B*)**.

In each pooling round, participants were instructed to pool artificial samples from five individual sample tubes to a corresponding pooling tube, guided by either paper or device instructions. Breaks were allowed between rounds and sessions to prevent fatigue, but participants were required to complete all 16 rounds each day unless technical issues, such as significant device wire disconnections, occurred.

To ensure fair comparisons between instructional tools both within subjects and across protocol groups, several standardized approaches were implemented for the pooling exercises. Before the start of paper-assisted pooling on Day 1, we recommended participants to read the paper instructions beforehand, though it was not mandatory. Once they felt comfortable with the instructions, they could start the paper-assisted pooling at their own discretion. During the paper-assisted pooling, participants were free to refer to the paper instructions at any time.

Participants were encouraged to ask a researcher for clarification if any steps were confusing or unclear during the pooling with either paper or device instructions. However, researchers did not provide interventions for high-performance sample pooling. For example, they did not teach specific volume-transfer skills for accurate and reliable pooling artificial samples, even when participants inquired about such techniques. Instead, researchers reminded participants of the mechanism of dual-bulb pipettes and encouraged them to develop their own skills independently.

These measures were designed to create a comfortable environment, promote a more natural interaction with the instructions for the pooling exercises, and ensuring that learning and training stemmed from the instructional tools rather than from researcher input. Additionally, these approaches aimed to alleviate any potential anxiety or nervousness participants might feel about being evaluated on personal attributes such as intelligence or cognitive abilities. By implementing these standardized approaches, the study sought to provide a fair and comprehensive comparison between paper and device-assisted pooling instructions.

**Method S15. Data collection during pooling exercises**

For the number of handling errors, a researcher monitored participants' handling errors throughout the pooling exercises. Only uncorrected handling errors were recorded based on a handling error check sheet **(Supplementary Fig. S23)**. These uncorrected handling errors included (1) correctable errors that were not corrected by participants or were corrected only after compromising the biosafety and sample integrity, (2) fundamentally uncorrectable errors that permanently compromise sample traceability, integrity, and reliability, and (3) correctable errors that were not corrected during the procedure, though they could have been safely remedied without compromising biosafety or sample integrity **(Supplementary Note S4)**.

For pooling performance during the paper-assisted pooling, the customized data logger was used for time-series weight measurements (100 milliseconds interval) and documentation of scanned barcodes. The data was saved to individual text files on an SD card, with each file corresponding to a specific participant. When participants failed to scan tube barcodes, a researcher manually recorded both individual and pooling tube barcodes on the scorecard. These missed scans not only resulted in missing barcode data but also affected weight measurements, as barcode scanning was required to tare the scale between samples. Without taring, the scale recorded cumulative weights rather than individual sample volumes, requiring manual tracking to determine actual sample volumes transferred. For each pooling round, a researcher recorded the start and end timestamps to calculate each participant’s completion time for the pooling process.

During the device-assisted pooling, the instructional pooling device recorded four key elements on an SD card: weight measurements of pooled samples, tube barcode scans, detectable handling errors, and the user’s completion time for the pooling process. For weight measurements, the device followed a systematic process: it tared the pooling tube during individual pipette unwrapping, monitored weight changes every 100 milliseconds, validate the weight measurements of sample dispensing, and detected tube stabilization (CV ≤1%) after the dispensing. Once stable, the device sampled weights every 200 milliseconds and calculated the average weight to estimate the actual pooled sample volume. When participants accidentally removed volume from pooling tubes before adding new samples, the device recorded both negative stabilized weights (pre-transfer) and final weights (post-transfer) of pooling tube as well as time-series weight values. In these cases, weights of actual pooled sample were manually calculated from these records. To ensure data integrity, the device maintained a running log of all operations in case of unexpected power loss or system interruption. Critical events, such as error detections and weight measurements, were immediately written to the SD card rather than stored in temporary memory.

**Method S16. Post-experimental procedures**

On Day 1, all participants responded to the demographic survey, followed by the Kirkpatrick Training Evaluation Questionnaire **(KTEQ; Supplementary Fig. S8c)**. The subsequent steps differed by protocol: Protocol 1 participants completed the Post-Study System Usefulness Questionnaire (PSSUQ) for paper instructions (PSSUQ-Paper), while Protocol 2 participants completed both the PSSUQ for the device (PSSUQ-Device) and the PSSUQ-Paper, in that order. These PSSUQ versions were modified from the original^5^ and tailed to our specific sample pooling schemes **(Supplementary Tables S6–7)**.

On Day 2, Protocol 1 participants responded to both the PSSUQ-Device and the Post-Training Evaluation Questionnaire (PTEQ), while Protocol 2 participants responded only to the PTEQ. Questionnaire items 1–3 were asked to all participants who participated in Day-2 exercises. All Day-2 participants answered PTEQ items 1–3 **(Supplementary Fig. S8d–f)**.

All surveys were prepared using the online survey platform, Qualtrics, and delivered through QR codes that direct to online survey links for questionnaire. After the pooling exercises, participants were guided to scan QR codes to respond to the questionnaire. Participants received compensation ($30–$60) after completing the surveys on each experiment day.

**Method S17. Data analysis in the user study**

All device data were extracted and analyzed from device log files and user pooling data files automatically recorded by the customized data logger and instructional pooling device. All data extraction and organization were performed using Microsoft Excel (version 2408). All statistical analyses were implemented in Python (version 3.11.10) with statsmodels (version 0.14.4), scikit-learn (version 1.5.1), scipy (version 1.9.3), and dabest^6^ (version 2024.3.29) packages.

To test statistical differences in usability between paper instructions and device instructions, we used paired, one-sided Wilcoxon signed-rank tests for non-normally distributed PSSUQ scores shown in **Fig. 3*C*,*D*** and **Supplementary Figs. S8a,b** and **S9**. The PSSUQ scores for SysUse, InfoQual, and InterQual were calculated by averaging scores of items 1–8, items 9–15, and items 16–18, respectively, per participant. To avoid Type I errors from the multiple comparisons, *P* values were corrected using the Benjamini-Hochberg correction method (5% false discovery rate). The statistical analysis results are presented in **Supplementary Table S8**.

To evaluate whether the majority (defined as ≥75%) of participants responded to Likert score 4 or 5 for KTEQ items and PTEQ item 1, we used one-sided Binomial tests (threshold: 75%) for Protocol 1 and 2 groups shown **Supplementary Fig. S8c,d**. To test statistical differences in Likert scores between PTEQ items 2 and 3, we used paired, one-sided Wilcoxon signed-rank tests. To avoid Type I errors from the multiple comparisons, *P* values were corrected using the Benjamini-Hochberg correction method (5% false discovery rate). The statistical analysis results are presented in **Supplementary Table S9**.

Using recorded weight data, we assessed the volume-transfer performance using two metrics: accuracy and precision. First, volume-transfer accuracy (in percentage, %) for pooled sample from each individual tube was calculated using the formula:

Acc_pool_ [%] of each pooled sample = (1 − |the mass for 300 µL − the measured weight (in mass units, mg) of pooled sample|/the mass for 300 µL) × 100 [%]

Then, the volume-transfer accuracy (defined as Acc_pool_ in **Figs. 4*C*,*D*** and **5*C*,*D***) for each pooling round was calculated by averaging all accuracy values for samples pooled into each pool. The volume-transfer precision for each pooling round was determined using the coefficient of variation (CV_pool_):

CV_pool_ [%] = (standard deviation/average of measured weight (in mg) of samples pooled in a pool) × 100 [%]

To assess how instructional tools (the paper and the device instructions) influenced volume-transfer performance, we calculated the average volume-transfer accuracy (Avg. Acc_pool_) for each pooling session by collapsing and averaging all accuracy data across four corresponding pooling rounds. For within-subject analysis, we conducted paired, one-tailed *t*-tests and then calculated Hedges’ g effect sizes to evaluate whether the average accuracy improved in subsequent pooling sessions relative to the initial baseline session (#1, BASE). Using the dabest library, we generated Gardner-Altman plots with effect sizes and their bootstrap 95% confidence intervals with 5,000 iterations, shown in **Figs. 4*G*–*H*** and **5*G*–*H***. The statistical analysis and calculated results are presented in **Supplementary Tables S12 and S22**.

Using manually recorded uncorrected handling errors, we assessed the handling practices of participants. The number of uncorrected handling errors per pooling round (defined as *err*_all_ in **Figs. 4*A*,*B*** and **5*A*,*B***) was calculated by the summation of all uncorrected errors from the records on scoreboards. Depending on the severity level **(Supplementary Table S10)**, we classified the uncorrected errors into severe errors and minor errors **(Supplementary Fig. S11)**. To assess how instructional tools influenced handling practices, we collapsed all data of the number of uncorrected errors across four pooling rounds and did summation of the errors for each pooling session. For within-subject analysis, we conducted paired, one-tailed Wilcoxon signed-rank tests and calculated matched-pairs rank biserial correlation coefficients^7^, *r*_C_, to evaluate whether the number of uncorrected errors (Σ*err*_all_, Σ*err*_minor_, and Σ*err*_severe_) decreased in subsequent pooling sessions relative to the initial baseline session (#1, BASE). Using the dabest library, we generated Gardner-Altman plots with effect sizes and their bootstrap 95% confidence intervals with 5,000 iterations, shown in **Figs. 4*E*–*F*** and **5*E*–*F*** and **Supplementary Fig. S12**. The statistical analysis and calculated results are presented in **Supplementary Tables S11, S19–21, and S23–24**.

Considering two volume-transfer metrics (Acc_pool_≥80% and CV_pool_≤25%), we counted the number of pools through high-performance volume transfer (defined as High-Quality Pools in **Supplementary Note S6**) across four pooling rounds per session. We conducted paired, one-tailed Wilcoxon signed-rank tests and calculated matched-pairs rank biserial correlation coefficients, *r*_C_, to evaluate whether the number of pooling rounds with high-performance volume-transfer increased in subsequent sessions relative to the initial baseline session (#1, BASE). Using the dabest library, we generated Gardner-Altman plots with effect sizes and their bootstrap 95% confidence intervals with 5,000 iterations, shown in **Supplementary Fig. S16**. The statistical analysis and calculated results are presented in **Supplementary Tables S30–31**.

Considering high-performance volume-transfer and the number of uncorrected errors, we counted the number of high-performed pooling rounds (Acc_pool_≥80%, CV_pool_≤25%, zero number of uncorrected errors) across four pooling rounds per session. We conducted paired, one-tailed Wilcoxon signed-rank tests and calculated matched-pairs rank biserial correlation coefficients, *r*_C_, to evaluate whether the number of pooling rounds with high-performance pooling increased in subsequent sessions relative to the initial baseline session (#1, BASE). Using the dabest library, we generated Gardner-Altman plots with effect sizes and their bootstrap 95% confidence intervals with 5,000 iterations, shown in **Fig. 6*A*–*D***. The statistical analysis results are presented in **Supplementary Tables S33–34**.

For all paired statistical tests, the data from Protocol 2 users who did not participate in the Day-2 pooling exercise were not included. For one-tailed Wilcoxon signed-rank tests, zero differences were included in the ranking process, and the zero ranks were split between positive and negative ones. Levels of significance, α, was set at 0.05.

To control for the increased risk of Type I errors that occurs when conducting multiple statistical tests (i.e., one-sided paired *t*-tests and one-sided Wilcoxon signed-rank tests), we complemented our analyses with linear mixed-effects models, which allowed for simultaneous testing of all effects. The linear mixed-effects models included fixed effects of ‘Session’ and ‘Group’ as well as their interaction. ‘Group’ was coded as a categorical variable with two levels (Protocol 1 and Protocol 2). The ‘Session’ variable was defined differently for each analysis: five levels (sessions #1, #2, #3, #6, and #7) for assistive effects and three levels (sessions #1, #4, and #8) for training effects. Prior to analyses, the dependent variables, ‘Performance,’ were transformed. The average volume-transfer accuracy (Avg. Acc_pool_) was logit-transformed to address the bounded nature of accuracy measures and ensure the assumption of linearity was met. The non-normally distributed data (e.g., Σ*err*_all_, High-Performed Pooling Rounds, etc.) were transformed by Yeo-Jonhson transformation to ensure normality. After the transformation, the analyses were performed by using the following model specification:

Performance ~ Session + Group + Session × Group + (1|Participant)

where Performance was the dependent variable (e.g., Avg. Acc_pool_ etc.), Session and Group were categorical predictors, and (1|Participant) represents random intercepts for participants. To maintain consistency with each Gardner-Altman plot, either Protocol 1 or Protocol 2 served as the reference category in the dummy coding scheme, with session #1 (BASE) consistently serving as the reference category for Session. Model parameters were estimated using maximum likelihood estimation (MLE) with the Powell optimization method (maximum 2,000 iterations). The statistical analysis results are presented in **Supplementary Tables S11–12, S19–24, S30–31, and S33–34**.

To investigate the changes in completion time (*t*_pool_) across pooling rounds, linear mixed-effects models were fitted separately for each Protocol group. The models included fixed effects of ‘Device’ and ‘Pooling Round’ as well as their interaction. ‘Device’ was coded as a binary predictor (paper=0, device=1) to distinguish between paper-assisted and device-assisted pooling rounds. Prior to the analyses, completion time was standardized within each Protocol group. The model specification was:

Completion Time ~ Device + Pooling Round + Device × Pooling Round + (1|Participant)

where Completion Time was the standardized outcome for completion time taken to complete the pooling process in each round. Device was a binary predictor. The same model parameters (e.g., MLE and optimization) were used. To assess the rate of change in completion time during device-assisted pooling rounds, linear contrasts were constructed by summing the coefficients for Pooling Round and Device × Pooling Round. The combined effect and its exact *P* value were estimated using a Wald z-test. The statistical analysis results are presented in **Supplementary Tables S13 and S25**.

To examine how completion time influenced performance metrics across pooling rounds, linear mixed-effects models, while controlling for completion time, were fitted within each Protocol group. Models included fixed effects of ‘Device,’ ‘Pooling Round,’ ‘Completion Time,’ and their two-way interactions. Prior to analyses, the dependent variables, ‘Performance,’ were transformed: Acc_pool_ was logit-transformed, and error metrics (i.e., *err*_all_, *err*_minor_, and *err*_severe_) were Yeo-Jonhson transformed. The model specification was:

Performance ~ Device + Pooling Round + Completion Time + Device × Pooling Round + Device × Completion Time + (1|Participant)

where Performance was the dependent variable (e.g., Acc_pool_ etc.). The same model parameters (e.g., MLE and optimization) were used. Linear contrasts were used to estimate (1) the progression of performance metrics across device-assisted pooling rounds (Pooling Round + Device × Pooling Round) and (2) the relationship between completion time and performance metrics under device-assisted conditions (Completion Time and Device × Completion Time). The statistical analysis results are presented in **Supplementary Tables S14–15 and S26–27**.

To investigate whether differences in completion time affected the original session-based analyses, linear mixed-effects model analyses were fitted with completion time as a covariate. The models included fixed effects of ‘Session,’ ‘Group,’ ‘Completion Time,’ and ‘Session × Group’ interaction. The same dummy coding approach was used with the primary analyses. The model specification was:

Performance ~ Completion Time + Session + Group + Session × Group + (1|Participant)

where Performance was the dependent variable (e.g., Avg. Acc_pool_ etc.), Completion Time was the standardized variable of average Completion Time across each session (1 session = 4 rounds). The same model parameters (e.g., MLE and optimization) were used. The statistical analysis results are presented in **Supplementary Tables S16–17 and S28–29**.

To examine the assistive and training effectiveness of instructional pooling device on the performance of individual Protocol 2 participants, we conducted McNemar’s Exact tests in the analyses for high-quality pools and high-performed pooling rounds. Each 2x2 contingency table was designed to examine how many participants improved or decreased their performance from the initial session (#1, BASE) to subsequent sessions (#2–#8) as the effects from the instructional device. “High” and “Low” in the tables were defined as individual participants who achieved high-performance volume-transfer or high-performance sample pooling for ≥2 and <2 rounds, respectively, in the pooling session. To avoid Type I errors from the multiple comparisons, *P* values were corrected using the Benjamini-Hochberg correction method with 5% false discovery rate. The statistical analysis results are presented in **Supplementary Tables S32 and S35**.

To test the statistical differences in the rates of *Incorrect* pools and *Invalid* pools between paper-assisted and device-assisted pooling among Protocol 2 participants, we used one-sided Fisher’s Exact tests. The null hypothesis was that paper-assisted pooling would produce equal or fewer errors than device-assisted pooling. The complete contingency tables and statistical analysis results are presented in **Supplementary Table S36**.

**Method S18. Clinical STH samples and commercial stool samples**

A panel of 159 clinical stool samples (500 mg each) was prepared at Smith College (Northampton, MA, USA) and shipped to California Institute of Technology (Pasadena, CA, USA). The samples were collected from children aged 22 months to 12 years (mean age = 57 months) enrolled in the WASH Benefits Bangladesh trial between May 2015 and May 2016 (Clinicaltrials.gov NCT01590095)^8^.

Initial screening at Smith College using real-time PCR revealed that among the 159 samples, 70 were positive for *Ascaris lumbricoides* alone, six were positive for both *A. lumbricoides* and at least one other soil-transmitted helminth (STH) species (*Necator americanus*, *Trichuris trichiura*, *Ancylostoma duodenale*, *A. ceylanicum*, or *Strongyloides stercoralis*), and 83 were negative for *A. lumbricoides*. Among 159 samples, four samples were excluded from this study: three negative samples and one positive sample (pre-screened Cq value = 23.042) due to the absence of actual fecal matter. Of the remaining 155 samples, four positive samples (pre-screened Cq values of 18.388, 24.829, 24.867, and 30.92) were used for testing the qPCR assay. Additionally, six samples were excluded from the pooling study due to discrepancies in the reported versus observed sample positivity. Overall, the exclusions resulted in a final dataset of 145 samples, comprising 71 positive and 74 negative samples. Of the 71 positive samples, 21 were selected to include weak positives (pre-screened Cq values ≥ 30) while maintaining the Cq distribution of samples with pre-screened Cq values ≤ 30. From the 74 samples negative for *A. lumbricoides*, 21 were randomly selected, of which at least 12 were positive for at least one STH species (*Necator americanus*, *Trichuris trichiura*, *Ancylostoma duodenale*, *A. ceylanicum*, or *Strongyloides stercoralis*) based on Smith College's qPCR analysis.

Each pool consisted of one clinical sample combined with four negative stool samples obtained from unique healthy individuals (Medix Biochemica, Inc., Cat# 991-18-1). All negative samples were confirmed to be free of *Ascaris lumbricoides* DNA through triplicates of DNA extraction with technical triplicate on qPCR analysis.

**Method S19. 5-sample pooling and DNA extraction**

Before starting the validation, two levels of randomization were implemented to simulate real-world conditions and eliminate potential handling bias. First, we randomized the sequence in which clinical samples were processed for pooling, ensuring that samples with different infection intensities (strong positives, moderate positives, weak positives, and negatives, as determined by Cq values) were tested in random order. Second, for each pooling round, we randomly assigned the position of the clinical sample tube among the five slots on the pooling device, with the remaining four slots filled by commercial negative sample tubes. This dual randomization strategy prevented any systematic bias in sample handling or processing.

To validate the utility of our instructional pooling device in its intended use case, we simulated the expected workflow where device-trained personnel would perform sample pooling on the device, followed by skilled clinical technicians conducting downstream analysis. In this study, one device-trained laboratory personnel performed all pooling processes using the instructional device, while a different laboratory technician conducted the subsequent DNA extraction and qPCR analysis. To prevent bias, the pooling operator was blinded to the infection status and intensity (Cq values) of the clinical samples before pooling.

Pooling experiments followed a validated STH pooling protocol^8^ with minor modifications **(Supplementary Fig. S24a)**, using the MP Bio Fast DNA SPIN Kit for Soil (MP Biomedicals; Santa Ana, CA, USA). Each 500-mg stool sample was divided into two approximately 250-mg aliquots in Lysing Matrix E tubes containing beads, lysis buffer (728 µL sodium phosphate buffer and 122 µL MT buffer), and 200 µL 95% ethanol. The suspensions were homogenized using a bead beater (Beadbug 6, Benchmark Scientific, Inc.; Sayreville, NJ, USA) at 7.0 m/s for 90 seconds. After centrifugation (Centrifuge 5420, Eppendorf; Nijmegen, Netherlands) at 14,000 x g for 10 minutes, the supernatants were recombined into a 12-mm sample-collection tube.

Stool suspensions from commercial negative samples were prepared in advance and stored at –80 °C until needed for pooling with clinical stool suspensions. The commercial stool suspensions were heated at 70 °C for 10 minutes before pooling to remove salt precipitation. For each pooling round, five 12-mm sample-collection tubes containing homogeneous suspensions (one clinical sample and four commercial negative samples) were prepared. Using 300-µL dual-bulb pipettes, pooling was performed on the instructional pooling device, with data recorded separately for each clinical sample. The resulting ~1.5 mL pool was split equally between two Lysing Matrix E tubes (containing beads only) and homogenized using the same bead-beating conditions described above.

For DNA extraction, different aliquoting strategies were used based on infection intensity of clinical samples. For pools and individual suspensions from clinical positive samples with pre-screened Cq values ≥ 30, two 300-µL aliquots were prepared in Lysing Matrix E tubes containing lysis buffer. For all other pools and individual suspensions, one 300-µL aliquot was prepared. DNA extraction followed a validated extraction protocol^9,10^, beginning with an additional bead-beating step (representing the second homogenization for individual samples and third for pooled samples). Internal amplification control (IAC) plasmid was spiked into each extraction according to the established protocol to ensure extraction performance^10^ **(Supplementary Fig. S24b)**. The extracted DNA was either analyzed by qPCR on the same day or stored at –80°C until analysis.

**Method S20. qPCR analysis**

The qPCR analysis for detecting *Ascaris lumbricoides* and IAC followed a published protocol^10,11^. For all extracted DNA, qPCR triplicates were run for 50 cycles using CFX96 Real-Time System (Bio-Rad Laboratories, Inc.; Hercules, CA, USA). No-template control using nuclease free water was included in all qPCR plates. Individual samples are considered positive if all triplicates have Cq values below 50. For weak positive clinical samples, Cq values from both extraction replicates were included in the analysis. With the same protocol, extractions of stool from healthy human were validated to be negative to *Ascaris lumbricoides.* Limit of blank was tested by running qPCR targeting *Ascaris lumbricoides* DNA on 20 replicates of extracted elution of the commercial healthy human stool **(Supplementary Fig. S24c)**.

**Method S21. Data analysis in the device validation study**

Like the user study, all device data were extracted and analyzed from device log files and user pooling data files automatically recorded by the instructional pooling device. All data extraction and organization were performed using Microsoft Excel (version 2408). All statistical analyses were implemented in Python (version 3.11.10) with scikit-learn (version 1.5.1), scipy (version 1.9.3), and dabest^6^ (version 2024.3.29) packages.

To test the statistical differences in the weight of transferred liquid across homogenized stool suspensions, we used the Kruskal-Wallis test for non-normally distributed data confirmed by the Shapiro-Wilk Normality test. The statistical analysis results are presented in **Supplementary Table S37**.

Using the dabest library, we generated Gardner-Altman plots with the mean differences in Cq values, as effect sizes, and their bootstrap 95% confidence intervals with 5,000 iterations, shown in **Fig. 8*F***. To evaluate the correlation of Cq values between individual and pooled positive samples, we performed the linear regression and calculated the Pearson’s *r* coefficient.

**Method S22. Determination of PPA and NPA**

We assessed the diagnostic performance of pooled qPCR testing with the instructional device, relative to our individual qPCR testing. We created the 2x2 contingency matrix **(Fig. 8*G*)** to evaluate the agreement in the diagnostic results between individual testing, considered as the ‘ground truth’ here, and pooled testing. From the matrix table, we calculated positive percent agreement (PPA) and negative percent agreement (NPA) using the formula:

PPA = the number of detected positive pools / the number of total positive clinical samples

NPA = the number of detected negative pools / the number of total negative clinical samples
